# Safety and immunogenicity of PanChol, a single-dose live-attenuated oral cholera vaccine: results from a phase 1a, double-blind, randomized, placebo-controlled trial

**DOI:** 10.1101/2025.09.12.25335655

**Authors:** Deborah R. Leitner, Stephen R. Walsh, Masataka Suzuki, Michaël Desjardins, Alisse Hannaford, Amy C. Sherman, Hannah Levine, Lena Carr, Elliot Hammerness, Akina Osaki, Emily Sullivan, Bryan Wang, George I. Balazs, Jun Bai Park Chang, Damien M. Slater, Nirajan Puri, Carole J. Kuehl, Wilbur H. Chen, Jason B. Harris, Steven Piantadosi, Lindsey R. Baden, Matthew K. Waldor, the PanChol study group

**Affiliations:** Division of Infectious Diseases, Brigham and Women’s Hospital, Boston, MA, USA; Department of Microbiology, Harvard Medical School, Boston, MA, USA; Department of Medicine, Harvard Medical School, Boston, MA, USA; Division of Infectious Diseases, Massachusetts General Hospital, Boston, MA, USA; Department of Pediatrics, Harvard Medical School, Boston, MA, USA; Center for Clinical Investigation, Brigham and Women’s Hospital, Boston, MA, USA; Center for Vaccine Development and Global Health, University of Maryland School of Medicine, Baltimore, Maryland, USA; Department of Surgery, Brigham and Women’s Hospital, Harvard Medical School, Boston, MA, USA

## Abstract

**Background:** Current whole-cell killed oral cholera vaccines have utility but require multiple doses and have limited efficacy in young children. PanChol is a single-dose live-attenuated cholera vaccine derived from the current seventh pandemic *Vibrio cholerae* O1 strain. It co-expresses Inaba and Ogawa antigens, over-expresses the non-toxic cholera toxin B subunit, and is designed to minimize reactogenicity and prevent toxigenic reversion. We assessed safety and immunogenicity in a first-in-human trial.

**Methods:** In a dose-escalation phase at Brigham and Women’s Hospital (Boston, MA, USA), seven cohorts received one dose of 10^4^-10^10^ colony-forming-units (CFU) PanChol. Two dosing groups of 2×10^7^ and 2×10^8^ were subsequently evaluated in a double-blind, placebo-controlled module. Fecal shedding was assessed until day five; safety and immunogenicity were monitored for six months. This trial is registered with ClinicalTrials.gov, NCT05657782.

**Findings:** Between Dec 2022 and Feb 2025, 57 healthy adults were enrolled (dose-escalation: n=21; expansion n=28 vaccine and n=8 placebo recipients). PanChol was safe and well-tolerated at all doses, and no safety concerns were identified including no vaccine-related serious adverse events. In the dose-escalation phase, 81% (17/21) of participants had 39 unsolicited adverse events (AE). In the randomized module, at least one AE occurred in 64% (9/14) at 10^7^ dose and in all 10^8^ (13/13) and placebo (7/7) recipients. Most AEs were mild and only four were >grade 2 (all unrelated to vaccine). Shedding was detected in 44 recipients of ≥10^5^ CFU, with no relationship to dose. All 45 vaccinees given ≥10^5^ CFU seroconverted vibriocidal antibodies to both serotypes, with comparable mean titers across doses. IgM responses targeting Inaba or Ogawa polysaccharides were detected in 44 and 41 vaccinees respectively, and anti-toxin IgG responses were measured in 21 vaccinees. Antibody lymphocyte supernatant assays demonstrated mucosal IgA to these antigens and to colonization factor, TcpA.

**Interpretation:** A single oral dose of PanChol induced 100% vibriocidal seroconversion over a 100,000-fold dose range with no safety concerns. These findings support further development of PanChol as a new tool for cholera prevention, including studies in endemic settings and in children.

**Funding:** Wellcome Trust.

**Research in context:** *Evidence before this study:* Cholera remains a global public health threat; killed oral cholera vaccines (OCVs) are important for control. However, they have limited efficacy in young children and require multiple doses for maximum efficacy. Live-attenuated OCVs, like natural infection, may induce protective immunity with a single dose. While other live-attenuated OCVs have been developed, none are WHO prequalified. We searched PubMed from inception to July 2025 for studies evaluating OCVs using the terms “live oral cholera vaccine trial O1”, yielding 25 non-review clinical trial articles. All tested live vaccines were derived from the extinct classical *V. cholerae* biotype or early El Tor biotype strains and were Inaba serotype. None were engineered to be resistant to reversion to toxigenicity.

*Added value of this study:* This first-in-human trial establishes that PanChol, a live-attenuated OCVs engineered from the current global pandemic El Tor *V. cholerae* O1 strain, is safe and immunogenic in an adult population in Boston, USA. Unlike previous live-attenuated vaccines, PanChol expresses both Inaba and Ogawa serotype antigens, is engineered for enhanced genetic stability, and resists toxigenic reversion. PanChol shedding, a marker for vaccine replication in the intestine, was detectable across doses 10^5^-10^10^ CFU. Whole genome sequencing of PanChol isolated from vaccinees’ stool confirmed the vaccine’s genomic stability. Across all doses, 100% of vaccinees seroconverted to both Inaba and Ogawa serotypes, demonstrating potent immunogenicity.

*Implications of all the available evidence:* PanChol’s favorable safety profile and immunogenicity support additional development as a new agent for cholera control. Since PanChol is derived from the current pandemic strain, and natural infection stimulates more potent immunity to cholera than killed vaccines, PanChol may offer effective single-dose protection for children. It may be beneficial for reactive vaccination campaigns and for alleviating the global shortage of killed OCVs. These positive results warrant the establishment of the vaccine’s safety and immunogenicity in cholera endemic settings and age de-escalation trials.

## Introduction

Vaccines are a key tool for the prevention and elimination of cholera, an acute and potentially rapidly lethal diarrheal disease^1^. It is estimated that there are 1·3-4 million cases of cholera and 21,000-143,000 deaths annually, with at least 1 billion people at risk of disease^1^. Cholera is caused by ingestion of cholera-toxin producing strains of *Vibrio cholerae*, a curved gram-negative rod. *V. cholerae* possesses an unusual capacity to colonize the human small intestine. While proliferating in the small intestine, the pathogen secretes cholera toxin. Genes encoding this AB5 type toxin, *ctxAB*, are not an ancestral part of the *V. cholerae* genome but instead are located within a prophage (CTXϕ) that was acquired by horizontal gene transfer^2^. Amongst diarrheal diseases, cholera is distinctive because it causes global pandemics in which a single strain becomes predominant throughout the globe. All recorded cholera pandemics, including the ongoing seventh pandemic, have been caused by the O1 serogroup, which is divided into two major serotypes – Inaba and Ogawa.

At present, the only World Health Organization cholera vaccines approved for use in cholera endemic areas or in outbreaks are oral killed whole-cell vaccines that consist of killed Inaba and Ogawa *V. cholerae*^3^. Clinical trials have shown that these vaccines, which primarily represent oral administration of *V. cholerae* LPS, confer an average of two dose protective efficacy of 55% with some studies showing rapid waning efficacy after two years^4^. While an advance for cholera control, these vaccines have important limitations, including the requirement of two doses for maximum efficacy and their limited efficacy (less than 30%) in children less than five years of age, the group most susceptible to death from cholera^4^.

Studies of both naturally occurring and experimentally induced human cholera have revealed that infection stimulates protective immunity against subsequent cholera^5,6^. These observations have inspired the creation of live-attenuated cholera vaccines. Although previous live-attenuated vaccines such as Vaxchora^7^ and Peru-15^8^ have shown promise as single-dose vaccines to engender robust and protective immune responses including children <5 years of age^9,10^, they have important shortcomings. Vaxchora was created in the now extinct and more virulent classical *V. cholerae* biotype and is capable of reversion to toxigenicity.

PanChol, named for pandemic cholera vaccine, is a novel live-attenuated cholera vaccine derived from the *V. cholerae* O1 strain that is representative of the current pandemic El Tor *V. cholerae* biotype, the etiology of the vast majority of global cholera^11^. PanChol was extensively engineered to ensure its biosafety for individuals and communities and protect its genetic stability while leaving intact its capacity for colonization (table 1)^11,12^. PanChol expresses both Inaba and Ogawa serotype antigens^12^ and over-expresses the B subunit of cholera toxin (CT-B) because immune responses to this non-toxic part of cholera toxin may confer some protection against cholera as well as diarrhea from enterotoxigenic *E. coli* (ETEC), which expresses a related toxin^13^. In addition to deletion of *ctxA* and other potential toxins linked to reactogenicity^14^, PanChol is resistant to reversion to toxigenicity because it contains deletions of the entire CTX prophage, the site, where the CTXϕ genome integrates into the *V. cholerae* chromosome, and *recA*, a gene linked to recombination. Moreover, a CRISPR system that recognizes and degrades *ctxA* was introduced into the PanChol genome, rendering the vaccine strain resistant to re-acquisition of this cholera toxin gene by means of horizontal gene transfer. In pre-clinical studies in infant rabbits and mice, PanChol was safe, immunogenic, and elicited protection in lethal challenge models^12^. Here, we report results of a first-in-human trial of a single dose of PanChol in healthy volunteers.

**Table 1:** Genetic design features of PanChol.

| Design principles | Genetic features of PanChol |
| --- | --- |
| **Target antigens** |  |
| Globally predominant strain | A derivative of 2010 Haiti outbreak |
| Bivalent expression of Inaba and Ogawa serotypes | Hypomorphic WbeT methylase |
| Provide short-term protection vs ETEC | Overexpression of CT-B (*ctxB*) |
| **Safety** |  |
| Remove capacity to make cholera toxin | Deletion of *ctxAB* and entire CTX prophage |
| Eliminate reactogenicity | Deletion of flagellins, *hlyA* and *rtxA* |
| Sensitive to antibiotics, incapable of transfer | Deletion of most of SXT mobile element |
| **Genetic stability (no reversion)** |  |
| Eliminate capacity to regain CTX phage | Deletion of CTX phage attachment site |
| Prevent acquisition of *ctxA* gene | CRISPR targets *ctxA* for destruction |
| Eliminate capacity for recombination | Deletion of *recA* |

## Methods

### Study Design

A first-in-human study was conducted at Brigham and Women’s Hospital (Boston, MA, USA) to evaluate the safety, tolerability, and immunogenicity of PanChol in healthy adults. The research protocol was approved by the Mass General Brigham Healthcare Institutional Review Board and Institutional Biosafety Committee and conducted under United States Food and Drug Administration (FDA) Investigational New Drug. 15 participants, three per dose, were first enrolled in an open-label fixed dose-ranging module to evaluate safety (range of PanChol 10^6^-10^10^ colony-forming-units (CFU)), followed by a placebo-controlled expansion module (figure 1). Prior to each log 10 dose-escalation from 10^6^ to 10^10^, the study team reviewed reactogenicity (solicited) and unsolicited adverse events (AEs) reported by participants to ensure that no dose-limiting toxicities (defined as grade 3 in accordance with the FDA Toxicity Grading Scale for Healthy Adult and Adolescent Volunteers Enrolled in Preventive Vaccine Clinical Trials, September 2007 and deemed related to vaccination) had occurred. A Data and Safety Monitoring Board was convened and reviewed unblinded data from the dose-escalation cohort prior to commencement of the expansion module, where a larger number of participants were given 2×10^7^ (n=14), 2×10^8^ CFU (n=14) or placebo (n=8). The 10^4^ (n=3) and 10^5^ CFU (n=3) PanChol groups were added via an amendment following completion of the dose-expansion cohort and were conducted in a dose-de-escalation fashion to determine the minimum dose of PanChol, which could lead to colonization and immunogenicity. This study is registered with ClinicalTrials.gov, NCT05657782.

**Figure 1:**
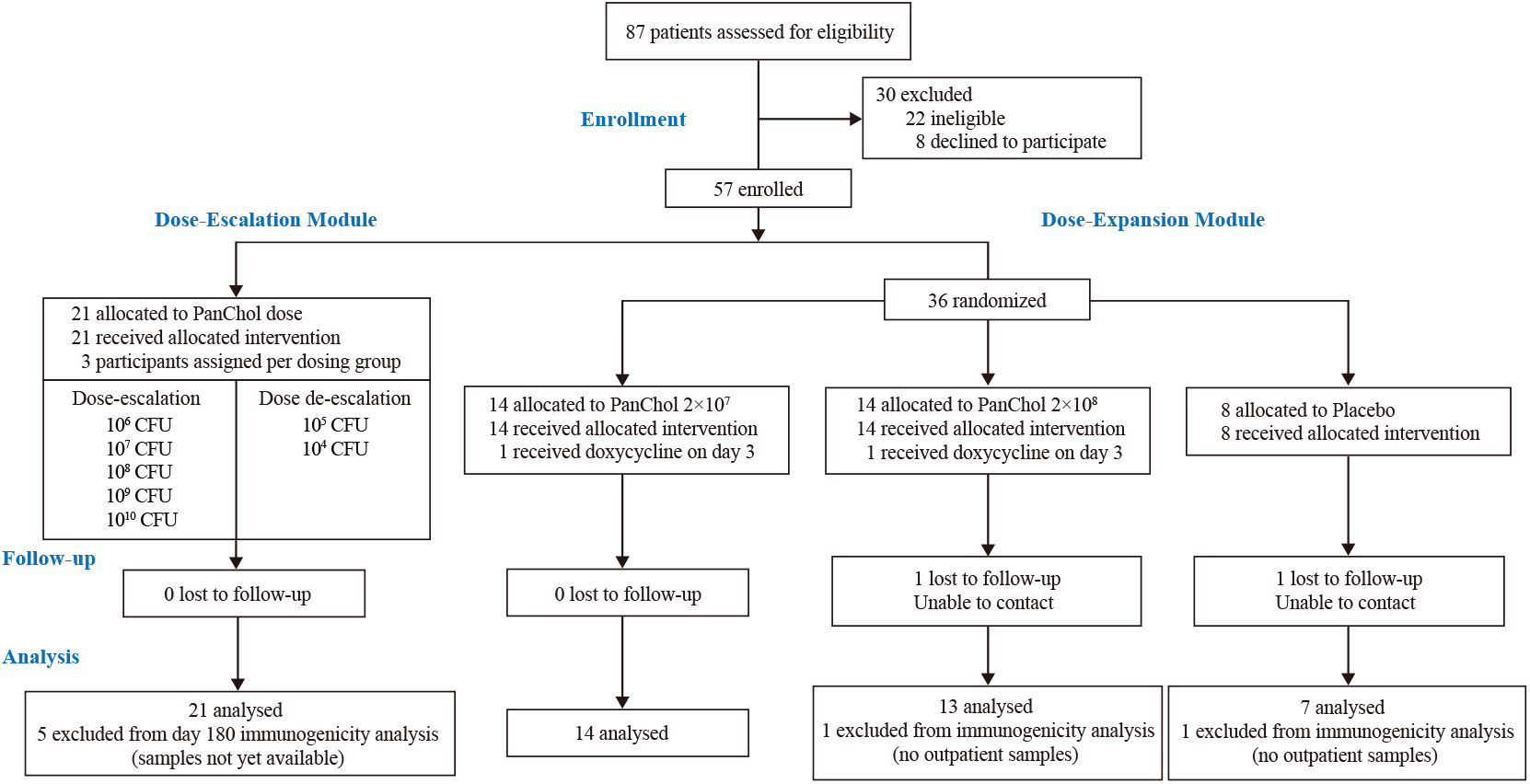
Trial profile.

### Participants

Healthy adults, aged 18-55 years old, were recruited through the MassGeneral Brigham Rally volunteer portal. Volunteers were screened and enrolled if they were deemed medically healthy and met eligibility criteria. Participants were excluded if they had a history of gastrointestinal (GI) disorders, recent fever, GI illness or diarrhea, prior cholera infection or vaccination, abnormal stool patterns, recent antibiotic use or vaccine administration or known allergy to doxycycline or related antibiotics. The complete inclusion and exclusion criteria are listed in the supplementary appendix. Participants of childbearing potential agreed to acceptable methods of contraception as outlined by the study procedures for at least 28 days prior to vaccination and until three months after receipt of the study product.

### Randomization and Masking

For the dose-expansion module, randomized assignments were generated in advance and placed into a REDCap randomization database^15^. Assignments were dispensed one by one by the Data Coordinating Centre (DCC), under the Center for Clinical Investigation at Brigham and Women’s Hospital or pharmacy staff using outlined procedures and REDCap’s dedicated randomization module. The list of assignments was generated from a custom program written by the DCC statistician in Mathematica, version 13^16^ using blocked random assignments with a hidden block size. With this system, 14 participants were randomized to 2×10^7^ CFU of PanChol, 14 participants randomized to 2×10^8^ CFU of PanChol, and eight participants randomized to placebo (matching diluent) in a double-blind fashion.

Participants, investigators, monitors, and endpoint assessors were blinded until all study visits were completed for the dose-expansion module. The pharmacists who dispensed and prepared the study product and the pharmacy monitor were not blinded. PanChol and placebo were prepared in opaque amber bottles by the pharmacy to maintain masking.

### Procedures

After written informed consent was obtained and eligibility was confirmed, participants were enrolled and admitted to the Center for Clinical Investigation at Brigham and Women’s Hospital and dosed once with PanChol or placebo. PanChol was produced under standards of good manufacturing practice at Walter Reed Army Institute of Research (Lot #2081) and stored at -80°C. Vaccine viability, monitored via CFU enumeration, remained intact over the course of the study duration. PanChol was diluted to the desired concentration in sodium bicarbonate buffer (2·5 g sodium bicarbonate [USP], 1·6 g ascorbic acid [USP], and 0·2 g lactose [NF] dissolved in 100 mL sterile water for irrigation) and all doses were confirmed by independent titration at the time of each volunteer dosing. Blood and stool samples were collected throughout the hospital period, vital signs were checked daily, and reactogenicity and unsolicited AEs were ascertained. Blood was taken for immunogenicity studies and for safety tests. Participants were administered doxycycline (initial dose 200 mg, followed by 100 mg twice a day for four days) five days after receipt of the study product to prevent potential shedding of PanChol organisms and discharged seven days after receipt of the study product if *V. cholerae* stool cultures were negative. Follow-up occurred on days 15, 29, 57, 85 and 180, with additional blood and stool collections and unsolicited AE assessments. AEs were graded in accordance with the FDA Toxicity Grading Scale for Healthy Adult and Adolescent Volunteers Enrolled in Preventive Vaccine Clinical Trials, September 2007. See supplementary appendix for details of laboratory procedures.

### Outcomes

The primary safety outcome was to estimate the incidence of solicited and unsolicited adverse events, including serious adverse events, following PanChol vaccination. The primary immunogenicity outcome was to evaluate the seroconversion (4-fold rise in titer over baseline) of vibriocidal titers to both Inaba and Ogawa *V. cholerae* at 14 days post-vaccination (day 15).

Secondary outcomes included the magnitude of pre- and post-vaccination serum vibriocidal titers to both Inaba and Ogawa *V. cholerae*, as well as direct assessment of stool shedding of PanChol organisms using both quantitative and qualitative stool cultures.

Exploratory outcomes included measurements of IgM, IgG, and IgA antibodies in serum and antibody secreting cell responses (antibodies in lymphocyte supernatant (ALS)/plasmablast) targeting Inaba- and Ogawa-specific polysaccharides (OSP), CT-B and TcpA via multiplex bead assay (Luminex), assessing the genetic stability of PanChol as well as PanChol-related changes in the fecal microbiome.

### Statistical Analysis

No formal sample size calculation was performed. Based on experience from previous studies with other cholera vaccines, the chosen cohort sizes were considered sufficient to meet the objectives of the study while minimizing unnecessary exposure. If no AEs were observed in a cohort the size of our expansion cohort (n=28), the upper one-sided 95% exact binomial confidence bound on the event rate would be approximately 10%. Primary analysis for safety analyses included all participants who received the study product. The incidence of any AE (solicited and unsolicited) was determined. Numbers of participants with AE, overall, by grade and relationship with vaccination were calculated. For both dose-escalation and dose-expansion modules, immunogenicity data were presented as the frequency and magnitude of antibody responses. Comparison of peak vibriocidal titers between participants who received PanChol doses 10^7^ or 10^8^ CFU and those who received placebo were performed using Mann-Whitney test with a two-sided 5% type I error rate (GraphPad Prism version 10).

## Results

Enrollment was conducted between Dec 13, 2022, and Feb 7, 2025. A total of 57 participants, 18-55 years old, were enrolled in the dose-escalation module (n=21) and the expansion (n=36) module (figure 1). Approximately half the cohort was male (n=27, 47%), the majority were White (n=35, 61%) and not Hispanic or Latino (n=51, 90%) (table S1). Two participants were lost to follow up for the outpatient period and their immunogenicity data were censored. Two participants withdrew early in the inpatient period and started doxycycline on day 3; stool was not collected after this point. Almost all (45/49, 92%) participants who received doses 10^6^ and above or placebo completed the six months of follow-up.

PanChol was safe and generally well-tolerated at all doses administered (figure 2; figure S1; table S2 and S3). In the dose-escalation module, in which doses of 10^4^ to 10^10^ CFU were administered, 87% (N=20) of the solicited AEs during the first five days of the inpatient period were graded as mild and 13% (N=3) graded as moderate (all diarrhea, one each in 10^5^, 10^7^ and 10^9^ dose groups, table S2). No dose-limiting toxicities were noted during dose-escalation. An increase in gastrointestinal symptoms, particularly nausea, was noted late in the inpatient period (table S2 and S3), which was attributed to doxycycline administration.

**Figure 2:**
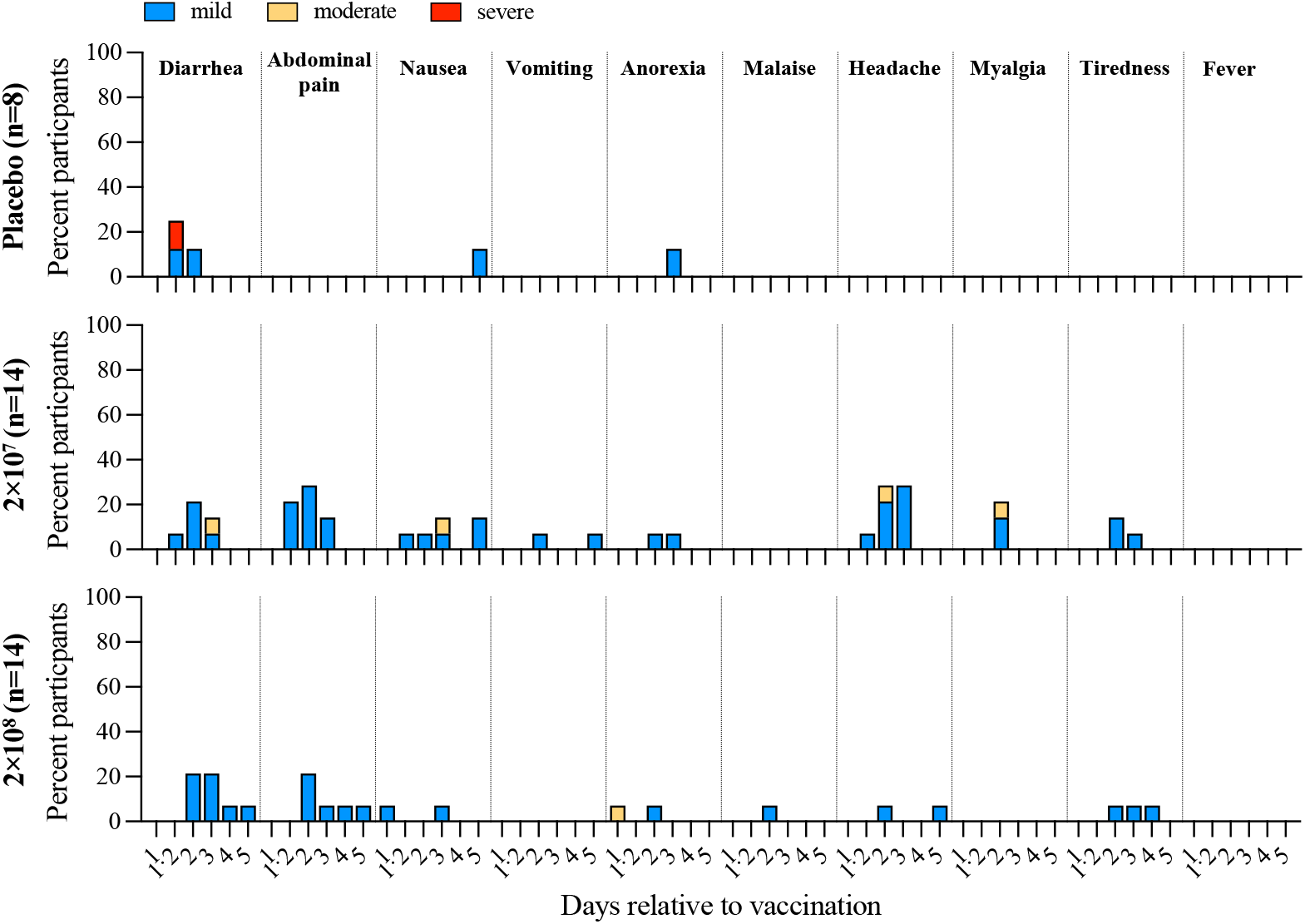
Solicited adverse events among study participants in the dose-expansion module. Shown are recorded adverse events from hospital admission (D1) until administration of doxycycline (D5) for participants dosed with placebo (top), 2×10^7^ (middle) or 2×10^8^ CFU (bottom). The Day 1·2 point represents ∼4hr post-vaccination. Two study participants started doxycycline on day three (2×10^7^: n=1; 2×10^8^: n=1). Solicited adverse events included diarrhea, abdominal pain, nausea, vomiting, anorexia, malaise, headache, myalgia, tiredness and fever. Grading of adverse events is indicated by color; blue: mild; yellow: moderate; red: severe.

No severe solicited AEs were reported in PanChol vaccinees in the placebo-controlled dose-expansion module. There was one severe solicited AE, diarrhea, in a placebo recipient observed in the hours following administration of placebo buffer. In this module, most solicited AEs (89%, 50/56) were mild with similar frequencies in recipients of the two vaccine doses (figure 2).

In total, among the 49 vaccine recipients, 31% (n=15) of participants had no solicited AEs during the first five days of the inpatient period. In the remaining vaccine recipients, there were 74 solicited AEs, 89% (N=66) of which were graded as mild with the remaining graded as moderate (figure 2; figure S1; table S2 and S3). With one exception, solicited AEs were transient and limited to 2 days.

47 participants (82%) reported 110 unsolicited AEs during the study follow-up period (table S4AB). Only one of these AEs (mild “gassy sensation”) was deemed related to the vaccine (dose-expansion module, 10^7^ group, and occurred on day 3 and resolved within a day). Of the 109 unrelated unsolicited AEs, the majority 72 (66%) were graded mild, while 33 (30%) were moderate, 2 (1·8%) were severe, and 2 (1·8%) were graded as life-threatening.

In the dose-escalation phase, 81% (17/21) of the participants had 39 unsolicited AEs. In the randomized module at least one AE occurred in 64% (9/14) at 10^7^ dose, 100% (13/13) at 10^8^ dose, and 100% (7/7) placebo recipients. There were three SAEs and four >grade 2 AEs which were: hospitalization for depression (dose-escalation module, 10^9^ group, day 29, grade 3 and SAE), increased AST (randomized module, placebo recipient, grade 3, day 11), hospitalization for depression (randomized module, 10^8^ group, day 39, grade 4 and SAE) and a fall (dose-escalation module, 10^9^, day 33, grade 4 and SAE).

Post vaccination, daily PanChol shedding was monitored by counting CFU in fecal samples until day 5 when doxycycline was given. In the dose-escalation module, PanChol bacteria were detected in the stool of all vaccinees dosed with 10^5^-10^10^ CFU. However, only 1 of 3 recipients of 10^4^ CFU shed PanChol bacteria, suggesting that the minimal colonization dose of PanChol when administered with buffer lies between 10^4^ and 10^5^ CFU.

PanChol shedding was not detected in any placebo recipient and was detected in 44/46 (96%) participants who received doses of 10^5^-10^10^ CFU (figure 3A; table S5). Two of the recipients of the highest dose, 10^10^ CFU, started excreting PanChol several hours after vaccination, likely representing direct passage of the vaccine through the intestine. At the other doses, fecal shedding generally became detectable 24 hours post vaccination, with similar numbers of PanChol CFU detected after three days (figure 3B; table S5). Lack of correlation between vaccine dose and fecal CFU suggests that if PanChol establishes a replicative niche in the intestine, the abundance of vaccine shedding is independent of dose (analogous to a vaccinia ‘take’ for the smallpox vaccine). In most vaccinees (n=37/42, 88%), shedding remained detectable throughout the inpatient period prior to doxycycline administration. The morning following the first dose of doxycycline administration all stool cultures were PanChol negative in the hospital CLIA approved, clinical microbiology laboratory, except for two samples which were culture negative later that day or the next day (one each).

**Figure 3:**
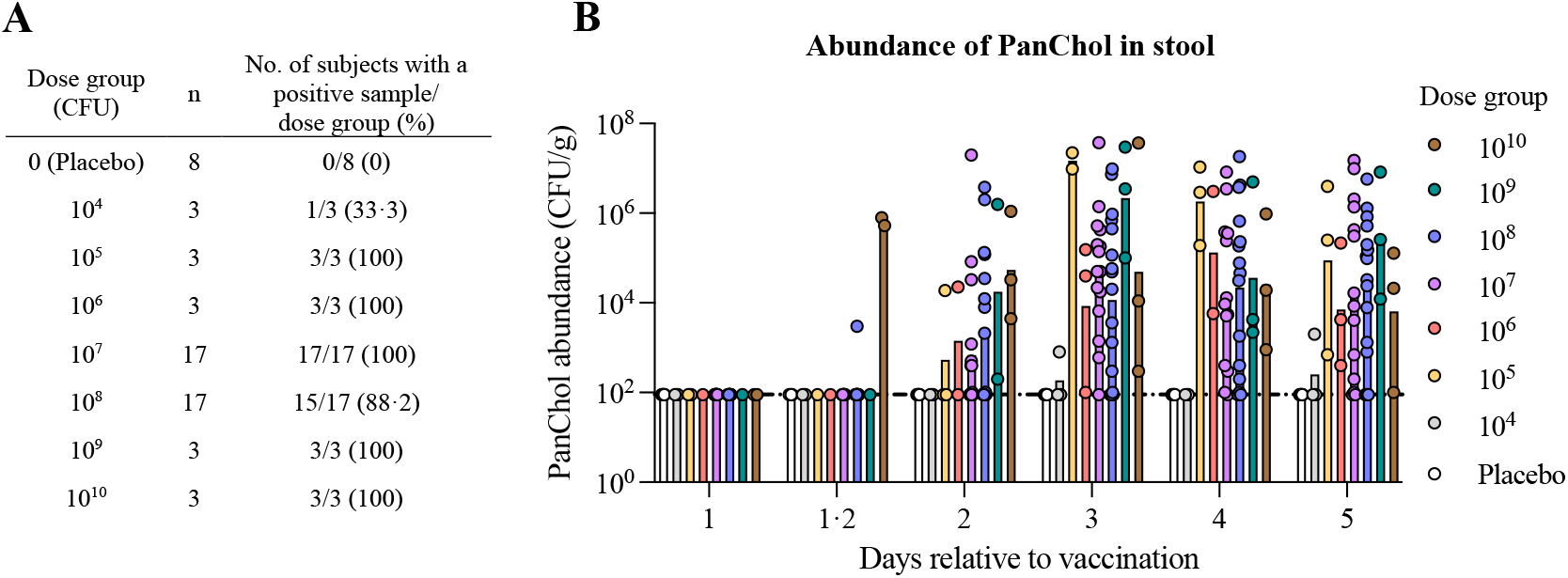
PanChol shedding in feces. A) Percentage of participants in each dose group with PanChol detected in at least one stool culture within the first four days post vaccination. B) The abundance of PanChol CFU in stool samples collected once daily during the first four days post vaccination. The colors distinguish the different doses, and the non-colored bar represents placebo recipients The day 1·2 point represents ∼4hr post-vaccination. Two study participants started doxycycline on day three (2×10^7^: n=1; 2×10^8^: n=1) and no stool specimen were collected afterwards. The limit of detection is 100 CFU/g and no shedding is presented as 90 CFU/g. Bars indicate geometric means.

Since PanChol is a bivalent vaccine expressing both the Inaba and Ogawa antigens, we evaluated whether the vaccine stably expressed both serotypes during its passage through the intestinal tract using slide agglutination assays with specific antisera targeting Inaba or Ogawa *V. cholerae*. All tested colonies derived from fecal samples (n=44/44, 100%), including through day four post vaccination, agglutinated with both antisera, reflecting the stability of the bivalent phenotype.

Whole genome sequencing was performed to probe genetic stability on samples from participants’ stool obtained three or four days following vaccination by comparison to the vaccine strain sequence. In samples prepared from five participants (10^6^ (n=1), 10^7^ (n=3) and 10^8^ (n=1) CFU), only a single nucleotide polymorphism (at position 690612) was detected in one individual, suggesting that the PanChol genome is highly stable during passage through and replication in the human gut.

Although PanChol was detectable in fecal samples from most vaccinees by plating colonies on selective media, the vaccine organisms represented a very small fraction of the fecal microbiota during the first five days post-vaccination and were only detectable in 16S rRNA analyses of fecal samples from recipients receiving 10^10^ PanChol CFU (figure S2A). Furthermore, the vaccine did not have a detectable effect on the composition of the microbiota-based analyses of 16S rRNA-sequences. Stool samples from nine vaccinees (10^6^ (n=1); 10^7^ (n=2); 10^8^ (n=3); 10^9^ (n=1); 10^10^ (n=2)) compared to three placebo recipients showed minimal changes in participants’ microbiota diversity during the inpatient period with a comparable highest dissimilarity index (Bray Curtis) (figure S2BC). Thus, PanChol intestinal colonization appears to have a negligible impact on the intestinal microbial community.

Serum vibriocidal antibody titers using isogenic Inaba and Ogawa *V. cholerae* O1 strains as targets^17^ were used to evaluate the immunogenicity of a single PanChol dose. All vaccinees administered 10^5^–10^10^ PanChol CFU developed vibriocidal antibody titers against both serotypes, including the two vaccine recipients without detectable shedding in the 10^8^ dose group, 14 days post-vaccination, whereas no response was detectable in placebo recipients (figure 4; table S6). All (n=45, 100%) vaccinees had a ≥4-fold increase (seroconversion) in vibriocidal antibody titers, comparing baseline to the individuals’ peak responses, against both serotypes regardless of baseline titers (table S6). Similar vibriocidal antibody titers were detected in all vaccinees over the six-log dose range studied, suggesting that regardless of vaccine dose, vaccine replication in the intestine allowed comparable immune responses.

**Figure 4:**
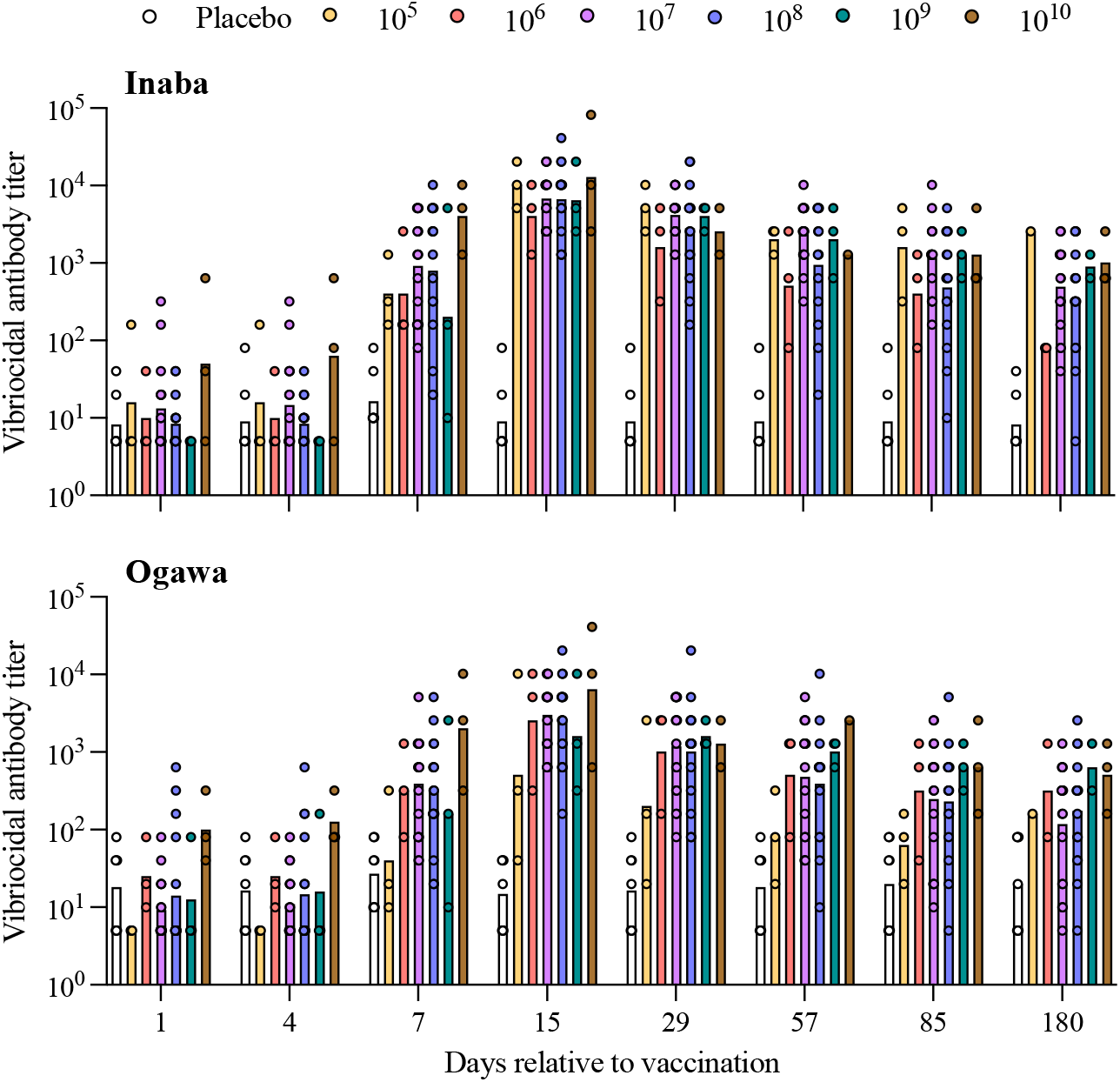
Serum vibriocidal antibody responses to Inaba and Ogawa *V. cholerae*. Temporal course of vibriocidal antibody titers starting at baseline (D1) until study end (D180) to Inaba (top) and Ogawa (bottom) *V. cholerae*. The colors distinguish the different doses, and the non-colored bar represents placebo recipients (D0-D85 n=54; D180 n=52). Bars represent geometric means.

Vaccine induced vibriocidal titers became detectable at day seven and generally peaked 14 days after vaccination (figure 4). At day 15, vibriocidal antibody responses to both serotypes were significantly induced compared to placebo recipients in vaccinees dosed with 2×10^7^ or 2×10^8^ CFU (table S7). Titers remained elevated up to at least six months post-vaccination in most participants and remained approximately 37-fold above baseline for Inaba (37·1x for the 10^7^ and 39·9x for 10^8^ dose groups, respectively) and approximately 12-fold above baseline levels for Ogawa (12·8x for the 10^7^ and 12·4x for 10^8^ dose groups, respectively) (figure 4; table S7). Overall, PanChol induced comparable levels of serotype-specific vibriocidal responses, although a trend towards higher Inaba antibody titers was observable (day 15 geometric mean titer (GMT) (95% confidence interval (CI)) for Inaba 6811 (4567-10157) and 6640 (3815-11555) in response to the 10^7^ and 10^8^ CFU dose of PanChol; day 15 GMT (95%CI) for Ogawa 3013 (1778-5107) and 2451 (1241-4842) in response to the 10^7^ and 10^8^ CFU dose of PanChol). At most doses, both the mean of the individual peak serum vibriocidal antibody response (mean peak titer) and the mean peak fold increase in titer was approximately twice as great for Inaba compared to Ogawa (table S8).

Antigen- and isotype-specific responses to PanChol in sera from vaccine and placebo recipients were monitored using paramagnetic beads coated with Inaba and Ogawa OSP, CT-B, and TcpA, the major subunit of *V. cholerae*’s chief intestinal colonization factor^18^. Biobanked serum from a previous trial including sera from participants challenged with a virulent *V. cholerae* Inaba strain (N16961) and recipients of Vaxchora, a different live-attenuated cholera vaccine^19^, were analyzed concurrently for comparison. Importantly, IgM, IgG and IgA responses to Inaba and Ogawa OSP and CT-B subunit were detected in most vaccinees and were similar to those seen in experimentally induced cholera from participants of the Vaxchora challenge study (figure 5 A-I; table S9). Serum antibody responses to TcpA were not as potently induced compared to the other antigens (figure 5 J-L). Compared to Vaxchora, which is derived from an Inaba strain of classical *V. cholerae*, PanChol, expresses both serotype antigens, induced more robust antibody responses to Ogawa-specific OSP and similar responses to Inaba OSP (figure 5 A-F).

**Figure 5:**
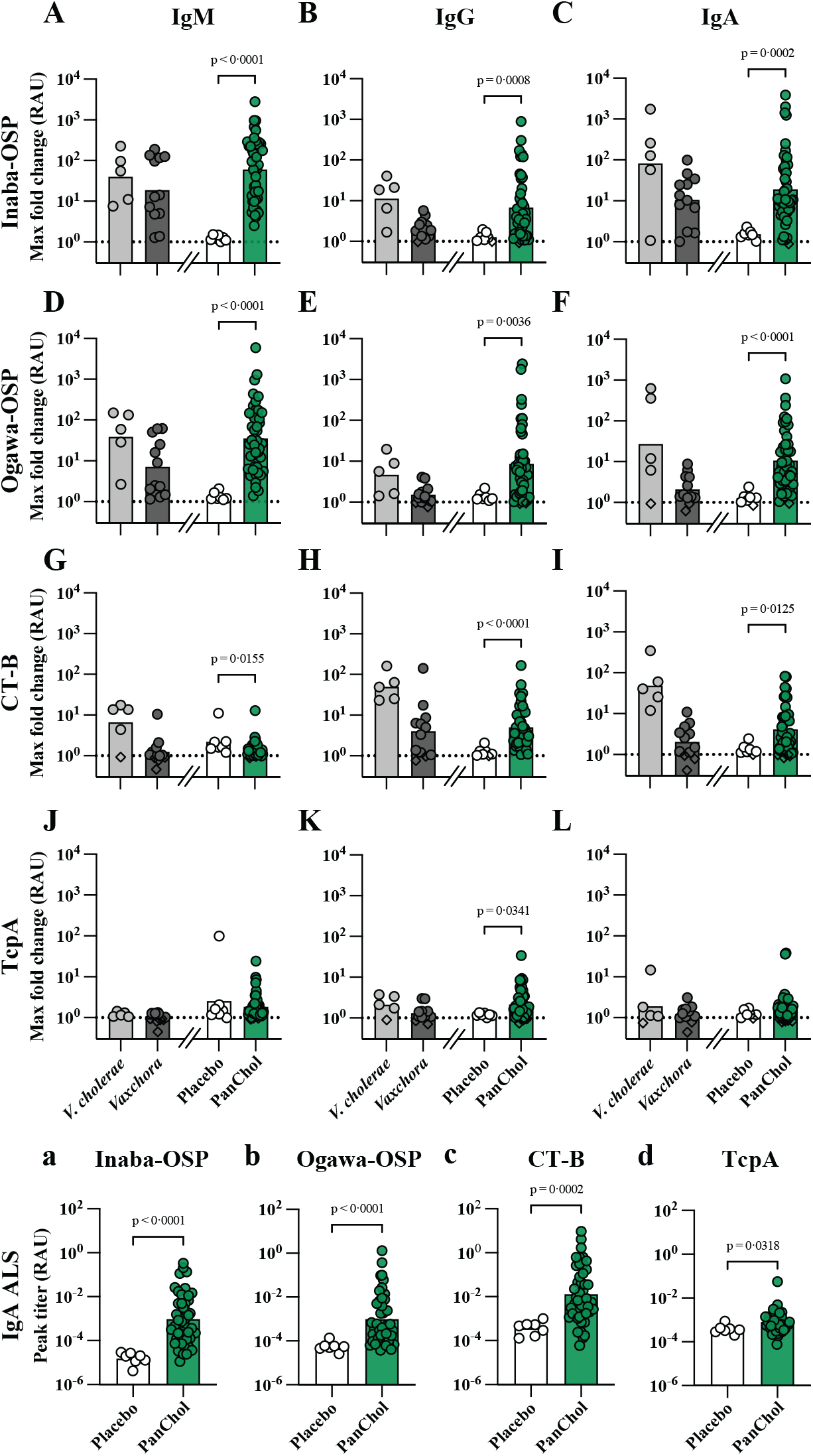
Antigen- and isotype-specific immune responses to Inaba (A, B, C, a), and Ogawa (D,E, F, b) OSP, CT-B (G, H, I, c) and TcpA (J, K, L, d) in serum (A-L) and in antibody lymphocyte supernatant (a-d). A-L) Maximum fold change of the antigen- and isotype-specific immune response to the indicated antigens. Biobanked sera from a previous trial of Vaxchora, a different live-attenuated cholera vaccine^19^ were also analyzed for peak antigen- and isotype-specific responses in A-L. The *V. cholerae* group in A-L represents data from placebo participants that received the Inaba wild-type *V. cholerae* challenge strain (N16961) in the Vaxchora trial. Values were calculated by comparing each individual’s baseline titer to their peak response. If no baseline data (day 1) were available, the next reported data (day 4 or day 7) was used for calculation. Closed circles represent an increase in the respective isotype specific immune response, diamonds represent a fold change below 1 and the dotted line indicates a fold change of 1. Statistical comparison of placebo vs PanChol was performed using a Mann-Whitney test. *V. cholerae*:5; Vaxchora:12; PanChol Placebo:7; PanChol vaccine:45. a-d) Peak IgA responses to indicated antigens in antibody lymphocyte supernatant of PanChol and placebo recipients. Values were calculated using each individual’s peak response. The PanChol group represents combined data from all vaccine recipients. Statistical comparison of placebo vs PanChol was performed using a Mann-Whitney test. Placebo:7; PanChol:45. For all graphs, bars represent geometric means, and data is presented as relative antibody units (RAU).

Antigen-specific antibodies in lymphocyte supernatant (ALS), a surrogate marker for mucosal immune responses^20^, were measured using beads coated with the aforementioned four *V. cholerae* antigens to evaluate antigen-and isotype-specific mucosal immune responses to PanChol. PanChol vaccinees had significantly induced IgA ALS responses to all four tested antigens (figure 5 a-d; table S10). IgM ALS responses were restricted to Inaba and Ogawa OSP, whereas IgG ALS responses were confined to CT-B (figure S3; table S10).

## Discussion

Cholera continues to threaten public health in many parts of the world and vaccines are increasingly recognized as important tools for cholera control^1^. Large scale clinical trials have established the utility of whole-cell killed OCVs in reducing disease but have also revealed limited efficacy in young children^21,22^. However, natural infection with *V. cholerae* generally leads to protection against subsequent disease, even in young children^5,23^. Protection from natural infection may also be more durable than that elicited by killed OCVs^5,6^. PanChol, a live vaccine engineered from a current ‘wave 3’ clinical isolate of El Tor *V. cholerae*^*11*^, was created to harness the superior immunogenicity engendered by *V. cholerae* replication in the small intestine compared to that elicited by killed OCVs. Here, we report the first-in-human study of PanChol safety and immunogenicity in healthy adults. A single oral dose of PanChol was generally well-tolerated and safe up to the highest administered dose of 10^10^ CFU. Vaccine shedding, a marker for vaccine replication in the intestine was detectable across a dose range spanning from 10^5^ to 10^10^ CFU. Notably, PanChol was immunogenic at all doses across this 100,000-fold dose range, eliciting seroconverting vibriocidal titers to both Inaba and Ogawa serotypes in every vaccinee.

PanChol was safe and generally well-tolerated in the participants in this trial and exhibited a similar reactogenicity profile as Peru-15, another El Tor-based live attenuated vaccine that underwent clinical trials in both the USA and LMIC^8,10,24,25^. This is in contrast to several early trials of different live-attenuated vaccines based on El Tor *V. cholerae*, where an unacceptable frequency of reactogenic diarrhea was observed^14^, even though these vaccines lacked *ctxA*, the enzymatic component of cholera toxin. Precise mechanistic understanding of the etiology of the reactogenicity of these vaccines has been elusive, but subsequent studies have suggested a link between *V. cholerae* motility and reactogenic diarrhea. Peru-15, a non-motile Inaba El Tor-based vaccine did not cause significant reactogenic diarrhea^8,10,24,25^. PanChol is also non-motile; all five *V. cholerae* flagellin genes were deleted from the PanChol genome because pre-clinical studies suggested that reactogenic diarrhea was dependent on flagellins, perhaps due to TLR-5-dependent inflammation^26^.

Fecal shedding of PanChol was detected in 96% (44/46) of recipients given ≥10^5^ CFU indicating that the vaccine strain has a substantial capacity to overcome human barriers that can limit *V. cholerae* infection and establish intestinal colonization. Only 1 of the 3 recipients of 10^4^ CFU dose shed the vaccine, suggesting that the infectious dose of the vaccine lies between 10^4^ and 10^5^ CFU, similar to the infectious dose for N16961, the wild-type El Tor *V. cholerae* strain that has often been used as a challenge strain for testing vaccine efficacy^27^. The high fraction of vaccinees who shed PanChol over a 100,000-fold dose range reflects the vaccine strain’s capacity for intestinal colonization and contrasts with that observed for Vaxchora, a live-attenuated vaccine created from a classical *V. cholerae* strain and licensed in the USA for travelers, which was only shed by 11% of vaccinees receiving a dose of ∼4×10^8^ CFU^7^.

In contrast to Vaxchora, PanChol was specifically engineered to be resistant to toxigenic reversion and to have genetic stability. Whole genome sequencing confirmed *in vivo* genetic stability despite intestinal replication. Furthermore, despite detectable shedding, microbiome analyses suggest minimal impact on the intestinal microbiome, indicating that the intestinal replication of the vaccine is likely not nearly as robust as wild-type *V. cholerae*^*28*^.

PanChol’s capacity for intestinal colonization may underlie its potent immunogenicity given the substantial increase in vaccine antigens delivered. PanChol led to 100% of vaccinees seroconverting vibriocidal titers to both *V. cholerae* serotypes across doses of 10^5^ to 10^10^ CFU. Such high levels of seroconversion to both serotypes have not been observed in previous trials of single-dose live attenuated OCVs or multiple-dose killed OCVs^7,8,24,29^. Geometric mean vibriocidal titers elicited by PanChol also compare favorably with those elicited by other live-attenuated vaccines tested in North Americans^7,8,24^. Although PanChol expresses both the Inaba and Ogawa antigens, there was a trend towards higher Inaba vs Ogawa vibriocidal titers, suggesting that, at least in the context of PanChol vaccination, Inaba is more immunogenic. Nevertheless, the bivalent PanChol formulation appears to have been beneficial since IgM, IgG and IgA anti-Ogawa OSP titers elicited by PanChol were greater than those detected after immunization with Vaxchora, an Inaba vaccine. There was also a tendency of PanChol to stimulate greater IgA titers targeting CT-B than Vaxchora. There is some debate about the importance of immunity to cholera toxin in protection from cholera, but large-scale clinical trials of a killed vaccine with the addition of CT-B (Dukoral) demonstrated that this non-toxic subunit of cholera enhanced short term protection from cholera as well ETEC^13^. However, infection with wild-type *V. cholerae* stimulated greater IgA titers to CT-B than PanChol potentially due to the adjuvant activity of CT-A. As observed in previous studies of immune responses in cholera patients and with other live-attenuated vaccines, serum antibody and ALS titers to TcpA, the major subunit of *V. cholerae*’s most important colonization factor, were more modestly induced by PanChol^30^. Such antibodies, which are not stimulated by killed whole cell vaccines because TCP is not expressed, may provide augmented protection against cholera^18^.

Limitations of this study include its small size and conduct at a single institution in a non-endemic setting. Several vaccines, including the oral live-attenuated rotavirus vaccines, have reduced efficacy in LMIC^31^. A critical next step in the development of PanChol is to investigate its safety and immunogenicity in a cholera endemic region. A Phase 1b study of PanChol is being planned in Lusaka, Zambia, where participants will not receive doxycycline to clear the vaccine until 14 days after vaccination. This will enable increased understanding of the duration of PanChol shedding, which was curtailed in the current study on day five, and evaluation of whether longer duration of intestinal colonization could increase immunogenicity. Future studies should address the durability of immune responses elicited by PanChol beyond six months and evaluate the safety and immunogenicity of PanChol in children, a key and vulnerable population that is essential to target for cholera control. Moreover, the capacity of PanChol to induce rapid protection even prior to stimulating adaptive immunity, as observed in experimental animals^11^, should be assessed in humans. The potential value of a second PanChol dose is currently under investigation.

A single-dose oral live-attenuated vaccine such as PanChol could become an important tool for global cholera control and close key gaps left by killed OCVs. In contrast to current killed OCVs and potential future parental subunit vaccines, PanChol, a derivative of the currently circulating wave 3 El Tor *V. cholerae*, will closely mimic natural infection and express nearly the full panoply of *V. cholerae* antigens while replicating in the small intestine. Moreover, unlike killed vaccines, which only have marginal efficacy in young children, natural infection stimulates more potent antibody and memory responses to Inaba and Ogawa OSP^32^, which is thought to be the major protective antigen for immunity to cholera. A single-dose vaccine such as PanChol could also be beneficial given the current shortage of killed OCVs and may be particularly valuable in reactive vaccination campaigns to curtail cholera outbreaks, thereby strengthening global cholera control efforts.

## Data Availability

Deidentified data, the study protocol, statistical code, statistical analysis plan, and/or informed consent form
may be shared upon request to the corresponding author.

## Contributors

MKW, LRB, WHC, and SP conceptualized the study. DRL, SRW, MS, AH, ACS, AO, ES, BW, KGD, GIB, RD, AAM, DMS, and LRB were involved in the data collection. DRL, SRW, LRB, and MKW had access to the data and verified the data. DRL, SRW, MS, and NP did the statistical analysis. DRL, MS, and DMS were involved in the laboratory assay development and conducted the laboratory analyses for the study. KAD, LAP, and AE were involved in sample preparation. SRW, MD, AH, ACS, HL, LC, EH, AH, and JBPC were involved in the clinical organization of this study. MD, SRW, LRB were involved in protocol development and CJK, MD, LRB, and MKW were involved in IND development. BP and NP were data managers. JBH, XL, BP, LRB and MKW supervised aspects of this study. All authors contributed to the data interpretation, provided critical review, and approved the original version of the manuscript. All authors had final responsibility for the decision to submit for publication.

## Data sharing

Deidentified data, the study protocol, statistical code, statistical analysis plan, and/or informed consent form may be shared upon request to the corresponding author.

## Acknowledgements

The authors wish to thank the participants for their involvement in the study. The authors wish to thank Jon A. Gothing, Kim Gaffney, Erin Curry, Virginia Dimalanta, Cynthia Mompremier, Jacquelyn Koser, Rhonda Furlonge, Virginia Cyr, Roselande Muscade, Vivian Tran, Lori Golden, Heather Rios, Caitlin Sugrue, Kevin M. Zinchuk, Stephanie Manners, Charles M. Kelly III, Emily Koleske, Augusto R. (Trey) DeLeon III, Chidiogo Nwakoby, Nicholas Morreale, Anna Piermattei, Esther Haddad, Humberto Licona, Abboud Kaok, Xi Zhang, Hannah Drackley, Caleb Lambert, DSMB members (Kathy Neuzil, Jerome Kim (Chair), Holly Janes) and RSG members (Pierre Ballard, Jeanette Hayes, Chad Porter, Laura B. Martin, John J. Mekalanos, Raphael Dolin, Mark Riddle). Thank you to the Waldor lab for their feedback on the project and comments on the manuscript.

This study is supported by Wellcome Trust (218443/Z/19/Z) to MKW and LRB. MKW is supported by NIH RO1 AI042347-29 and the Howard Hughes Medical Institute (HHMI). LRB is supported by NIAID 5-UM1-AI-069412-19 and the Harvard Clinical and Translational Science Center (1UL1TR002541-01). AH is supported by NIH T32 grant AI007061. JBH is supported by NIAID R01AI179917.

