## Supplementary Appendix for "Safety and immunogenicity of PanChol, a single-dose live-attenuated oral cholera vaccine: results from a phase 1a, double-blind, randomized, placebo-controlled trial"

‡Centre hospitalier de l'Université de Montréal, Montreal, Qc, Canada

**PanChol study group**

August Heithoff, BS; Katherine G. Daily, BA; Ruchika Dehinwal, PhD; Alexander A. Morano, PhD; Kimberly A. Dufresne, BS; Lindsey A. Parisi, BS; Aidan Eustace, BS; Bonnie Piantadosi, MPH; Xiaofang Li, MD

**Full eligibility criteria**

To be eligible to participate in this study healthy volunteers must meet the following criteria within 56 days of Study Day 1 or at the time point specified in the individual eligibility criterion listed.

Inclusion Criteria

1. Must have given written informed consent (signed and dated) and any other authorizations required by local law and be able to comply with all study requirements.
2. Healthy adults aged from 18 to 55 years old.
3. Considered healthy, as judged by the clinical investigator, according to medical history, physical examination, vital signs, screening laboratories, and medication history.
4. Capable of understanding, consenting, and complying with the entire study protocol including the inpatient period.
5. Female participants must be non-pregnant and non-lactating and either
    a. surgically sterile (history of bilateral ligation, bilateral salpingectomy, bilateral oophorectomy, total hysterectomy) or postmenopausal (defined as as amenorrhea for at least 12 consecutive months before screening without an alternative medical cause)
    b. be of child-bearing potential and practicing an acceptable method of contraception or abstaining from all activities that could result in pregnancy for at least 28 days before vaccination until 3 months after receiving the IP.

       Acceptable methods of contraception include barrier methods (such as condom, diaphragm, or cervical cap used in conjunction with spermicide), intrauterine device, hormonal contraception (that may be taken or administered by oral, intravaginal, transdermal, subdermal or IM route), vasectomized partner (the vasectomized partner should be the sole partner for that participant)

Exclusion Criteria

1. Confirmed or suspected immunosuppressive condition, as a result of a disease (e.g., primary immune deficiency, malignancy, HIV infection) or have taken any systemic immunosuppressive therapy within 6 months of enrollment.
2. Pregnant or lactating women.
3. History of gastrointestinal (GI) disorder, such as previous major GI surgery, malabsorption, or any chronic GI disorders that would interfere, according to the investigator, with the IP.
4. Acute GI or febrile illness within 7 days of enrollment.
5. Have any acute or chronic medical condition that, in the opinion of the investigator, would make vaccination unsafe or interfere with the evaluation of immune response to study vaccination.
6. History of cholera vaccination.
7. History of cholera infection.
8. Abnormal stool pattern, defined as < 3 or >21 stools per week.
9. Serious allergic reaction to PanChol or placebo component (sodium bicarbonate, lactose, ascorbic acid)
10. Use of any systemic antibiotics within 1 month of PanChol administration.
11. Receipt of a live vaccine in the previous 4 weeks or planned in the 4 weeks following enrollment.
12. Receipt of a killed or subunit (non-live) vaccine in the previous 2 weeks or planned in the 2 weeks following enrollment.
13. Individuals who do not speak English will not be enrolled into this trial. This study involves more than minimal risk and no prospect of direct benefit for participants. Additionally, a subject who did not speak English may not be able to easily communicate safety concerns in a timely fashion to the study investigators
14. Childcare workers with direct contact with children ≤ 2 years of age
15. Individuals whose occupation involves handling of food

16. Healthcare workers who have direct contact with patients who are immunodeficient, HIV-positive, or have an unstable medical condition
17. Use laxatives regularly
18. Have diarrhea within 48 hours before enrollment
19. Have a history of hypersensitivity to any of the tetracyclines
20. Have a history of hypersensitivity to streptomycin or any aminoglycoside due to the known cross-sensitivity of patients to drugs in this class.
21. Individuals who have a household member who are immunodeficient, HIV-positive, or have an unstable medical condition.

**Quantification of PanChol bacteria**

Vaccine: Every vaccine dose was independently titrated by plating on Luria-Bertani agar containing streptomycin (200 μg/mL) and X-gal (40μg/mL) to determine the number of PanChol CFU per 100 ml.

Stool sample: During the inpatient period (day 1-5), one stool sample per day per study participant was collected and serial dilutions were directly plated without enrichment on thiosulfate-citrate-bile salts-sucrose (TCBS) agar (Sigma) containing streptomycin (200 μg/mL) to determine the number of PanChol CFU per gram of stool. Collected stool samples were stored at room temperature and plated within an hour after collection. If no stool specimen was produced, a rectal swab was obtained and streaked on TCBS plates to monitor for the presence of PanChol. During the inpatient period, PanChol CFU isolated from fecal samples were tested for agglutination with specific antisera targeting Inaba or Ogawa *V. cholerae* (Difco Laboratories).

**PanChol genetic stability**

Genomic DNA was isolated from PanChol CFU from the oral suspension used for vaccination and from fecal samples obtained three- or four-days following vaccination with $10^6$ (n=1), $10^7$ (n=3) and $10^8$ (n=1) CFU using the GeneJet gDNA Isolation Kit (Thermo Fisher Scientific). Whole genome sequencing was carried out at the SeqCenter, Pittsburgh. Reads were mapped to the PanChol genome (PRJNA793890) with Snippy v4.6.0 (https://github.com/tseemann/snippy) using freebayes v1.3.6 (https://github.com/freebayes/freebayes) with a minimum read coverage of 4, minimum base quality of 13, a mapping quality of 60, and a requirement of 75% read concordance[1]. Differences in the sequences of the genomes of the ingested and shed vaccine were identified by comparing the mapping results of the ingested PanChol genome with those from the shed PanChol genome.

**16S rRNA amplicon sequence and analysis**

Stool samples were collected from participants and stored at -80 °C until use. Stool samples from participants in module 1 ($10^6$ n=1; $10^7$ n=2; $10^8$: n=3; $10^9$: n=1; $10^{10}$: n=2; placebo: n=3) were used for 16S rRNA sequencing if pre-vaccine comparison samples were available. Bacterial DNA was extracted from ~250mg of stool using DNeasy PowerLyzer PowerSoil Kit (Qiagen: 12855-50) and purified with a spin column. The V3-V4 region of 16S rRNA was PCR amplified from 12·5 ng of genomic DNA using Phusion DNA polymerase (New England Biolab: M0531S) with 341F+adaptor (5'-TCG TCG GCA GCG TCA GAT GTG TAT AAG AGA CAG CCT ACG GGN GGC WGC AG-3') and 805R+adaptor (5'-GTC TCG TGG GCT CGG AGA TGT GTA TAA GAG ACA GGA CTA CHV GGG TAT CTA ATC C-3') primers. PCR amplicons were purified using Ampure XP beads (Beckman Coulter: No: A63881) and were subjected to a 2nd PCR using Phusion DNA polymerase with the Nextera XT Index Kit v2 Set A and D primers (Illumina). Each PCR amplicon was purified using Ampure XP beads and the concentration of the purified amplicon was determined with Qubit. Paired-end reads (301 cycles each) of a 650 pM pooled amplicon mix was sequenced using a NextSeq™ 1000/2000 P1 XLEAP-SBS™ Reagent Kit (600 Cycles).

Sequence results were analyzed using Qiime2 version 2025.5[2]. Briefly, fastq files were demultiplexed and adaptor sequences were trimmed using cutadapt. Forward and reverse sequences were merged using the "v-search merge-pairs" command, and low-quality reads were filtered with the "quality-filter q-score" command with default settings. Sequences were denoised using "qiime deblur denoise-16S" function with a parameter "--p-trim-length 401", since >98% of sequences were more than 401 nt in length with quality scores of 40. Sequence read depth in all samples ranged from 64,841 to 320,337. Shannon index was calculated using the "qiime2 diversity core-metrics-phylogenetic" command with sampling-depth 64800. Bray-Curtis distance was calculated with the "qiime2 diversity beta-group-significance" command.

**Vibriocidal antibody assay**

Serum samples were collected before vaccination and on day 4, 7, 15, 29, 57, 85 and 180 days post vaccination. Quantification of serum vibriocidal antibodies using the isogenic Inaba and Ogawa ZChol *V. cholerae* strain*s* as targets was performed as described[3,4]. Briefly, the ZChol strains were grown in brain heart infusion (BHI) media (BD Difco), harvested and resuspended in sterile normal saline. A mixture of guinea pig complement (Sigma) and the respective target strains was added to a 96-well plate containing serial dilutions of heat-inactivated serum samples from vaccine and placebo recipients (1:10 starting dilution). After 60 min at 37°C, BHI was added to the wells which were then incubated at 37°C to assess the vibriocidal antibody activity. Bacterial growth was measured using the BioTek Epoch2 microplate reader at an optical density of 595 nm. The titers of vibriocidal Inaba and Ogawa antibodies were reported as the highest dilution of serum resulting in a ≥50% reduction in target optical density compared to control wells without serum. A titer of 5 was assigned when no vibriocidal antibody responses were detected. The mouse monoclonal antibody 432A.1G8.G1.H12 targeting *V. cholerae* O1 OSP[5] was used as a positive control; a ≥4-fold increase in titer relative to baseline was defined as seroconversion.

**Multiplex Bead Assay for isotype- and antigen-specific antibody responses**

Serum samples were diluted 100-fold (final test concentration of 1:1000). Samples were tested in duplicate on a FlexMap3D instrument (Luminex, Austin, Texas; xPonent software v.4.3.1). A standard curve, prepared from a pool of convalescent-phase cholera patient sera, was run on each plate and relative antibody units (RAU) were predicted from averaged NetMFI values, using 4-parameter logistic regression (GraphPad Prism, v.10.4.0). A detailed version of the protocol can be found at: dx.doi.org/10.17504/protocols.io.3byl4b1x8vo5/v1

A random selection of anonymized samples from a previously reported clinical trial (http://clinicaltrials.gov/show/NCT01895855), during which consent was obtained for secondary use in additional immunologic evaluations, were included for comparison. Samples were selected from 3 cohorts: Day 1 and 39 samples from a placebo group challenged at Day 11, Day 1 and Day 11 samples from a vaccinated group challenged at Day 11, and Day 1, Day 11, and Day 91 samples from a vaccinated group challenged at Day 91[6].

**Antibody in lymphocyte supernatant (ALS) assay**

Participant blood samples (~24 mL) were collected and mixed in a sodium citrate CPT tube (BD Vacutainer 362761). After centrifugation at 1,800 × g for 30 min the plasma was removed by aspiration. The cells were transferred to a 50 mL conical tube and washed twice in 45 mL PBS centrifugated at 300 × g for 10 min. The cells were resuspended in 10 mL of PBS, and live cells were counted using 3% Acetic acid with methylene blue (Stem Cell Technologies, 07060) and a hemocytometer. After centrifugation at 300 x g for 10 min, cells were resuspended in fetal bovine serum (FBS) at a concentration of 10 million cells/mL and were kept at 4 °C Ten million cells were centrifuged at 300 × g for 10 min and then resuspended in 1 mL of RPMI1640 medium (Gibco, 11875119) containing 10% FBS, 50 mg/mL gentamicin (Gibco, 15710-072) and cultured in a 24-well plate in 5% $CO_2$ incubator at 37 °C for 48 hours. The culture supernatant was collected, centrifuged at 300 × g for 10 min to remove cells, and stored at -80 °C until subsequent Luminex analysis, where they were analyzed at 10-fold, and when needed, 2-fold dilutions.

**Figure S1: Solicited adverse events among study participants in the dose-escalation module.** Shown are recorded adverse events from hospital admission (day 1) until administration of doxycycline (day 5) for recipients of PanChol doses from $10^4$ to $10^{10}$ CFU. The 1·2 point is ~4hr post-vaccination. Solicited adverse events included diarrhea, abdominal pain, nausea, vomiting, anorexia, malaise, headache, myalgia, tiredness and fever. Grading of adverse events is indicated by color; blue: mild; yellow: moderate.

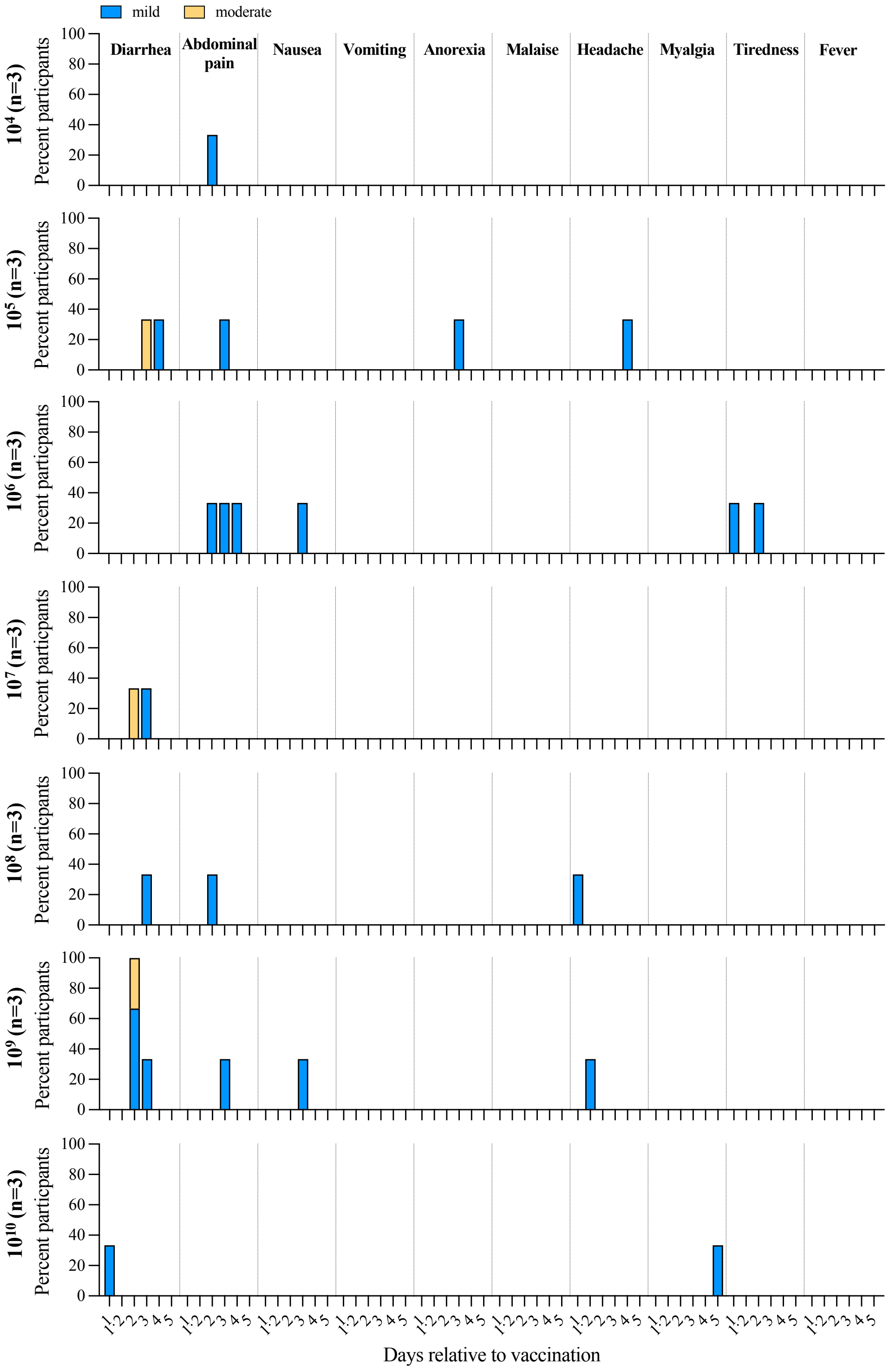

**Figure S2: Microbiome composition following PanChol ingestion.** A) Fraction of Vibrio species among total sequence reads. B) Change in the Shannon index in stool samples from individual participants; follow-up is day 180. C) Maximal Bray-Curtis dissimilarity index based on comparison of a pre-dose stool specimen to samples during the inpatient period. Data represented as mean with standard deviation. $10^6$ n=1; $10^7$ n=2; $10^8$: n=3; $10^9$: n=1; $10^{10}$: n=2; placebo n=3.

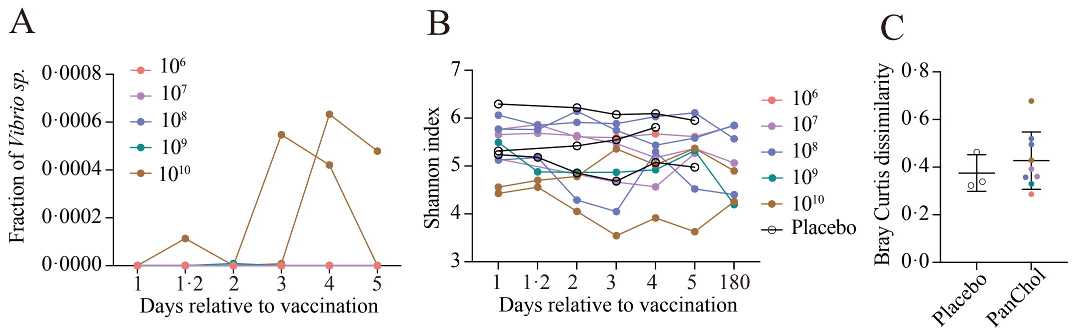

**Figure S3: Peak IgM and IgG specific immune responses to Inaba (A, B), and Ogawa (C, D) specific polysaccharides (OSP), cholera toxin B subunit, CT-B (E, F) and TcpA (G, H) in antibody lymphocyte supernatant of vaccine and placebo recipients.** PanChol group, combined data of all vaccine recipients. Statistical comparison of placebo vs PanChol was performed using a Mann-Whitney test. Placebo: ≥4; PanChol: ≥41. For all graphs, bars represent geometric means, and data is presented as relative antibody units (RAU).

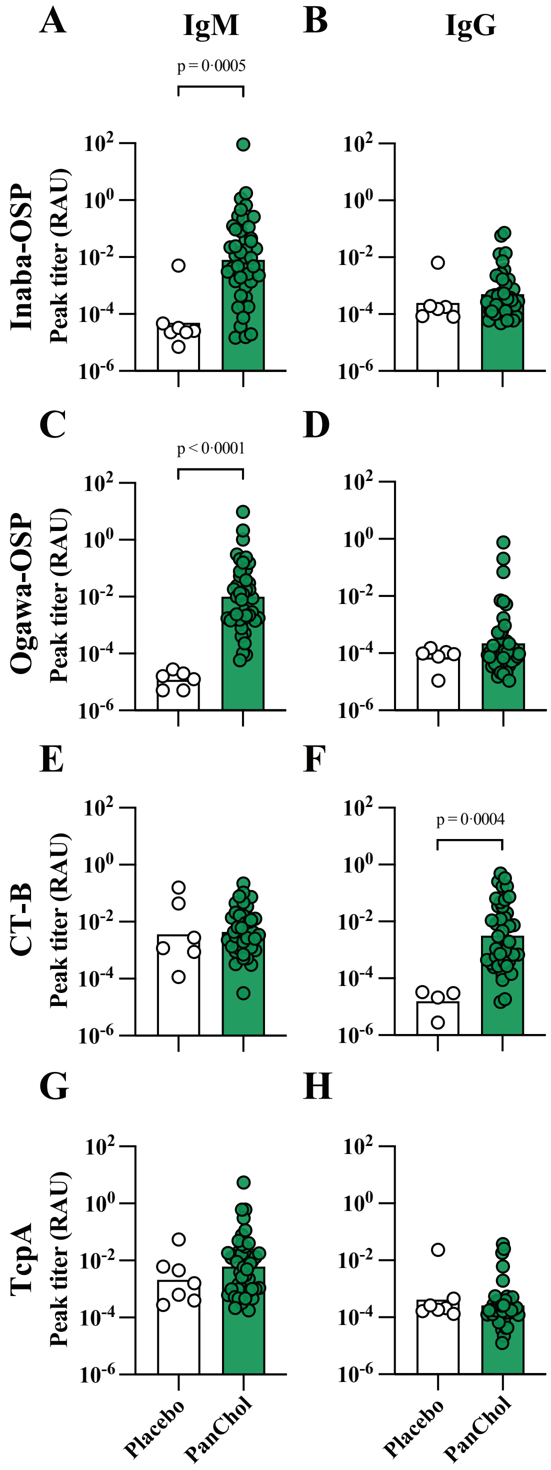

**Table S1: Baseline characteristics**

| | Total | Dose-escalation | Dose-expansion | | |
|---|---|---|---|---|---|
| | | $10^4$- $10^{10}$ | $2\times10^7$ | $2\times10^8$ | Placebo |
| **Study participants** (n) | 57 | 21 | 14 | 14 | 8 |
| **Age at consent** (median years, range) | 29 (18-55) | 37·1 (23·5 – 53·8) | 29·1 (18·4 – 54·6) | 28·1 (18·6 – 55·9) | 29·1 (20·7 – 50·7) |
| **Male Sex** (n, %) | 27 (47·3) | 11 (52·3) | 5 (35·7) | 7 (50·0) | 4 (50·0) |
| **Ethnicity** (n, %) | | | | | |
| Hispanic or Latino | 5 (8·8) | 1 (4·8) | 2 (14·3) | 1 (7·1) | 1 (12·5) |
| Not Hispanic or Latino | 51 (89·5) | 20 (95·2) | 12 (85·7) | 13 (92·9) | 6 (75·0) |
| Not Reported | 1 (1·8) | 0 (0·0) | 0 (0·0) | 0 (0·0) | 1 (12·5) |
| **Race** (n, %) | | | | | |
| Asian | 8 (14·0) | 3 (14·3) | 0 (0·0) | 4 (28·6) | 1 (12·5) |
| Black or African American | 13 (22·8) | 7 (33·3) | 2 (14·3) | 2 (14·3) | 2 (25·0) |
| White | 35 (61·4) | 11 (52·4) | 12 (85·7) | 8 (57·1) | 4 (50·0) |
| Not Reported | 1 (1·8) | 0 (0·0) | 0 (0·0) | 0 (0·0) | 1 (12·5) |
| **Gender Identity** (n, %) | | | | | |
| Male | 25 (43·9) | 10 (47·6) | 5 (35·7) | 7 (50·0) | 3 (37·5) |
| Female | 22 (38·6) | 9 (42·9) | 4 (28·6) | 6 (42·9) | 3 (37·5) |
| Transgender | 8 (14·0) | 2 (9·5) | 4 (28·6) | 1 (7·1) | 1 (12·5) |
| Other | 2 (3·5) | 0 (0·0) | 1 (7·1) | 0 (0·0) | 1 (12·5) |

**Table S2: Solicited adverse events reported from participants of the dose-escalation module during the inpatient period (day 1-7).**

| | No. of subjects with symptom/total no. of subjects in the dosing group during the inpatient period (dose-escalation module) | | | | | | |
|---|---|---|---|---|---|---|---|
| **Symptom** | **$10^4$ CFU** | **$10^5$ CFU** | **$10^6$ CFU** | **$10^7$ CFU** | **$10^8$ CFU** | **$10^9$ CFU** | **$10^{10}$ CFU** |
| Diarrhea | | | | | | | |
| Mild grade | 0/3 | 0/3 | 0/3 | 1/3 | 1/3 | 2/3 | 1/3 |
| Moderate grade | 0/3 | 1/3 | 0/3 | 1/3 | 0/3 | 1/3 | 0/3 |
| Severe grade | 0/3 | 0/3 | 0/3 | 0/3 | 0/3 | 0/3 | 0/3 |
| Abdominal pain | | | | | | | |
| Mild grade | 1/3 | 1/3 | 2/3 | 0/3 | 1/3 | 1/3 | 0/3 |
| Moderate grade | 0/3 | 0/3 | 0/3 | 0/3 | 0/3 | 0/3 | 0/3 |
| Nausea | | | | | | | |
| Mild grade | 1/3 | 0/3 | 2/3 | 1/3 | 0/3 | 1/3 | 0/3 |
| Moderate grade | 0/3 | 0/3 | 0/3 | 0/3 | 0/3 | 0/3 | 0/3 |
| Vomiting | | | | | | | |
| Mild grade | 0/3 | 0/3 | 1/3 | 1/3 | 0/3 | 0/3 | 0/3 |
| Moderate grade | 0/3 | 0/3 | 0/3 | 0/3 | 0/3 | 0/3 | 0/3 |
| Anorexia | | | | | | | |
| Mild grade | 0/3 | 1/3 | 0/3 | 0/3 | 0/3 | 0/3 | 0/3 |
| Moderate grade | 0/3 | 0/3 | 0/3 | 0/3 | 0/3 | 0/3 | 0/3 |
| Malaise | | | | | | | |
| Mild grade | 0/3 | 0/3 | 0/3 | 0/3 | 0/3 | 0/3 | 0/3 |
| Moderate grade | 0/3 | 0/3 | 0/3 | 0/3 | 0/3 | 0/3 | 0/3 |
| Headache | | | | | | | |
| Mild grade | 0/3 | 1/3 | 0/3 | 0/3 | 1/3 | 1/3 | 0/3 |
| Moderate grade | 0/3 | 0/3 | 0/3 | 0/3 | 0/3 | 0/3 | 0/3 |
| Myalgia | | | | | | | |
| Mild grade | 0/3 | 0/3 | 0/3 | 0/3 | 0/3 | 0/3 | 1/3 |
| Moderate grade | 0/3 | 0/3 | 0/3 | 0/3 | 0/3 | 0/3 | 0/3 |
| Tiredness | | | | | | | |
| Mild grade | 0/3 | 0/3 | 2/3 | 0/3 | 0/3 | 0/3 | 0/3 |
| Moderate grade | 0/3 | 0/3 | 0/3 | 0/3 | 0/3 | 0/3 | 0/3 |
| Fever | | | | | | | |
| Mild grade | 0/3 | 0/3 | 0/3 | 0/3 | 0/3 | 0/3 | 0/3 |
| Moderate grade | 0/3 | 0/3 | 0/3 | 0/3 | 0/3 | 0/3 | 0/3 |

**Table S3: Solicited adverse events reported from participants of the dose-expansion module during the inpatient period (day 1-7).**

| | No. of subjects with symptom/total no. of subjects in group during the inpatient period (dose-expansion module) | | |
|---|---|---|---|
| **Symptom** | **Placebo** | **$2\times10^7$ CFU** | **$2\times10^8$ CFU** |
| Diarrhea | | | |
| Mild grade | 2/8 | 4/14 | 6/14 |
| Moderate grade | 0/8 | 1/14 | 0/14 |
| Severe grade | 1/8 | 0/14 | 0/14 |
| Abdominal pain | | | |
| Mild grade | 0/8 | 5/14 | 5/14 |
| Moderate grade | 0/8 | 0/14 | 0/14 |
| Nausea | | | |
| Mild grade | 2/8 | 7/14 | 4/14 |
| Moderate grade | 0/8 | 1/14 | 0/14 |
| Vomiting | | | |
| Mild grade | 1/8 | 2/14 | 0/14 |
| Moderate grade | 0/8 | 0/14 | 0/14 |
| Anorexia | | | |
| Mild grade | 1/8 | 2/14 | 1/14 |
| Moderate grade | 0/8 | 0/14 | 1/14 |
| Malaise | | | |
| Mild grade | 0/8 | 0/14 | 2/14 |
| Moderate grade | 0/8 | 0/14 | 0/14 |
| Headache | | | |
| Mild grade | 0/8 | 6/14 | 2/14 |
| Moderate grade | 0/8 | 2/14 | 0/14 |
| Myalgia | | | |
| Mild grade | 0/8 | 2/14 | 0/14 |
| Moderate grade | 0/8 | 1/14 | 0/14 |
| Tiredness | | | |
| Mild grade | 0/8 | 2/14 | 3/14 |
| Moderate grade | 0/8 | 0/14 | 0/14 |
| Fever | | | |
| Mild grade | 0/8 | 0/14 | 0/14 |
| Moderate grade | 0/8 | 0/14 | 0/14 |

**Table S4A: Unsolicited adverse events greater than grade 2**

| Pub-ID | Dose | Module | Verbatim Term | Days since vaccination | Maximum Severity Grade[1] | Relatedness |
|---|---|---|---|---|---|---|
| 100-08 | $10^9$ | 1[2] | Fall | 33 | 4 | Not related |
| 100-12 | $10^9$ | 1 | Depression - SAE | 27 | 3 | Not related |
| 100-44 | $2 \times 10^8$ | 3[3] | Major Depression Episode - SAE | 39 | 4 | Not related |
| 100-40 | Placebo | 3 | Increased AST | 11 | 3 | Not related |

[1]Graded according to the FDA Toxicity Grading Scale for Healthy Adult and Adolescent Volunteers Enrolled in Preventive Vaccine Clinical Trials (September 2007); [2]dose-escalation module; [3]dose-expansion module

### Table S4B: Unsolicited adverse events grade 2 and below

| Pub-ID | Dose | Module | Verbatim Term | Days since vaccination | Maximum Severity Grade[1] | Relatedness |
|---|---|---|---|---|---|---|
| 100-01 | $10^6$ | 1[2] | Tachypnea | 2 | 1 | Not related |
| 100-01 | $10^6$ | 1 | Nausea and Vomiting | 5 | 1 | Not related |
| 100-01 | $10^6$ | 1 | Menstrual Cramps | 3 | 1 | Not related |
| 100-02 | $10^6$ | 1 | Tachypnea | 2 | 1 | Not related |
| 100-02 | $10^6$ | 1 | Elevated Bilirubin | 3 | 1 | Not related |
| 100-02 | $10^6$ | 1 | Chills | 0 | 1 | Not related |
| 100-02 | $10^6$ | 1 | Sprained ankle - Right | 130 | 2 | Not related |
| 100-03 | $10^6$ | 1 | Tachypnea | 2 | 1 | Not related |
| 100-04 | $10^7$ | 1 | Diarrhea | 0 | 2 | Not related |
| 100-04 | $10^7$ | 1 | Tachypnea | 2 | 1 | Not related |
| 100-04 | $10^7$ | 1 | Tachypnea | 5 | 1 | Not related |
| 100-05 | $10^8$ | 1 | Tachypnea | 1 | 1 | Not related |
| 100-06 | $10^7$ | 1 | Nausea and Vomiting | 5 | 1 | Not related |
| 100-06 | $10^7$ | 1 | Elevated Absolute Eosinophil Count | 6 | 1 | Not related |
| 100-07 | $10^7$ | 1 | Increased ALT | 6 | 2 | Not related |
| 100-09 | $10^8$ | 1 | Thyroid Nodule | 43 | 1 | Not related |
| 100-10 | $10^8$ | 1 | Systolic Hypertension | 0 | 2 | Not related |
| 100-10 | $10^8$ | 1 | Systolic Hypertension | 4 | 2 | Not related |
| 100-11 | $10^9$ | 1 | Systolic Hypertension | 2 | 1 | Not related |
| 100-11 | $10^9$ | 1 | Elevated ALT | 6 | 1 | Not related |
| 100-11 | $10^9$ | 1 | Elevated AST | 6 | 1 | Not related |
| 100-13 | $10^{10}$ | 1 | Decreased Absolute Neutrophil Count | 3 | 1 | Not related |
| 100-13 | $10^{10}$ | 1 | Decreased White Blood Cell Count | 3 | 1 | Not related |
| 100-19 | $2 \times 10^8$ | 3[3] | Tachypnea | 1 | 1 | Not related |
| 100-19 | $2 \times 10^8$ | 3 | Increased Total Bilirubin | 1 | 1 | Not related |

| Pub-ID | Dose | Module | Verbatim Term | Days since vaccination | Maximum Severity Grade[1] | Relatedness |
|---|---|---|---|---|---|---|
| 100-19 | $2 \times 10^8$ | 3 | Decreased Absolute Neutrophil Count | 3 | 2 | Not related |
| 100-19 | $2 \times 10^8$ | 3 | Decreased Heart Rate | 4 | 2 | Not related |
| 100-20 | $2 \times 10^7$ | 3 | Decreased Absolute Neutrophil Count | 3 | 1 | Not related |
| 100-20 | $2 \times 10^7$ | 3 | Upper Respiratory Infection | 66 | 2 | Not related |
| 100-21 | $2 \times 10^7$ | 3 | Sore throat | 1 | 2 | Not related |
| 100-21 | $2 \times 10^7$ | 3 | Headache | 1 | 1 | Not related |
| 100-21 | $2 \times 10^7$ | 3 | Dehydration | 42 | 1 | Not related |
| 100-22 | $2 \times 10^8$ | 3 | Tachypnea | 1 | 1 | Not related |
| 100-22 | $2 \times 10^8$ | 3 | Tachypnea | 6 | 1 | Not related |
| 100-23 | $2 \times 10^7$ | 3 | Tachypnea | 1 | 1 | Not related |
| 100-23 | $2 \times 10^7$ | 3 | Elevated ALT | 6 | 1 | Not related |
| 100-23 | $2 \times 10^7$ | 3 | Increased Total Bilirubin | 29 | 1 | Not related |
| 100-24 | Placebo | 3 | Tachypnea | 0 | 2 | Not related |
| 100-24 | Placebo | 3 | Diastolic Hypertension | 1 | 1 | Not related |
| 100-24 | Placebo | 3 | Decreased Hemoglobin | 14 | 1 | Not related |
| 100-25 | $2 \times 10^8$ | 3 | Headache | 1 | 1 | Not related |
| 100-26 | Placebo | 3 | Decreased Hemoglobin | 22 | 1 | Not related |
| 100-27 | Placebo | 3 | Decreased Hemoglobin | 16 | 2 | Not related |
| 100-28 | $2 \times 10^8$ | 3 | Tachypnea | 2 | 1 | Not related |
| 100-28 | $2 \times 10^8$ | 3 | Increased White Blood Cells | 1 | 1 | Not related |
| 100-28 | $2 \times 10^8$ | 3 | Upper Respiratory Infection | 13 | 2 | Not related |
| 100-29 | $2 \times 10^8$ | 3 | Decreased Absolute Neutrophil Count | 3 | 1 | Not related |
| 100-30 | Placebo | 3 | Tachypnea | 2 | 1 | Not related |
| 100-30 | Placebo | 3 | Elevated White Blood Cell Count | 6 | 1 | Not related |
| 100-31 | $2 \times 10^7$ | 3 | Gassy abdominal sensation | 3 | 1 | Related |

| Pub-ID | Dose | Module | Verbatim Term | Days since vaccination | Maximum Severity Grade[1] | Relatedness |
|---|---|---|---|---|---|---|
| 100-31 | $2 \times 10^7$ | 3 | Tachypnea | 1 | 1 | Not related |
| 100-31 | $2 \times 10^7$ | 3 | Decreased WBC | 29 | 1 | Not related |
| 100-31 | $2 \times 10^7$ | 3 | Decreased Neutrophils | 29 | 2 | Not related |
| 100-31 | $2 \times 10^7$ | 3 | Decreased Absolute Neutrophil Count | 29 | 2 | Not related |
| 100-32 | $2 \times 10^8$ | 3 | Lowered Hemoglobin | 0 | 2 | Not related |
| 100-32 | $2 \times 10^8$ | 3 | Lowered Hemoglobin | 31 | 2 | Not related |
| 100-33 | $2 \times 10^8$ | 3 | Decreased Absolute Neutrophil Count | 1 | 2 | Not related |
| 100-33 | $2 \times 10^8$ | 3 | Decreased White Blood Cells | 3 | 1 | Not related |
| 100-33 | $2 \times 10^8$ | 3 | Decreased White Blood Cells | 27 | 1 | Not related |
| 100-34 | $2 \times 10^8$ | 3 | Tachypnea | 0 | 1 | Not related |
| 100-34 | $2 \times 10^8$ | 3 | Orthostatic tachycardia | 2 | 1 | Not related |
| 100-34 | $2 \times 10^8$ | 3 | Increased Total Bilirubin | 1 | 2 | Not related |
| 100-34 | $2 \times 10^8$ | 3 | Lowered Absolute Neutrophil Count | 11 | 1 | Not related |
| 100-34 | $2 \times 10^8$ | 3 | Kidney Infection | 28 | 2 | Not related |
| 100-35 | Placebo | 3 | Tachypnea | 2 | 1 | Not related |
| 100-35 | Placebo | 3 | COVID-19 Infection | 18 | 2 | Not related |
| 100-35 | Placebo | 3 | Pneumonia | 115 | 2 | Not related |
| 100-36 | Placebo | 3 | Tachypnea | 2 | 1 | Not related |
| 100-37 | $2 \times 10^7$ | 3 | Systolic hypertension | 5 | 1 | Not related |
| 100-37 | $2 \times 10^7$ | 3 | Tachypnea | 1 | 1 | Not related |
| 100-38 | $2 \times 10^7$ | 3 | Increased White Blood Cells | 1 | 1 | Not related |
| 100-38 | $2 \times 10^7$ | 3 | Increased Sodium | 1 | 2 | Not related |
| 100-38 | $2 \times 10^7$ | 3 | COVID infection | 36 | 1 | Not related |
| 100-38 | $2 \times 10^7$ | 3 | Hemorrhoids | 122 | 1 | Not related |

| Pub-ID | Dose | Module | Verbatim Term | Days since vaccination | Maximum Severity Grade[1] | Relatedness |
|---|---|---|---|---|---|---|
| 100-39 | $2 \times 10^7$ | 3 | Right great toe abrasion | 6 | 1 | Not related |
| 100-40 | Placebo | 3 | Tachypnea | 0 | 1 | Not related |
| 100-40 | Placebo | 3 | Upper Respiratory Infection | 7 | 1 | Not related |
| 100-40 | Placebo | 3 | Increased ALT | 11 | 1 | Not related |
| 100-41 | $2 \times 10^8$ | 3 | Diastolic hypertension | 2 | 2 | Not related |
| 100-41 | $2 \times 10^8$ | 3 | Increased ALT | 3 | 1 | Not related |
| 100-41 | $2 \times 10^8$ | 3 | Increased AST | 3 | 1 | Not related |
| 100-42 | $2 \times 10^8$ | 3 | Decreased hemoglobin | 13 | 1 | Not related |
| 100-42 | $2 \times 10^8$ | 3 | Hyperkalemia | 26 | 2 | Not related |
| 100-43 | $2 \times 10^8$ | 3 | Urticaria | 44 | 2 | Not related |
| 100-14 | $10^4$ | 1 | Decreased Absolute Neutrophil Count | 1 | 1 | Not related |
| 100-14 | $10^4$ | 1 | Tachypnea | 0 | 1 | Not related |
| 100-45 | $2 \times 10^7$ | 3 | External Hemorrhoids | 2 | 2 | Not related |
| 100-45 | $2 \times 10^7$ | 3 | Headaches | 6 | 2 | Not related |
| 100-45 | $2 \times 10^7$ | 3 | Upper Respiratory Illness | 21 | 2 | Not related |
| 100-45 | $2 \times 10^7$ | 3 | Upper respiratory illness | 142 | 1 | Not related |
| 100-46 | $2 \times 10^7$ | 3 | Tachycardia | 1 | 1 | Not related |
| 100-46 | $2 \times 10^7$ | 3 | Elevated White Blood Cells | 1 | 2 | Not related |
| 100-15 | $10^5$ | 1 | Decreased Hemoglobin | 16 | 1 | Not related |
| 100-47 | $2 \times 10^8$ | 3 | Chills | 2 | 1 | Not related |
| 100-47 | $2 \times 10^8$ | 3 | Gassy GI sensation | 4 | 1 | Not related |

| Pub-ID | Dose | Module | Verbatim Term | Days since vaccination | Maximum Severity Grade[1] | Relatedness |
|---|---|---|---|---|---|---|
| 100-16 | $10^5$ | 1 | Lowered Absolute Lymphocytes | 3 | 2 | Not related |
| 100-16 | $10^5$ | 1 | Pruritus of the extremities | 46 | 1 | Not related |
| 100-17 | $10^5$ | 1 | Viral Upper Respiratory Tract Syndrome | 2 | 2 | Not related |
| 100-17 | $10^5$ | 1 | Tachypnea | 2 | 1 | Not related |
| 100-17 | $10^5$ | 1 | Elevated AST | 14 | 1 | Not related |
| 100-17 | $10^5$ | 1 | Decreased Absolute Neutrophil Count | 28 | 1 | Not related |
| 100-18 | $10^4$ | 1 | Tachypnea | 1 | 1 | Not related |
| 100-18 | $10^4$ | 1 | Decreased Hemoglobin | 0 | 2 | Not related |
| 100-18 | $10^4$ | 1 | Decreased Lymphocytes | 1 | 2 | Not related |
| 100-18 | $10^4$ | 1 | Leukopenia | 26 | 1 | Not related |
| 100-18 | $10^4$ | 1 | Decreased Absolute Neutrophil Count | 26 | 2 | Not related |

[1]Graded according to the FDA Toxicity Grading Scale for Healthy Adult and Adolescent Volunteers Enrolled in Preventive Vaccine Clinical Trials (September 2007); [2]dose-escalation module; [3]dose-expansion module

**Table S5: Quantification of PanChol bacteria in fecal samples derived from vaccine and placebo recipients.**
Dark blue: no sample; light blue: negative rectal swab; orange: positive rectal swab.

**Dose-Escalation Module**

| | Abundace of PanChol (CFU/g) | | | | | | |
|---|---|---|---|---|---|---|---|
| **Dose (CFU)** | **1** | **1·2** | **2** | **3** | **4** | **5** | **Notes** |
| $10^4$ | 9·00E+01 | 9·00E+01 | 9·00E+01 | 8·00E+02 | 9·00E+01 | 2·00E+03 | |
| $10^4$ | 9·00E+01 | 9·00E+01 | 9·00E+01 | 9·00E+01 | 9·00E+01 | 9·00E+01 | |
| $10^4$ | 9·00E+01 | 9·00E+01 | 9·00E+01 | 9·00E+01 | 9·00E+01 | 9·00E+01 | |
| $10^5$ | 9·00E+01 | | 9·00E+01 | | 1·07E+07 | 2·50E+05 | |
| $10^5$ | 9·00E+01 | 9·00E+01 | 9·00E+01 | 9·64E+06 | 1·90E+05 | 7·00E+02 | |
| $10^5$ | 9·00E+01 | | 1·90E+04 | 2·24E+07 | 2·90E+06 | 3·99E+06 | |
| $10^6$ | | | | 4·00E+04 | | 2·17E+05 | |
| $10^6$ | 9·00E+01 | 9·00E+01 | 9·00E+01 | 1·00E+02 | 5·80E+03 | 4·00E+02 | |
| $10^6$ | | | 2·25E+04 | 1·54E+05 | 3·08E+06 | 4·20E+03 | |
| $10^7$ | | | 9·00E+01 | 9·00E+01 | 3·80E+05 | | |
| $10^7$ | 9·00E+01 | 9·00E+01 | 9·00E+01 | 4·30E+05 | 8·20E+06 | 1·50E+07 | |
| $10^7$ | 9·00E+01 | | | 1·76E+04 | 1·30E+04 | 7·00E+02 | |
| $10^8$ | 9·00E+01 | 9·00E+01 | 1·00E+02 | 3·00E+02 | 4·00E+02 | 3·30E+04 | |
| $10^8$ | 9·00E+01 | 9·00E+01 | 3·50E+04 | 4·90E+04 | 1·80E+03 | 2·41E+04 | |
| $10^8$ | 9·00E+01 | 9·00E+01 | 1·20E+05 | 3·60E+03 | 1·88E+05 | 1·30E+06 | |
| $10^9$ | 9·00E+01 | 9·00E+01 | 1·56E+06 | 1·00E+05 | 4·20E+03 | 1·21E+04 | |
| $10^9$ | | | 2·00E+02 | 3·49E+06 | 5·00E+06 | 2·60E+05 | |
| $10^9$ | | | | 3·00E+07 | 2·20E+03 | 8·30E+06 | |
| $10^{10}$ | 9·00E+01 | 7·90E+05 | 4·40E+03 | 3·00E+02 | 9·00E+02 | 1·00E+02 | |
| $10^{10}$ | 9·00E+01 | 5·30E+05 | 3·30E+04 | 3·67E+07 | 1·93E+04 | 1·29E+05 | |
| $10^{10}$ | | | 1·10E+06 | 1·10E+04 | 9·70E+05 | 2·10E+04 | |

**Dose- Expansion Module**

| | Abundace of PanChol (CFU/g) | | | | | | |
|---|---|---|---|---|---|---|---|
| **Dose (CFU)** | **1** | **1·2** | **2** | **3** | **4** | **5** | **Notes** |
| 0 | 9·00E+01 | | 9·00E+01 | 9·00E+01 | | 9·00E+01 | |
| 0 | 9·00E+01 | | 9·00E+01 | 9·00E+01 | 9·00E+01 | 9·00E+01 | |
| 0 | 9·00E+01 | | | | | | |
| 0 | 9·00E+01 | 9·00E+01 | 9·00E+01 | 9·00E+01 | 9·00E+01 | 9·00E+01 | |
| 0 | | 9·00E+01 | 9·00E+01 | 9·00E+01 | 9·00E+01 | 9·00E+01 | |
| 0 | 9·00E+01 | 9·00E+01 | 9·00E+01 | 9·00E+01 | 9·00E+01 | 9·00E+01 | |
| 0 | 9·00E+01 | | 9·00E+01 | 9·00E+01 | 9·00E+01 | 9·00E+01 | |
| 0 | 9·00E+01 | 9·00E+01 | 9·00E+01 | 9·00E+01 | 9·00E+01 | 9·00E+01 | |
| $2\times10^7$ | 9·00E+01 | 9·00E+01 | 4·00E+02 | 1·40E+03 | 3·00E+02 | 9·00E+01 | |
| $2\times10^7$ | 9·00E+01 | 9·00E+01 | 9·00E+01 | 5·00E+04 | 2·42E+05 | 4·30E+05 | |
| $2\times10^7$ | | | 9·00E+01 | 6·00E+02 | | | |
| $2\times10^7$ | 9·00E+01 | | 5·00E+02 | 5·20E+05 | 4·00E+02 | 1·66E+04 | |
| $2\times10^7$ | 9·00E+01 | 9·00E+01 | 4·00E+02 | 1·80E+05 | 9·30E+03 | 4·10E+03 | |
| $2\times10^7$ | 9·00E+01 | | 9·00E+01 | 5·00E+04 | 3·60E+05 | 9·80E+06 | |
| $2\times10^7$ | 9·00E+01 | 9·00E+01 | 1·97E+07 | 2·00E+05 | | | early discharge |
| $2\times10^7$ | 9·00E+01 | 9·00E+01 | 1·00E+02 | 6·60E+03 | 9·00E+01 | 2·08E+06 | |
| $2\times10^7$ | 9·00E+01 | | 1·20E+03 | 3·76E+07 | 3·50E+06 | 8·50E+03 | |
| $2\times10^7$ | 9·00E+01 | 9·00E+01 | 3·30E+04 | 2·20E+04 | 5·50E+03 | 3·10E+05 | |
| $2\times10^7$ | 9·00E+01 | | 8·30E+04 | 1·40E+05 | | 1·37E+06 | |
| $2\times10^7$ | 9·00E+01 | | 9·00E+01 | | | 1·00E+02 | |
| $2\times10^7$ | 9·00E+01 | 9·00E+01 | 9·00E+01 | 1·40E+06 | 5·10E+03 | 2·00E+02 | |
| $2\times10^7$ | 9·00E+01 | 9·00E+01 | 9·00E+01 | | 1·00E+02 | 9·00E+01 | |
| $2\times10^8$ | 9·00E+01 | 9·00E+01 | 9·00E+01 | 9·00E+01 | 9·00E+01 | 9·00E+01 | |
| $2\times10^8$ | | | 9·00E+01 | 2·00E+04 | 4·60E+04 | 1·10E+05 | |
| $2\times10^8$ | 9·00E+01 | | 1·33E+05 | 1·16E+05 | 7·80E+04 | 9·70E+04 | |
| $2\times10^8$ | | 9·00E+01 | | 9·00E+01 | | 1·50E+05 | |
| $2\times10^8$ | | 9·00E+01 | 8·00E+03 | 9·00E+01 | 1·00E+02 | 8·00E+02 | |
| $2\times10^8$ | 9·00E+01 | 3·00E+03 | 3·76E+06 | 7·00E+05 | 3·12E+04 | 8·30E+05 | |
| $2\times10^8$ | 9·00E+01 | | 9·00E+01 | 4·50E+05 | 1·82E+07 | 5·20E+05 | |
| $2\times10^8$ | 9·00E+01 | 9·00E+01 | 2·01E+06 | 7·40E+06 | 4·20E+06 | 5·80E+06 | |
| $2\times10^8$ | 9·00E+01 | 9·00E+01 | 1·00E+02 | 9·50E+05 | 3·80E+06 | 2·00E+05 | |
| $2\times10^8$ | 9·00E+01 | 9·00E+01 | 9·00E+01 | 9·00E+01 | 9·00E+01 | 9·00E+01 | |
| $2\times10^8$ | 9·00E+01 | 9·00E+01 | 2·10E+03 | 5·80E+04 | 2·00E+02 | 9·00E+01 | |
| $2\times10^8$ | 9·00E+01 | 9·00E+01 | 9·00E+01 | 1·30E+03 | 2·30E+05 | 1·30E+03 | |
| $2\times10^8$ | 9·00E+01 | 9·00E+01 | 9·00E+01 | 9·70E+06 | | | early discharge |
| $2\times10^8$ | 9·00E+01 | 9·00E+01 | 1·23E+04 | 1·00E+02 | 6·60E+05 | 7·80E+03 | |

no sample
negative rectal swab
positive rectal swab

Limit of detection: 100 CFU/g
No shedding is presented as 90CFU/g.

**Table S6: Serum vibriocidal antibody responses (VAT) to Inaba and Ogawa *V. cholerae* O1.**
Dark blue: no serum sample.

**Dose-Escalation Module**

| Inaba (VAT) | | | | | | | | | |
|---|---|---|---|---|---|---|---|---|---|
| Dose (CFU) | D1 | D4 | D7 | D15 | D29 | D57 | D85 | D180 | max |
| $10^5$ | 160 | 160 | 320 | 10240 | 5120 | 2560 | 5120 | 2560 | 10240 |
| $10^5$ | 5 | 5 | 160 | 5120 | 2560 | 2560 | 2560 | | 5120 |
| $10^5$ | 5 | 5 | 1280 | 20480 | 10240 | 1280 | 320 | | 20480 |
| $10^6$ | 40 | 40 | 2560 | 10240 | 5120 | 2560 | 1280 | | 10240 |
| $10^6$ | 5 | 5 | 160 | 1280 | 320 | 80 | 80 | 80 | 1280 |
| $10^6$ | 5 | 5 | 160 | 5120 | 2560 | 640 | 640 | 80 | 5120 |
| $10^7$ | 5 | 5 | 160 | 2560 | 5120 | 5120 | 2560 | 1280 | 5120 |
| $10^7$ | 5 | 5 | 320 | 5120 | 2560 | 2560 | 320 | 160 | 5120 |
| $10^7$ | 5 | 5 | 160 | 5120 | 2560 | 640 | 320 | 40 | 5120 |
| $10^8$ | 5 | 5 | 20 | 1280 | 640 | 80 | 40 | 40 | 1280 |
| $10^8$ | 20 | 10 | 2560 | 10240 | 5120 | 1280 | 640 | 320 | 10240 |
| $10^8$ | 40 | 20 | 2560 | 10240 | 5120 | 1280 | 640 | 640 | 10240 |
| $10^9$ | 5 | 5 | 5120 | 20480 | 5120 | 5120 | 1280 | 1280 | 20480 |
| $10^9$ | 5 | 5 | 10 | 2560 | 5120 | 2560 | 2560 | | 5120 |
| $10^9$ | 5 | 5 | 160 | 5120 | 2560 | 640 | 640 | 640 | 5120 |
| $10^{10}$ | 5 | 5 | 1280 | 2560 | 1280 | | 640 | 640 | 2560 |
| $10^{10}$ | 640 | 640 | 10240 | 81920 | | | 5120 | 2560 | 81920 |
| $10^{10}$ | 40 | 80 | 5120 | 10240 | 5120 | 1280 | 640 | 640 | 10240 |

| Fold change to baseline Inaba | | | | | | | | | |
|---|---|---|---|---|---|---|---|---|---|
| Dose (CFU) | D1 | D4 | D7 | D15 | D29 | D57 | D85 | D180 | max FC |
| $10^5$ | 1 | 1 | 2 | 64 | 32 | 16 | 32 | 16 | 64 |
| $10^5$ | 1 | 1 | 32 | 1024 | 512 | 512 | 512 | | 1024 |
| $10^5$ | 1 | 1 | 256 | 4096 | 2048 | 256 | 64 | | 4096 |
| $10^6$ | 1 | 1 | 64 | 256 | 128 | 64 | 32 | | 256 |
| $10^6$ | 1 | 1 | 32 | 256 | 64 | 16 | 16 | 16 | 256 |
| $10^6$ | 1 | 1 | 32 | 1024 | 512 | 128 | 128 | 16 | 1024 |
| $10^7$ | 1 | 1 | 32 | 512 | 1024 | 1024 | 512 | 256 | 1024 |
| $10^7$ | 1 | 1 | 64 | 1024 | 512 | 512 | 64 | 32 | 1024 |
| $10^7$ | 1 | 1 | 32 | 1024 | 512 | 128 | 64 | 8 | 1024 |
| $10^8$ | 1 | 1 | 4 | 256 | 128 | 16 | 8 | 8 | 256 |
| $10^8$ | 1 | 0·5 | 128 | 512 | 256 | 64 | 32 | 16 | 512 |
| $10^8$ | 1 | 0·5 | 64 | 256 | 128 | 32 | 16 | 16 | 256 |
| $10^9$ | 1 | 1 | 1024 | 4096 | 1024 | 1024 | 256 | 256 | 4096 |
| $10^9$ | 1 | 1 | 2 | 512 | 1024 | 512 | 512 | | 1024 |
| $10^9$ | 1 | 1 | 32 | 1024 | 512 | 128 | 128 | 128 | 1024 |
| $10^{10}$ | 1 | 1 | 256 | 512 | 256 | | 128 | 128 | 512 |
| $10^{10}$ | 1 | 1 | 16 | 128 | | | 8 | 4 | 128 |
| $10^{10}$ | 1 | 2 | 128 | 256 | 128 | 32 | 16 | 16 | 256 |

**Dose- Expansion Module**

| Inaba (VAT) | | | | | | | | | |
|---|---|---|---|---|---|---|---|---|---|
| Dose (CFU) | D1 | D4 | D7 | D15 | D29 | D57 | D85 | D180 | max |
| 0 | 5 | 5 | 10 | 5 | 5 | 5 | 5 | 5 | 10 |
| 0 | 5 | 5 | 10 | 5 | 5 | 5 | 5 | 5 | 10 |
| 0 | 20 | 20 | 40 | 20 | 20 | 20 | 20 | 20 | 40 |
| 0 | 5 | 5 | 10 | 5 | 5 | 5 | 5 | 5 | 10 |
| 0 | 40 | 80 | 80 | 80 | 80 | 80 | 80 | 40 | 80 |
| 0 | 5 | 5 | 10 | 5 | 5 | 5 | 5 | 5 | 10 |
| 0 | 5 | 5 | 10 | 5 | 5 | 5 | 5 | 5 | 10 |
| $2\times10^7$ | 320 | 320 | 5120 | 10240 | 5120 | 5120 | 2560 | 2560 | 10240 |
| $2\times10^7$ | 5 | 5 | 160 | 2560 | 2560 | 1280 | 320 | 80 | 2560 |
| $2\times10^7$ | 20 | 40 | 5120 | 20480 | 5120 | 2560 | 1280 | 640 | 20480 |
| $2\times10^7$ | 40 | 40 | 640 | 10240 | 5120 | 2560 | 1280 | 1280 | 10240 |
| $2\times10^7$ | 5 | 5 | 2560 | 10240 | 5120 | 2560 | 1280 | 320 | 10240 |
| $2\times10^7$ | 160 | 160 | 1280 | 5120 | 5120 | 5120 | 5120 | 1280 | 5120 |
| $2\times10^7$ | 20 | | 160 | 2560 | 2560 | 1280 | 1280 | | 2560 |
| $2\times10^7$ | 10 | 10 | 80 | 2560 | 1280 | 1280 | 640 | 320 | 2560 |
| $2\times10^7$ | 5 | 5 | 1280 | 2560 | 1280 | 320 | 160 | 80 | 2560 |
| $2\times10^7$ | 10 | 20 | 5120 | 20480 | 5120 | 1280 | 1280 | 1280 | 20480 |
| $2\times10^7$ | 5 | 5 | 2560 | 10240 | 5120 | 5120 | 2560 | 1280 | 10240 |
| $2\times10^7$ | 5 | 5 | 1280 | 10240 | 10240 | 10240 | 10240 | 2560 | 10240 |
| $2\times10^7$ | 20 | 20 | 5120 | 20480 | 10240 | 5120 | 1280 | 320 | 20480 |
| $2\times10^7$ | 20 | 40 | 2560 | 10240 | 10240 | 2560 | | 640 | 10240 |
| $2\times10^8$ | 10 | 10 | 2560 | 10240 | 5120 | 5120 | 2560 | 1280 | 10240 |
| $2\times10^8$ | 5 | 5 | 320 | 5120 | 2560 | 320 | 160 | 80 | 5120 |
| $2\times10^8$ | 40 | 40 | 5120 | 20480 | 10240 | 5120 | 2560 | 2560 | 20480 |
| $2\times10^8$ | 5 | 5 | 320 | 2560 | 1280 | 2560 | 1280 | 640 | 2560 |
| $2\times10^8$ | 10 | 10 | 5120 | 40960 | 20480 | 5120 | 2560 | 2560 | 40960 |
| $2\times10^8$ | 5 | 5 | 5120 | 10240 | | | | | 10240 |
| $2\times10^8$ | 5 | 5 | 40 | 1280 | 320 | 160 | 80 | 80 | 1280 |
| $2\times10^8$ | 5 | 5 | 20 | 1280 | 160 | 20 | 10 | 5 | 1280 |
| $2\times10^8$ | 5 | 5 | 5120 | 10240 | 5120 | 2560 | 1280 | 640 | 10240 |
| $2\times10^8$ | 10 | 10 | 10240 | 20480 | 20480 | 5120 | 5120 | 2560 | 20480 |
| $2\times10^8$ | 5 | 5 | 160 | 5120 | 2560 | 640 | 320 | 320 | 5120 |
| $2\times10^8$ | 5 | 20 | 640 | 5120 | 1280 | | 320 | 320 | 5120 |
| $2\times10^8$ | 10 | 10 | 1280 | 10240 | 5120 | 1280 | 640 | 320 | 10240 |

| Fold change to baseline Inaba | | | | | | | | | |
|---|---|---|---|---|---|---|---|---|---|
| Dose (CFU) | D1 | D4 | D7 | D15 | D29 | D57 | D85 | D180 | max FC |
| 0 | 1 | 1 | 2 | 1 | 1 | 1 | 1 | 1 | 2 |
| 0 | 1 | 1 | 2 | 1 | 1 | 1 | 1 | 1 | 2 |
| 0 | 1 | 1 | 2 | 1 | 1 | 1 | 1 | 1 | 2 |
| 0 | 1 | 1 | 2 | 1 | 1 | 1 | 1 | 1 | 2 |
| 0 | 1 | 2 | 2 | 2 | 2 | 2 | 2 | 1 | 2 |
| 0 | 1 | 1 | 2 | 1 | 1 | 1 | 1 | 1 | 2 |
| 0 | 1 | 1 | 2 | 1 | 1 | 1 | 1 | 1 | 2 |
| $2\times10^7$ | 1 | 1 | 16 | 32 | 16 | 16 | 8 | 8 | 32 |
| $2\times10^7$ | 1 | 1 | 32 | 512 | 512 | 256 | 64 | 16 | 512 |
| $2\times10^7$ | 1 | 2 | 256 | 1024 | 256 | 128 | 64 | 32 | 1024 |
| $2\times10^7$ | 1 | 1 | 16 | 256 | 128 | 64 | 32 | 32 | 256 |
| $2\times10^7$ | 1 | 1 | 512 | 2048 | 1024 | 512 | 256 | 64 | 2048 |
| $2\times10^7$ | 1 | 1 | 8 | 32 | 32 | 32 | 32 | 8 | 32 |
| $2\times10^7$ | 1 | | 8 | 128 | 128 | 64 | 64 | | 128 |
| $2\times10^7$ | 1 | 1 | 8 | 256 | 128 | 128 | 64 | 32 | 256 |
| $2\times10^7$ | 1 | 1 | 256 | 512 | 256 | 64 | 32 | 16 | 512 |
| $2\times10^7$ | 1 | 2 | 512 | 2048 | 512 | 128 | 128 | 128 | 2048 |
| $2\times10^7$ | 1 | 1 | 512 | 2048 | 1024 | 1024 | 512 | 256 | 2048 |
| $2\times10^7$ | 1 | 1 | 256 | 2048 | 2048 | 2048 | 2048 | 512 | 2048 |
| $2\times10^7$ | 1 | 1 | 256 | 1024 | 512 | 256 | 64 | 16 | 1024 |
| $2\times10^7$ | 1 | 2 | 128 | 512 | 512 | 128 | | 32 | 512 |
| $2\times10^8$ | 1 | 1 | 256 | 1024 | 512 | 512 | 256 | 128 | 1024 |
| $2\times10^8$ | 1 | 1 | 64 | 1024 | 512 | 64 | 32 | 16 | 1024 |
| $2\times10^8$ | 1 | 1 | 128 | 512 | 256 | 128 | 64 | 64 | 512 |
| $2\times10^8$ | 1 | 1 | 64 | 512 | 256 | 512 | 256 | 128 | 512 |
| $2\times10^8$ | 1 | 1 | 512 | 4096 | 2048 | 512 | 256 | 256 | 4096 |
| $2\times10^8$ | 1 | 1 | 1024 | 2048 | | | | | 2048 |
| $2\times10^8$ | 1 | 1 | 8 | 256 | 64 | 32 | 16 | 16 | 256 |
| $2\times10^8$ | 1 | 1 | 4 | 256 | 32 | 4 | 2 | 1 | 256 |
| $2\times10^8$ | 1 | 1 | 1024 | 2048 | 1024 | 512 | 256 | 128 | 2048 |
| $2\times10^8$ | 1 | 1 | 1024 | 2048 | 2048 | 512 | 512 | 256 | 2048 |
| $2\times10^8$ | 1 | 1 | 32 | 1024 | 512 | 128 | 64 | 64 | 1024 |
| $2\times10^8$ | 1 | 4 | 128 | 1024 | 256 | | 64 | 64 | 1024 |
| $2\times10^8$ | 1 | 1 | 128 | 1024 | 512 | 128 | 64 | 32 | 1024 |

no serum sample

**Dose-Escalation Module**

| Ogawa (VAT) | | | | | | | | | |
|---|---|---|---|---|---|---|---|---|---|
| Dose (CFU) | D1 | D4 | D7 | D15 | D29 | D57 | D85 | D180 | max |
| $10^5$ | 5 | 5 | 20 | 320 | 160 | 80 | 160 | 160 | 320 |
| $10^5$ | 5 | 5 | 10 | 40 | 20 | 20 | 20 | | 40 |
| $10^5$ | 5 | 5 | 320 | 10240 | 2560 | 320 | 80 | | 10240 |
| $10^6$ | 80 | 80 | 1280 | 5120 | 2560 | 1280 | 640 | | 5120 |
| $10^6$ | 20 | 20 | 80 | 320 | 160 | 80 | 40 | 80 | 320 |
| $10^6$ | 10 | 10 | 320 | 10240 | 2560 | 1280 | 1280 | 1280 | 10240 |
| $10^7$ | 20 | 20 | 1280 | 5120 | 5120 | 2560 | 1280 | 640 | 5120 |
| $10^7$ | 40 | 40 | 1280 | 5120 | 5120 | 1280 | 640 | 160 | 5120 |
| $10^7$ | 5 | 5 | 40 | 2560 | 640 | 160 | 160 | 20 | 2560 |
| $10^8$ | 80 | 80 | 160 | 5120 | 1280 | 640 | 320 | 320 | 5120 |
| $10^8$ | 5 | 5 | 160 | 5120 | 1280 | 640 | 640 | 320 | 5120 |
| $10^8$ | 640 | 640 | 640 | 2560 | 1280 | 640 | 640 | 1280 | 2560 |
| $10^9$ | 5 | 5 | 2560 | 10240 | 2560 | 1280 | 320 | 320 | 10240 |
| $10^9$ | 5 | 5 | 10 | 320 | 1280 | 1280 | 1280 | | 1280 |
| $10^9$ | 80 | 160 | 160 | 1280 | 1280 | 640 | 640 | 1280 | 1280 |
| $10^{10}$ | 80 | 80 | 320 | 640 | 640 | | 160 | 160 | 640 |
| $10^{10}$ | 320 | 320 | 2560 | 40960 | | | 2560 | 1280 | 40960 |
| $10^{10}$ | 40 | 80 | 10240 | 10240 | 2560 | 2560 | 640 | 640 | 10240 |

| Fold change to baseline Ogawa | | | | | | | | | |
|---|---|---|---|---|---|---|---|---|---|
| Dose (CFU) | D1 | D4 | D7 | D15 | D29 | D57 | D85 | D180 | max FC |
| $10^5$ | 1 | 1 | 4 | 64 | 32 | 16 | 32 | 32 | 64 |
| $10^5$ | 1 | 1 | 2 | 8 | 4 | 4 | 4 | | 8 |
| $10^5$ | 1 | 1 | 64 | 2048 | 512 | 64 | 16 | | 2048 |
| $10^6$ | 1 | 1 | 16 | 64 | 32 | 16 | 8 | | 64 |
| $10^6$ | 1 | 1 | 4 | 16 | 8 | 4 | 2 | 4 | 16 |
| $10^6$ | 1 | 1 | 32 | 1024 | 256 | 128 | 128 | 128 | 1024 |
| $10^7$ | 1 | 1 | 64 | 256 | 256 | 128 | 64 | 32 | 256 |
| $10^7$ | 1 | 1 | 32 | 128 | 128 | 32 | 16 | 4 | 128 |
| $10^7$ | 1 | 1 | 8 | 512 | 128 | 32 | 32 | 4 | 512 |
| $10^8$ | 1 | 1 | 2 | 64 | 16 | 8 | 4 | 4 | 64 |
| $10^8$ | 1 | 1 | 32 | 1024 | 256 | 128 | 128 | 64 | 1024 |
| $10^8$ | 1 | 1 | 1 | 4 | 2 | 1 | 1 | 2 | 4 |
| $10^9$ | 1 | 1 | 512 | 2048 | 512 | 256 | 64 | 64 | 2048 |
| $10^9$ | 1 | 1 | 2 | 64 | 256 | 256 | 256 | | 256 |
| $10^9$ | 1 | 2 | 2 | 16 | 16 | 8 | 8 | 16 | 16 |
| $10^{10}$ | 1 | 1 | 4 | 8 | 8 | | 2 | 2 | 8 |
| $10^{10}$ | 1 | 1 | 8 | 128 | | | 8 | 4 | 128 |
| $10^{10}$ | 1 | 2 | 256 | 256 | 64 | 64 | 16 | 16 | 256 |

**Dose- Expansion Module**

| Ogawa (VAT) | | | | | | | | | |
|---|---|---|---|---|---|---|---|---|---|
| Dose (CFU) | D1 | D4 | D7 | D15 | D29 | D57 | D85 | D180 | max |
| 0 | 5 | 5 | 10 | 5 | 5 | 5 | 5 | 5 | 10 |
| 0 | 5 | 5 | 10 | 5 | 5 | 5 | 5 | 5 | 10 |
| 0 | 80 | 40 | 80 | 40 | 40 | 40 | 40 | 80 | 80 |
| 0 | 40 | 80 | 80 | 40 | 80 | 80 | 80 | 80 | 80 |
| 0 | 40 | 40 | 40 | 40 | 40 | 40 | 40 | 20 | 40 |
| 0 | 40 | 20 | 40 | 20 | 20 | 40 | 80 | 80 | 80 |
| 0 | 5 | 5 | 10 | 5 | 5 | 5 | 5 | 5 | 10 |
| $2\times10^7$ | 20 | 20 | 5120 | 10240 | 2560 | 2560 | 2560 | 1280 | 10240 |
| $2\times10^7$ | 80 | 80 | 640 | 10240 | 5120 | 5120 | 2560 | 640 | 10240 |
| $2\times10^7$ | 10 | 10 | 1280 | 5120 | 2560 | 1280 | 640 | 320 | 5120 |
| $2\times10^7$ | 10 | 20 | 80 | 1280 | 160 | 40 | 20 | 20 | 1280 |
| $2\times10^7$ | 5 | 5 | 1280 | 5120 | 2560 | 640 | 320 | 80 | 5120 |
| $2\times10^7$ | 5 | 5 | 640 | 2560 | 640 | 80 | 80 | 10 | 2560 |
| $2\times10^7$ | 5 | | 40 | 640 | 80 | 80 | 20 | | 640 |
| $2\times10^7$ | 20 | 40 | 80 | 1280 | 2560 | 1280 | 640 | 160 | 2560 |
| $2\times10^7$ | 5 | 5 | 320 | 1280 | 320 | 40 | 20 | 10 | 1280 |
| $2\times10^7$ | 5 | 10 | 1280 | 10240 | 5120 | 1280 | 1280 | 320 | 10240 |
| $2\times10^7$ | 5 | 5 | 640 | 5120 | 1280 | 1280 | 640 | 320 | 5120 |
| $2\times10^7$ | 5 | 5 | 80 | 640 | 320 | 320 | 320 | 320 | 640 |
| $2\times10^7$ | 5 | 5 | 160 | 640 | 160 | 40 | 10 | 5 | 640 |
| $2\times10^7$ | 5 | 5 | 640 | 10240 | 5120 | 2560 | | 640 | 10240 |
| $2\times10^8$ | 5 | 5 | 1280 | 5120 | 2560 | 2560 | 640 | 320 | 5120 |
| $2\times10^8$ | 5 | 5 | 640 | 2560 | 2560 | 640 | 320 | 160 | 2560 |
| $2\times10^8$ | 5 | 5 | 320 | 2560 | 1280 | 320 | 160 | 40 | 2560 |
| $2\times10^8$ | 160 | 160 | 640 | 1280 | 1280 | 640 | 640 | 1280 | 1280 |
| $2\times10^8$ | 5 | 5 | 160 | 640 | 320 | 80 | 40 | 40 | 640 |
| $2\times10^8$ | 20 | 20 | 1280 | 10240 | | | | | 10240 |
| $2\times10^8$ | 5 | 5 | 40 | 640 | 160 | 80 | 80 | 80 | 640 |
| $2\times10^8$ | 5 | 5 | 20 | 640 | 80 | 10 | 5 | 5 | 640 |
| $2\times10^8$ | 5 | 5 | 2560 | 10240 | 2560 | 1280 | 1280 | 640 | 10240 |
| $2\times10^8$ | 320 | 160 | 5120 | 20480 | 20480 | 10240 | 5120 | 2560 | 20480 |
| $2\times10^8$ | 5 | 5 | 320 | 5120 | 5120 | 640 | 320 | 320 | 5120 |
| $2\times10^8$ | 5 | 20 | 160 | 1280 | 640 | | 160 | 80 | 1280 |
| $2\times10^8$ | 5 | 5 | 20 | 160 | 80 | 40 | 20 | 20 | 160 |

| Fold change to baseline Ogawa | | | | | | | | | |
|---|---|---|---|---|---|---|---|---|---|
| Dose (CFU) | D1 | D4 | D7 | D15 | D29 | D57 | D85 | D180 | max FC |
| 0 | 1 | 1 | 2 | 1 | 1 | 1 | 1 | 1 | 2 |
| 0 | 1 | 1 | 2 | 1 | 1 | 1 | 1 | 1 | 2 |
| 0 | 1 | 0·5 | 1 | 0·5 | 0·5 | 0·5 | 0·5 | 1 | 1 |
| 0 | 1 | 2 | 2 | 1 | 2 | 2 | 2 | 2 | 2 |
| 0 | 1 | 1 | 1 | 1 | 1 | 1 | 1 | 0·5 | 1 |
| 0 | 1 | 0·5 | 1 | 0·5 | 0·5 | 1 | 2 | 2 | 2 |
| 0 | 1 | 1 | 2 | 1 | 1 | 1 | 1 | 1 | 2 |
| $2\times10^7$ | 1 | 1 | 256 | 512 | 128 | 128 | 128 | 64 | 512 |
| $2\times10^7$ | 1 | 1 | 8 | 128 | 64 | 64 | 32 | 8 | 128 |
| $2\times10^7$ | 1 | 1 | 128 | 512 | 256 | 128 | 64 | 32 | 512 |
| $2\times10^7$ | 1 | 2 | 8 | 128 | 16 | 4 | 2 | 2 | 128 |
| $2\times10^7$ | 1 | 1 | 256 | 1024 | 512 | 128 | 64 | 16 | 1024 |
| $2\times10^7$ | 1 | 1 | 128 | 512 | 128 | 16 | 16 | 2 | 512 |
| $2\times10^7$ | 1 | | 8 | 128 | 16 | 16 | 4 | | 128 |
| $2\times10^7$ | 1 | 2 | 4 | 64 | 128 | 64 | 32 | 8 | 128 |
| $2\times10^7$ | 1 | 1 | 64 | 256 | 64 | 8 | 4 | 2 | 256 |
| $2\times10^7$ | 1 | 2 | 256 | 2048 | 1024 | 256 | 256 | 64 | 2048 |
| $2\times10^7$ | 1 | 1 | 128 | 1024 | 256 | 256 | 128 | 64 | 1024 |
| $2\times10^7$ | 1 | 1 | 16 | 128 | 64 | 64 | 64 | 64 | 128 |
| $2\times10^7$ | 1 | 1 | 32 | 128 | 32 | 8 | 2 | 1 | 128 |
| $2\times10^7$ | 1 | 1 | 128 | 2048 | 1024 | 512 | | 128 | 2048 |
| $2\times10^8$ | 1 | 1 | 256 | 1024 | 512 | 512 | 128 | 64 | 1024 |
| $2\times10^8$ | 1 | 1 | 128 | 512 | 512 | 128 | 64 | 32 | 512 |
| $2\times10^8$ | 1 | 1 | 64 | 512 | 256 | 64 | 32 | 8 | 512 |
| $2\times10^8$ | 1 | 1 | 4 | 8 | 8 | 4 | 4 | 8 | 8 |
| $2\times10^8$ | 1 | 1 | 32 | 128 | 64 | 16 | 8 | 8 | 128 |
| $2\times10^8$ | 1 | 1 | 64 | 512 | | | | | 512 |
| $2\times10^8$ | 1 | 1 | 8 | 128 | 32 | 16 | 16 | 16 | 128 |
| $2\times10^8$ | 1 | 1 | 4 | 128 | 16 | 2 | 1 | 1 | 128 |
| $2\times10^8$ | 1 | 1 | 512 | 2048 | 512 | 256 | 256 | 128 | 2048 |
| $2\times10^8$ | 1 | 0·5 | 16 | 64 | 64 | 32 | 16 | 8 | 64 |
| $2\times10^8$ | 1 | 1 | 64 | 1024 | 1024 | 128 | 64 | 64 | 1024 |
| $2\times10^8$ | 1 | 4 | 32 | 256 | 128 | | 32 | 16 | 256 |
| $2\times10^8$ | 1 | 1 | 4 | 32 | 16 | 8 | 4 | 4 | 32 |

**Table S7: Kinetics of mean vibriocidal titers to both serotypes in participants in the dose-expansion module.** Geometric mean titers with 95% confidence intervals are presented at baseline and day 7, 15, 29 and 180 post-vaccination. * p<0·0001 representing the comparison of day 15 vibriocidal titers of placebo and vaccine recipients dosed either with $10^7$ or $10^8$ CFU (Mann-Whitney test).

| Serotype | Dose group (CFU) | n | GMT (95% CI) post vaccination (days) | | | | |
|---|---|---|---|---|---|---|---|
| | | | Baseline | 7 | 15 | 29 | 180 |
| Inaba | 0 (Placebo) | 7 | 8·2 (3·7-18·3) | 16·4 (7·3-36·7) | 9·1 (3·3-24·8) | 9·1 (3·3-24·8) | 8·2 (3·7-18·3) |
| | $10^7$ | 17 | 13·3 (6·9-25·6) | 923·7 (433·2-1970) | 6811 (4567-10157)* | 4176 (3009-5795) | 493·5 (248·5-980) |
| | $10^8$ | 16 | 8·4 (5·7-12·5) | 794·8 (262-2411) | 6640 (3815-11555)* | 2808 (1288-6122) | 335·1 (128·8-872·2) |
| Ogawa | 0 (Placebo) | 7 | 18·1 (5·8-56·4) | 26·9 (11-65·9) | 14·9 (5·6-39·2) | 16·4 (5·5-48·9) | 20 (5·5-72·1) |
| | $10^7$ | 17 | 9·2 (5·9-14·5) | 392·4 (186·7-824·8) | 3013 (1778-5107)* | 1180 (566·6-2457) | 118·1 (46-303·6) |
| | $10^8$ | 16 | 14·1 (5·6-35·7) | 306·4 (129·9-722·7) | 2451 (1241-4842)* | 1016 (437·8-2358) | 175·5 (67·9-453·7) |

GMT, geometric mean titer; CI, confidence interval; **p*< 0·0001, comparison of vibriocidal antibody titers of placebo and vaccine recipients on D15 (Mann-Whitney test)

**Table S8: Mean peak vibriocidal titers and mean maximum fold increase in vibriocidal titers by dose group.** Mean peak fold increase in titer was calculated using each participant's baseline and peak titer.

| Serotype | Dose group (CFU) | n | Mean peak titer | Mean peak fold increase | Serotype | Dose group (CFU) | n | Mean peak titer | Mean peak fold increase |
|---|---|---|---|---|---|---|---|---|---|
| **Inaba** | 0 (Placebo) | 7 | 24·3 | 2 | **Ogawa** | 0 (Placebo) | 7 | 44·3 | 1·7 |
| | $10^5$ | 3 | 11947 | 1728 | | $10^5$ | 3 | 3533 | 706·7 |
| | $10^6$ | 3 | 5547 | 512 | | $10^6$ | 3 | 5227 | 368 |
| | $10^7$ | 17 | 9035 | 914·8 | | $10^7$ | 17 | 4631 | 564·7 |
| | $10^8$ | 16 | 10320 | 1120 | | $10^8$ | 16 | 4610 | 466·8 |
| | $10^9$ | 3 | 10240 | 2048 | | $10^9$ | 3 | 4267 | 773·3 |
| | $10^{10}$ | 3 | 31573 | 298·7 | | $10^{10}$ | 3 | 17280 | 130·7 |

**Table S9: Isotype and antigen specific immune responses in sera derived from vaccine and placebo recipients.** Dark blue: no serum sample; yellow: below limit of detection.

**Dose-Escalation Module**

| | Inaba IgM (RAU) | | | | | | | | |
|---|---|---|---|---|---|---|---|---|---|
| Dose (CFU) | D1 | D4 | D7 | D15 | D29 | D57 | D85 | D180 | max |
| $10^5$ | 0·0175 | 0·0144 | 0·0306 | 0·1644 | 0·0958 | 0·1024 | 0·1103 | 0·1715 | 0·1715 |
| $10^5$ | 0·0126 | 0·0095 | 0·0162 | 0·1764 | 0·1516 | 0·0761 | 0·1024 | | 0·1764 |
| $10^5$ | 0·1793 | 0·1833 | 0·3245 | 3·1615 | 1·4471 | 0·3311 | 0·1909 | | 3·1615 |
| $10^6$ | 0·0363 | 0·0276 | 0·1923 | 9·3973 | 5·2374 | 1·3277 | 0·7381 | | 9·3973 |
| $10^6$ | 0·0561 | 0·0682 | 0·0965 | 0·1408 | 0·0768 | 0·0737 | 0·0640 | 0·0437 | 0·1408 |
| $10^6$ | 0·0093 | 0·0068 | 0·0456 | 1·6252 | 0·6483 | 0·3039 | 0·2319 | 0·0299 | 1·6252 |
| $10^7$ | 0·0414 | 0·0369 | 0·0648 | 0·2198 | 0·1506 | 0·1381 | 0·1072 | 0·0439 | 0·2198 |
| $10^7$ | 0·0124 | 0·0179 | 0·0505 | 2·1703 | 1·6044 | 0·4923 | 0·1157 | 0·0277 | 2·1703 |
| $10^7$ | 0·0768 | 0·0790 | 0·0545 | 0·9049 | 0·3188 | 0·1712 | 0·1207 | 0·0696 | 0·9049 |
| $10^8$ | 0·0089 | 0·0076 | 0·0075 | 0·2221 | 0·1372 | 0·0391 | 0·0228 | 0·0177 | 0·2221 |
| $10^8$ | 0·1379 | 0·1208 | 0·1937 | 1·8950 | 0·5626 | 0·3126 | 0·0977 | 0·0854 | 1·8950 |
| $10^8$ | 0·0656 | 0·0631 | 0·6425 | 3·9774 | 1·8408 | 0·1766 | 0·0958 | 0·1381 | 3·9774 |
| $10^9$ | 0·0441 | 0·0434 | 2·4562 | 43·1117 | 10·3921 | 1·4913 | 0·4487 | 0·2506 | 43·1117 |
| $10^9$ | 0·0347 | 0·0346 | 0·0243 | 0·3168 | 0·6364 | 0·7431 | 1·4028 | | 1·4028 |
| $10^9$ | 0·0590 | 0·0575 | 0·0717 | 0·5801 | 0·1343 | 0·1043 | 0·0990 | 0·0846 | 0·5801 |
| $10^{10}$ | 0·0375 | 0·0431 | 0·0617 | 0·1547 | 0·1696 | | 0·0773 | 0·0613 | 0·1696 |
| $10^{10}$ | 0·1880 | 0·2335 | 3·0780 | 67·9807 | | | 4·6855 | 1·0156 | 67·9807 |
| $10^{10}$ | 0·0426 | 0·0453 | 3·3315 | 9·4811 | 2·6635 | 1·1948 | 0·3265 | 0·1855 | 9·4811 |

| | Fold change Inaba IgM | | | | | | | | |
|---|---|---|---|---|---|---|---|---|---|
| Dose (CFU) | D1 | D4 | D7 | D15 | D29 | D57 | D85 | D180 | max FC |
| $10^5$ | 1·00 | 0·82 | 1·75 | 9·40 | 5·48 | 5·85 | 6·31 | 9·81 | 9·81 |
| $10^5$ | 1·00 | 0·75 | 1·29 | 14·03 | 12·06 | 6·06 | 8·14 | | 14·03 |
| $10^5$ | 1·00 | 1·02 | 1·81 | 17·63 | 8·07 | 1·85 | 1·06 | | 17·63 |
| $10^6$ | 1·00 | 0·76 | 5·29 | 258·60 | 144·13 | 36·54 | 20·31 | | 258·60 |
| $10^6$ | 1·00 | 1·22 | 1·72 | 2·51 | 1·37 | 1·31 | 1·14 | 0·78 | 2·51 |
| $10^6$ | 1·00 | 0·73 | 4·88 | 174·07 | 69·43 | 32·55 | 24·83 | 3·20 | 174·07 |
| $10^7$ | 1·00 | 0·89 | 1·57 | 5·31 | 3·64 | 3·34 | 2·59 | 1·06 | 5·31 |
| $10^7$ | 1·00 | 1·44 | 4·07 | 174·76 | 129·20 | 39·64 | 9·32 | 2·23 | 174·76 |
| $10^7$ | 1·00 | 1·03 | 0·71 | 11·78 | 4·15 | 2·23 | 1·57 | 0·91 | 11·78 |
| $10^8$ | 1·00 | 0·85 | 0·84 | 24·93 | 15·40 | 4·39 | 2·56 | 1·98 | 24·93 |
| $10^8$ | 1·00 | 0·88 | 1·40 | 13·74 | 4·08 | 2·27 | 0·71 | 0·62 | 13·74 |
| $10^8$ | 1·00 | 0·96 | 9·80 | 60·65 | 28·07 | 2·69 | 1·46 | 2·11 | 60·65 |
| $10^9$ | 1·00 | 0·98 | 55·69 | 977·49 | 235·62 | 33·81 | 10·17 | 5·68 | 977·49 |
| $10^9$ | 1·00 | 1·00 | 0·70 | 9·13 | 18·35 | 21·42 | 40·44 | | 40·44 |
| $10^9$ | 1·00 | 0·98 | 1·22 | 9·83 | 2·28 | 1·77 | 1·68 | 1·43 | 9·83 |
| $10^{10}$ | 1·00 | 1·15 | 1·65 | 4·13 | 4·53 | | 2·06 | 1·64 | 4·53 |
| $10^{10}$ | 1·00 | 1·24 | 16·37 | 361·61 | | | 24·92 | 5·40 | 361·61 |
| $10^{10}$ | 1·00 | 1·06 | 78·23 | 222·63 | 62·54 | 28·06 | 7·67 | 4·36 | 222·63 |

**Dose-Expansion Module**

| | Inaba IgM (RAU) | | | | | | | | |
|---|---|---|---|---|---|---|---|---|---|
| Dose (CFU) | D1 | D4 | D7 | D15 | D29 | D57 | D85 | D180 | max |
| 0 | 0·1556 | 0·1464 | 0·1584 | 0·1284 | 0·1609 | 0·1492 | 0·1502 | 0·1872 | 0·1872 |
| 0 | 0·0211 | 0·0208 | 0·0191 | 0·0195 | 0·0177 | 0·0213 | 0·0131 | 0·0140 | 0·0213 |
| 0 | 0·0627 | 0·0622 | 0·0801 | 0·0736 | 0·0683 | 0·0492 | 0·0421 | 0·0236 | 0·0801 |
| 0 | 0·1555 | 0·1616 | 0·1718 | 0·1349 | 0·1278 | 0·1087 | 0·0582 | 0·0623 | 0·1718 |
| 0 | 0·0812 | 0·0851 | 0·1115 | 0·1079 | 0·1197 | 0·0603 | 0·0527 | 0·0341 | 0·1197 |
| 0 | 0·1112 | 0·1201 | 0·1267 | 0·0696 | 0·0476 | 0·0736 | 0·0760 | 0·1049 | 0·1267 |
| 0 | 0·0373 | 0·0315 | 0·0445 | 0·0364 | 0·0566 | 0·0495 | 0·0297 | 0·0525 | 0·0566 |
| $2\times10^7$ | 0·2693 | 0·2433 | 0·5189 | 16·4138 | 5·4016 | 1·0056 | 0·6938 | 0·5244 | 16·4138 |
| $2\times10^7$ | 0·0400 | 0·0392 | 0·0891 | 11·0863 | 8·2969 | 2·8677 | 1·2419 | 0·2575 | 11·0863 |
| $2\times10^7$ | 0·0803 | 0·0948 | 2·6074 | 18·0558 | 8·2150 | 1·9866 | 0·7150 | 0·3734 | 18·0558 |
| $2\times10^7$ | 0·1078 | 0·1480 | 0·1288 | 0·6363 | 0·2056 | 0·1478 | 0·1271 | 0·0346 | 0·6363 |
| $2\times10^7$ | 0·1156 | 0·0962 | 0·2580 | 7·8914 | 3·3725 | 0·8086 | 0·5265 | 0·1212 | 7·8914 |
| $2\times10^7$ | 0·0365 | 0·0378 | 0·0385 | 0·3952 | 0·1945 | 0·0633 | 0·0645 | 0·0205 | 0·3952 |
| $2\times10^7$ | 0·1209 | | 0·1357 | 0·8424 | 0·5591 | 0·3802 | 0·2052 | | 0·8424 |
| $2\times10^7$ | 0·0975 | 0·1157 | 0·2102 | 17·1489 | 10·1604 | 4·5261 | 2·4544 | 0·5908 | 17·1489 |
| $2\times10^7$ | 0·0456 | 0·0541 | 0·3353 | 5·4943 | 2·8543 | 0·2710 | 0·1302 | 0·0696 | 5·4943 |
| $2\times10^7$ | 0·1890 | 0·2335 | 0·4092 | 10·6095 | 2·8424 | 0·8038 | 0·5760 | 0·3796 | 10·6095 |
| $2\times10^7$ | 0·0423 | 0·0670 | 0·1116 | 0·9685 | 0·9707 | 0·5488 | 0·4045 | 0·2794 | 0·9707 |
| $2\times10^7$ | 0·0272 | 0·0239 | 0·0892 | 2·0537 | 2·1348 | 1·5186 | 1·1463 | 0·6743 | 2·1348 |
| $2\times10^7$ | 0·0332 | 0·0415 | 1·8596 | 31·6258 | 14·0009 | 2·8981 | 0·6714 | 0·0838 | 31·6258 |
| $2\times10^7$ | 0·0398 | 0·0284 | 0·3938 | 5·9937 | 4·4841 | 1·6582 | | 0·4012 | 5·9937 |
| $2\times10^8$ | 0·1083 | 0·1016 | 2·8587 | 31·4981 | 15·7439 | 3·2397 | 1·5242 | 0·7598 | 31·4981 |
| $2\times10^8$ | 0·0125 | 0·0141 | 0·0262 | 1·1775 | 0·3358 | 0·0738 | 0·0570 | 0·0283 | 1·1775 |
| $2\times10^8$ | 0·1293 | 0·1374 | 1·4948 | 33·0181 | 9·6768 | 3·9240 | 1·0271 | 0·3311 | 33·0181 |
| $2\times10^8$ | 0·2113 | 0·2248 | 0·2040 | 0·6589 | 0·8768 | 0·5750 | 0·3167 | 0·1348 | 0·8768 |
| $2\times10^8$ | 0·0649 | 0·0764 | 0·2231 | 7·8785 | 3·8783 | 0·2816 | 0·2252 | 0·1384 | 7·8785 |
| $2\times10^8$ | 0·1192 | 0·1281 | 0·1424 | 2·0385 | | | | | 2·0385 |
| $2\times10^8$ | 0·2210 | 0·2278 | 0·2289 | 1·1330 | 0·2619 | 0·0940 | 0·0640 | 0·0484 | 1·1330 |
| $2\times10^8$ | 0·0196 | 0·0151 | 0·0151 | 0·3434 | 0·1100 | 0·0211 | 0·0145 | 0·0043 | 0·3434 |
| $2\times10^8$ | 0·0449 | 0·0388 | 1·3228 | 30·0340 | 10·2997 | 2·1627 | 1·4575 | 0·7240 | 30·0340 |
| $2\times10^8$ | 0·0143 | 0·0166 | 2·0431 | 39·7839 | 23·1500 | 7·7571 | 3·7617 | 1·7124 | 39·7839 |
| $2\times10^8$ | 0·0100 | 0·0112 | 0·0353 | 3·0476 | 1·8180 | 0·5164 | 0·2710 | 0·1474 | 3·0476 |
| $2\times10^8$ | 0·0136 | 0·0173 | 0·1995 | 2·4154 | 0·8180 | | 0·1286 | 0·1154 | 2·4154 |
| $2\times10^8$ | 0·0042 | 0·0049 | 0·2048 | 2·3459 | 1·2405 | 0·3463 | 0·2207 | 0·0872 | 2·3459 |

| | Fold change Inaba IgM | | | | | | | | |
|---|---|---|---|---|---|---|---|---|---|
| Dose (CFU) | D1 | D4 | D7 | D15 | D29 | D57 | D85 | D180 | max FC |
| 0 | 1·00 | 0·94 | 1·02 | 0·82 | 1·03 | 0·96 | 0·97 | 1·20 | 1·20 |
| 0 | 1·00 | 0·99 | 0·91 | 0·93 | 0·84 | 1·01 | 0·62 | 0·66 | 1·01 |
| 0 | 1·00 | 0·99 | 1·28 | 1·17 | 1·09 | 0·78 | 0·67 | 0·38 | 1·28 |
| 0 | 1·00 | 1·04 | 1·10 | 0·87 | 0·82 | 0·70 | 0·37 | 0·40 | 1·10 |
| 0 | 1·00 | 1·05 | 1·37 | 1·33 | 1·47 | 0·74 | 0·65 | 0·42 | 1·47 |
| 0 | 1·00 | 1·08 | 1·14 | 0·63 | 0·43 | 0·66 | 0·68 | 0·94 | 1·14 |
| 0 | 1·00 | 0·85 | 1·20 | 0·98 | 1·52 | 1·33 | 0·80 | 1·41 | 1·52 |
| $2\times10^7$ | 1·00 | 0·90 | 1·93 | 60·96 | 20·06 | 3·73 | 2·58 | 1·95 | 60·96 |
| $2\times10^7$ | 1·00 | 0·98 | 2·22 | 276·88 | 207·21 | 71·62 | 31·02 | 6·43 | 276·88 |
| $2\times10^7$ | 1·00 | 1·18 | 32·48 | 224·95 | 102·35 | 24·75 | 8·91 | 4·65 | 224·95 |
| $2\times10^7$ | 1·00 | 1·37 | 1·19 | 5·90 | 1·91 | 1·37 | 1·18 | 0·32 | 5·90 |
| $2\times10^7$ | 1·00 | 0·83 | 2·23 | 68·25 | 29·17 | 6·99 | 4·55 | 1·05 | 68·25 |
| $2\times10^7$ | 1·00 | 1·03 | 1·05 | 10·81 | 5·32 | 1·73 | 1·77 | 0·56 | 10·81 |
| $2\times10^7$ | 1·00 | | 1·12 | 6·97 | 4·63 | 3·15 | 1·70 | | 6·97 |
| $2\times10^7$ | 1·00 | 1·19 | 2·16 | 175·87 | 104·20 | 46·42 | 25·17 | 6·06 | 175·87 |
| $2\times10^7$ | 1·00 | 1·19 | 7·36 | 120·58 | 62·64 | 5·95 | 2·86 | 1·53 | 120·58 |
| $2\times10^7$ | 1·00 | 1·24 | 2·16 | 56·13 | 15·04 | 4·25 | 3·05 | 2·01 | 56·13 |
| $2\times10^7$ | 1·00 | 1·58 | 2·64 | 22·87 | 22·92 | 12·96 | 9·55 | 6·60 | 22·92 |
| $2\times10^7$ | 1·00 | 0·88 | 3·28 | 75·43 | 78·41 | 55·78 | 42·11 | 24·77 | 78·41 |
| $2\times10^7$ | 1·00 | 1·25 | 56·05 | 953·26 | 422·01 | 87·35 | 20·24 | 2·52 | 953·26 |
| $2\times10^7$ | 1·00 | 0·71 | 9·90 | 150·66 | 112·71 | 41·68 | | 10·08 | 150·66 |
| $2\times10^8$ | 1·00 | 0·94 | 26·38 | 290·71 | 145·31 | 29·90 | 14·07 | 7·01 | 290·71 |
| $2\times10^8$ | 1·00 | 1·14 | 2·10 | 94·56 | 26·97 | 5·93 | 4·57 | 2·27 | 94·56 |
| $2\times10^8$ | 1·00 | 1·06 | 11·56 | 255·29 | 74·82 | 30·34 | 7·94 | 2·56 | 255·29 |
| $2\times10^8$ | 1·00 | 1·06 | 0·97 | 3·12 | 4·15 | 2·72 | 1·50 | 0·64 | 4·15 |
| $2\times10^8$ | 1·00 | 1·18 | 3·44 | 121·41 | 59·77 | 4·34 | 3·47 | 2·13 | 121·41 |
| $2\times10^8$ | 1·00 | 1·07 | 1·20 | 17·10 | | | | | 17·10 |
| $2\times10^8$ | 1·00 | 1·03 | 1·04 | 5·13 | 1·18 | 0·43 | 0·29 | 0·22 | 5·13 |
| $2\times10^8$ | 1·00 | 0·77 | 0·77 | 17·52 | 5·61 | 1·08 | 0·74 | 0·22 | 17·52 |
| $2\times10^8$ | 1·00 | 0·86 | 29·47 | 669·00 | 229·42 | 48·17 | 32·47 | 16·13 | 669·00 |
| $2\times10^8$ | 1·00 | 1·16 | 143·35 | 2791·22 | 1624·19 | 544·24 | 263·92 | 120·14 | 2791·22 |
| $2\times10^8$ | 1·00 | 1·13 | 3·54 | 305·70 | 182·36 | 51·80 | 27·19 | 14·78 | 305·70 |
| $2\times10^8$ | 1·00 | 1·27 | 14·66 | 177·48 | 60·10 | | 9·45 | 8·48 | 177·48 |
| $2\times10^8$ | 1·00 | 1·15 | 48·37 | 554·02 | 292·97 | 81·78 | 52·11 | 20·59 | 554·02 |

no serum sample

| | RAU | | |
|---|---|---|---|
| | D1 | D39 | |
| *V. cholerae* | 0·0146 | 0·1628 | |
| | 0·0700 | 6·7315 | |
| | 0·0300 | 0·2255 | |
| | 0·2800 | 15·4587 | |
| | 0·0411 | 9·2681 | |
| | D1 | D11 | |
| Vaxchora D11 Challenge | 0·0092 | 1·7460 | |
| | 0·1711 | 0·2180 | |
| | 0·0105 | 1·0184 | |
| | 0·0231 | 0·1162 | |
| | 0·0513 | 0·2466 | |
| | 0·0275 | 3·7451 | |
| | D1 | D11 | D91 |
| Vaxchora D91 Challenge | 0·0083 | 0·0602 | 0·0159 |
| | 0·0573 | 0·8117 | 0·0542 |
| | 0·0011 | 0·1309 | 0·0124 |
| | 0·2200 | 0·2229 | 0·3015 |
| | 0·1382 | 1·7817 | 0·1984 |
| | 0·0047 | 0·5876 | 0·0221 |

| | RAU fold change | | | max FC |
|---|---|---|---|---|
| | D1 | D39 | | |
| *V. cholerae* | 1·00 | 11·17 | | 11·17 |
| | 1·00 | 96·15 | | 96·15 |
| | 1·00 | 7·52 | | 7·52 |
| | 1·00 | 55·20 | | 55·20 |
| | 1·00 | 225·62 | | 225·62 |
| | D1 | D11 | | |
| Vaxchora D11 Challenge | 1·00 | 188·92 | | 188·92 |
| | 1·00 | 1·27 | | 1·27 |
| | 1·00 | 96·76 | | 96·76 |
| | 1·00 | 5·04 | | 5·04 |
| | 1·00 | 4·80 | | 4·80 |
| | 1·00 | 136·23 | | 136·23 |
| | D1 | D11 | D91 | |
| Vaxchora D91 Challenge | 1·00 | 7·26 | 1·92 | 7·26 |
| | 1·00 | 14·17 | 0·95 | 14·17 |
| | 1·00 | 117·29 | 11·13 | 117·29 |
| | 1·00 | 1·01 | 1·37 | 1·37 |
| | 1·00 | 12·89 | 1·44 | 12·89 |
| | 1·00 | 125·07 | 4·70 | 125·07 |

**Dose-Escalation Module**

| | Inaba IgG (RAU) | | | | | | | | |
|---|---|---|---|---|---|---|---|---|---|
| Dose (CFU) | D1 | D4 | D7 | D15 | D29 | D57 | D85 | D180 | max |
| $10^5$ | 0·0194 | 0·0115 | 0·0177 | 0·0163 | 0·0139 | 0·0100 | 0·0174 | 0·0441 | 0·0441 |
| $10^5$ | 0·0140 | 0·0130 | 0·0205 | 0·0660 | 0·0423 | 0·0433 | 0·0456 | | 0·0660 |
| $10^5$ | 0·0250 | 0·0239 | 0·0268 | 22·2367 | 9·4033 | 1·7544 | 1·3084 | | 22·2367 |
| $10^6$ | 0·0307 | 0·0318 | 0·0297 | 1·0718 | 0·6311 | 0·6985 | 0·7886 | | 1·0718 |
| $10^6$ | 0·0241 | 0·0286 | 0·0338 | 0·0497 | 0·0590 | 0·1282 | 0·1726 | 0·2262 | 0·2262 |
| $10^6$ | 0·0430 | 0·0421 | 0·1310 | 5·2376 | 2·9138 | 1·4099 | 0·7167 | 0·2842 | 5·2376 |
| $10^7$ | 0·1248 | 0·1254 | 0·1109 | 0·1214 | 0·1443 | 0·3133 | 0·3248 | 0·8199 | 0·8199 |
| $10^7$ | 0·0544 | 0·0684 | 0·0786 | 0·1541 | 0·0871 | 0·0655 | 0·0909 | 0·0746 | 0·1541 |
| $10^7$ | 0·0455 | 0·0438 | 0·0472 | 0·1068 | 0·0739 | 0·0560 | 0·0619 | 0·0675 | 0·1068 |
| $10^8$ | 0·1150 | 0·1121 | 0·1150 | 0·1074 | 0·1214 | 0·1020 | 0·1503 | 0·7338 | 0·7338 |
| $10^8$ | 0·0977 | 0·0909 | 0·0793 | 0·8510 | 0·4097 | 0·3496 | 0·1479 | 0·1644 | 0·8510 |
| $10^8$ | 0·1265 | 0·0877 | 0·1044 | 0·1197 | 0·1271 | 0·0704 | 0·1687 | 0·4659 | 0·4659 |
| $10^9$ | 0·0480 | 0·0641 | 0·0705 | 0·0725 | 0·0746 | 0·0610 | 0·0870 | 0·1516 | 0·1516 |
| $10^9$ | 0·0307 | 0·0058 | 0·0203 | 0·0352 | 0·1442 | 0·2464 | 1·3151 | | 1·3151 |
| $10^9$ | 0·0528 | 0·0450 | 0·0434 | 0·0573 | 0·0535 | 0·0505 | 0·0580 | 0·0808 | 0·0808 |
| $10^{10}$ | 0·0318 | 0·0361 | 0·0466 | 0·1116 | 0·0372 | | 0·0490 | 0·0774 | 0·1116 |
| $10^{10}$ | 0·0168 | 0·0157 | 0·0208 | 0·0580 | | | 0·0265 | 0·0168 | 0·0580 |
| $10^{10}$ | 0·0815 | 0·0801 | 0·1056 | 0·1120 | 0·1153 | 0·1305 | 0·1632 | 0·2353 | 0·2353 |

| | Fold change Inaba IgG | | | | | | | | |
|---|---|---|---|---|---|---|---|---|---|
| Dose (CFU) | D1 | D4 | D7 | D15 | D29 | D57 | D85 | D180 | max FC |
| $10^5$ | 1·00 | 0·59 | 0·91 | 0·84 | 0·72 | 0·52 | 0·90 | 2·28 | 2·28 |
| $10^5$ | 1·00 | 0·92 | 1·46 | 4·71 | 3·01 | 3·08 | 3·25 | | 4·71 |
| $10^5$ | 1·00 | 0·95 | 1·07 | 890·25 | 376·46 | 70·24 | 52·38 | | 890·25 |
| $10^6$ | 1·00 | 1·03 | 0·97 | 34·86 | 20·53 | 22·72 | 25·65 | | 34·86 |
| $10^6$ | 1·00 | 1·19 | 1·40 | 2·06 | 2·45 | 5·32 | 7·16 | 9·39 | 9·39 |
| $10^6$ | 1·00 | 0·98 | 3·05 | 121·87 | 67·80 | 32·80 | 16·68 | 6·61 | 121·87 |
| $10^7$ | 1·00 | 1·00 | 0·89 | 0·97 | 1·16 | 2·51 | 2·60 | 6·57 | 6·57 |
| $10^7$ | 1·00 | 1·26 | 1·44 | 2·83 | 1·60 | 1·20 | 1·67 | 1·37 | 2·83 |
| $10^7$ | 1·00 | 0·96 | 1·04 | 2·34 | 1·62 | 1·23 | 1·36 | 1·48 | 2·34 |
| $10^8$ | 1·00 | 0·97 | 1·00 | 0·93 | 1·06 | 0·89 | 1·31 | 6·38 | 6·38 |
| $10^8$ | 1·00 | 0·93 | 0·81 | 8·71 | 4·19 | 3·58 | 1·51 | 1·68 | 8·71 |
| $10^8$ | 1·00 | 0·69 | 0·83 | 0·95 | 1·00 | 0·56 | 1·33 | 3·68 | 3·68 |
| $10^9$ | 1·00 | 1·33 | 1·47 | 1·51 | 1·55 | 1·27 | 1·81 | 3·16 | 3·16 |
| $10^9$ | 1·00 | 0·19 | 0·66 | 1·15 | 4·69 | 8·01 | 42·77 | | 42·77 |
| $10^9$ | 1·00 | 0·85 | 0·82 | 1·09 | 1·01 | 0·96 | 1·10 | 1·53 | 1·53 |
| $10^{10}$ | 1·00 | 1·13 | 1·47 | 3·51 | 1·17 | | 1·54 | 2·43 | 3·51 |
| $10^{10}$ | 1·00 | 0·94 | 1·24 | 3·46 | | | 1·58 | 1·00 | 3·46 |
| $10^{10}$ | 1·00 | 0·98 | 1·29 | 1·37 | 1·41 | 1·60 | 2·00 | 2·89 | 2·89 |

**Dose- Expansion Module**

| | Inaba IgG (RAU) | | | | | | | | |
|---|---|---|---|---|---|---|---|---|---|
| Dose (CFU) | D1 | D4 | D7 | D15 | D29 | D57 | D85 | D180 | max |
| 0 | 0·0267 | 0·0398 | 0·0320 | 0·0341 | 0·0386 | 0·0347 | 0·0426 | 0·0524 | 0·0524 |
| 0 | 0·1160 | 0·1445 | 0·1264 | 0·1043 | 0·1103 | 0·1073 | 0·0973 | 0·0931 | 0·1445 |
| 0 | 0·0664 | 0·0784 | 0·0824 | 0·0678 | 0·0791 | 0·0789 | 0·0689 | 0·0092 | 0·0824 |
| 0 | 0·2507 | 0·2676 | 0·2513 | 0·2255 | 0·2755 | 0·0386 | 0·0278 | 0·0312 | 0·2755 |
| 0 | 0·1818 | 0·1676 | 0·1799 | 0·1884 | 0·1738 | 0·0482 | 0·0439 | 0·0378 | 0·1884 |
| 0 | 0·0483 | 0·0444 | 0·0477 | 0·0060 | 0·0025 | 0·0094 | 0·0055 | 0·0047 | 0·0483 |
| 0 | 0·0040 | 0·0047 | 0·0055 | 0·0068 | 0·0046 | 0·0021 | 0·0016 | 0·0051 | 0·0068 |
| $2×10^7$ | 0·1302 | 0·0912 | 0·0878 | 0·1040 | 0·1040 | 0·1079 | 0·1079 | 0·1450 | 0·1450 |
| $2×10^7$ | 0·0764 | 0·0837 | 0·0866 | 23·1134 | 12·3467 | 4·3538 | 2·2251 | 0·7230 | 23·1134 |
| $2×10^7$ | 0·0458 | 0·0412 | 0·0329 | 0·0632 | 0·0504 | 0·0390 | 0·0299 | 0·0378 | 0·0632 |
| $2×10^7$ | 0·0418 | 0·0450 | 0·0398 | 0·0416 | 0·0329 | 0·0412 | 0·0171 | 0·0112 | 0·0450 |
| $2×10^7$ | 0·0787 | 0·0764 | 0·0832 | 0·9016 | 0·4984 | 0·3909 | 0·4295 | 0·8024 | 0·9016 |
| $2×10^7$ | 0·0975 | 0·0796 | 0·0797 | 0·0682 | 0·0923 | 0·0857 | 0·1040 | 0·0207 | 0·1040 |
| $2×10^7$ | 0·0792 | | 0·0683 | 0·1018 | 0·1085 | 0·0951 | 0·0936 | | 0·1085 |
| $2×10^7$ | 0·0243 | 0·0199 | 0·0250 | 0·0364 | 0·0374 | 0·0095 | | 0·0055 | 0·0374 |
| $2×10^7$ | 0·1160 | 0·0981 | 0·0961 | 0·1058 | 0·0981 | 0·2887 | 0·3418 | 0·8353 | 0·8353 |
| $2×10^7$ | 0·1253 | 0·1253 | 0·1472 | 0·1523 | 0·0485 | 0·0716 | 0·1027 | 0·5494 | 0·5494 |
| $2×10^7$ | 0·0122 | 0·0089 | 0·0154 | 0·0230 | 0·0269 | 0·0356 | 0·0625 | 0·2420 | 0·2420 |
| $2×10^7$ | 0·0525 | 0·0504 | 0·0557 | 0·0580 | 0·0670 | 0·0694 | 0·0906 | 0·3351 | 0·3351 |
| $2×10^7$ | 0·0192 | 0·0231 | 0·0167 | 0·0257 | 0·0299 | 0·0279 | 0·0230 | 0·0249 | 0·0299 |
| $2×10^7$ | 0·0055 | 0·0047 | 0·0095 | 0·2504 | 0·2310 | 0·1352 | | 0·1527 | 0·2504 |
| $2×10^8$ | 0·0620 | 0·0594 | 0·0720 | 0·0946 | 0·1004 | 0·2894 | 0·8113 | 0·9170 | 0·9170 |
| $2×10^8$ | 0·0411 | 0·0379 | 0·0373 | 0·0293 | 0·0337 | 0·0360 | 0·0337 | 0·0362 | 0·0411 |
| $2×10^8$ | 0·0404 | 0·0400 | 0·0490 | 0·0629 | 0·0616 | 0·1773 | 0·6905 | 1·5288 | 1·5288 |
| $2×10^8$ | 0·1246 | 0·1342 | 0·1285 | 0·1217 | 0·1325 | 0·2392 | 0·2033 | 0·6327 | 0·6327 |
| $2×10^8$ | 0·0362 | 0·0217 | 0·0232 | 0·0500 | 0·0446 | 0·0021 | 0·0152 | 0·0391 | 0·0500 |
| $2×10^8$ | 0·0221 | 0·0258 | 0·0190 | 0·0245 | | | | | 0·0258 |
| $2×10^8$ | 0·0772 | 0·0852 | 0·0840 | 0·0931 | 0·0201 | 0·1163 | 0·2465 | 0·3705 | 0·3705 |
| $2×10^8$ | 0·0151 | 0·0247 | 0·0254 | 0·0125 | 0·0007 | | | 0·0004 | 0·0254 |
| $2×10^8$ | 0·0675 | 0·0481 | 0·0747 | 0·0678 | 0·0381 | 0·0319 | 0·0474 | 0·2701 | 0·2701 |
| $2×10^8$ | 0·0382 | 0·0457 | 0·0319 | 0·0405 | 0·0518 | 0·0441 | 0·0380 | 0·0448 | 0·0518 |
| $2×10^8$ | 0·0087 | 0·0076 | 0·0066 | 0·0142 | 0·0083 | 0·2790 | 0·6279 | 1·1848 | 1·1848 |
| $2×10^8$ | 0·0207 | 0·0218 | 0·0257 | 3·5286 | 2·3629 | | 2·0667 | 2·7097 | 3·5286 |
| $2×10^8$ | 0·0021 | 0·0011 | 0·0064 | 0·0099 | 0·0062 | 0·1304 | 0·1182 | 0·2579 | 0·2579 |

| | Fold change Inaba IgG | | | | | | | | |
|---|---|---|---|---|---|---|---|---|---|
| Dose (CFU) | D1 | D4 | D7 | D15 | D29 | D57 | D85 | D180 | max FC |
| 0 | 1·00 | 1·49 | 1·20 | 1·28 | 1·45 | 1·30 | 1·60 | 1·96 | 1·96 |
| 0 | 1·00 | 1·25 | 1·09 | 0·90 | 0·95 | 0·93 | 0·84 | 0·80 | 1·25 |
| 0 | 1·00 | 1·18 | 1·24 | 1·02 | 1·19 | 1·19 | 1·04 | 0·14 | 1·24 |
| 0 | 1·00 | 1·07 | 1·00 | 0·90 | 1·10 | 0·15 | 0·11 | 0·12 | 1·10 |
| 0 | 1·00 | 0·92 | 0·99 | 1·04 | 0·96 | 0·26 | 0·24 | 0·21 | 1·04 |
| 0 | 1·00 | 0·92 | 0·99 | 0·13 | 0·05 | 0·19 | 0·11 | 0·10 | 0·99 |
| 0 | 1·00 | 1·19 | 1·37 | 1·69 | 1·14 | 0·53 | 0·40 | 1·28 | 1·69 |
| $2×10^7$ | 1·00 | 0·70 | 0·67 | 0·80 | 0·80 | 0·83 | 0·83 | 1·11 | 1·11 |
| $2×10^7$ | 1·00 | 1·10 | 1·13 | 302·55 | 161·62 | 56·99 | 29·13 | 9·46 | 302·55 |
| $2×10^7$ | 1·00 | 0·90 | 0·72 | 1·38 | 1·10 | 0·85 | 0·65 | 0·83 | 1·38 |
| $2×10^7$ | 1·00 | 1·08 | 0·95 | 1·00 | 0·79 | 0·99 | 0·41 | 0·27 | 1·08 |
| $2×10^7$ | 1·00 | 0·97 | 1·06 | 11·45 | 6·33 | 4·96 | 5·45 | 10·19 | 11·45 |
| $2×10^7$ | 1·00 | 0·82 | 0·82 | 0·70 | 0·95 | 0·88 | 1·07 | 0·21 | 1·07 |
| $2×10^7$ | 1·00 | | 0·86 | 1·28 | 1·37 | 1·20 | 1·18 | | 1·37 |
| $2×10^7$ | 1·00 | 0·82 | 1·03 | 1·50 | 1·54 | 0·39 | | 0·23 | 1·54 |
| $2×10^7$ | 1·00 | 0·85 | 0·83 | 0·91 | 0·85 | 2·49 | 2·95 | 7·20 | 7·20 |
| $2×10^7$ | 1·00 | 1·00 | 1·17 | 1·21 | 0·39 | 0·57 | 0·82 | 4·38 | 4·38 |
| $2×10^7$ | 1·00 | 0·73 | 1·27 | 1·89 | 2·21 | 2·93 | 5·14 | 19·89 | 19·89 |
| $2×10^7$ | 1·00 | 0·96 | 1·06 | 1·11 | 1·28 | 1·32 | 1·73 | 6·39 | 6·39 |
| $2×10^7$ | 1·00 | 1·20 | 0·87 | 1·34 | 1·55 | 1·45 | 1·19 | 1·29 | 1·55 |
| $2×10^7$ | 1·00 | 0·87 | 1·74 | 45·65 | 42·11 | 24·65 | | 27·83 | 45·65 |
| $2×10^8$ | 1·00 | 0·96 | 1·16 | 1·53 | 1·62 | 4·67 | 13·09 | 14·80 | 14·80 |
| $2×10^8$ | 1·00 | 0·92 | 0·91 | 0·71 | 0·82 | 0·88 | 0·82 | 0·88 | 0·92 |
| $2×10^8$ | 1·00 | 0·99 | 1·21 | 1·55 | 1·52 | 4·38 | 17·08 | 37·81 | 37·81 |
| $2×10^8$ | 1·00 | 1·08 | 1·03 | 0·98 | 1·06 | 1·92 | 1·63 | 5·08 | 5·08 |
| $2×10^8$ | 1·00 | 0·60 | 0·64 | 1·38 | 1·23 | 0·06 | 0·42 | 1·08 | 1·38 |
| $2×10^8$ | 1·00 | 1·17 | 0·86 | 1·11 | | | | | 1·17 |
| $2×10^8$ | 1·00 | 1·10 | 1·09 | 1·20 | 0·26 | 1·51 | 3·19 | 4·80 | 4·80 |
| $2×10^8$ | 1·00 | 1·64 | 1·69 | 0·83 | 0·05 | | | 0·03 | 1·69 |
| $2×10^8$ | 1·00 | 0·71 | 1·11 | 1·01 | 0·56 | 0·47 | 0·70 | 4·00 | 4·00 |
| $2×10^8$ | 1·00 | 1·20 | 0·84 | 1·06 | 1·36 | 1·16 | 0·99 | 1·17 | 1·36 |
| $2×10^8$ | 1·00 | 0·88 | 0·76 | 1·63 | 0·96 | 32·11 | 72·27 | 136·36 | 136·36 |
| $2×10^8$ | 1·00 | 1·05 | 1·24 | 170·19 | 113·97 | | 99·68 | 130·69 | 170·19 |
| $2×10^8$ | 1·00 | 0·50 | 3·01 | 4·64 | 2·92 | 61·29 | 55·58 | 121·20 | 121·20 |

below limit of detection

| | RAU | | |
|---|---|---|---|
| | D1 | D39 | |
| *V. cholerae* | 0·0455 | 0·2975 | |
| | 0·0376 | 0·7930 | |
| | 0·1779 | 0·3023 | |
| | 0·1150 | 2·1244 | |
| | 0·0455 | 1·8440 | |
| | D1 | D11 | |
| Vaxchora D11 Challenge | 0·1321 | 0·3282 | |
| | 0·1684 | 0·1647 | |
| | 0·5171 | 0·7018 | |
| | 0·0921 | 0·1038 | |
| | 0·0909 | 0·3974 | |
| | 0·0759 | 0·4323 | |
| | D1 | D11 | D91 |
| Vaxchora D91 Challenge | 0·0348 | 0·0634 | 0·0480 |
| | 0·1944 | 0·3842 | 0·1990 |
| | 0·0896 | 0·1679 | 0·1421 |
| | 0·2020 | 0·2733 | 0·2222 |
| | 0·0627 | 0·1877 | 0·1080 |
| | 0·0560 | 0·1443 | 0·0940 |

| | RAU fold change | | | max FC |
|---|---|---|---|---|
| | D1 | D39 | | |
| *V. cholerae* | 1·00 | 6·53 | | 6·53 |
| | 1·00 | 21·09 | | 21·09 |
| | 1·00 | 1·70 | | 1·70 |
| | 1·00 | 18·47 | | 18·47 |
| | 1·00 | 40·49 | | 40·49 |
| | D1 | D11 | | |
| Vaxchora D11 Challenge | 1·00 | 2·48 | | 2·48 |
| | 1·00 | 0·98 | | 0·98 |
| | 1·00 | 1·36 | | 1·36 |
| | 1·00 | 1·13 | | 1·13 |
| | 1·00 | 4·37 | | 4·37 |
| | 1·00 | 5·69 | | 5·69 |
| | D1 | D11 | D91 | |
| Vaxchora D91 Challenge | 1·00 | 1·82 | 1·38 | 1·82 |
| | 1·00 | 1·98 | 1·02 | 1·98 |
| | 1·00 | 1·87 | 1·59 | 1·87 |
| | 1·00 | 1·35 | 1·10 | 1·35 |
| | 1·00 | 3·00 | 1·72 | 3·00 |
| | 1·00 | 2·58 | 1·68 | 2·58 |

**Dose-Escalation Module**

| Inaba IgA (RAU) | | | | | | | | | |
|---|---|---|---|---|---|---|---|---|---|
| **Dose (CFU)** | **D1** | **D4** | **D7** | **D15** | **D29** | **D57** | **D85** | **D180** | **max** |
| $10^5$ | 0·0017 | 0·0042 | 0·0087 | 0·0129 | 0·0130 | 0·0143 | 0·0128 | 0·0106 | 0·0143 |
| $10^5$ | 0·0175 | 0·0139 | 0·0190 | 0·0196 | 0·0191 | 0·0186 | 0·0193 | | 0·0196 |
| $10^5$ | 0·0140 | 0·0167 | 0·0194 | 0·4121 | 0·1029 | 0·0513 | 0·0761 | | 0·4121 |
| $10^6$ | 0·0013 | 0·0025 | 0·0111 | 2·5844 | 0·1975 | 0·0918 | 0·0783 | | 2·5844 |
| $10^6$ | 0·0191 | 0·0208 | 0·1421 | 1·1625 | 0·2677 | 0·1284 | 0·1007 | 0·0819 | 1·1625 |
| $10^6$ | 0·0017 | 0·0035 | 0·0047 | 2·0900 | 0·3196 | 0·0641 | 0·0184 | 0·0145 | 2·0900 |
| $10^7$ | 0·0044 | 0·0036 | 0·0035 | 0·0058 | 0·0119 | 0·0157 | 0·0223 | 0·0231 | 0·0231 |
| $10^7$ | 0·0035 | 0·0034 | 0·0113 | 0·0153 | 0·0120 | 0·0078 | 0·0036 | 0·0005 | 0·0153 |
| $10^7$ | 0·0096 | 0·0090 | 0·0067 | 0·1184 | 0·0181 | 0·0071 | 0·0082 | 0·0009 | 0·1184 |
| $10^8$ | 0·0032 | 0·0032 | 0·0036 | 0·0111 | 0·0047 | 0·0077 | 0·0097 | 0·0206 | 0·0206 |
| $10^8$ | 0·0090 | 0·0093 | 0·0058 | 0·0321 | 0·0128 | 0·0110 | 0·0352 | 0·0462 | 0·0462 |
| $10^8$ | 0·0043 | 0·0028 | 0·1524 | 1·0628 | 0·1708 | 0·0417 | 0·0482 | 0·0480 | 1·0628 |
| $10^9$ | 0·0073 | 0·0055 | 0·0219 | 0·3534 | 0·1117 | 0·0405 | 0·0177 | 0·0136 | 0·3534 |
| $10^9$ | 0·0241 | 0·0228 | 0·0268 | 0·0271 | 0·1569 | 0·1230 | 0·2330 | | 0·2330 |
| $10^9$ | 0·0059 | 0·0052 | 0·0067 | 0·0107 | 0·0125 | 0·0115 | 0·0125 | 0·0193 | 0·0193 |
| $10^{10}$ | 0·0006 | 0·0003 | 0·0022 | 0·0066 | 0·0030 | | 0·0027 | 0·0028 | 0·0066 |
| $10^{10}$ | 0·0075 | 0·0083 | 0·0319 | 0·2563 | | | 0·1147 | 0·0367 | 0·2563 |
| $10^{10}$ | 0·0008 | 0·0005 | 0·0403 | 1·1487 | 0·1491 | 0·0439 | 0·0373 | 0·0443 | 1·1487 |

| Fold change Inaba IgA | | | | | | | | | |
|---|---|---|---|---|---|---|---|---|---|
| **Dose (CFU)** | **D1** | **D4** | **D7** | **D15** | **D29** | **D57** | **D85** | **D180** | **max FC** |
| $10^5$ | 1·00 | 2·49 | 5·14 | 7·61 | 7·62 | 8·38 | 7·52 | 6·21 | 8·38 |
| $10^5$ | 1·00 | 0·79 | 1·08 | 1·12 | 1·09 | 1·06 | 1·10 | | 1·12 |
| $10^5$ | 1·00 | 1·19 | 1·38 | 29·36 | 7·33 | 3·66 | 5·42 | | 29·36 |
| $10^6$ | 1·00 | 1·95 | 8·60 | 1999·18 | 152·81 | 71·01 | 60·58 | | 1999·18 |
| $10^6$ | 1·00 | 1·09 | 7·45 | 60·95 | 14·03 | 6·73 | 5·28 | 4·29 | 60·95 |
| $10^6$ | 1·00 | 2·05 | 2·77 | 1240·03 | 189·62 | 38·05 | 10·93 | 8·61 | 1240·03 |
| $10^7$ | 1·00 | 0·83 | 0·78 | 1·30 | 2·69 | 3·55 | 5·05 | 5·22 | 5·22 |
| $10^7$ | 1·00 | 0·98 | 3·26 | 4·43 | 3·47 | 2·25 | 1·05 | 0·14 | 4·43 |
| $10^7$ | 1·00 | 0·93 | 0·69 | 12·30 | 1·88 | 0·74 | 0·85 | 0·10 | 12·30 |
| $10^8$ | 1·00 | 1·00 | 1·12 | 3·46 | 1·48 | 2·42 | 3·02 | 6·44 | 6·44 |
| $10^8$ | 1·00 | 1·04 | 0·65 | 3·58 | 1·43 | 1·23 | 3·93 | 5·15 | 5·15 |
| $10^8$ | 1·00 | 0·66 | 35·80 | 249·75 | 40·15 | 9·80 | 11·33 | 11·28 | 249·75 |
| $10^9$ | 1·00 | 0·75 | 2·99 | 48·36 | 15·28 | 5·54 | 2·42 | 1·86 | 48·36 |
| $10^9$ | 1·00 | 0·95 | 1·11 | 1·12 | 6·51 | 5·11 | 9·68 | | 9·68 |
| $10^9$ | 1·00 | 0·88 | 1·13 | 1·82 | 2·12 | 1·94 | 2·12 | 3·26 | 3·26 |
| $10^{10}$ | 1·00 | 0·45 | 3·57 | 10·80 | 4·87 | | 4·38 | 4·59 | 10·80 |
| $10^{10}$ | 1·00 | 1·11 | 4·28 | 34·35 | | | 15·38 | 4·92 | 34·35 |
| $10^{10}$ | 1·00 | 0·59 | 49·77 | 1419·07 | 184·15 | 54·20 | 46·14 | 54·76 | 1419·07 |

**Dose- Expansion Module**

| Inaba IgA (RAU) | | | | | | | | | |
|---|---|---|---|---|---|---|---|---|---|
| **Dose (CFU)** | **D1** | **D4** | **D7** | **D15** | **D29** | **D57** | **D85** | **D180** | **max** |
| 0 | 0·0168 | 0·0216 | 0·0216 | 0·0181 | 0·0216 | 0·0180 | 0·0221 | 0·0139 | 0·0221 |
| 0 | 0·0082 | 0·0033 | 0·0118 | 0·0059 | 0·0083 | 0·0052 | 0·0116 | 0·0088 | 0·0118 |
| 0 | 0·0088 | 0·0142 | 0·0042 | 0·0071 | 0·0124 | 0·0098 | 0·0108 | 0·0047 | 0·0142 |
| 0 | 0·0795 | 0·0644 | 0·0818 | 0·0584 | 0·0395 | 0·0107 | 0·0085 | 0·0069 | 0·0818 |
| 0 | 0·0049 | 0·0041 | 0·0111 | 0·0077 | 0·0094 | 0·0065 | 0·0057 | 0·0050 | 0·0111 |
| 0 | 0·0082 | 0·0142 | 0·0080 | 0·0050 | 0·0052 | 0·0054 | 0·0055 | 0·0043 | 0·0142 |
| 0 | 0·0095 | 0·0073 | 0·0093 | 0·0081 | 0·0142 | 0·0077 | 0·0028 | 0·0071 | 0·0142 |
| $2\times10^7$ | 0·0097 | 0·0152 | 0·0281 | 0·1799 | 0·0504 | 0·0448 | 0·0218 | 0·0154 | 0·1799 |
| $2\times10^7$ | 0·0104 | 0·0136 | 0·0384 | 40·0331 | 8·0326 | 0·7279 | 0·4657 | 0·2318 | 40·0331 |
| $2\times10^7$ | 0·0265 | 0·0265 | 0·1081 | 0·2800 | 0·1750 | 0·0802 | 0·0552 | 0·0322 | 0·2800 |
| $2\times10^7$ | 0·0238 | 0·0195 | 0·0218 | 0·0144 | 0·0193 | 0·0295 | 0·0201 | 0·0080 | 0·0295 |
| $2\times10^7$ | 0·0189 | 0·0186 | 0·0289 | 3·2011 | 0·3853 | 0·2678 | 0·3879 | 0·3264 | 3·2011 |
| $2\times10^7$ | 0·0191 | 0·0195 | 0·0311 | 0·3841 | 0·1760 | 0·0554 | 0·0458 | 0·0238 | 0·3841 |
| $2\times10^7$ | 0·0368 | | 0·0244 | 0·0315 | 0·0404 | 0·0308 | 0·0196 | | 0·0404 |
| $2\times10^7$ | 0·0121 | 0·0214 | 0·0243 | 0·0884 | 0·0831 | 0·0326 | 0·0224 | 0·0097 | 0·0884 |
| $2\times10^7$ | 0·0195 | 0·0237 | 0·0392 | 0·1994 | 0·1552 | 0·0686 | 0·0632 | 0·0543 | 0·1994 |
| $2\times10^7$ | 0·0242 | 0·0295 | 0·0356 | 2·4514 | 0·1111 | 0·0458 | 0·0363 | 0·0226 | 2·4514 |
| $2\times10^7$ | 0·0067 | 0·0066 | 0·0283 | 0·3527 | 0·2621 | 0·1573 | 0·1492 | 0·1323 | 0·3527 |
| $2\times10^7$ | 0·0092 | 0·0106 | 0·0156 | 0·0133 | 0·0197 | 0·0291 | 0·0382 | 0·0624 | 0·0624 |
| $2\times10^7$ | 0·0028 | 0·0015 | 0·0236 | 0·2150 | 0·0751 | 0·0211 | 0·0125 | 0·0050 | 0·2150 |
| $2\times10^7$ | 0·0056 | 0·0048 | 0·0345 | 0·7314 | 0·2480 | 0·0788 | | 0·0346 | 0·7314 |
| $2\times10^8$ | 0·0342 | 0·0382 | 0·0710 | 0·1857 | 0·1686 | 0·1321 | 0·0872 | 0·0665 | 0·1857 |
| $2\times10^8$ | 0·0411 | 0·0500 | 0·0457 | 0·0543 | 0·0550 | 0·0523 | 0·0420 | 0·0243 | 0·0550 |
| $2\times10^8$ | 0·0079 | 0·0045 | 0·0138 | 0·9273 | 0·0891 | 0·0669 | 0·0599 | 0·0542 | 0·9273 |
| $2\times10^8$ | 0·0300 | 0·0358 | 0·0464 | 0·0716 | 0·0928 | 0·1226 | 0·0552 | 0·0590 | 0·1226 |
| $2\times10^8$ | 0·0049 | 0·0037 | 0·0074 | 0·0306 | 0·0197 | 0·0065 | 0·0047 | 0·0029 | 0·0306 |
| $2\times10^8$ | 0·0171 | 0·0160 | 0·0142 | 0·1651 | | | | | 0·1651 |
| $2\times10^8$ | 0·0067 | 0·0017 | 0·0034 | 0·0178 | 0·0110 | 0·0119 | 0·0141 | 0·0106 | 0·0178 |
| $2\times10^8$ | 0·0112 | 0·0047 | 0·0100 | 0·0097 | 0·0044 | 0·0059 | 0·0070 | 0·0073 | 0·0112 |
| $2\times10^8$ | 0·0743 | 0·0677 | 0·1973 | 0·6637 | 0·2397 | 0·0980 | 0·0731 | 0·0505 | 0·6637 |
| $2\times10^8$ | 0·0071 | 0·0113 | 0·0158 | 0·1213 | 0·0951 | 0·0520 | 0·0477 | 0·0315 | 0·1213 |
| $2\times10^8$ | 0·0033 | 0·0041 | 0·0047 | 0·0287 | 0·0165 | 0·0219 | 0·0172 | 0·0195 | 0·0287 |
| $2\times10^8$ | 0·0127 | 0·0247 | 0·0257 | 0·8704 | 0·1689 | | 0·0551 | 0·0531 | 0·8704 |
| $2\times10^8$ | 0·0042 | 0·0036 | 0·0082 | 0·0137 | 0·0111 | 0·0165 | 0·0205 | 0·0471 | 0·0471 |

| Fold change Inaba IgA | | | | | | | | | |
|---|---|---|---|---|---|---|---|---|---|
| **Dose (CFU)** | **D1** | **D4** | **D7** | **D15** | **D29** | **D57** | **D85** | **D180** | **max FC** |
| 0 | 1·00 | 1·29 | 1·29 | 1·08 | 1·29 | 1·07 | 1·32 | 0·83 | 1·32 |
| 0 | 1·00 | 0·40 | 1·45 | 0·72 | 1·01 | 0·63 | 1·42 | 1·07 | 1·45 |
| 0 | 1·00 | 1·60 | 0·48 | 0·80 | 1·40 | 1·11 | 1·22 | 0·53 | 1·60 |
| 0 | 1·00 | 0·81 | 1·03 | 0·73 | 0·50 | 0·13 | 0·11 | 0·09 | 1·03 |
| 0 | 1·00 | 0·85 | 2·29 | 1·58 | 1·94 | 1·34 | 1·18 | 1·04 | 2·29 |
| 0 | 1·00 | 1·74 | 0·98 | 0·61 | 0·64 | 0·66 | 0·67 | 0·53 | 1·74 |
| 0 | 1·00 | 0·76 | 0·98 | 0·85 | 1·49 | 0·81 | 0·30 | 0·75 | 1·49 |
| $2\times10^7$ | 1·00 | 1·57 | 2·90 | 18·56 | 5·20 | 4·62 | 2·24 | 1·58 | 18·56 |
| $2\times10^7$ | 1·00 | 1·31 | 3·70 | 3859·95 | 774·50 | 70·18 | 44·90 | 22·35 | 3859·95 |
| $2\times10^7$ | 1·00 | 1·00 | 4·08 | 10·58 | 6·61 | 3·03 | 2·08 | 1·22 | 10·58 |
| $2\times10^7$ | 1·00 | 0·82 | 0·92 | 0·60 | 0·81 | 1·24 | 0·85 | 0·34 | 1·24 |
| $2\times10^7$ | 1·00 | 0·99 | 1·53 | 169·62 | 20·42 | 14·19 | 20·56 | 17·30 | 169·62 |
| $2\times10^7$ | 1·00 | 1·02 | 1·62 | 20·09 | 9·20 | 2·90 | 2·39 | 1·25 | 20·09 |
| $2\times10^7$ | 1·00 | | 0·66 | 0·86 | 1·10 | 0·84 | 0·53 | | 1·10 |
| $2\times10^7$ | 1·00 | 1·78 | 2·01 | 7·32 | 6·89 | 2·70 | 1·86 | 0·80 | 7·32 |
| $2\times10^7$ | 1·00 | 1·22 | 2·02 | 10·25 | 7·98 | 3·52 | 3·25 | 2·79 | 10·25 |
| $2\times10^7$ | 1·00 | 1·22 | 1·47 | 101·27 | 4·59 | 1·89 | 1·50 | 0·93 | 101·27 |
| $2\times10^7$ | 1·00 | 0·99 | 4·25 | 52·97 | 39·37 | 23·63 | 22·41 | 19·87 | 52·97 |
| $2\times10^7$ | 1·00 | 1·15 | 1·70 | 1·44 | 2·14 | 3·16 | 4·14 | 6·78 | 6·78 |
| $2\times10^7$ | 1·00 | 0·55 | 8·38 | 76·18 | 26·61 | 7·49 | 4·42 | 1·77 | 76·18 |
| $2\times10^7$ | 1·00 | 0·85 | 6·12 | 129·71 | 43·99 | 13·98 | | 6·13 | 129·71 |
| $2\times10^8$ | 1·00 | 1·12 | 2·08 | 5·43 | 4·93 | 3·86 | 2·55 | 1·95 | 5·43 |
| $2\times10^8$ | 1·00 | 1·22 | 1·11 | 1·32 | 1·34 | 1·27 | 1·02 | 0·59 | 1·34 |
| $2\times10^8$ | 1·00 | 0·57 | 1·75 | 117·10 | 11·25 | 8·45 | 7·56 | 6·85 | 117·10 |
| $2\times10^8$ | 1·00 | 1·19 | 1·55 | 2·38 | 3·09 | 4·08 | 1·84 | 1·96 | 4·08 |
| $2\times10^8$ | 1·00 | 0·75 | 1·53 | 6·30 | 4·06 | 1·33 | 0·97 | 0·60 | 6·30 |
| $2\times10^8$ | 1·00 | 0·94 | 0·83 | 9·64 | | | | | 9·64 |
| $2\times10^8$ | 1·00 | 0·25 | 0·51 | 2·64 | 1·63 | 1·76 | 2·09 | 1·57 | 2·64 |
| $2\times10^8$ | 1·00 | 0·42 | 0·89 | 0·86 | 0·39 | 0·53 | 0·62 | 0·65 | 0·89 |
| $2\times10^8$ | 1·00 | 0·91 | 2·66 | 8·93 | 3·23 | 1·32 | 0·98 | 0·68 | 8·93 |
| $2\times10^8$ | 1·00 | 1·57 | 2·21 | 16·98 | 13·31 | 7·28 | 6·68 | 4·41 | 16·98 |
| $2\times10^8$ | 1·00 | 1·24 | 1·43 | 8·66 | 4·98 | 6·60 | 5·17 | 5·87 | 8·66 |
| $2\times10^8$ | 1·00 | 1·94 | 2·02 | 68·60 | 13·31 | | 4·34 | 4·18 | 68·60 |
| $2\times10^8$ | 1·00 | 0·85 | 1·95 | 3·27 | 2·63 | 3·93 | 4·87 | 11·20 | 11·20 |

| | **RAU** | | |
|---|---|---|---|
| | D1 | D39 | |
| *V. cholerae* | 0·0082 | 14·2989 | |
| | 0·0110 | 0·6328 | |
| | 0·0375 | 0·0401 | |
| | 0·0309 | 3·9626 | |
| | 0·0288 | 7·4963 | |
| | D1 | D11 | |
| Vaxchora D11 Challenge | 0·0130 | 0·1043 | |
| | 0·0580 | 0·0587 | |
| | 0·1280 | 1·7380 | |
| | 0·0130 | 0·0212 | |
| | 0·0159 | 0·5441 | |
| | 0·0209 | 0·5283 | |
| | D1 | D11 | D91 |
| Vaxchora D91 Challenge | 0·0019 | 0·0025 | 0·0034 |
| | 0·0064 | 0·6443 | 0·0206 |
| | 0·0031 | 0·1453 | 0·0080 |
| | 0·0262 | 0·1789 | 0·0357 |
| | 0·0113 | 0·2759 | 0·0466 |
| | 0·0046 | 0·0476 | 0·0186 |

| | **RAU fold change** | | | **max FC** |
|---|---|---|---|---|
| | D1 | D39 | | |
| *V. cholerae* | 1·00 | 1750·61 | | 1750·61 |
| | 1·00 | 57·63 | | 57·63 |
| | 1·00 | 1·07 | | 1·07 |
| | 1·00 | 128·30 | | 128·30 |
| | 1·00 | 260·21 | | 260·21 |
| | D1 | D11 | | |
| Vaxchora D11 Challenge | 1·00 | 8·01 | | 8·01 |
| | 1·00 | 1·01 | | 1·01 |
| | 1·00 | 13·58 | | 13·58 |
| | 1·00 | 1·63 | | 1·63 |
| | 1·00 | 34·29 | | 34·29 |
| | 1·00 | 25·32 | | 25·32 |
| | D1 | D11 | D91 | |
| Vaxchora D91 Challenge | 1·00 | 1·29 | 1·74 | 1·74 |
| | 1·00 | 99·92 | 3·20 | 99·92 |
| | 1·00 | 46·30 | 2·56 | 46·30 |
| | 1·00 | 6·83 | 1·36 | 6·83 |
| | 1·00 | 24·46 | 4·13 | 24·46 |
| | 1·00 | 10·25 | 4·01 | 10·25 |

**Dose-Escalation Module**

| Ogawa IgM (RAU) | | | | | | | | | |
|---|---|---|---|---|---|---|---|---|---|
| Dose (CFU) | D1 | D4 | D7 | D15 | D29 | D57 | D85 | D180 | max |
| $10^5$ | 0·0333 | 0·0166 | 0·0468 | 0·4103 | 0·2014 | 0·1370 | 0·1197 | 0·1685 | 0·4103 |
| $10^5$ | 0·0079 | 0·0053 | 0·0039 | 0·0253 | 0·0345 | 0·0236 | 0·0244 | | 0·0345 |
| $10^5$ | 0·1259 | 0·1316 | 0·1319 | 0·1754 | 0·1359 | 0·0987 | 0·1069 | | 0·1754 |
| $10^6$ | 0·1044 | 0·0873 | 0·5409 | 5·6475 | 2·6043 | 0·8547 | 0·5172 | | 5·6475 |
| $10^6$ | 0·0076 | 0·0101 | 0·0146 | 0·0554 | 0·0366 | 0·0262 | 0·0176 | 0·0118 | 0·0554 |
| $10^6$ | 0·0118 | 0·0099 | 0·1641 | 11·3709 | 3·0592 | 1·2220 | 0·9650 | 0·2275 | 11·3709 |
| $10^7$ | 0·0223 | 0·0188 | 0·1992 | 5·0226 | 2·3810 | 1·3490 | 0·7292 | 0·2145 | 5·0226 |
| $10^7$ | 0·0162 | 0·0169 | 0·1614 | 2·8529 | 2·1542 | 0·8579 | 0·2989 | 0·0738 | 2·8529 |
| $10^7$ | 0·0303 | 0·0357 | 0·0286 | 0·1678 | 0·0812 | 0·0567 | 0·0516 | 0·0404 | 0·1678 |
| $10^8$ | 0·0108 | 0·0106 | 0·0247 | 1·1431 | 0·6306 | 0·2353 | 0·1882 | 0·0874 | 1·1431 |
| $10^8$ | 0·0210 | 0·0167 | 0·0389 | 1·1895 | 0·3692 | 0·1398 | 0·0621 | 0·0468 | 1·1895 |
| $10^8$ | 0·0105 | 0·0097 | 0·0278 | 0·2695 | 0·1200 | 0·0312 | 0·0247 | 0·0433 | 0·2695 |
| $10^9$ | 0·0046 | 0·0014 | 0·2686 | 2·0038 | 0·5962 | 0·1294 | 0·0429 | 0·0289 | 2·0038 |
| $10^9$ | 0·0121 | 0·0103 | 0·0047 | 0·0218 | 0·0600 | 0·0707 | 0·1580 | | 0·1580 |
| $10^9$ | 0·0414 | 0·0428 | 0·0558 | 1·0099 | 0·2168 | 0·1780 | 0·2207 | 0·5674 | 1·0099 |
| $10^{10}$ | 0·0149 | 0·0185 | 0·0305 | 0·0897 | 0·0726 | | 0·0350 | 0·0397 | 0·0897 |
| $10^{10}$ | 0·0903 | 0·0963 | 0·3222 | 1·4568 | | | 0·1764 | 0·0956 | 1·4568 |
| $10^{10}$ | 0·0277 | 0·0272 | 3·7863 | 10·4964 | 2·6434 | 0·9112 | 0·2412 | 0·1330 | 10·4964 |

| Fold change Ogawa IgM | | | | | | | | | |
|---|---|---|---|---|---|---|---|---|---|
| Dose (CFU) | D1 | D4 | D7 | D15 | D29 | D57 | D85 | D180 | max FC |
| $10^5$ | 1·00 | 0·50 | 1·41 | 12·33 | 6·05 | 4·12 | 3·60 | 5·06 | 12·33 |
| $10^5$ | 1·00 | 0·66 | 0·49 | 3·18 | 4·34 | 2·97 | 3·07 | | 4·34 |
| $10^5$ | 1·00 | 1·05 | 1·05 | 1·39 | 1·08 | 0·78 | 0·85 | | 1·39 |
| $10^6$ | 1·00 | 0·84 | 5·18 | 54·09 | 24·94 | 8·19 | 4·95 | | 54·09 |
| $10^6$ | 1·00 | 1·32 | 1·91 | 7·25 | 4·79 | 3·43 | 2·30 | 1·55 | 7·25 |
| $10^6$ | 1·00 | 0·83 | 13·85 | 959·72 | 258·21 | 103·14 | 81·45 | 19·20 | 959·72 |
| $10^7$ | 1·00 | 0·84 | 8·92 | 224·79 | 106·56 | 60·38 | 32·63 | 9·60 | 224·79 |
| $10^7$ | 1·00 | 1·04 | 9·99 | 176·59 | 133·34 | 53·10 | 18·50 | 4·57 | 176·59 |
| $10^7$ | 1·00 | 1·18 | 0·95 | 5·55 | 2·68 | 1·87 | 1·71 | 1·33 | 5·55 |
| $10^8$ | 1·00 | 0·99 | 2·29 | 106·22 | 58·59 | 21·86 | 17·48 | 8·12 | 106·22 |
| $10^8$ | 1·00 | 0·79 | 1·85 | 56·77 | 17·62 | 6·67 | 2·97 | 2·23 | 56·77 |
| $10^8$ | 1·00 | 0·93 | 2·65 | 25·74 | 11·46 | 2·98 | 2·36 | 4·13 | 25·74 |
| $10^9$ | 1·00 | 0·30 | 58·98 | 440·01 | 130·91 | 28·41 | 9·42 | 6·34 | 440·01 |
| $10^9$ | 1·00 | 0·85 | 0·39 | 1·81 | 4·98 | 5·86 | 13·10 | | 13·10 |
| $10^9$ | 1·00 | 1·03 | 1·35 | 24·39 | 5·24 | 4·30 | 5·33 | 13·70 | 24·39 |
| $10^{10}$ | 1·00 | 1·24 | 2·04 | 6·01 | 4·86 | | 2·34 | 2·66 | 6·01 |
| $10^{10}$ | 1·00 | 1·07 | 3·57 | 16·14 | | | 1·95 | 1·06 | 16·14 |
| $10^{10}$ | 1·00 | 0·98 | 136·74 | 379·08 | 95·47 | 32·91 | 8·71 | 4·80 | 379·08 |

**Dose- Expansion Module**

| Ogawa IgM (RAU) | | | | | | | | | |
|---|---|---|---|---|---|---|---|---|---|
| Dose (CFU) | D1 | D4 | D7 | D15 | D29 | D57 | D85 | D180 | max |
| 0 | 0·0393 | 0·0358 | 0·0348 | 0·0367 | 0·0377 | 0·0342 | 0·0435 | 0·0471 | 0·0471 |
| 0 | 0·0088 | 0·0091 | 0·0086 | 0·0105 | 0·0061 | 0·0083 | 0·0059 | 0·0071 | 0·0105 |
| 0 | 0·0719 | 0·0775 | 0·0804 | 0·0742 | 0·0839 | 0·0658 | 0·0437 | 0·0696 | 0·0839 |
| 0 | 0·1632 | 0·1702 | 0·1805 | 0·1455 | 0·1612 | 0·3393 | 0·1692 | 0·1141 | 0·3393 |
| 0 | 0·0540 | 0·0500 | 0·0668 | 0·0635 | 0·0683 | 0·0289 | 0·0343 | 0·0208 | 0·0683 |
| 0 | 0·0922 | 0·1003 | 0·1033 | 0·0557 | 0·0377 | 0·0592 | 0·0632 | 0·0818 | 0·1033 |
| 0 | 0·0906 | 0·0742 | 0·1104 | 0·1006 | 0·1381 | 0·1450 | 0·0575 | 0·1512 | 0·1512 |
| $2\times10^7$ | 0·1768 | 0·2049 | 0·2108 | 0·3254 | 0·2775 | 0·1785 | 0·2517 | 0·2646 | 0·3254 |
| $2\times10^7$ | 0·1969 | 0·2122 | 0·9912 | 27·7280 | 16·6929 | 6·0782 | 2·9698 | 1·0815 | 27·7280 |
| $2\times10^7$ | 0·0432 | 0·0467 | 0·5574 | 3·7278 | 2·1210 | 0·7035 | 0·2760 | 0·1294 | 3·7278 |
| $2\times10^7$ | 0·0437 | 0·0644 | 0·0617 | 0·2062 | 0·0742 | 0·0659 | 0·0589 | 0·0152 | 0·2062 |
| $2\times10^7$ | 0·0403 | 0·0353 | 0·2376 | 2·7542 | 1·0344 | 0·2921 | 0·1780 | 0·0335 | 2·7542 |
| $2\times10^7$ | 0·0172 | 0·0164 | 0·2717 | 1·6725 | 1·0500 | 0·2376 | 0·1750 | 0·0246 | 1·6725 |
| $2\times10^7$ | 0·1087 | | 0·1302 | 0·6912 | 0·2820 | 0·1763 | 0·1095 | | 0·6912 |
| $2\times10^7$ | 0·0959 | 0·1022 | 0·2270 | 16·7601 | 10·6496 | 4·9654 | 2·6575 | 0·5372 | 16·7601 |
| $2\times10^7$ | 0·0022 | 0·0051 | 0·0032 | 0·0154 | 0·0114 | 0·0082 | 0·0085 | 0·0032 | 0·0154 |
| $2\times10^7$ | 0·0858 | 0·1065 | 1·3789 | 24·7046 | 7·7335 | 1·8281 | 1·1008 | 0·6928 | 24·7046 |
| $2\times10^7$ | 0·0331 | 0·0444 | 0·1095 | 0·8820 | 0·8879 | 0·4942 | 0·2892 | 0·2063 | 0·8879 |
| $2\times10^7$ | 0·0156 | 0·0164 | 0·0286 | 0·2562 | 0·2084 | 0·2080 | 0·2200 | 0·1866 | 0·2562 |
| $2\times10^7$ | 0·0189 | 0·0166 | 0·0284 | 0·0553 | 0·0299 | 0·0231 | 0·0218 | 0·0261 | 0·0553 |
| $2\times10^7$ | 0·0386 | 0·0231 | 0·2341 | 4·3560 | 3·6247 | 1·5172 | | 0·3788 | 4·3560 |
| $2\times10^8$ | 0·0566 | 0·0566 | 0·0772 | 0·2420 | 0·2532 | 0·1062 | 0·0787 | 0·0577 | 0·2532 |
| $2\times10^8$ | 0·0153 | 0·0151 | 0·0294 | 0·5252 | 0·2339 | 0·0668 | 0·0481 | 0·0203 | 0·5252 |
| $2\times10^8$ | 0·0376 | 0·0315 | 0·1289 | 1·1659 | 0·2460 | 0·1327 | 0·0897 | 0·0267 | 1·1659 |
| $2\times10^8$ | 0·2280 | 0·2315 | 0·5721 | 1·1660 | 1·1261 | 0·9070 | 1·1108 | 1·0286 | 1·1660 |
| $2\times10^8$ | 0·0325 | 0·0395 | 0·1203 | 0·6408 | 0·4840 | 0·0893 | 0·0596 | 0·0434 | 0·6408 |
| $2\times10^8$ | 0·0538 | 0·0548 | 0·1047 | 3·6779 | | | | | 3·6779 |
| $2\times10^8$ | 0·0769 | 0·0931 | 0·1095 | 1·0439 | 0·6109 | 0·2945 | 0·2604 | 0·2080 | 1·0439 |
| $2\times10^8$ | 0·0069 | 0·0077 | 0·0063 | 0·2231 | 0·0679 | 0·0151 | 0·0140 | 0·0057 | 0·2231 |
| $2\times10^8$ | 0·0331 | 0·0269 | 1·5740 | 42·8939 | 10·5323 | 2·3211 | 1·5355 | 0·7635 | 42·8939 |
| $2\times10^8$ | 0·1143 | 0·1262 | 1·7239 | 16·8965 | 10·2668 | 2·4002 | 0·9532 | 0·3970 | 16·8965 |
| $2\times10^8$ | 0·0095 | 0·0134 | 0·5602 | 56·7692 | 20·5979 | 2·6296 | 0·7343 | 0·0936 | 56·7692 |
| $2\times10^8$ | 0·0105 | 0·0118 | 0·2336 | 1·6031 | 0·6253 | | 0·2294 | 0·1994 | 1·6031 |
| $2\times10^8$ | 0·0070 | 0·0029 | 0·0050 | 0·0103 | 0·0061 | 0·0135 | 0·0131 | 0·0156 | 0·0156 |

| Fold change Ogawa IgM | | | | | | | | | |
|---|---|---|---|---|---|---|---|---|---|
| Dose (CFU) | D1 | D4 | D7 | D15 | D29 | D57 | D85 | D180 | max FC |
| 0 | 1·00 | 0·91 | 0·89 | 0·93 | 0·96 | 0·87 | 1·11 | 1·20 | 1·20 |
| 0 | 1·00 | 1·04 | 0·98 | 1·19 | 0·70 | 0·94 | 0·67 | 0·80 | 1·19 |
| 0 | 1·00 | 1·08 | 1·12 | 1·03 | 1·17 | 0·92 | 0·61 | 0·97 | 1·17 |
| 0 | 1·00 | 1·04 | 1·11 | 0·89 | 0·99 | 2·08 | 1·04 | 0·70 | 2·08 |
| 0 | 1·00 | 0·93 | 1·24 | 1·18 | 1·27 | 0·54 | 0·63 | 0·39 | 1·27 |
| 0 | 1·00 | 1·09 | 1·12 | 0·60 | 0·41 | 0·64 | 0·69 | 0·89 | 1·12 |
| 0 | 1·00 | 0·82 | 1·22 | 1·11 | 1·52 | 1·60 | 0·63 | 1·67 | 1·67 |
| $2\times10^7$ | 1·00 | 1·16 | 1·19 | 1·84 | 1·57 | 1·01 | 1·42 | 1·50 | 1·84 |
| $2\times10^7$ | 1·00 | 1·08 | 5·03 | 140·81 | 84·77 | 30·87 | 15·08 | 5·49 | 140·81 |
| $2\times10^7$ | 1·00 | 1·08 | 12·91 | 86·33 | 49·12 | 16·29 | 6·39 | 3·00 | 86·33 |
| $2\times10^7$ | 1·00 | 1·47 | 1·41 | 4·71 | 1·70 | 1·51 | 1·35 | 0·35 | 4·71 |
| $2\times10^7$ | 1·00 | 0·88 | 5·90 | 68·40 | 25·69 | 7·25 | 4·42 | 0·83 | 68·40 |
| $2\times10^7$ | 1·00 | 0·96 | 15·84 | 97·50 | 61·21 | 13·85 | 10·20 | 1·44 | 97·50 |
| $2\times10^7$ | 1·00 | | 1·20 | 6·36 | 2·59 | 1·62 | 1·01 | | 6·36 |
| $2\times10^7$ | 1·00 | 1·07 | 2·37 | 174·80 | 111·07 | 51·79 | 27·72 | 5·60 | 174·80 |
| $2\times10^7$ | 1·00 | 2·30 | 1·43 | 6·99 | 5·20 | 3·74 | 3·86 | 1·43 | 6·99 |
| $2\times10^7$ | 1·00 | 1·24 | 16·07 | 287·88 | 90·12 | 21·30 | 12·83 | 8·07 | 287·88 |
| $2\times10^7$ | 1·00 | 1·34 | 3·31 | 26·63 | 26·80 | 14·92 | 8·73 | 6·23 | 26·80 |
| $2\times10^7$ | 1·00 | 1·05 | 1·83 | 16·41 | 13·34 | 13·32 | 14·09 | 11·95 | 16·41 |
| $2\times10^7$ | 1·00 | 0·88 | 1·50 | 2·93 | 1·59 | 1·22 | 1·15 | 1·38 | 2·93 |
| $2\times10^7$ | 1·00 | 0·60 | 6·07 | 112·93 | 93·97 | 39·33 | | 9·82 | 112·93 |
| $2\times10^8$ | 1·00 | 1·00 | 1·37 | 4·28 | 4·48 | 1·88 | 1·39 | 1·02 | 4·48 |
| $2\times10^8$ | 1·00 | 0·99 | 1·93 | 34·38 | 15·31 | 4·37 | 3·15 | 1·33 | 34·38 |
| $2\times10^8$ | 1·00 | 0·84 | 3·43 | 31·00 | 6·54 | 3·53 | 2·39 | 0·71 | 31·00 |
| $2\times10^8$ | 1·00 | 1·02 | 2·51 | 5·11 | 4·94 | 3·98 | 4·87 | 4·51 | 5·11 |
| $2\times10^8$ | 1·00 | 1·22 | 3·70 | 19·73 | 14·90 | 2·75 | 1·83 | 1·34 | 19·73 |
| $2\times10^8$ | 1·00 | 1·02 | 1·95 | 68·37 | | | | | 68·37 |
| $2\times10^8$ | 1·00 | 1·21 | 1·42 | 13·58 | 7·95 | 3·83 | 3·39 | 2·71 | 13·58 |
| $2\times10^8$ | 1·00 | 1·10 | 0·91 | 32·11 | 9·77 | 2·17 | 2·01 | 0·82 | 32·11 |
| $2\times10^8$ | 1·00 | 0·81 | 47·59 | 1297·03 | 318·48 | 70·19 | 46·43 | 23·09 | 1297·03 |
| $2\times10^8$ | 1·00 | 1·10 | 15·08 | 147·84 | 89·83 | 21·00 | 8·34 | 3·47 | 147·84 |
| $2\times10^8$ | 1·00 | 1·41 | 58·96 | 5974·49 | 2167·76 | 276·75 | 77·28 | 9·86 | 5974·49 |
| $2\times10^8$ | 1·00 | 1·12 | 22·29 | 152·97 | 59·67 | | 21·89 | 19·03 | 152·97 |
| $2\times10^8$ | 1·00 | 0·42 | 0·72 | 1·48 | 0·88 | 1·94 | 1·88 | 2·24 | 2·24 |

| | RAU | | |
|---|---|---|---|
| | D1 | D39 | |
| *V. cholerae* | 0·0088 | 0·5204 | |
| | 0·0349 | 5·2353 | |
| | 0·0079 | 0·0210 | |
| | 0·1538 | 20·3912 | |
| | 0·0378 | 1·0988 | |
| | D1 | D11 | |
| Vaxchora D11 Challenge | 0·0103 | 0·6372 | |
| | 0·0904 | 0·1051 | |
| | 0·0066 | 0·3332 | |
| | 0·0057 | 0·0140 | |
| | 0·0296 | 0·0386 | |
| | 0·1025 | 0·2432 | |
| | D1 | D11 | D91 |
| Vaxchora D91 Challenge | 0·0028 | 0·0696 | 0·0173 |
| | 0·0113 | 0·1024 | 0·0114 |
| | 0·0013 | 0·0799 | 0·0131 |
| | 0·0637 | 0·1315 | 0·1167 |
| | 0·0710 | 0·1048 | 0·0700 |
| | 0·0013 | 0·0202 | 0·0020 |

| | RAU fold change | | | max FC |
|---|---|---|---|---|
| | D1 | D39 | | |
| *V. cholerae* | 1·00 | 59·12 | | 59·12 |
| | 1·00 | 149·89 | | 149·89 |
| | 1·00 | 2·65 | | 2·65 |
| | 1·00 | 132·54 | | 132·54 |
| | 1·00 | 29·06 | | 29·06 |
| | D1 | D11 | | |
| Vaxchora D11 Challenge | 1·00 | 62·14 | | 62·14 |
| | 1·00 | 1·16 | | 1·16 |
| | 1·00 | 50·26 | | 50·26 |
| | 1·00 | 2·46 | | 2·46 |
| | 1·00 | 1·30 | | 1·30 |
| | 1·00 | 2·37 | | 2·37 |
| | D1 | D11 | D91 | |
| Vaxchora D91 Challenge | 1·00 | 25·20 | 6·26 | 25·20 |
| | 1·00 | 9·09 | 1·01 | 9·09 |
| | 1·00 | 60·81 | 9·95 | 60·81 |
| | 1·00 | 2·06 | 1·83 | 2·06 |
| | 1·00 | 1·48 | 0·99 | 1·48 |
| | 1·00 | 16·07 | 1·63 | 16·07 |

**Dose-Escalation Module**

| Ogawa IgG (RAU) | | | | | | | | | |
|---|---|---|---|---|---|---|---|---|---|
| **Dose (CFU)** | **D1** | **D4** | **D7** | **D15** | **D29** | **D57** | **D85** | **D180** | **max** |
| $10^5$ | 0·0064 | 0·0005 | 0·0019 | 0·0053 | 0·0060 | 0·0028 | 0·0039 | 0·0094 | 0·0094 |
| $10^5$ | 0·0001 | | 0·0035 | 0·0030 | 0·0042 | 0·0030 | 0·0057 | | 0·0057 |
| $10^5$ | 0·0032 | 0·0049 | 0·0054 | 0·0470 | 0·0259 | 0·0064 | 0·0133 | | 0·0470 |
| $10^6$ | 0·0028 | 0·0071 | 0·0078 | 0·0347 | 0·0216 | 0·0239 | 0·0278 | | 0·0347 |
| $10^6$ | 0·0105 | 0·0071 | 0·0028 | 0·0119 | 0·0035 | 0·0085 | 0·0057 | 0·0092 | 0·0119 |
| $10^6$ | 0·0211 | 0·0199 | 0·1323 | 50·8877 | 23·5763 | 8·6894 | 3·6844 | 0·8868 | 50·8877 |
| $10^7$ | 0·0163 | 0·0144 | 0·0131 | 0·0284 | 0·0294 | 0·0702 | 0·0931 | 0·1666 | 0·1666 |
| $10^7$ | 0·0057 | 0·0085 | 0·0092 | 0·0239 | 0·0205 | 0·0169 | 0·0289 | 0·0359 | 0·0359 |
| $10^7$ | 0·0169 | 0·0138 | 0·0181 | 0·0181 | 0·0157 | 0·0169 | 0·0125 | 0·0134 | 0·0181 |
| $10^8$ | 0·0131 | 0·0119 | 0·0105 | 0·0144 | 0·0138 | 0·0119 | 0·0251 | 0·0616 | 0·0616 |
| $10^8$ | 0·0078 | 0·0092 | 0·0105 | 0·0560 | 0·0239 | 0·0228 | 0·0156 | 0·0187 | 0·0560 |
| $10^8$ | 0·0150 | 0·0175 | 0·0216 | 0·0175 | 0·0157 | 0·0134 | 0·0470 | 0·1584 | 0·1584 |
| $10^9$ | 0·0085 | 0·0057 | 0·0092 | 0·0181 | 0·0131 | 0·0129 | 0·0171 | 0·0223 | 0·0223 |
| $10^9$ | 0·0071 | 0·0078 | 0·0112 | 0·0090 | 0·0396 | 0·0995 | 0·4516 | | 0·4516 |
| $10^9$ | 0·0156 | 0·0107 | 0·0156 | 0·0161 | 0·0101 | 0·0166 | 0·0156 | 0·0282 | 0·0282 |
| $10^{10}$ | 0·0043 | 0·0090 | 0·0766 | 1·4250 | 1·1861 | | 0·3381 | 0·1189 | 1·4250 |
| $10^{10}$ | 0·0025 | 0·0067 | 0·0084 | 0·0140 | | | 0·0049 | 0·0055 | 0·0140 |
| $10^{10}$ | 0·0134 | 0·0134 | 0·0084 | 0·0140 | 0·0129 | 0·0192 | 0·0182 | 0·0258 | 0·0258 |

| Fold change Ogawa IgG | | | | | | | | | |
|---|---|---|---|---|---|---|---|---|---|
| **Dose (CFU)** | **D1** | **D4** | **D7** | **D15** | **D29** | **D57** | **D85** | **D180** | **max FC** |
| $10^5$ | 1·00 | 0·08 | 0·30 | 0·83 | 0·94 | 0·43 | 0·61 | 1·47 | 1·47 |
| $10^5$ | 1·00 | | 66·62 | 58·41 | 80·25 | 58·41 | 109·76 | | 109·76 |
| $10^5$ | 1·00 | 1·53 | 1·71 | 14·77 | 8·14 | 2·01 | 4·18 | | 14·77 |
| $10^6$ | 1·00 | 2·58 | 2·83 | 12·57 | 7·84 | 8·67 | 10·07 | | 12·57 |
| $10^6$ | 1·00 | 0·68 | 0·26 | 1·13 | 0·33 | 0·81 | 0·54 | 0·87 | 1·13 |
| $10^6$ | 1·00 | 0·94 | 6·28 | 2415·90 | 1119·29 | 412·53 | 174·92 | 42·10 | 2415·90 |
| $10^7$ | 1·00 | 0·89 | 0·81 | 1·74 | 1·81 | 4·31 | 5·72 | 10·23 | 10·23 |
| $10^7$ | 1·00 | 1·49 | 1·61 | 4·21 | 3·60 | 2·97 | 5·08 | 6·31 | 6·31 |
| $10^7$ | 1·00 | 0·82 | 1·07 | 1·07 | 0·93 | 1·00 | 0·74 | 0·79 | 1·07 |
| $10^8$ | 1·00 | 0·90 | 0·80 | 1·10 | 1·05 | 0·90 | 1·91 | 4·69 | 4·69 |
| $10^8$ | 1·00 | 1·18 | 1·35 | 7·17 | 3·06 | 2·92 | 1·99 | 2·39 | 7·17 |
| $10^8$ | 1·00 | 1·16 | 1·44 | 1·16 | 1·04 | 0·89 | 3·12 | 10·53 | 10·53 |
| $10^9$ | 1·00 | 0·67 | 1·08 | 2·13 | 1·55 | 1·51 | 2·02 | 2·62 | 2·62 |
| $10^9$ | 1·00 | 1·10 | 1·57 | 1·26 | 5·57 | 13·99 | 63·49 | | 63·49 |
| $10^9$ | 1·00 | 0·69 | 1·00 | 1·03 | 0·65 | 1·07 | 1·00 | 1·81 | 1·81 |
| $10^{10}$ | 1·00 | 2·07 | 17·67 | 328·57 | 273·49 | | 77·96 | 27·42 | 328·57 |
| $10^{10}$ | 1·00 | 2·65 | 3·33 | 5·52 | | | 1·95 | 2·19 | 5·52 |
| $10^{10}$ | 1·00 | 1·00 | 0·63 | 1·04 | 0·96 | 1·43 | 1·36 | 1·92 | 1·92 |

**Dose- Expansion Module**

| Ogawa IgG (RAU) | | | | | | | | | |
|---|---|---|---|---|---|---|---|---|---|
| **Dose (CFU)** | **D1** | **D4** | **D7** | **D15** | **D29** | **D57** | **D85** | **D180** | **max** |
| 0 | 0·0127 | 0·0148 | 0·0192 | 0·0096 | 0·0178 | 0·0194 | 0·0099 | 0·0196 | 0·0196 |
| 0 | 0·0383 | 0·0465 | 0·0391 | 0·0286 | 0·0328 | 0·0313 | 0·0333 | 0·0306 | 0·0465 |
| 0 | 0·0283 | 0·0269 | 0·0244 | 0·0310 | 0·0342 | 0·0326 | 0·0364 | 0·0039 | 0·0364 |
| 0 | 0·0813 | 0·0877 | 0·0848 | 0·0743 | 0·0829 | 0·0175 | 0·0094 | 0·0080 | 0·0877 |
| 0 | 0·0495 | 0·0586 | 0·0528 | 0·0532 | 0·0471 | 0·0001 | 0·0023 | | 0·0586 |
| 0 | 0·0249 | 0·0205 | 0·0306 | 0·0015 | 0·0015 | 0·0032 | 0·0002 | | 0·0306 |
| 0 | 0·0018 | | | 0·0039 | | 0·0019 | | 0·0004 | 0·0039 |
| $2\times10^7$ | 0·0895 | 0·0598 | 0·0569 | 0·0592 | 0·0618 | 0·0629 | 0·0611 | 0·0833 | 0·0895 |
| $2\times10^7$ | 0·0403 | 0·0324 | 0·0726 | 1·8673 | 1·5346 | 1·0516 | 0·6851 | 0·1925 | 1·8673 |
| $2\times10^7$ | 0·0211 | 0·0172 | 0·0160 | 0·0246 | 0·0326 | 0·0207 | 0·0242 | 0·0152 | 0·0326 |
| $2\times10^7$ | 0·0253 | 0·0258 | 0·0255 | 0·0182 | 0·0262 | 0·0242 | 0·0176 | 0·0028 | 0·0262 |
| $2\times10^7$ | 0·0423 | 0·0439 | 0·0355 | 0·3180 | 0·1977 | 0·1358 | 0·1093 | 0·0389 | 0·3180 |
| $2\times10^7$ | 0·0260 | 0·0229 | 0·0228 | 0·0319 | 0·0352 | 0·0263 | 0·0296 | 0·0038 | 0·0352 |
| $2\times10^7$ | 0·0641 | | 0·0590 | 0·0837 | 0·0740 | 0·0740 | 0·0464 | | 0·0837 |
| $2\times10^7$ | 0·0170 | 0·0154 | 0·0144 | 0·0215 | 0·0244 | 0·0008 | | 0·0001 | 0·0244 |
| $2\times10^7$ | 0·0590 | 0·0600 | 0·0571 | 0·0542 | 0·0527 | 0·0101 | 0·0132 | 0·0322 | 0·0600 |
| $2\times10^7$ | 0·0405 | 0·0430 | 0·0395 | 0·1063 | 0·0354 | 0·0750 | 0·0952 | 0·3584 | 0·3584 |
| $2\times10^7$ | | | 0·0005 | | 0·0046 | 0·0047 | 0·0126 | 0·0256 | 0·0256 |
| $2\times10^7$ | 0·0064 | 0·0030 | 0·0036 | 0·0006 | 0·0089 | 0·0077 | 0·0104 | 0·0434 | 0·0434 |
| $2\times10^7$ | | 0·0004 | 0·0049 | 0·0056 | 0·0016 | 0·0015 | 0·0016 | 0·0016 | 0·0056 |
| $2\times10^7$ | 0·0001 | 0·0012 | 0·0001 | 0·0620 | 0·0617 | 0·0335 | | 0·0919 | 0·0919 |
| $2\times10^8$ | 0·0310 | 0·0349 | 0·0469 | 0·0688 | 0·0458 | 0·0647 | 0·1801 | 0·3126 | 0·3126 |
| $2\times10^8$ | 0·0040 | 0·0153 | 0·0078 | 0·0078 | 0·0076 | 0·0086 | 0·0106 | 0·0082 | 0·0153 |
| $2\times10^8$ | 0·0355 | 0·0318 | 0·0326 | 0·0321 | 0·0345 | 0·0701 | 0·2209 | 0·2314 | 0·2314 |
| $2\times10^8$ | 0·0452 | 0·0397 | 0·0403 | 0·0700 | 0·0710 | 0·0954 | 0·0721 | 0·1552 | 0·1552 |
| $2\times10^8$ | 0·0110 | 0·0180 | 0·0313 | 1·2342 | 0·9557 | 0·1816 | 0·0954 | 0·0180 | 1·2342 |
| $2\times10^8$ | 0·0164 | 0·0094 | 0·0042 | 0·0156 | | | | | 0·0164 |
| $2\times10^8$ | 0·0408 | 0·0468 | 0·0452 | 0·1236 | 0·0327 | 0·0625 | 0·0894 | 0·0484 | 0·1236 |
| $2\times10^8$ | 0·0127 | 0·0069 | 0·0062 | 0·0211 | 0·0029 | 0·0045 | | 0·0008 | 0·0211 |
| $2\times10^8$ | 0·0293 | 0·0244 | 0·0283 | 0·0301 | 0·0151 | 0·0097 | 0·0433 | 0·2118 | 0·2118 |
| $2\times10^8$ | 0·0085 | 0·0108 | 0·0097 | 0·1293 | 0·1229 | 0·0586 | 0·0311 | 0·0184 | 0·1293 |
| $2\times10^8$ | 0·0020 | | 0·0028 | 0·0276 | 0·0193 | 0·0709 | 0·2478 | 0·5072 | 0·5072 |
| $2\times10^8$ | 0·0038 | 0·0043 | 0·0008 | 0·0050 | 0·0007 | | 0·0107 | 0·0227 | 0·0227 |
| $2\times10^8$ | 0·0008 | 0·0007 | 0·0014 | 0·0060 | 0·0026 | 0·0669 | 0·0638 | 0·0367 | 0·0669 |

| Fold change Ogawa IgG | | | | | | | | | |
|---|---|---|---|---|---|---|---|---|---|
| **Dose (CFU)** | **D1** | **D4** | **D7** | **D15** | **D29** | **D57** | **D85** | **D180** | **max FC** |
| 0 | 1·00 | 1·17 | 1·51 | 0·76 | 1·40 | 1·53 | 0·78 | 1·54 | 1·54 |
| 0 | 1·00 | 1·21 | 1·02 | 0·75 | 0·86 | 0·82 | 0·87 | 0·80 | 1·21 |
| 0 | 1·00 | 0·95 | 0·86 | 1·10 | 1·21 | 1·15 | 1·29 | 0·14 | 1·29 |
| 0 | 1·00 | 1·08 | 1·04 | 0·91 | 1·02 | 0·21 | 0·12 | 0·10 | 1·08 |
| 0 | 1·00 | 1·18 | 1·07 | 1·08 | 0·95 | 0·00 | 0·05 | | 1·18 |
| 0 | 1·00 | 0·82 | 1·23 | 0·06 | 0·06 | 0·13 | 0·01 | | 1·23 |
| 0 | 1·00 | | | 2·20 | | 1·08 | | 0·21 | 2·20 |
| $2\times10^7$ | 1·00 | 0·67 | 0·64 | 0·66 | 0·69 | 0·70 | 0·68 | 0·93 | 0·93 |
| $2\times10^7$ | 1·00 | 0·80 | 1·80 | 46·33 | 38·07 | 26·09 | 17·00 | 4·78 | 46·33 |
| $2\times10^7$ | 1·00 | 0·82 | 0·76 | 1·17 | 1·55 | 0·98 | 1·15 | 0·72 | 1·55 |
| $2\times10^7$ | 1·00 | 1·02 | 1·01 | 0·72 | 1·03 | 0·96 | 0·70 | 0·11 | 1·03 |
| $2\times10^7$ | 1·00 | 1·04 | 0·84 | 7·52 | 4·68 | 3·21 | 2·59 | 0·92 | 7·52 |
| $2\times10^7$ | 1·00 | 0·88 | 0·88 | 1·23 | 1·35 | 1·01 | 1·14 | 0·14 | 1·35 |
| $2\times10^7$ | 1·00 | | 0·92 | 1·31 | 1·15 | 1·15 | 0·72 | | 1·31 |
| $2\times10^7$ | 1·00 | 0·91 | 0·84 | 1·26 | 1·43 | 0·05 | | 0·00 | 1·43 |
| $2\times10^7$ | 1·00 | 1·02 | 0·97 | 0·92 | 0·89 | 0·17 | 0·22 | 0·55 | 1·02 |
| $2\times10^7$ | 1·00 | 1·06 | 0·98 | 2·63 | 0·87 | 1·85 | 2·35 | 8·86 | 8·86 |
| $2\times10^7$ | | | 1·00 | | 9·48 | 9·77 | 25·99 | 52·81 | 52·81 |
| $2\times10^7$ | 1·00 | 0·48 | 0·56 | 0·09 | 1·39 | 1·21 | 1·63 | 6·78 | 6·78 |
| $2\times10^7$ | | 1·00 | 12·89 | 14·72 | 4·31 | 3·95 | 4·31 | 4·31 | 14·72 |
| $2\times10^7$ | 1·00 | 23·53 | 1·00 | 1190·09 | 1184·97 | 643·75 | | 1762·98 | 1762·98 |
| $2\times10^8$ | 1·00 | 1·13 | 1·52 | 2·22 | 1·48 | 2·09 | 5·82 | 10·10 | 10·10 |
| $2\times10^8$ | 1·00 | 3·81 | 1·94 | 1·94 | 1·89 | 2·15 | 2·64 | 2·05 | 3·81 |
| $2\times10^8$ | 1·00 | 0·90 | 0·92 | 0·90 | 0·97 | 1·98 | 6·22 | 6·52 | 6·52 |
| $2\times10^8$ | 1·00 | 0·88 | 0·89 | 1·55 | 1·57 | 2·11 | 1·59 | 3·43 | 3·43 |
| $2\times10^8$ | 1·00 | 1·64 | 2·85 | 112·55 | 87·15 | 16·56 | 8·70 | 1·64 | 112·55 |
| $2\times10^8$ | 1·00 | 0·57 | 0·26 | 0·95 | | | | | 0·95 |
| $2\times10^8$ | 1·00 | 1·15 | 1·11 | 3·03 | 0·80 | 1·53 | 2·19 | 1·19 | 3·03 |
| $2\times10^8$ | 1·00 | 0·54 | 0·48 | 1·66 | 0·23 | 0·35 | | 0·07 | 1·66 |
| $2\times10^8$ | 1·00 | 0·83 | 0·97 | 1·03 | 0·51 | 0·33 | 1·48 | 7·24 | 7·24 |
| $2\times10^8$ | 1·00 | 1·27 | 1·14 | 15·16 | 14·41 | 6·88 | 3·65 | 2·16 | 15·16 |
| $2\times10^8$ | 1·00 | | 1·35 | 13·47 | 9·41 | 34·57 | 120·87 | 247·44 | 247·44 |
| $2\times10^8$ | 1·00 | 1·15 | 0·22 | 1·34 | 0·19 | | 2·84 | 6·06 | 6·06 |
| $2\times10^8$ | 1·00 | 0·85 | 1·62 | 7·14 | 3·12 | 79·75 | 76·02 | 43·77 | 79·75 |

| | RAU | | |
|---|---|---|---|
| | D1 | D39 | |
| *V. cholerae* | 0·0466 | 0·2631 | |
| | 0·0352 | 0·3114 | |
| | 0·0432 | 0·0614 | |
| | 0·0578 | 1·1366 | |
| | 0·0437 | 0·0710 | |
| | D1 | D11 | |
| Vaxchora D11 Challenge | 0·0922 | 0·1624 | |
| | 0·1894 | 0·1845 | |
| | 0·1549 | 0·1549 | |
| | 0·0935 | 0·0994 | |
| | 0·1142 | 0·1459 | |
| | 0·1397 | 0·1065 | |
| | D1 | D11 | D91 |
| Vaxchora D91 Challenge | 0·0398 | 0·0500 | 0·0476 |
| | 0·0105 | 0·0432 | 0·0393 |
| | 0·0627 | 0·1316 | 0·1042 |
| | 0·0978 | 0·1542 | 0·1010 |
| | 0·0256 | 0·0951 | 0·0990 |
| | 0·0300 | 0·0413 | 0·0383 |

| | RAU fold change | | | max FC |
|---|---|---|---|---|
| | D1 | D39 | | |
| *V. cholerae* | 1·00 | 5·64 | | 5·64 |
| | 1·00 | 8·84 | | 8·84 |
| | 1·00 | 1·42 | | 1·42 |
| | 1·00 | 19·66 | | 19·66 |
| | 1·00 | 1·62 | | 1·62 |
| | D1 | D11 | | |
| Vaxchora D11 Challenge | 1·00 | 1·76 | | 1·76 |
| | 1·00 | 0·97 | | 0·97 |
| | 1·00 | 1·00 | | 1·00 |
| | 1·00 | 1·06 | | 1·06 |
| | 1·00 | 1·28 | | 1·28 |
| | 1·00 | 0·76 | | 0·76 |
| | D1 | D11 | D91 | |
| Vaxchora D91 Challenge | 1·00 | 1·26 | 1·20 | 1·26 |
| | 1·00 | 4·11 | 3·73 | 4·11 |
| | 1·00 | 2·10 | 1·66 | 2·10 |
| | 1·00 | 1·58 | 1·03 | 1·58 |
| | 1·00 | 3·71 | 3·87 | 3·87 |
| | 1·00 | 1·38 | 1·28 | 1·38 |

**Dose-Escalation Module**

| Ogawa IgA (RAU) | | | | | | | | | |
|---|---|---|---|---|---|---|---|---|---|
| **Dose (CFU)** | **D1** | **D4** | **D7** | **D15** | **D29** | **D57** | **D85** | **D180** | **max** |
| $10^{5}$ | 0·0071 | 0·0130 | 0·0121 | 0·0604 | 0·0316 | 0·0213 | 0·0177 | 0·0174 | 0·0604 |
| $10^{5}$ | 0·0260 | 0·0242 | 0·0273 | 0·0231 | 0·0269 | 0·0233 | 0·0260 | | 0·0273 |
| $10^{5}$ | 0·0230 | 0·0220 | 0·0230 | 0·0450 | 0·0209 | 0·0195 | 0·0200 | | 0·0450 |
| $10^{6}$ | 0·0096 | 0·0092 | 0·0327 | 0·5537 | 0·0727 | 0·0362 | 0·0301 | | 0·5537 |
| $10^{6}$ | 0·0478 | 0·0285 | 0·0548 | 0·0754 | 0·0395 | 0·0305 | 0·0278 | 0·0233 | 0·0754 |
| $10^{6}$ | 0·0089 | 0·0134 | 0·0511 | 3·2380 | 0·4718 | 0·1249 | 0·0492 | 0·0281 | 3·2380 |
| $10^{7}$ | 0·0073 | 0·0103 | 0·0106 | 0·0428 | 0·0560 | 0·0516 | 0·0443 | 0·0315 | 0·0560 |
| $10^{7}$ | 0·0169 | 0·0165 | 0·1036 | 0·2986 | 0·0840 | 0·0241 | 0·0205 | 0·0050 | 0·2986 |
| $10^{7}$ | 0·0117 | 0·0101 | 0·0067 | 0·0231 | 0·0146 | 0·0088 | 0·0097 | 0·0002 | 0·0231 |
| $10^{8}$ | 0·0154 | 0·0175 | 0·0144 | 0·0722 | 0·0329 | 0·0252 | 0·0265 | 0·0174 | 0·0722 |
| $10^{8}$ | 0·0133 | 0·0118 | 0·0203 | 0·5481 | 0·0898 | 0·0212 | 0·0158 | 0·0133 | 0·5481 |
| $10^{8}$ | 0·0040 | 0·0023 | 0·0063 | 0·0389 | 0·0139 | 0·0116 | 0·0117 | 0·0156 | 0·0389 |
| $10^{9}$ | 0·0101 | 0·0111 | 0·0194 | 0·0394 | 0·0226 | 0·0164 | 0·0108 | 0·0094 | 0·0394 |
| $10^{9}$ | 0·0566 | 0·0457 | 0·0473 | 0·0187 | 0·1704 | 0·1148 | 0·1923 | | 0·1923 |
| $10^{9}$ | 0·0026 | 0·0033 | 0·0026 | 0·0091 | 0·0115 | 0·0066 | 0·0050 | 0·0100 | 0·0115 |
| $10^{10}$ | 0·0003 | 0·0010 | 0·0087 | 0·0070 | 0·0050 | | 0·0014 | 0·0014 | 0·0087 |
| $10^{10}$ | 0·0101 | 0·0108 | 0·0126 | 0·0417 | | | 0·0233 | 0·0136 | 0·0417 |
| $10^{10}$ | 0·0009 | 0·0016 | 0·0103 | 0·1074 | 0·0256 | 0·0117 | 0·0168 | 0·0307 | 0·1074 |

| Fold change Ogawa IgA | | | | | | | | | |
|---|---|---|---|---|---|---|---|---|---|
| **Dose (CFU)** | **D1** | **D4** | **D7** | **D15** | **D29** | **D57** | **D85** | **D180** | **max FC** |
| $10^{5}$ | 1·00 | 1·83 | 1·70 | 8·52 | 4·46 | 3·01 | 2·50 | 2·45 | 8·52 |
| $10^{5}$ | 1·00 | 0·93 | 1·05 | 0·89 | 1·03 | 0·90 | 1·00 | | 1·05 |
| $10^{5}$ | 1·00 | 0·96 | 1·00 | 1·95 | 0·91 | 0·85 | 0·87 | | 1·95 |
| $10^{6}$ | 1·00 | 0·95 | 3·40 | 57·67 | 7·57 | 3·77 | 3·14 | | 57·67 |
| $10^{6}$ | 1·00 | 0·60 | 1·15 | 1·58 | 0·83 | 0·64 | 0·58 | 0·49 | 1·58 |
| $10^{6}$ | 1·00 | 1·50 | 5·71 | 362·13 | 52·76 | 13·96 | 5·50 | 3·14 | 362·13 |
| $10^{7}$ | 1·00 | 1·40 | 1·45 | 5·84 | 7·64 | 7·04 | 6·05 | 4·30 | 7·64 |
| $10^{7}$ | 1·00 | 0·98 | 6·14 | 17·70 | 4·98 | 1·43 | 1·22 | 0·30 | 17·70 |
| $10^{7}$ | 1·00 | 0·86 | 0·58 | 1·97 | 1·24 | 0·75 | 0·83 | 0·02 | 1·97 |
| $10^{8}$ | 1·00 | 1·14 | 0·94 | 4·70 | 2·14 | 1·64 | 1·73 | 1·13 | 4·70 |
| $10^{8}$ | 1·00 | 0·89 | 1·53 | 41·36 | 6·78 | 1·60 | 1·19 | 1·00 | 41·36 |
| $10^{8}$ | 1·00 | 0·57 | 1·57 | 9·69 | 3·47 | 2·88 | 2·93 | 3·89 | 9·69 |
| $10^{9}$ | 1·00 | 1·10 | 1·92 | 3·89 | 2·24 | 1·62 | 1·06 | 0·92 | 3·89 |
| $10^{9}$ | 1·00 | 0·81 | 0·84 | 0·33 | 3·01 | 2·03 | 3·40 | | 3·40 |
| $10^{9}$ | 1·00 | 1·27 | 1·01 | 3·52 | 4·48 | 2·55 | 1·96 | 3·88 | 4·48 |
| $10^{10}$ | 1·00 | 3·13 | 26·62 | 21·55 | 15·48 | | 4·17 | 4·25 | 26·62 |
| $10^{10}$ | 1·00 | 1·07 | 1·26 | 4·15 | | | 2·32 | 1·36 | 4·15 |
| $10^{10}$ | 1·00 | 1·67 | 10·90 | 113·41 | 27·04 | 12·30 | 17·71 | 32·41 | 113·41 |

**Dose- Expansion Module**

| Ogawa IgA (RAU) | | | | | | | | | |
|---|---|---|---|---|---|---|---|---|---|
| **Dose (CFU)** | **D1** | **D4** | **D7** | **D15** | **D29** | **D57** | **D85** | **D180** | **max** |
| 0 | 0·0255 | 0·0266 | 0·0276 | 0·0316 | 0·0320 | 0·0304 | 0·0357 | 0·0260 | 0·0357 |
| 0 | 0·0234 | 0·0166 | 0·0200 | 0·0179 | 0·0168 | 0·0157 | 0·0166 | 0·0170 | 0·0234 |
| 0 | 0·0242 | 0·0270 | 0·0207 | 0·0174 | 0·0217 | 0·0247 | 0·0148 | 0·0146 | 0·0270 |
| 0 | 0·0370 | 0·0455 | 0·0479 | 0·0312 | 0·0281 | 0·0173 | 0·0141 | 0·0111 | 0·0479 |
| 0 | 0·0118 | 0·0086 | 0·0155 | 0·0284 | 0·0104 | 0·0126 | 0·0107 | 0·0064 | 0·0284 |
| 0 | 0·0185 | 0·0191 | 0·0179 | 0·0081 | 0·0068 | 0·0079 | 0·0086 | 0·0061 | 0·0191 |
| 0 | 0·0133 | 0·0110 | 0·0148 | 0·0141 | 0·0179 | 0·0115 | 0·0072 | 0·0092 | 0·0179 |
| $2\times10^{7}$ | 0·0102 | 0·0179 | 0·0125 | 0·0567 | 0·0201 | 0·0149 | 0·0240 | 0·0176 | 0·0567 |
| $2\times10^{7}$ | 0·0195 | 0·0251 | 0·0330 | 2·4397 | 0·6934 | 0·2000 | 0·1342 | 0·0554 | 2·4397 |
| $2\times10^{7}$ | 0·0220 | 0·0257 | 0·0363 | 0·1226 | 0·0756 | 0·0379 | 0·0265 | 0·0135 | 0·1226 |
| $2\times10^{7}$ | 0·0373 | 0·0273 | 0·0195 | 0·0247 | 0·0296 | 0·0354 | 0·0283 | 0·0124 | 0·0373 |
| $2\times10^{7}$ | 0·0201 | 0·0188 | 0·0695 | 1·6153 | 0·2724 | 0·0582 | 0·0526 | 0·0365 | 1·6153 |
| $2\times10^{7}$ | 0·0257 | 0·0304 | 0·3470 | 2·3296 | 1·0663 | 0·2061 | 0·1426 | 0·0345 | 2·3296 |
| $2\times10^{7}$ | 0·0367 | | 0·0291 | 0·2740 | 0·0698 | 0·0341 | 0·0220 | | 0·2740 |
| $2\times10^{7}$ | 0·0180 | 0·0305 | 0·1172 | 0·3785 | 0·1655 | 0·0450 | 0·0327 | 0·0188 | 0·3785 |
| $2\times10^{7}$ | 0·0180 | 0·0201 | 0·0210 | 0·0217 | 0·0233 | 0·0220 | 0·0231 | 0·0255 | 0·0255 |
| $2\times10^{7}$ | 0·0182 | 0·0186 | 0·0480 | 4·3940 | 0·2034 | 0·0976 | 0·0708 | 0·0499 | 4·3940 |
| $2\times10^{7}$ | 0·0084 | 0·0059 | 0·0339 | 0·1923 | 0·1320 | 0·0507 | 0·0439 | 0·0443 | 0·1923 |
| $2\times10^{7}$ | 0·0136 | 0·0152 | 0·0268 | 0·0120 | 0·0196 | 0·0278 | 0·0388 | 0·0360 | 0·0388 |
| $2\times10^{7}$ | 0·0023 | 0·0012 | 0·0051 | 0·0130 | 0·0104 | 0·0052 | 0·0062 | 0·0008 | 0·0130 |
| $2\times10^{7}$ | 0·0105 | 0·0088 | 0·0446 | 0·9925 | 0·3131 | 0·1013 | | 0·0494 | 0·9925 |
| $2\times10^{8}$ | 0·0381 | 0·0526 | 0·0344 | 0·0664 | 0·0580 | 0·0419 | 0·0364 | 0·0401 | 0·0664 |
| $2\times10^{8}$ | 0·0411 | 0·0520 | 0·0488 | 0·0508 | 0·0596 | 0·0554 | 0·0457 | 0·0207 | 0·0596 |
| $2\times10^{8}$ | 0·0079 | 0·0118 | 0·0108 | 0·0266 | 0·0192 | 0·0237 | 0·0200 | 0·0179 | 0·0266 |
| $2\times10^{8}$ | 0·0309 | 0·0257 | 0·3772 | 0·5365 | 0·2586 | 0·0842 | 0·0426 | 0·0286 | 0·5365 |
| $2\times10^{8}$ | 0·0097 | 0·0123 | 0·0450 | 0·1886 | 0·0898 | 0·0093 | 0·0042 | 0·0002 | 0·1886 |
| $2\times10^{8}$ | 0·0192 | 0·0099 | 0·0092 | 0·0321 | | | | | 0·0321 |
| $2\times10^{8}$ | 0·0149 | 0·0135 | 0·0160 | 0·1561 | 0·0239 | 0·0198 | 0·0201 | 0·0155 | 0·1561 |
| $2\times10^{8}$ | 0·0170 | 0·0126 | 0·0141 | 0·0272 | 0·0081 | 0·0066 | 0·0103 | 0·0107 | 0·0272 |
| $2\times10^{8}$ | 0·0388 | 0·0413 | 0·1759 | 0·7248 | 0·2783 | 0·1112 | 0·0821 | 0·0548 | 0·7248 |
| $2\times10^{8}$ | 0·0212 | 0·0232 | 1·3754 | 22·6371 | 7·5668 | 0·4771 | 0·2122 | 0·1139 | 22·6371 |
| $2\times10^{8}$ | 0·0086 | 0·0082 | 0·0170 | 0·2312 | 0·1066 | 0·0606 | 0·0284 | 0·0115 | 0·2312 |
| $2\times10^{8}$ | 0·0224 | 0·0393 | 0·0316 | 0·2160 | 0·0495 | | 0·0342 | 0·0294 | 0·2160 |
| $2\times10^{8}$ | 0·0056 | 0·0052 | 0·0088 | 0·0061 | 0·0100 | 0·0078 | 0·0133 | 0·0157 | 0·0157 |

| Fold change Ogawa IgA | | | | | | | | | |
|---|---|---|---|---|---|---|---|---|---|
| **Dose (CFU)** | **D1** | **D4** | **D7** | **D15** | **D29** | **D57** | **D85** | **D180** | **max FC** |
| 0 | 1·00 | 1·04 | 1·08 | 1·24 | 1·25 | 1·19 | 1·40 | 1·02 | 1·40 |
| 0 | 1·00 | 0·71 | 0·85 | 0·76 | 0·72 | 0·67 | 0·71 | 0·72 | 0·85 |
| 0 | 1·00 | 1·12 | 0·86 | 0·72 | 0·90 | 1·02 | 0·61 | 0·60 | 1·12 |
| 0 | 1·00 | 1·23 | 1·29 | 0·84 | 0·76 | 0·47 | 0·38 | 0·30 | 1·29 |
| 0 | 1·00 | 0·73 | 1·32 | 2·41 | 0·88 | 1·07 | 0·91 | 0·55 | 2·41 |
| 0 | 1·00 | 1·03 | 0·97 | 0·44 | 0·37 | 0·43 | 0·47 | 0·33 | 1·03 |
| 0 | 1·00 | 0·83 | 1·11 | 1·06 | 1·34 | 0·86 | 0·54 | 0·69 | 1·34 |
| $2\times10^{7}$ | 1·00 | 1·75 | 1·22 | 5·55 | 1·97 | 1·46 | 2·35 | 1·72 | 5·55 |
| $2\times10^{7}$ | 1·00 | 1·29 | 1·69 | 124·90 | 35·50 | 10·24 | 6·87 | 2·84 | 124·90 |
| $2\times10^{7}$ | 1·00 | 1·17 | 1·65 | 5·57 | 3·43 | 1·72 | 1·20 | 0·61 | 5·57 |
| $2\times10^{7}$ | 1·00 | 0·73 | 0·52 | 0·66 | 0·79 | 0·95 | 0·76 | 0·33 | 0·95 |
| $2\times10^{7}$ | 1·00 | 0·93 | 3·46 | 80·26 | 13·54 | 2·89 | 2·61 | 1·81 | 80·26 |
| $2\times10^{7}$ | 1·00 | 1·18 | 13·51 | 90·72 | 41·52 | 8·02 | 5·55 | 1·34 | 90·72 |
| $2\times10^{7}$ | 1·00 | | 0·79 | 7·48 | 1·90 | 0·93 | 0·60 | | 7·48 |
| $2\times10^{7}$ | 1·00 | 1·69 | 6·50 | 20·99 | 9·18 | 2·50 | 1·81 | 1·04 | 20·99 |
| $2\times10^{7}$ | 1·00 | 1·12 | 1·17 | 1·21 | 1·29 | 1·22 | 1·28 | 1·41 | 1·41 |
| $2\times10^{7}$ | 1·00 | 1·02 | 2·64 | 241·67 | 11·19 | 5·37 | 3·89 | 2·75 | 241·67 |
| $2\times10^{7}$ | 1·00 | 0·70 | 4·03 | 22·84 | 15·68 | 6·02 | 5·21 | 5·26 | 22·84 |
| $2\times10^{7}$ | 1·00 | 1·12 | 1·97 | 0·88 | 1·44 | 2·05 | 2·85 | 2·65 | 2·85 |
| $2\times10^{7}$ | 1·00 | 0·54 | 2·24 | 5·71 | 4·57 | 2·31 | 2·74 | 0·35 | 5·71 |
| $2\times10^{7}$ | 1·00 | 0·83 | 4·25 | 94·39 | 29·78 | 9·63 | | 4·70 | 94·39 |
| $2\times10^{8}$ | 1·00 | 1·38 | 0·90 | 1·74 | 1·52 | 1·10 | 0·95 | 1·05 | 1·74 |
| $2\times10^{8}$ | 1·00 | 1·27 | 1·19 | 1·24 | 1·45 | 1·35 | 1·11 | 0·50 | 1·45 |
| $2\times10^{8}$ | 1·00 | 1·50 | 1·37 | 3·39 | 2·45 | 3·02 | 2·54 | 2·28 | 3·39 |
| $2\times10^{8}$ | 1·00 | 0·83 | 12·20 | 17·36 | 8·37 | 2·72 | 1·38 | 0·93 | 17·36 |
| $2\times10^{8}$ | 1·00 | 1·27 | 4·64 | 19·45 | 9·27 | 0·96 | 0·43 | 0·02 | 19·45 |
| $2\times10^{8}$ | 1·00 | 0·51 | 0·48 | 1·67 | | | | | 1·67 |
| $2\times10^{8}$ | 1·00 | 0·90 | 1·07 | 10·46 | 1·60 | 1·32 | 1·35 | 1·04 | 10·46 |
| $2\times10^{8}$ | 1·00 | 0·74 | 0·83 | 1·60 | 0·48 | 0·39 | 0·61 | 0·63 | 1·60 |
| $2\times10^{8}$ | 1·00 | 1·07 | 4·54 | 18·70 | 7·18 | 2·87 | 2·12 | 1·41 | 18·70 |
| $2\times10^{8}$ | 1·00 | 1·10 | 65·00 | 1069·80 | 357·60 | 22·55 | 10·03 | 5·38 | 1069·80 |
| $2\times10^{8}$ | 1·00 | 0·95 | 1·97 | 26·80 | 12·35 | 7·02 | 3·29 | 1·33 | 26·80 |
| $2\times10^{8}$ | 1·00 | 1·76 | 1·41 | 9·65 | 2·21 | | 1·53 | 1·31 | 9·65 |
| $2\times10^{8}$ | 1·00 | 0·92 | 1·57 | 1·08 | 1·78 | 1·39 | 2·38 | 2·80 | 2·80 |

| | RAU | | |
|---|---|---|---|
| | D1 | D39 | |
| *V. cholerae* | 0·0502 | 31·3893 | |
| | 0·0360 | 0·4287 | |
| | 0·0368 | 0·0345 | |
| | 0·0243 | 8·6786 | |
| | 0·0516 | 0·3164 | |
| | D1 | D11 | |
| Vaxchora D11 Challenge | 0·0669 | 0·0869 | |
| | 0·1473 | 0·2058 | |
| | 0·1351 | 0·1881 | |
| | 0·0433 | 0·0399 | |
| | 0·0608 | 0·3761 | |
| | 0·1111 | 0·0684 | |
| | D1 | D11 | D91 |
| Vaxchora D91 Challenge | 0·0087 | 0·0118 | 0·0136 |
| | 0·0100 | 0·0434 | 0·0169 |
| | 0·0310 | 0·2712 | 0·0328 |
| | 0·0211 | 0·0541 | 0·0252 |
| | 0·0167 | 0·0550 | 0·0704 |
| | 0·0249 | 0·0324 | 0·0271 |

| | RAU fold change | | | max FC |
|---|---|---|---|---|
| | D1 | D39 | | |
| *V. cholerae* | 1·00 | 625·15 | | 625·15 |
| | 1·00 | 11·92 | | 11·92 |
| | 1·00 | 0·94 | | 0·94 |
| | 1·00 | 356·90 | | 356·90 |
| | 1·00 | 6·14 | | 6·14 |
| | D1 | D11 | | |
| Vaxchora D11 Challenge | 1·00 | 1·30 | | 1·30 |
| | 1·00 | 1·40 | | 1·40 |
| | 1·00 | 1·39 | | 1·39 |
| | 1·00 | 0·92 | | 0·92 |
| | 1·00 | 6·19 | | 6·19 |
| | 1·00 | 0·62 | | 0·62 |
| | D1 | D11 | D91 | |
| Vaxchora D91 Challenge | 1·00 | 1·35 | 1·56 | 1·56 |
| | 1·00 | 4·36 | 1·69 | 4·36 |
| | 1·00 | 8·76 | 1·06 | 8·76 |
| | 1·00 | 2·57 | 1·19 | 2·57 |
| | 1·00 | 3·30 | 4·23 | 4·23 |
| | 1·00 | 1·30 | 1·09 | 1·30 |

**Dose-Escalation Module**

| CT-B IgM (RAU) | | | | | | | | | |
|---|---|---|---|---|---|---|---|---|---|
| **Dose (CFU)** | **D1** | **D4** | **D7** | **D15** | **D29** | **D57** | **D85** | **D180** | **max** |
| $10^5$ | 48·7681 | 39·8867 | 49·3276 | 35·5923 | 33·7757 | 31·5007 | 27·3105 | 31·9335 | 49·3276 |
| $10^5$ | 4·3177 | 4·2725 | 4·8235 | 4·8335 | 8·8920 | 6·5285 | 8·5346 | | 8·8920 |
| $10^5$ | 9·5358 | 8·8401 | 10·8047 | 9·4074 | 9·4989 | 8·8229 | 9·5727 | | 10·8047 |
| $10^6$ | 5·5944 | 5·1347 | 5·2430 | 4·7094 | 5·0782 | 6·1324 | 5·4690 | | 6·1324 |
| $10^6$ | 41·6655 | 56·4419 | 62·8863 | 41·8381 | 29·2414 | 41·7635 | 36·6352 | 38·4355 | 62·8863 |
| $10^6$ | 16·0650 | 13·7872 | 18·7657 | 15·3953 | 15·8055 | 14·7220 | 18·9365 | 8·8397 | 18·9365 |
| $10^7$ | 39·1511 | 35·5127 | 45·5692 | 48·1872 | 40·8617 | 49·7729 | 63·0023 | 25·0933 | 63·0023 |
| $10^7$ | 5·3346 | 5·6100 | 4·8418 | 5·1347 | 6·4255 | 5·6917 | 5·3980 | 4·6634 | 6·4255 |
| $10^7$ | 8·2430 | 8·8641 | 6·6112 | 8·0020 | 7·3857 | 7·6316 | 8·1349 | 4·3412 | 8·8641 |
| $10^8$ | 6·7071 | 5·9575 | 4·7675 | 4·0417 | 4·3163 | 4·8582 | 5·0863 | 4·1777 | 6·7071 |
| $10^8$ | 9·0076 | 8·2534 | 8·4373 | 9·3846 | 8·2047 | 9·2457 | 3·5076 | 3·7795 | 9·3846 |
| $10^8$ | 9·6409 | 9·7854 | 11·8589 | 10·1366 | 9·8659 | 9·8469 | 9·6533 | 16·0359 | 16·0359 |
| $10^9$ | 6·3882 | 5·9345 | 6·7990 | 5·4847 | 5·4296 | 3·1768 | 3·2137 | 4·0957 | 6·7990 |
| $10^9$ | 6·5520 | 6·0337 | 4·9401 | 4·3706 | 6·3251 | 4·5692 | 9·7566 | | 9·7566 |
| $10^9$ | 5·7821 | 6·3789 | 7·0435 | 6·9148 | 6·3504 | 6·7480 | 7·6381 | 6·8802 | 7·6381 |
| $10^{10}$ | 3·9379 | 4·5920 | 5·6449 | 7·2536 | 6·5116 | | 4·8836 | 5·6129 | 7·2536 |
| $10^{10}$ | 8·7717 | 10·6467 | 7·0027 | 7·2599 | | | 6·7480 | 7·3569 | 10·6467 |
| $10^{10}$ | 10·7689 | 10·1323 | 13·0508 | 24·3296 | 11·4982 | 8·5427 | 9·4011 | 8·0897 | 24·3296 |

| Fold change CT-B IgM | | | | | | | | | |
|---|---|---|---|---|---|---|---|---|---|
| **Dose (CFU)** | **D1** | **D4** | **D7** | **D15** | **D29** | **D57** | **D85** | **D180** | **max FC** |
| $10^5$ | 1·00 | 0·82 | 1·01 | 0·73 | 0·69 | 0·65 | 0·56 | 0·65 | 1·01 |
| $10^5$ | 1·00 | 0·99 | 1·12 | 1·12 | 2·06 | 1·51 | 1·98 | | 2·06 |
| $10^5$ | 1·00 | 0·93 | 1·13 | 0·99 | 1·00 | 0·93 | 1·00 | | 1·13 |
| $10^6$ | 1·00 | 0·92 | 0·94 | 0·84 | 0·91 | 1·10 | 0·98 | | 1·10 |
| $10^6$ | 1·00 | 1·35 | 1·51 | 1·00 | 0·70 | 1·00 | 0·88 | 0·92 | 1·51 |
| $10^6$ | 1·00 | 0·86 | 1·17 | 0·96 | 0·98 | 0·92 | 1·18 | 0·55 | 1·18 |
| $10^7$ | 1·00 | 0·91 | 1·16 | 1·23 | 1·04 | 1·27 | 1·61 | 0·64 | 1·61 |
| $10^7$ | 1·00 | 1·05 | 0·91 | 0·96 | 1·20 | 1·07 | 1·01 | 0·87 | 1·20 |
| $10^7$ | 1·00 | 1·08 | 0·80 | 0·97 | 0·90 | 0·93 | 0·99 | 0·53 | 1·08 |
| $10^8$ | 1·00 | 0·89 | 0·71 | 0·60 | 0·64 | 0·72 | 0·76 | 0·62 | 0·89 |
| $10^8$ | 1·00 | 0·92 | 0·94 | 1·04 | 0·91 | 1·03 | 0·39 | 0·42 | 1·04 |
| $10^8$ | 1·00 | 1·01 | 1·23 | 1·05 | 1·02 | 1·02 | 1·00 | 1·66 | 1·66 |
| $10^9$ | 1·00 | 0·93 | 1·06 | 0·86 | 0·85 | 0·50 | 0·50 | 0·64 | 1·06 |
| $10^9$ | 1·00 | 0·92 | 0·75 | 0·67 | 0·97 | 0·70 | 1·49 | | 1·49 |
| $10^9$ | 1·00 | 1·10 | 1·22 | 1·20 | 1·10 | 1·17 | 1·32 | 1·19 | 1·32 |
| $10^{10}$ | 1·00 | 1·17 | 1·43 | 1·84 | 1·65 | | 1·24 | 1·43 | 1·84 |
| $10^{10}$ | 1·00 | 1·21 | 0·80 | 0·83 | | | 0·77 | 0·84 | 1·21 |
| $10^{10}$ | 1·00 | 0·94 | 1·21 | 2·26 | 1·07 | 0·79 | 0·87 | 0·75 | 2·26 |

**Dose- Expansion Module**

| CT-B IgM (RAU) | | | | | | | | | |
|---|---|---|---|---|---|---|---|---|---|
| **Dose (CFU)** | **D1** | **D4** | **D7** | **D15** | **D29** | **D57** | **D85** | **D180** | **max** |
| 0 | 1·9038 | 2·0065 | 2·2448 | 1·9413 | 2·2285 | 2·3885 | 21·1801 | 4·1024 | 21·1801 |
| 0 | 1·6485 | 1·8299 | 1·8140 | 1·9127 | 2·6778 | 1·9566 | 1·4712 | 1·4018 | 2·6778 |
| 0 | 4·3432 | 4·6302 | 4·9423 | 4·4139 | 4·3243 | 3·4401 | 2·7802 | 7·1160 | 7·1160 |
| 0 | 37·2078 | 41·6211 | 44·4041 | 37·9207 | 36·9638 | 83·2086 | 69·7915 | 68·7031 | 83·2086 |
| 0 | 3·2106 | 2·8629 | 3·6935 | 3·2758 | 3·4934 | 6·3596 | 6·0827 | 6·9474 | 6·9474 |
| 0 | 5·5765 | 5·6905 | 5·8598 | 7·9676 | 4·6655 | 7·9362 | 7·4938 | 8·4349 | 8·4349 |
| 0 | 14·3808 | 12·9317 | 16·0618 | 11·9490 | 17·4674 | 14·1165 | 11·5488 | 16·8740 | 17·4674 |
| $2\times10^7$ | 24·0857 | 24·1834 | 21·4040 | 23·0546 | 22·5363 | 18·7744 | 19·5633 | 24·7112 | 24·7112 |
| $2\times10^7$ | 5·6271 | 5·5908 | 5·6401 | 4·3040 | 4·1883 | 4·4226 | 5·8814 | 4·3649 | 5·8814 |
| $2\times10^7$ | 6·1090 | 6·6454 | 5·6103 | 5·8151 | 5·4235 | 5·0607 | 5·2953 | 4·9259 | 6·6454 |
| $2\times10^7$ | 12·5464 | 19·5569 | 16·0379 | 14·8629 | 12·9106 | 15·0548 | 12·0859 | 13·2007 | 19·5569 |
| $2\times10^7$ | 9·8241 | 9·3126 | 11·3472 | 12·2269 | 10·4226 | 8·4987 | 9·2273 | 8·1421 | 12·2269 |
| $2\times10^7$ | 3·6131 | 3·4433 | 3·3486 | 3·1904 | 3·3057 | 3·0901 | 3·3617 | 9·5358 | 9·5358 |
| $2\times10^7$ | 9·0962 | | 8·4625 | 8·1234 | 8·1771 | 9·3977 | 18·0794 | | 18·0794 |
| $2\times10^7$ | 3·2106 | 3·5431 | 3·5207 | 10·9079 | 7·9098 | 11·3101 | 7·7351 | 41·5786 | 41·5786 |
| $2\times10^7$ | 3·2641 | 3·7977 | 3·5367 | 3·3007 | 3·3519 | 6·0213 | 4·6752 | 4·8941 | 6·0213 |
| $2\times10^7$ | 13·6033 | 15·2247 | 17·7578 | 13·9511 | 21·6282 | 19·7118 | 20·6870 | 26·1957 | 26·1957 |
| $2\times10^7$ | 11·9490 | 17·8840 | 18·8798 | 12·9802 | 14·8129 | 13·6263 | 13·4999 | 12·1080 | 18·8798 |
| $2\times10^7$ | 39·4900 | 40·1532 | 55·3359 | 38·9665 | 40·4886 | 43·5495 | 47·5105 | 46·2821 | 55·3359 |
| $2\times10^7$ | 64·4998 | 70·6733 | 76·7717 | 70·3415 | 70·4519 | 71·1180 | 69·6820 | 71·7896 | 76·7717 |
| $2\times10^7$ | 6·8508 | 5·3157 | 7·5534 | 6·5812 | 7·6136 | 6·5680 | | 5·3596 | 7·6136 |
| $2\times10^8$ | 4·7533 | 4·7087 | 4·8929 | 6·8490 | 4·5766 | 4·2280 | 4·7129 | 4·4082 | 6·8490 |
| $2\times10^8$ | 82·5645 | 84·1662 | 93·3131 | 95·9979 | 83·2462 | 88·2217 | 83·0182 | 60·5049 | 95·9979 |
| $2\times10^8$ | 7·1517 | 7·1885 | 7·2182 | 7·5208 | 6·5330 | 6·1216 | 7·9833 | 7·5534 | 7·9833 |
| $2\times10^8$ | 11·7598 | 12·0657 | 10·7723 | 6·4975 | 7·5173 | 9·0996 | 8·8401 | 6·5548 | 12·0657 |
| $2\times10^8$ | 18·3957 | 20·2094 | 20·4380 | 13·0529 | 14·9143 | 20·7239 | 24·5568 | 25·9235 | 25·9235 |
| $2\times10^8$ | 7·1030 | 6·8610 | 6·9043 | 6·5893 | | | | | 7·1030 |
| $2\times10^8$ | 9·6334 | 11·8608 | 10·6117 | 23·7578 | 27·7874 | 15·2005 | 16·7208 | 11·8590 | 27·7874 |
| $2\times10^8$ | 2·7511 | 2·4392 | 2·5143 | 2·2992 | 3·0127 | 3·3581 | 4·6460 | 3·8846 | 4·6460 |
| $2\times10^8$ | 5·1993 | 4·6006 | 4·8530 | 3·8354 | 4·9144 | 4·2367 | 4·1658 | 4·5310 | 5·1993 |
| $2\times10^8$ | 3·2580 | 3·8272 | 3·1469 | 2·7476 | 2·2739 | 2·4538 | 2·6581 | 2·4868 | 3·8272 |
| $2\times10^8$ | 12·5256 | 15·6546 | 16·2095 | 17·0904 | 15·8279 | 15·7410 | 16·3285 | 16·9356 | 17·0904 |
| $2\times10^8$ | 3·9849 | 3·5973 | 2·7174 | 2·6817 | 2·7476 | | 2·6172 | 3·9680 | 3·9849 |
| $2\times10^8$ | 2·6876 | 2·4429 | 3·4909 | 3·3292 | 3·9596 | 3·4984 | 3·2934 | 3·9012 | 3·9596 |

| Fold change CT-B IgM | | | | | | | | | |
|---|---|---|---|---|---|---|---|---|---|
| **Dose (CFU)** | **D1** | **D4** | **D7** | **D15** | **D29** | **D57** | **D85** | **D180** | **max FC** |
| 0 | 1·00 | 1·05 | 1·18 | 1·02 | 1·17 | 1·25 | 11·13 | 2·15 | 11·13 |
| 0 | 1·00 | 1·11 | 1·10 | 1·16 | 1·62 | 1·19 | 0·89 | 0·85 | 1·62 |
| 0 | 1·00 | 1·07 | 1·14 | 1·02 | 1·00 | 0·79 | 0·64 | 1·64 | 1·64 |
| 0 | 1·00 | 1·12 | 1·19 | 1·02 | 0·99 | 2·24 | 1·88 | 1·85 | 2·24 |
| 0 | 1·00 | 0·89 | 1·15 | 1·02 | 1·09 | 1·98 | 1·89 | 2·16 | 2·16 |
| 0 | 1·00 | 1·02 | 1·05 | 1·43 | 0·84 | 1·42 | 1·34 | 1·51 | 1·51 |
| 0 | 1·00 | 0·90 | 1·12 | 0·83 | 1·21 | 0·98 | 0·80 | 1·17 | 1·21 |
| $2\times10^7$ | 1·00 | 1·00 | 0·89 | 0·96 | 0·94 | 0·78 | 0·81 | 1·03 | 1·03 |
| $2\times10^7$ | 1·00 | 0·99 | 1·00 | 0·76 | 0·74 | 0·79 | 1·05 | 0·78 | 1·05 |
| $2\times10^7$ | 1·00 | 1·09 | 0·92 | 0·95 | 0·89 | 0·83 | 0·87 | 0·81 | 1·09 |
| $2\times10^7$ | 1·00 | 1·56 | 1·28 | 1·18 | 1·03 | 1·20 | 0·96 | 1·05 | 1·56 |
| $2\times10^7$ | 1·00 | 0·95 | 1·16 | 1·24 | 1·06 | 0·87 | 0·94 | 0·83 | 1·24 |
| $2\times10^7$ | 1·00 | 0·95 | 0·93 | 0·88 | 0·91 | 0·86 | 0·93 | 2·64 | 2·64 |
| $2\times10^7$ | 1·00 | | 0·93 | 0·89 | 0·90 | 1·03 | 1·99 | | 1·99 |
| $2\times10^7$ | 1·00 | 1·10 | 1·10 | 3·40 | 2·46 | 3·52 | 2·41 | 12·95 | 12·95 |
| $2\times10^7$ | 1·00 | 1·16 | 1·08 | 1·01 | 1·03 | 1·84 | 1·43 | 1·50 | 1·84 |
| $2\times10^7$ | 1·00 | 1·12 | 1·31 | 1·03 | 1·59 | 1·45 | 1·52 | 1·93 | 1·93 |
| $2\times10^7$ | 1·00 | 1·50 | 1·58 | 1·09 | 1·24 | 1·14 | 1·13 | 1·01 | 1·58 |
| $2\times10^7$ | 1·00 | 1·02 | 1·40 | 0·99 | 1·03 | 1·10 | 1·20 | 1·17 | 1·40 |
| $2\times10^7$ | 1·00 | 1·10 | 1·19 | 1·09 | 1·09 | 1·10 | 1·08 | 1·11 | 1·19 |
| $2\times10^7$ | 1·00 | 0·78 | 1·10 | 0·96 | 1·11 | 0·96 | | 0·78 | 1·11 |
| $2\times10^8$ | 1·00 | 0·99 | 1·03 | 1·44 | 0·96 | 0·89 | 0·99 | 0·93 | 1·44 |
| $2\times10^8$ | 1·00 | 1·02 | 1·13 | 1·16 | 1·01 | 1·07 | 1·01 | 0·73 | 1·16 |
| $2\times10^8$ | 1·00 | 1·01 | 1·01 | 1·05 | 0·91 | 0·86 | 1·12 | 1·06 | 1·12 |
| $2\times10^8$ | 1·00 | 1·03 | 0·92 | 0·55 | 0·64 | 0·77 | 0·75 | 0·56 | 1·03 |
| $2\times10^8$ | 1·00 | 1·10 | 1·11 | 0·71 | 0·81 | 1·13 | 1·33 | 1·41 | 1·41 |
| $2\times10^8$ | 1·00 | 0·97 | 0·97 | 0·93 | | | | | 0·97 |
| $2\times10^8$ | 1·00 | 1·23 | 1·10 | 2·47 | 2·88 | 1·58 | 1·74 | 1·23 | 2·88 |
| $2\times10^8$ | 1·00 | 0·89 | 0·91 | 0·84 | 1·10 | 1·22 | 1·69 | 1·41 | 1·69 |
| $2\times10^8$ | 1·00 | 0·88 | 0·93 | 0·74 | 0·95 | 0·81 | 0·80 | 0·87 | 0·95 |
| $2\times10^8$ | 1·00 | 1·17 | 0·97 | 0·84 | 0·70 | 0·75 | 0·82 | 0·76 | 1·17 |
| $2\times10^8$ | 1·00 | 1·25 | 1·29 | 1·36 | 1·26 | 1·26 | 1·30 | 1·35 | 1·36 |
| $2\times10^8$ | 1·00 | 0·90 | 0·68 | 0·67 | 0·69 | | 0·66 | 1·00 | 1·00 |
| $2\times10^8$ | 1·00 | 0·91 | 1·30 | 1·24 | 1·47 | 1·30 | 1·23 | 1·45 | 1·47 |

| | RAU | | |
|---|---|---|---|
| | D1 | D39 | |
| *V. cholerae* | 9·4893 | 128·5977 | |
| | 16·1163 | 283·2030 | |
| | 5·5553 | 24·5622 | |
| | 6·9451 | 6·4255 | |
| | 4·6343 | 62·9635 | |
| | D1 | D11 | |
| Vaxchora D11 Challenge | 3·0816 | 3·8228 | |
| | 26·6614 | 28·1467 | |
| | 4·2775 | 44·4870 | |
| | 5·8424 | 5·3346 | |
| | 6·0566 | 6·3096 | |
| | 7·1375 | 5·4690 | |
| | D1 | D11 | D91 |
| Vaxchora D91 Challenge | 4·4531 | 3·9351 | 3·9217 |
| | 55·3714 | 25·4225 | 14·9817 |
| | 1·8829 | 1·9988 | 1·9747 |
| | 29·8887 | 21·0560 | 47·4493 |
| | 5·8809 | 12·4505 | 5·9575 |
| | 5·8732 | 5·8885 | 5·1267 |

| | RAU fold change | | | max FC |
|---|---|---|---|---|
| | D1 | D39 | | |
| *V. cholerae* | 1·00 | 13·55 | | 13·55 |
| | 1·00 | 17·57 | | 17·57 |
| | 1·00 | 4·42 | | 4·42 |
| | 1·00 | 0·93 | | 0·93 |
| | 1·00 | 13·59 | | 13·59 |
| | D1 | D11 | | |
| Vaxchora D11 Challenge | 1·00 | 1·24 | | 1·24 |
| | 1·00 | 1·06 | | 1·06 |
| | 1·00 | 10·40 | | 10·40 |
| | 1·00 | 0·91 | | 0·91 |
| | 1·00 | 1·04 | | 1·04 |
| | 1·00 | 0·77 | | 0·77 |
| | D1 | D11 | D91 | |
| Vaxchora D91 Challenge | 1·00 | 0·88 | 0·88 | 0·88 |
| | 1·00 | 0·46 | 0·27 | 0·46 |
| | 1·00 | 1·06 | 1·05 | 1·06 |
| | 1·00 | 0·70 | 1·59 | 1·59 |
| | 1·00 | 2·12 | 1·01 | 2·12 |
| | 1·00 | 1·00 | 0·87 | 1·00 |

**Dose-Escalation Module**

| CT-B IgG (RAU) | | | | | | | | | |
|---|---|---|---|---|---|---|---|---|---|
| Dose (CFU) | D1 | D4 | D7 | D15 | D29 | D57 | D85 | D180 | max |
| $10^5$ | 0·1851 | 0·1728 | 0·1877 | 0·7514 | 0·7877 | 0·5807 | 0·5334 | 0·3417 | 0·7877 |
| $10^5$ | 0·0809 | 0·0862 | 0·1033 | 0·1042 | 0·1077 | 0·1244 | 0·1463 | | 0·1463 |
| $10^5$ | 0·1009 | 0·0964 | 0·0995 | 0·1234 | 0·2839 | 0·2199 | 0·2409 | | 0·2839 |
| $10^6$ | 0·2487 | 0·2378 | 0·2954 | 2·5102 | 1·3203 | 0·9238 | 0·8265 | | 2·5102 |
| $10^6$ | 0·2036 | 0·2579 | 0·3829 | 2·0417 | 1·1996 | 0·7454 | 0·4340 | 0·3820 | 2·0417 |
| $10^6$ | 0·7032 | 0·6544 | 0·8750 | 7·7320 | 5·0655 | 3·7722 | 2·3921 | 1·2175 | 7·7320 |
| $10^7$ | 0·4688 | 0·4671 | 0·4381 | 0·5281 | 0·5975 | 0·7482 | 0·5981 | 0·1927 | 0·7482 |
| $10^7$ | 0·3965 | 0·4883 | 0·4498 | 0·4878 | 0·5913 | 0·6205 | 0·5586 | 0·1227 | 0·6205 |
| $10^7$ | 0·1414 | 0·1231 | 0·1391 | 0·1863 | 0·1916 | 0·1672 | 0·1541 | 0·0677 | 0·1916 |
| $10^8$ | 0·1487 | 0·1438 | 0·1523 | 0·4715 | 0·7052 | 0·5336 | 0·7113 | 0·4438 | 0·7113 |
| $10^8$ | 0·2166 | 0·2025 | 0·2221 | 1·0638 | 1·0757 | 1·0950 | 0·5941 | 0·4704 | 1·0950 |
| $10^8$ | 0·1288 | 0·0918 | 0·2078 | 2·6963 | 2·8151 | 1·0140 | 0·8876 | 0·6960 | 2·8151 |
| $10^9$ | 1·4131 | 1·4954 | 3·1488 | 9·8135 | 6·7288 | 3·2378 | 2·9582 | 3·1486 | 9·8135 |
| $10^9$ | 0·0244 | 0·0321 | 0·0342 | 0·0373 | 0·0420 | 0·0396 | 0·0535 | | 0·0535 |
| $10^9$ | 0·0185 | 0·0201 | 0·0216 | 0·3092 | 0·1869 | 0·1736 | 0·1622 | 0·0960 | 0·3092 |
| $10^{10}$ | 0·0565 | 0·0616 | 0·0845 | 0·1218 | 0·1058 | | 0·1011 | 0·1718 | 0·1718 |
| $10^{10}$ | 0·4259 | 0·4580 | 0·4714 | 2·2594 | | | 0·9633 | 0·7051 | 2·2594 |
| $10^{10}$ | 0·1006 | 0·1022 | 0·2122 | 16·7337 | 12·3423 | 8·2762 | 4·9335 | 2·6583 | 16·7337 |

| Fold change CT-B IgG | | | | | | | | | |
|---|---|---|---|---|---|---|---|---|---|
| Dose (CFU) | D1 | D4 | D7 | D15 | D29 | D57 | D85 | D180 | max FC |
| $10^5$ | 1·00 | 0·93 | 1·01 | 4·06 | 4·26 | 3·14 | 2·88 | 1·85 | 4·26 |
| $10^5$ | 1·00 | 1·07 | 1·28 | 1·29 | 1·33 | 1·54 | 1·81 | | 1·81 |
| $10^5$ | 1·00 | 0·95 | 0·99 | 1·22 | 2·81 | 2·18 | 2·39 | | 2·81 |
| $10^6$ | 1·00 | 0·96 | 1·19 | 10·09 | 5·31 | 3·71 | 3·32 | | 10·09 |
| $10^6$ | 1·00 | 1·27 | 1·88 | 10·03 | 5·89 | 3·66 | 2·13 | 1·88 | 10·03 |
| $10^6$ | 1·00 | 0·93 | 1·24 | 11·00 | 7·20 | 5·36 | 3·40 | 1·73 | 11·00 |
| $10^7$ | 1·00 | 1·00 | 0·93 | 1·13 | 1·27 | 1·60 | 1·28 | 0·41 | 1·60 |
| $10^7$ | 1·00 | 1·23 | 1·13 | 1·23 | 1·49 | 1·56 | 1·41 | 0·31 | 1·56 |
| $10^7$ | 1·00 | 0·87 | 0·98 | 1·32 | 1·36 | 1·18 | 1·09 | 0·48 | 1·36 |
| $10^8$ | 1·00 | 0·97 | 1·02 | 3·17 | 4·74 | 3·59 | 4·78 | 2·98 | 4·78 |
| $10^8$ | 1·00 | 0·93 | 1·03 | 4·91 | 4·97 | 5·05 | 2·74 | 2·17 | 5·05 |
| $10^8$ | 1·00 | 0·71 | 1·61 | 20·93 | 21·85 | 7·87 | 6·89 | 5·40 | 21·85 |
| $10^9$ | 1·00 | 1·06 | 2·23 | 6·94 | 4·76 | 2·29 | 2·09 | 2·23 | 6·94 |
| $10^9$ | 1·00 | 1·31 | 1·40 | 1·53 | 1·72 | 1·62 | 2·19 | | 2·19 |
| $10^9$ | 1·00 | 1·08 | 1·17 | 16·71 | 10·10 | 9·38 | 8·76 | 5·19 | 16·71 |
| $10^{10}$ | 1·00 | 1·09 | 1·50 | 2·16 | 1·87 | | 1·79 | 3·04 | 3·04 |
| $10^{10}$ | 1·00 | 1·08 | 1·11 | 5·30 | | | 2·26 | 1·66 | 5·30 |
| $10^{10}$ | 1·00 | 1·02 | 2·11 | 166·32 | 122·68 | 82·26 | 49·04 | 26·42 | 166·32 |

**Dose- Expansion Module**

| CT-B IgG (RAU) | | | | | | | | | |
|---|---|---|---|---|---|---|---|---|---|
| Dose (CFU) | D1 | D4 | D7 | D15 | D29 | D57 | D85 | D180 | max |
| 0 | 0·0207 | 0·0221 | 0·0238 | 0·0219 | 0·0268 | 0·0220 | 0·0240 | 0·0253 | 0·0268 |
| 0 | 0·2358 | 0·2653 | 0·2537 | 0·2193 | 0·2353 | 0·2238 | 0·4874 | 0·3261 | 0·4874 |
| 0 | 0·1165 | 0·1143 | 0·1239 | 0·0971 | 0·1031 | 0·1504 | 0·1085 | 0·0127 | 0·1504 |
| 0 | 0·4453 | 0·4851 | 0·4485 | 0·4042 | 0·4928 | 0·2718 | 0·2181 | 0·2017 | 0·4928 |
| 0 | 0·0489 | 0·0454 | 0·0481 | 0·0486 | 0·0481 | 0·0394 | 0·0361 | 0·0318 | 0·0489 |
| 0 | 0·3057 | 0·3305 | 0·3172 | 0·2102 | 0·1737 | 0·2287 | 0·2015 | 0·2014 | 0·3305 |
| 0 | 0·0360 | 0·0292 | 0·0326 | 0·0344 | 0·0366 | 0·0279 | 0·0271 | 0·0319 | 0·0366 |
| $2\times10^7$ | 0·2704 | 0·2340 | 0·2596 | 0·9392 | 0·9417 | 0·7905 | 0·8374 | 0·7531 | 0·9417 |
| $2\times10^7$ | 0·1626 | 0·1855 | 0·2626 | 2·7783 | 1·7611 | 1·2094 | 0·8473 | 0·4446 | 2·7783 |
| $2\times10^7$ | 0·0162 | 0·0182 | 0·0147 | 0·0181 | 0·0168 | 0·0212 | 0·0217 | 0·0151 | 0·0217 |
| $2\times10^7$ | 0·2510 | 0·3196 | 0·2839 | 0·5873 | 0·5390 | 0·6067 | 0·3411 | 0·2853 | 0·6067 |
| $2\times10^7$ | 0·1200 | 0·1141 | 0·1027 | 0·3015 | 0·3656 | 0·3686 | 0·3377 | 0·1353 | 0·3686 |
| $2\times10^7$ | 0·1369 | 0·1239 | 0·1415 | 0·3064 | 0·3670 | 0·3559 | 0·3775 | 0·1573 | 0·3775 |
| $2\times10^7$ | 0·2377 | | 0·2680 | 0·2603 | 0·3696 | 0·4059 | 0·3165 | | 0·4059 |
| $2\times10^7$ | 0·2336 | 0·2500 | 0·3214 | 8·0040 | 6·4070 | 3·3133 | 1·2467 | 0·4366 | 8·0040 |
| $2\times10^7$ | 0·0452 | 0·0457 | 0·0434 | 0·0481 | 0·0755 | 0·0584 | 0·0480 | 0·0463 | 0·0755 |
| $2\times10^7$ | 0·3861 | 0·3261 | 0·4124 | 0·3529 | 0·2190 | 0·2078 | 0·1894 | 0·1526 | 0·4124 |
| $2\times10^7$ | 0·2151 | 0·2047 | 0·5198 | 2·4724 | 3·1328 | 1·4386 | 1·0090 | 0·9747 | 3·1328 |
| $2\times10^7$ | 0·1236 | 0·1224 | 0·1628 | 0·3996 | 0·4890 | 0·3307 | 0·3532 | 0·2204 | 0·4890 |
| $2\times10^7$ | 0·0412 | 0·0526 | 0·0512 | 0·0488 | 0·0577 | 0·0539 | 0·0516 | 0·0577 | 0·0577 |
| $2\times10^7$ | 0·0249 | 0·0240 | 0·0943 | 0·2416 | 0·2546 | 0·1883 | | 0·0766 | 0·2546 |
| $2\times10^8$ | 0·3399 | 0·3559 | 0·4152 | 0·4572 | 0·6718 | 0·7432 | 0·7926 | 0·5454 | 0·7926 |
| $2\times10^8$ | 0·0527 | 0·0523 | 0·0591 | 0·3493 | 0·2960 | 0·2615 | 0·1986 | 0·1038 | 0·3493 |
| $2\times10^8$ | 0·3011 | 0·3274 | 0·5401 | 0·8919 | 0·6974 | 0·5313 | 0·2221 | 0·2277 | 0·8919 |
| $2\times10^8$ | 0·2948 | 0·2736 | 0·4662 | 3·4480 | 3·2304 | 1·7476 | 0·9586 | 0·3630 | 3·4480 |
| $2\times10^8$ | 0·4245 | 0·4202 | 1·4819 | 14·5274 | 10·4192 | 1·8872 | 1·0869 | 0·4679 | 14·5274 |
| $2\times10^8$ | 0·2681 | 0·2411 | 0·4389 | 0·9189 | | | | | 0·9189 |
| $2\times10^8$ | 0·6990 | 0·8113 | 1·1032 | 36·8765 | 38·9655 | 34·6795 | 17·6769 | 1·9164 | 38·9655 |
| $2\times10^8$ | 0·0700 | 0·0573 | 0·0662 | 0·1361 | 0·2172 | 0·2038 | 0·2489 | 0·1916 | 0·2489 |
| $2\times10^8$ | 0·0327 | 0·0243 | 0·0240 | 0·0396 | 0·1132 | 0·1535 | 0·1436 | 0·1348 | 0·1535 |
| $2\times10^8$ | 0·1108 | 0·1329 | 0·3141 | 3·2014 | 1·8031 | 0·4703 | 0·2616 | 0·1984 | 3·2014 |
| $2\times10^8$ | 0·2612 | 0·2495 | 0·2437 | 0·2646 | 0·2605 | 0·2694 | 0·2644 | 0·2891 | 0·2891 |
| $2\times10^8$ | 0·0527 | 0·0486 | 0·0443 | 0·0367 | 0·0394 | | 0·0462 | 0·1069 | 0·1069 |
| $2\times10^8$ | 0·1031 | 0·0854 | 0·1343 | 0·2188 | 0·2080 | 0·1747 | 0·1543 | 0·1597 | 0·2188 |

| Fold change CT-B IgG | | | | | | | | | |
|---|---|---|---|---|---|---|---|---|---|
| Dose (CFU) | D1 | D4 | D7 | D15 | D29 | D57 | D85 | D180 | max FC |
| 0 | 1·00 | 1·07 | 1·15 | 1·06 | 1·30 | 1·06 | 1·16 | 1·22 | 1·30 |
| 0 | 1·00 | 1·12 | 1·08 | 0·93 | 1·00 | 0·95 | 2·07 | 1·38 | 2·07 |
| 0 | 1·00 | 0·98 | 1·06 | 0·83 | 0·89 | 1·29 | 0·93 | 0·11 | 1·29 |
| 0 | 1·00 | 1·09 | 1·01 | 0·91 | 1·11 | 0·61 | 0·49 | 0·45 | 1·11 |
| 0 | 1·00 | 0·93 | 0·98 | 0·99 | 0·98 | 0·81 | 0·74 | 0·65 | 0·99 |
| 0 | 1·00 | 1·08 | 1·04 | 0·69 | 0·57 | 0·75 | 0·66 | 0·66 | 1·08 |
| 0 | 1·00 | 0·81 | 0·91 | 0·96 | 1·02 | 0·78 | 0·75 | 0·89 | 1·02 |
| $2\times10^7$ | 1·00 | 0·87 | 0·96 | 3·47 | 3·48 | 2·92 | 3·10 | 2·79 | 3·48 |
| $2\times10^7$ | 1·00 | 1·14 | 1·62 | 17·09 | 10·83 | 7·44 | 5·21 | 2·73 | 17·09 |
| $2\times10^7$ | 1·00 | 1·12 | 0·91 | 1·12 | 1·04 | 1·31 | 1·34 | 0·93 | 1·34 |
| $2\times10^7$ | 1·00 | 1·27 | 1·13 | 2·34 | 2·15 | 2·42 | 1·36 | 1·14 | 2·42 |
| $2\times10^7$ | 1·00 | 0·95 | 0·86 | 2·51 | 3·05 | 3·07 | 2·81 | 1·13 | 3·07 |
| $2\times10^7$ | 1·00 | 0·91 | 1·03 | 2·24 | 2·68 | 2·60 | 2·76 | 1·15 | 2·76 |
| $2\times10^7$ | 1·00 | | 1·13 | 1·10 | 1·56 | 1·71 | 1·33 | | 1·71 |
| $2\times10^7$ | 1·00 | 1·07 | 1·38 | 34·26 | 27·43 | 14·18 | 5·34 | 1·87 | 34·26 |
| $2\times10^7$ | 1·00 | 1·01 | 0·96 | 1·06 | 1·67 | 1·29 | 1·06 | 1·02 | 1·67 |
| $2\times10^7$ | 1·00 | 0·84 | 1·07 | 0·91 | 0·57 | 0·54 | 0·49 | 0·40 | 1·07 |
| $2\times10^7$ | 1·00 | 0·95 | 2·42 | 11·49 | 14·56 | 6·69 | 4·69 | 4·53 | 14·56 |
| $2\times10^7$ | 1·00 | 0·99 | 1·32 | 3·23 | 3·96 | 2·68 | 2·86 | 1·78 | 3·96 |
| $2\times10^7$ | 1·00 | 1·28 | 1·24 | 1·18 | 1·40 | 1·31 | 1·25 | 1·40 | 1·40 |
| $2\times10^7$ | 1·00 | 0·96 | 3·79 | 9·70 | 10·22 | 7·56 | | 3·07 | 10·22 |
| $2\times10^8$ | 1·00 | 1·05 | 1·22 | 1·34 | 1·98 | 2·19 | 2·33 | 1·60 | 2·33 |
| $2\times10^8$ | 1·00 | 0·99 | 1·12 | 6·63 | 5·62 | 4·96 | 3·77 | 1·97 | 6·63 |
| $2\times10^8$ | 1·00 | 1·09 | 1·79 | 2·96 | 2·32 | 1·76 | 0·74 | 0·76 | 2·96 |
| $2\times10^8$ | 1·00 | 0·93 | 1·58 | 11·70 | 10·96 | 5·93 | 3·25 | 1·23 | 11·70 |
| $2\times10^8$ | 1·00 | 0·99 | 3·49 | 34·22 | 24·54 | 4·45 | 2·56 | 1·10 | 34·22 |
| $2\times10^8$ | 1·00 | 0·90 | 1·64 | 3·43 | | | | | 3·43 |
| $2\times10^8$ | 1·00 | 1·16 | 1·58 | 52·75 | 55·74 | 49·61 | 25·29 | 2·74 | 55·74 |
| $2\times10^8$ | 1·00 | 0·82 | 0·95 | 1·95 | 3·10 | 2·91 | 3·56 | 2·74 | 3·56 |
| $2\times10^8$ | 1·00 | 0·74 | 0·73 | 1·21 | 3·46 | 4·69 | 4·39 | 4·12 | 4·69 |
| $2\times10^8$ | 1·00 | 1·20 | 2·83 | 28·89 | 16·27 | 4·24 | 2·36 | 1·79 | 28·89 |
| $2\times10^8$ | 1·00 | 0·95 | 0·93 | 1·01 | 1·00 | 1·03 | 1·01 | 1·11 | 1·11 |
| $2\times10^8$ | 1·00 | 0·92 | 0·84 | 0·70 | 0·75 | | 0·88 | 2·03 | 2·03 |
| $2\times10^8$ | 1·00 | 0·83 | 1·30 | 2·12 | 2·02 | 1·69 | 1·50 | 1·55 | 2·12 |

| | RAU | | |
|---|---|---|---|
| | D1 | D39 | |
| *V. cholerae* | 0·2551 | 41·3854 | |
| | 1·9926 | 50·1770 | |
| | 3·6006 | 83·0713 | |
| | 0·1204 | 5·9248 | |
| | 0·4858 | 31·1167 | |
| | D1 | D11 | |
| Vaxchora D11 Challenge | 0·1724 | 1·4968 | |
| | 0·2182 | 0·2640 | |
| | 1·3189 | 23·1666 | |
| | 0·3618 | 0·3522 | |
| | 0·1503 | 0·1580 | |
| | 1·2351 | 0·9401 | |
| | D1 | D11 | D91 |
| Vaxchora D91 Challenge | 1·9207 | 2·6394 | 2·2632 |
| | 0·0626 | 0·0805 | 0·3951 |
| | 0·2442 | 1·4890 | 0·8181 |
| | 0·2854 | 0·3099 | 1·4233 |
| | 0·0554 | 7·8103 | 1·0327 |
| | 0·0950 | 0·3171 | 0·2646 |

| | RAU fold change | | | max FC |
|---|---|---|---|---|
| | D1 | D39 | | |
| *V. cholerae* | 1·00 | 162·24 | | 162·24 |
| | 1·00 | 25·18 | | 25·18 |
| | 1·00 | 23·07 | | 23·07 |
| | 1·00 | 49·21 | | 49·21 |
| | 1·00 | 64·06 | | 64·06 |
| | D1 | D11 | | |
| Vaxchora D11 Challenge | 1·00 | 8·68 | | 8·68 |
| | 1·00 | 1·21 | | 1·21 |
| | 1·00 | 17·56 | | 17·56 |
| | 1·00 | 0·97 | | 0·97 |
| | 1·00 | 1·05 | | 1·05 |
| | 1·00 | 0·76 | | 0·76 |
| | D1 | D11 | D91 | |
| Vaxchora D91 Challenge | 1·00 | 1·37 | 1·18 | 1·37 |
| | 1·00 | 1·29 | 6·31 | 6·31 |
| | 1·00 | 6·10 | 3·35 | 6·10 |
| | 1·00 | 1·09 | 4·99 | 4·99 |
| | 1·00 | 140·90 | 18·63 | 140·90 |
| | 1·00 | 3·34 | 2·78 | 3·34 |

**Dose-Escalation Module**

| CT-B IgA (RAU) | | | | | | | | | |
|---|---|---|---|---|---|---|---|---|---|
| **Dose (CFU)** | **D1** | **D4** | **D7** | **D15** | **D29** | **D57** | **D85** | **D180** | **max** |
| $10^5$ | 0·3726 | 0·3573 | 0·3412 | 2·5297 | 2·1222 | 1·1656 | 0·8222 | 0·5404 | 2·5297 |
| $10^5$ | 0·2759 | 0·2602 | 0·2710 | 0·2134 | 0·2405 | 0·2090 | 0·2694 | | 0·2759 |
| $10^5$ | 0·1653 | 0·1839 | 0·1947 | 0·2181 | 0·3444 | 0·2170 | 0·1917 | | 0·3444 |
| $10^6$ | 0·1793 | 0·2019 | 0·2462 | 1·4832 | 0·4823 | 0·3117 | 0·2598 | | 1·4832 |
| $10^6$ | 0·8621 | 0·6931 | 1·8598 | 5·9258 | 1·6387 | 0·8949 | 0·8354 | 0·7132 | 5·9258 |
| $10^6$ | 0·5409 | 0·7933 | 0·9187 | 5·4065 | 1·1732 | 0·8498 | 0·5125 | 0·5972 | 5·4065 |
| $10^7$ | 0·2508 | 0·2357 | 0·2379 | 0·5625 | 0·6848 | 0·6282 | 0·6772 | 0·3690 | 0·6848 |
| $10^7$ | 0·1069 | 0·0967 | 0·1018 | 0·1395 | 0·3223 | 0·2103 | 0·1928 | 0·1389 | 0·3223 |
| $10^7$ | 0·0797 | 0·0665 | 0·0703 | 0·0983 | 0·0887 | 0·0726 | 0·0766 | 0·0437 | 0·0983 |
| $10^8$ | 0·6446 | 0·7413 | 0·6444 | 16·0204 | 10·5840 | 5·1290 | 5·7179 | 4·4060 | 16·0204 |
| $10^8$ | 0·1603 | 0·1373 | 0·1451 | 0·5641 | 0·5567 | 0·2918 | 0·1614 | 0·1586 | 0·5641 |
| $10^8$ | 0·0869 | 0·0785 | 0·6308 | 3·1965 | 3·8580 | 1·0644 | 0·9288 | 0·8963 | 3·8580 |
| $10^9$ | 0·3562 | 0·3343 | 1·6580 | 2·3682 | 0·9774 | 0·5806 | 0·4668 | 0·4780 | 2·3682 |
| $10^9$ | 0·2926 | 0·2401 | 0·2452 | 0·1565 | 0·1601 | 0·1327 | 0·2276 | | 0·2926 |
| $10^9$ | 0·1338 | 0·1383 | 0·1279 | 3·6413 | 2·0362 | 1·6801 | 1·9280 | 0·9348 | 3·6413 |
| $10^{10}$ | 0·1308 | 0·1225 | 0·1451 | 0·1802 | 0·1769 | | 0·1501 | 0·1663 | 0·1802 |
| $10^{10}$ | 0·3379 | 0·3811 | 0·8379 | 2·9181 | | | 0·6284 | 0·5550 | 2·9181 |
| $10^{10}$ | 0·0146 | 0·0158 | 0·0614 | 1·2045 | 0·3742 | 0·2164 | 0·1506 | 0·1038 | 1·2045 |

| Fold change CT-B IgA | | | | | | | | | |
|---|---|---|---|---|---|---|---|---|---|
| **Dose (CFU)** | **D1** | **D4** | **D7** | **D15** | **D29** | **D57** | **D85** | **D180** | **max FC** |
| $10^5$ | 1·00 | 0·96 | 0·92 | 6·79 | 5·70 | 3·13 | 2·21 | 1·45 | 6·79 |
| $10^5$ | 1·00 | 0·94 | 0·98 | 0·77 | 0·87 | 0·76 | 0·98 | | 0·98 |
| $10^5$ | 1·00 | 1·11 | 1·18 | 1·32 | 2·08 | 1·31 | 1·16 | | 2·08 |
| $10^6$ | 1·00 | 1·13 | 1·37 | 8·27 | 2·69 | 1·74 | 1·45 | | 8·27 |
| $10^6$ | 1·00 | 0·80 | 2·16 | 6·87 | 1·90 | 1·04 | 0·97 | 0·83 | 6·87 |
| $10^6$ | 1·00 | 1·47 | 1·70 | 10·00 | 2·17 | 1·57 | 0·95 | 1·10 | 10·00 |
| $10^7$ | 1·00 | 0·94 | 0·95 | 2·24 | 2·73 | 2·50 | 2·70 | 1·47 | 2·73 |
| $10^7$ | 1·00 | 0·90 | 0·95 | 1·31 | 3·01 | 1·97 | 1·80 | 1·30 | 3·01 |
| $10^7$ | 1·00 | 0·83 | 0·88 | 1·23 | 1·11 | 0·91 | 0·96 | 0·55 | 1·23 |
| $10^8$ | 1·00 | 1·15 | 1·00 | 24·85 | 16·42 | 7·96 | 8·87 | 6·83 | 24·85 |
| $10^8$ | 1·00 | 0·86 | 0·91 | 3·52 | 3·47 | 1·82 | 1·01 | 0·99 | 3·52 |
| $10^8$ | 1·00 | 0·90 | 7·26 | 36·79 | 44·41 | 12·25 | 10·69 | 10·32 | 44·41 |
| $10^9$ | 1·00 | 0·94 | 4·66 | 6·65 | 2·74 | 1·63 | 1·31 | 1·34 | 6·65 |
| $10^9$ | 1·00 | 0·82 | 0·84 | 0·53 | 0·55 | 0·45 | 0·78 | | 0·84 |
| $10^9$ | 1·00 | 1·03 | 0·96 | 27·22 | 15·22 | 12·56 | 14·41 | 6·99 | 27·22 |
| $10^{10}$ | 1·00 | 0·94 | 1·11 | 1·38 | 1·35 | | 1·15 | 1·27 | 1·38 |
| $10^{10}$ | 1·00 | 1·13 | 2·48 | 8·64 | | | 1·86 | 1·64 | 8·64 |
| $10^{10}$ | 1·00 | 1·08 | 4·21 | 82·55 | 25·64 | 14·83 | 10·32 | 7·11 | 82·55 |

**Dose- Expansion Module**

| CT-B IgA (RAU) | | | | | | | | | |
|---|---|---|---|---|---|---|---|---|---|
| **Dose (CFU)** | **D1** | **D4** | **D7** | **D15** | **D29** | **D57** | **D85** | **D180** | **max** |
| 0 | 0·3728 | 0·3966 | 0·4158 | 0·4068 | 0·3949 | 0·3202 | 0·3370 | 0·3271 | 0·4158 |
| 0 | 0·3116 | 0·2779 | 0·2510 | 0·2603 | 0·2676 | 0·2388 | 0·7611 | 0·3122 | 0·7611 |
| 0 | 0·0940 | 0·1096 | 0·1058 | 0·0716 | 0·0914 | 0·0889 | 0·0848 | 0·0712 | 0·1096 |
| 0 | 0·6728 | 0·5666 | 0·6639 | 0·5694 | 0·4127 | 0·6243 | 0·4509 | 0·3932 | 0·6728 |
| 0 | 0·1500 | 0·1510 | 0·2092 | 0·1716 | 0·1543 | 0·2276 | 0·1799 | 0·1639 | 0·2276 |
| 0 | 0·1926 | 0·1904 | 0·1540 | 0·0656 | 0·0573 | 0·0667 | 0·0643 | 0·2580 | 0·2580 |
| 0 | 0·0355 | 0·0270 | 0·0302 | 0·0305 | 0·0421 | 0·0261 | 0·0135 | 0·0264 | 0·0421 |
| $2\times10^7$ | 0·1154 | 0·1039 | 0·1858 | 0·6233 | 0·3020 | 0·2182 | 0·1928 | 0·1875 | 0·6233 |
| $2\times10^7$ | 0·1141 | 0·1226 | 0·1998 | 0·9696 | 0·4559 | 0·2034 | 0·1781 | 0·1269 | 0·9696 |
| $2\times10^7$ | 0·1136 | 0·1118 | 0·0946 | 0·0988 | 0·0876 | 0·0946 | 0·1052 | 0·0868 | 0·1136 |
| $2\times10^7$ | 0·4119 | 0·4771 | 0·4350 | 0·5776 | 0·6048 | 0·6290 | 0·5024 | 0·2629 | 0·6290 |
| $2\times10^7$ | 0·3069 | 0·2525 | 0·2950 | 0·7328 | 0·5152 | 0·3268 | 0·2505 | 0·0800 | 0·7328 |
| $2\times10^7$ | 0·7462 | 0·8355 | 1·1789 | 1·7153 | 1·9611 | 1·4536 | 1·1870 | 0·4946 | 1·9611 |
| $2\times10^7$ | 3·1450 | | 2·4463 | 1·9511 | 2·4246 | 2·3152 | 2·9074 | | 3·1450 |
| $2\times10^7$ | 0·2049 | 0·2349 | 0·6504 | 8·3940 | 3·5016 | 1·1330 | 0·7455 | 0·4749 | 8·3940 |
| $2\times10^7$ | 0·1568 | 0·1670 | 0·1982 | 0·2319 | 0·2881 | 0·1776 | 0·1876 | 0·2153 | 0·2881 |
| $2\times10^7$ | 0·1900 | 0·1703 | 0·2023 | 0·2132 | 0·1562 | 0·1211 | 0·1049 | 0·1087 | 0·2132 |
| $2\times10^7$ | 0·2452 | 0·1704 | 0·5261 | 0·7290 | 0·8199 | 0·5833 | 0·5292 | 0·5925 | 0·8199 |
| $2\times10^7$ | 0·2642 | 0·2891 | 0·5162 | 1·7751 | 1·0531 | 0·5679 | 0·7933 | 0·5750 | 1·7751 |
| $2\times10^7$ | 0·0909 | 0·0769 | 0·1418 | 0·1846 | 0·1701 | 0·1403 | 0·1116 | 0·0972 | 0·1846 |
| $2\times10^7$ | 0·1108 | 0·1105 | 0·1421 | 0·1156 | 0·1217 | 0·1351 | | 0·0806 | 0·1421 |
| $2\times10^8$ | 0·4062 | 0·3961 | 0·3912 | 0·4350 | 0·5119 | 0·4048 | 0·3949 | 0·4537 | 0·5119 |
| $2\times10^8$ | 0·3935 | 0·5003 | 0·4594 | 1·8997 | 1·1156 | 0·7056 | 0·5847 | 0·3994 | 1·8997 |
| $2\times10^8$ | 0·2841 | 0·2746 | 0·5777 | 0·7595 | 0·4710 | 0·3310 | 0·1541 | 0·2079 | 0·7595 |
| $2\times10^8$ | 0·2052 | 0·1901 | 0·9017 | 2·9997 | 2·0807 | 0·9570 | 0·5812 | 0·2518 | 2·9997 |
| $2\times10^8$ | 0·1871 | 0·2304 | 1·0560 | 1·7897 | 0·9562 | 0·2732 | 0·1817 | 0·3731 | 1·7897 |
| $2\times10^8$ | 0·2684 | 0·2030 | 0·3895 | 0·4358 | | | | | 0·4358 |
| $2\times10^8$ | 0·1314 | 0·1533 | 0·5163 | 3·4261 | 0·5408 | 0·3038 | 0·2163 | 0·1358 | 3·4261 |
| $2\times10^8$ | 0·1947 | 0·1907 | 0·1785 | 0·2697 | 0·2964 | 0·2451 | 0·2574 | 0·2815 | 0·2964 |
| $2\times10^8$ | 0·5973 | 0·6631 | 0·5716 | 0·3010 | 1·1680 | 0·7038 | 0·5513 | 0·3865 | 1·1680 |
| $2\times10^8$ | 0·1867 | 0·2081 | 2·9560 | 14·7954 | 4·2018 | 0·4436 | 0·3394 | 0·2601 | 14·7954 |
| $2\times10^8$ | 0·2455 | 0·2626 | 0·2397 | 0·2603 | 0·2124 | 0·2594 | 0·2133 | 0·2298 | 0·2626 |
| $2\times10^8$ | 0·3270 | 0·3223 | 0·2473 | 0·1782 | 0·2273 | | 0·2230 | 0·3109 | 0·3270 |
| $2\times10^8$ | 0·0478 | 0·0356 | 0·0594 | 0·0763 | 0·0711 | 0·0727 | 0·0640 | 0·0978 | 0·0978 |

| Fold change CT-B IgA | | | | | | | | | |
|---|---|---|---|---|---|---|---|---|---|
| **Dose (CFU)** | **D1** | **D4** | **D7** | **D15** | **D29** | **D57** | **D85** | **D180** | **max FC** |
| 0 | 1·00 | 1·06 | 1·12 | 1·09 | 1·06 | 0·86 | 0·90 | 0·88 | 1·12 |
| 0 | 1·00 | 0·89 | 0·81 | 0·84 | 0·86 | 0·77 | 2·44 | 1·00 | 2·44 |
| 0 | 1·00 | 1·17 | 1·13 | 0·76 | 0·97 | 0·95 | 0·90 | 0·76 | 1·17 |
| 0 | 1·00 | 0·84 | 0·99 | 0·85 | 0·61 | 0·93 | 0·67 | 0·58 | 0·99 |
| 0 | 1·00 | 1·01 | 1·40 | 1·14 | 1·03 | 1·52 | 1·20 | 1·09 | 1·52 |
| 0 | 1·00 | 0·99 | 0·80 | 0·34 | 0·30 | 0·35 | 0·33 | 1·34 | 1·34 |
| 0 | 1·00 | 0·76 | 0·85 | 0·86 | 1·18 | 0·73 | 0·38 | 0·74 | 1·18 |
| $2\times10^7$ | 1·00 | 0·90 | 1·61 | 5·40 | 2·62 | 1·89 | 1·67 | 1·63 | 5·40 |
| $2\times10^7$ | 1·00 | 1·07 | 1·75 | 8·50 | 4·00 | 1·78 | 1·56 | 1·11 | 8·50 |
| $2\times10^7$ | 1·00 | 0·98 | 0·83 | 0·87 | 0·77 | 0·83 | 0·93 | 0·76 | 0·98 |
| $2\times10^7$ | 1·00 | 1·16 | 1·06 | 1·40 | 1·47 | 1·53 | 1·22 | 0·64 | 1·53 |
| $2\times10^7$ | 1·00 | 0·82 | 0·96 | 2·39 | 1·68 | 1·06 | 0·82 | 0·26 | 2·39 |
| $2\times10^7$ | 1·00 | 1·12 | 1·58 | 2·30 | 2·63 | 1·95 | 1·59 | 0·66 | 2·63 |
| $2\times10^7$ | 1·00 | | 0·78 | 0·62 | 0·77 | 0·74 | 0·92 | | 0·92 |
| $2\times10^7$ | 1·00 | 1·15 | 3·17 | 40·97 | 17·09 | 5·53 | 3·64 | 2·32 | 40·97 |
| $2\times10^7$ | 1·00 | 1·07 | 1·26 | 1·48 | 1·84 | 1·13 | 1·20 | 1·37 | 1·84 |
| $2\times10^7$ | 1·00 | 0·90 | 1·06 | 1·12 | 0·82 | 0·64 | 0·55 | 0·57 | 1·12 |
| $2\times10^7$ | 1·00 | 0·70 | 2·15 | 2·97 | 3·34 | 2·38 | 2·16 | 2·42 | 3·34 |
| $2\times10^7$ | 1·00 | 1·09 | 1·95 | 6·72 | 3·99 | 2·15 | 3·00 | 2·18 | 6·72 |
| $2\times10^7$ | 1·00 | 0·85 | 1·56 | 2·03 | 1·87 | 1·54 | 1·23 | 1·07 | 2·03 |
| $2\times10^7$ | 1·00 | 1·00 | 1·28 | 1·04 | 1·10 | 1·22 | | 0·73 | 1·28 |
| $2\times10^8$ | 1·00 | 0·98 | 0·96 | 1·07 | 1·26 | 1·00 | 0·97 | 1·12 | 1·26 |
| $2\times10^8$ | 1·00 | 1·27 | 1·17 | 4·83 | 2·84 | 1·79 | 1·49 | 1·01 | 4·83 |
| $2\times10^8$ | 1·00 | 0·97 | 2·03 | 2·67 | 1·66 | 1·17 | 0·54 | 0·73 | 2·67 |
| $2\times10^8$ | 1·00 | 0·93 | 4·40 | 14·62 | 10·14 | 4·66 | 2·83 | 1·23 | 14·62 |
| $2\times10^8$ | 1·00 | 1·23 | 5·65 | 9·57 | 5·11 | 1·46 | 0·97 | 1·99 | 9·57 |
| $2\times10^8$ | 1·00 | 0·76 | 1·45 | 1·62 | | | | | 1·62 |
| $2\times10^8$ | 1·00 | 1·17 | 3·93 | 26·07 | 4·12 | 2·31 | 1·65 | 1·03 | 26·07 |
| $2\times10^8$ | 1·00 | 0·98 | 0·92 | 1·38 | 1·52 | 1·26 | 1·32 | 1·45 | 1·52 |
| $2\times10^8$ | 1·00 | 1·11 | 0·96 | 0·50 | 1·96 | 1·18 | 0·92 | 0·65 | 1·96 |
| $2\times10^8$ | 1·00 | 1·11 | 15·83 | 79·23 | 22·50 | 2·38 | 1·82 | 1·39 | 79·23 |
| $2\times10^8$ | 1·00 | 1·07 | 0·98 | 1·06 | 0·87 | 1·06 | 0·87 | 0·94 | 1·07 |
| $2\times10^8$ | 1·00 | 0·99 | 0·76 | 0·55 | 0·70 | | 0·68 | 0·95 | 0·99 |
| $2\times10^8$ | 1·00 | 0·74 | 1·24 | 1·59 | 1·49 | 1·52 | 1·34 | 2·04 | 2·04 |

| | RAU | | |
|---|---|---|---|
| | D1 | D39 | |
| *V. cholerae* | 0·4752 | 165·3919 | |
| | 0·4201 | 25·5035 | |
| | 1·0947 | 28·0699 | |
| | 0·1965 | 2·3643 | |
| | 0·7002 | 28·4187 | |
| | D1 | D11 | |
| Vaxchora D11 Challenge | 0·3642 | 1·1842 | |
| | 0·8355 | 0·9830 | |
| | 1·0063 | 5·9184 | |
| | 0·3685 | 0·4190 | |
| | 0·3632 | 0·3457 | |
| | 0·5450 | 0·2240 | |
| | D1 | D11 | D91 |
| Vaxchora D91 Challenge | 6·0973 | 4·8230 | 3·8725 |
| | 0·1831 | 0·2750 | 0·2698 |
| | 0·2542 | 2·7882 | 0·4546 |
| | 0·2714 | 0·3339 | 0·6998 |
| | 1·8627 | 9·0015 | 2·1024 |
| | 0·2352 | 0·8027 | 0·2866 |

| | RAU fold change | | | max FC |
|---|---|---|---|---|
| | D1 | D39 | | |
| *V. cholerae* | 1·00 | 348·05 | | 348·05 |
| | 1·00 | 60·71 | | 60·71 |
| | 1·00 | 25·64 | | 25·64 |
| | 1·00 | 12·03 | | 12·03 |
| | 1·00 | 40·59 | | 40·59 |
| | D1 | D11 | | |
| Vaxchora D11 Challenge | 1·00 | 3·25 | | 3·25 |
| | 1·00 | 1·18 | | 1·18 |
| | 1·00 | 5·88 | | 5·88 |
| | 1·00 | 1·14 | | 1·14 |
| | 1·00 | 0·95 | | 0·95 |
| | 1·00 | 0·41 | | 0·41 |
| | D1 | D11 | D91 | |
| Vaxchora D91 Challenge | 1·00 | 0·79 | 0·64 | 0·79 |
| | 1·00 | 1·50 | 1·47 | 1·50 |
| | 1·00 | 10·97 | 1·79 | 10·97 |
| | 1·00 | 1·23 | 2·58 | 2·58 |
| | 1·00 | 4·83 | 1·13 | 4·83 |
| | 1·00 | 3·41 | 1·22 | 3·41 |

**Dose-Escalation Module**

| TcpA IgM (RAU) | | | | | | | | | |
|---|---|---|---|---|---|---|---|---|---|
| **Dose (CFU)** | **D1** | **D4** | **D7** | **D15** | **D29** | **D57** | **D85** | **D180** | **max** |
| $10^5$ | 5·2587 | 4·1776 | 5·9419 | 6·4290 | 4·9831 | 5·8443 | 3·8355 | 7·0887 | 7·0887 |
| $10^5$ | 3·2086 | 3·3258 | 3·9082 | 5·0454 | 3·6554 | 2·3154 | 3·3398 | | 5·0454 |
| $10^5$ | 3·9082 | 3·6478 | 4·6058 | 7·9350 | 5·8322 | 4·2920 | 4·0156 | | 7·9350 |
| $10^6$ | 3·8139 | 3·7482 | 3·2251 | 3·8981 | 4·0527 | 4·3650 | 4·6891 | | 4·6891 |
| $10^6$ | 10·1334 | 11·8599 | 14·2736 | 18·0176 | 10·1001 | 11·3339 | 10·1667 | 7·4940 | 18·0176 |
| $10^6$ | 2·3649 | 1·9469 | 3·2322 | 4·0306 | 2·6343 | 2·2290 | 2·8312 | 1·3580 | 4·0306 |
| $10^7$ | 5·1107 | 4·8071 | 6·1355 | 7·8613 | 5·9781 | 6·2450 | 7·5928 | 2·8706 | 7·8613 |
| $10^7$ | 6·4042 | 6·6429 | 4·5604 | 4·6209 | 4·9257 | 5·4379 | 24·6632 | 3·4505 | 24·6632 |
| $10^7$ | 6·0748 | 6·6802 | 5·1145 | 4·9949 | 5·8299 | 8·2963 | 7·6187 | 2·9487 | 8·2963 |
| $10^8$ | 2·0165 | 1·9573 | 1·3347 | 1·4026 | 1·6218 | 1·7654 | 1·7934 | 0·8548 | 2·0165 |
| $10^8$ | 8·3273 | 7·6748 | 7·7831 | 11·0424 | 8·1857 | 7·8788 | 3·4066 | 3·9152 | 11·0424 |
| $10^8$ | 5·1920 | 4·7919 | 8·9064 | 17·9204 | 11·5381 | 4·0667 | 3·7725 | 6·0999 | 17·9204 |
| $10^9$ | 5·3166 | 5·0991 | 6·6636 | 5·8020 | 5·7541 | 1·8579 | 2·0096 | 2·3369 | 6·6636 |
| $10^9$ | 6·1314 | 5·2426 | 3·1252 | 1·7204 | 2·7225 | 1·8108 | 5·5545 | | 6·1314 |
| $10^9$ | 3·5978 | 3·6459 | 3·6719 | 3·9039 | 3·4249 | 4·0895 | 4·2119 | 3·9378 | 4·2119 |
| $10^{10}$ | 2·5652 | 3·7800 | 4·1621 | 5·3890 | 5·7135 | | 3·1743 | 3·4249 | 5·7135 |
| $10^{10}$ | 3·7240 | 4·8458 | 2·6105 | 3·2828 | | | 4·0211 | 3·2429 | 4·8458 |
| $10^{10}$ | 2·0740 | 2·3989 | 2·9772 | 14·7887 | 6·8902 | 3·4249 | 7·1388 | 3·3300 | 14·7887 |

| Fold change TcpA IgM | | | | | | | | | |
|---|---|---|---|---|---|---|---|---|---|
| **Dose (CFU)** | **D1** | **D4** | **D7** | **D15** | **D29** | **D57** | **D85** | **D180** | **max FC** |
| $10^5$ | 1·00 | 0·79 | 1·13 | 1·22 | 0·95 | 1·11 | 0·73 | 1·35 | 1·35 |
| $10^5$ | 1·00 | 1·04 | 1·22 | 1·57 | 1·14 | 0·72 | 1·04 | | 1·57 |
| $10^5$ | 1·00 | 0·93 | 1·18 | 2·03 | 1·49 | 1·10 | 1·03 | | 2·03 |
| $10^6$ | 1·00 | 0·98 | 0·85 | 1·02 | 1·06 | 1·14 | 1·23 | | 1·23 |
| $10^6$ | 1·00 | 1·17 | 1·41 | 1·78 | 1·00 | 1·12 | 1·00 | 0·74 | 1·78 |
| $10^6$ | 1·00 | 0·82 | 1·37 | 1·70 | 1·11 | 0·94 | 1·20 | 0·57 | 1·70 |
| $10^7$ | 1·00 | 0·94 | 1·20 | 1·54 | 1·17 | 1·22 | 1·49 | 0·56 | 1·54 |
| $10^7$ | 1·00 | 1·04 | 0·71 | 0·72 | 0·77 | 0·85 | 3·85 | 0·54 | 3·85 |
| $10^7$ | 1·00 | 1·10 | 0·84 | 0·82 | 0·96 | 1·37 | 1·25 | 0·49 | 1·37 |
| $10^8$ | 1·00 | 0·97 | 0·66 | 0·70 | 0·80 | 0·88 | 0·89 | 0·42 | 0·97 |
| $10^8$ | 1·00 | 0·92 | 0·93 | 1·33 | 0·98 | 0·95 | 0·41 | 0·47 | 1·33 |
| $10^8$ | 1·00 | 0·92 | 1·72 | 3·45 | 2·22 | 0·78 | 0·73 | 1·17 | 3·45 |
| $10^9$ | 1·00 | 0·96 | 1·25 | 1·09 | 1·08 | 0·35 | 0·38 | 0·44 | 1·25 |
| $10^9$ | 1·00 | 0·86 | 0·51 | 0·28 | 0·44 | 0·30 | 0·91 | | 0·91 |
| $10^9$ | 1·00 | 1·01 | 1·02 | 1·09 | 0·95 | 1·14 | 1·17 | 1·09 | 1·17 |
| $10^{10}$ | 1·00 | 1·47 | 1·62 | 2·10 | 2·23 | | 1·24 | 1·34 | 2·23 |
| $10^{10}$ | 1·00 | 1·30 | 0·70 | 0·88 | | | 1·08 | 0·87 | 1·30 |
| $10^{10}$ | 1·00 | 1·16 | 1·44 | 7·13 | 3·32 | 1·65 | 3·44 | 1·61 | 7·13 |

**Dose- Expansion Module**

| TcpA IgM (RAU) | | | | | | | | | |
|---|---|---|---|---|---|---|---|---|---|
| **Dose (CFU)** | **D1** | **D4** | **D7** | **D15** | **D29** | **D57** | **D85** | **D180** | **max** |
| 0 | 2·9304 | 3·0452 | 3·2079 | 2·9148 | 3·4167 | 2·9956 | 3·7183 | 4·3321 | 4·3321 |
| 0 | 3·4061 | 3·2790 | 3·2711 | 2·9122 | 3·1895 | 3·4087 | 2·0424 | 1·7372 | 3·4087 |
| 0 | 7·7390 | 8·9383 | 9·0910 | 7·9886 | 8·7799 | 5·9140 | 4·5195 | 9·4721 | 9·4721 |
| 0 | 6·4380 | 7·2572 | 6·8331 | 5·5849 | 5·2566 | 95·5528 | 21·8979 | 639·7380 | 639·7380 |
| 0 | 7·1303 | 6·0041 | 8·8337 | 7·4576 | 8·1902 | 14·8303 | 13·1530 | 9·3920 | 14·8303 |
| 0 | 3·8286 | 4·1487 | 4·4836 | 3·7016 | 2·0014 | 3·5497 | 3·1749 | 3·9823 | 4·4836 |
| 0 | 13·6262 | 11·4129 | 16·0493 | 13·0952 | 21·2758 | 20·1420 | 10·0539 | 22·1101 | 22·1101 |
| $2\times10^7$ | 8·7925 | 7·5399 | 6·2555 | 7·7205 | 8·5561 | 6·3437 | 6·5030 | 8·2525 | 8·7925 |
| $2\times10^7$ | 2·4685 | 2·3664 | 3·0269 | 24·0990 | 11·7831 | 4·3129 | 4·9516 | 3·2105 | 24·0990 |
| $2\times10^7$ | 5·6683 | 6·7704 | 5·5964 | 7·0249 | 5·6625 | 4·9039 | 4·0615 | 4·3623 | 7·0249 |
| $2\times10^7$ | 3·4379 | 5·7779 | 4·8899 | 12·1735 | 5·4961 | 6·0682 | 4·7250 | 5·3576 | 12·1735 |
| $2\times10^7$ | 5·4446 | 4·5112 | 7·5461 | 24·6897 | 10·9158 | 4·7250 | 4·2854 | 4·5963 | 24·6897 |
| $2\times10^7$ | 1·7920 | 1·7970 | 1·6750 | 2·2800 | 2·9330 | 2·6121 | 3·0583 | 3·2979 | 3·2979 |
| $2\times10^7$ | 7·7513 | | 7·5308 | 6·5267 | 5·9053 | 6·9139 | 9·2733 | | 9·2733 |
| $2\times10^7$ | 2·2952 | 2·4660 | 2·2749 | 3·0269 | 2·8109 | 3·9082 | 3·1951 | 3·0110 | 3·9082 |
| $2\times10^7$ | 4·4836 | 4·9713 | 4·4036 | 4·2005 | 3·3160 | 3·5053 | 2·1967 | 4·1001 | 4·9713 |
| $2\times10^7$ | 13·2562 | 17·0566 | 22·7234 | 20·7857 | 13·5660 | 13·0090 | 12·4778 | 13·9939 | 22·7234 |
| $2\times10^7$ | 1·5834 | 2·2752 | 4·4003 | 13·5360 | 12·8385 | 5·9666 | 3·5796 | 2·6543 | 13·5360 |
| $2\times10^7$ | 4·8608 | 4·7219 | 6·7575 | 6·7857 | 6·4557 | 6·6877 | 6·4156 | 5·3687 | 6·7857 |
| $2\times10^7$ | 3·3894 | 3·6862 | 6·8709 | 9·3325 | 5·9666 | 3·9657 | 3·7405 | 3·4252 | 9·3325 |
| $2\times10^7$ | 4·4278 | 3·0432 | 5·2262 | 4·3730 | 4·6058 | 5·3798 | | 4·2831 | 5·3798 |
| $2\times10^8$ | 2·7617 | 2·8368 | 3·3160 | 8·9605 | 4·0234 | 2·5530 | 66·7972 | 3·9367 | 66·7972 |
| $2\times10^8$ | 11·5540 | 11·2724 | 14·1426 | 16·8902 | 13·4503 | 14·8323 | 11·1497 | 5·1660 | 16·8902 |
| $2\times10^8$ | 5·5276 | 5·4818 | 6·3820 | 7·7113 | 4·8171 | 8·1840 | 5·8201 | 5·1831 | 8·1840 |
| $2\times10^8$ | 15·8581 | 17·2279 | 15·8841 | 8·6662 | 11·4503 | 11·6470 | 9·2930 | 6·6738 | 17·2279 |
| $2\times10^8$ | 4·0343 | 4·4670 | 5·0839 | 5·1716 | 5·8300 | 4·6346 | 5·4133 | 4·7612 | 5·8300 |
| $2\times10^8$ | 10·3750 | 10·8758 | 11·8104 | 19·8849 | | | | | 19·8849 |
| $2\times10^8$ | 9·2444 | 11·2108 | 11·4131 | 13·4008 | 7·4836 | 5·6072 | 7·8195 | 6·4960 | 13·4008 |
| $2\times10^8$ | 1·7272 | 1·4152 | 1·4350 | 1·9144 | 2·2752 | 2·3925 | 2·5204 | 2·4717 | 2·5204 |
| $2\times10^8$ | 5·4132 | 4·9910 | 5·1376 | 8·0185 | 9·7375 | 7·6576 | 7·5622 | 8·3623 | 9·7375 |
| $2\times10^8$ | 2·5754 | 2·7236 | 2·3769 | 2·2015 | 1·8087 | 1·8754 | 1·8501 | 2·0329 | 2·7236 |
| $2\times10^8$ | 7·0153 | 9·3721 | 10·0539 | 10·0539 | 9·4120 | 7·7868 | 8·7036 | 9·4721 | 10·0539 |
| $2\times10^8$ | 1·8669 | 1·8838 | 1·4351 | 1·6775 | 1·5042 | | 1·4283 | 1·6168 | 1·8838 |
| $2\times10^8$ | 2·5368 | 1·9180 | 3·1415 | 3·0691 | 3·4109 | 3·0238 | 2·7003 | 2·7826 | 3·4109 |

| Fold change TcpA IgM | | | | | | | | | |
|---|---|---|---|---|---|---|---|---|---|
| **Dose (CFU)** | **D1** | **D4** | **D7** | **D15** | **D29** | **D57** | **D85** | **D180** | **max FC** |
| 0 | 1·00 | 1·04 | 1·09 | 0·99 | 1·17 | 1·02 | 1·27 | 1·48 | 1·48 |
| 0 | 1·00 | 0·96 | 0·96 | 0·85 | 0·94 | 1·00 | 0·60 | 0·51 | 1·00 |
| 0 | 1·00 | 1·15 | 1·17 | 1·03 | 1·13 | 0·76 | 0·58 | 1·22 | 1·22 |
| 0 | 1·00 | 1·13 | 1·06 | 0·87 | 0·82 | 14·84 | 3·40 | 99·37 | 99·37 |
| 0 | 1·00 | 0·84 | 1·24 | 1·05 | 1·15 | 2·08 | 1·84 | 1·32 | 2·08 |
| 0 | 1·00 | 1·08 | 1·17 | 0·97 | 0·52 | 0·93 | 0·83 | 1·04 | 1·17 |
| 0 | 1·00 | 0·84 | 1·18 | 0·96 | 1·56 | 1·48 | 0·74 | 1·62 | 1·62 |
| $2\times10^7$ | 1·00 | 0·86 | 0·71 | 0·88 | 0·97 | 0·72 | 0·74 | 0·94 | 0·97 |
| $2\times10^7$ | 1·00 | 0·96 | 1·23 | 9·76 | 4·77 | 1·75 | 2·01 | 1·30 | 9·76 |
| $2\times10^7$ | 1·00 | 1·19 | 0·99 | 1·24 | 1·00 | 0·87 | 0·72 | 0·77 | 1·24 |
| $2\times10^7$ | 1·00 | 1·68 | 1·42 | 3·54 | 1·60 | 1·77 | 1·37 | 1·56 | 3·54 |
| $2\times10^7$ | 1·00 | 0·83 | 1·39 | 4·53 | 2·00 | 0·87 | 0·79 | 0·84 | 4·53 |
| $2\times10^7$ | 1·00 | 1·00 | 0·93 | 1·27 | 1·64 | 1·46 | 1·71 | 1·84 | 1·84 |
| $2\times10^7$ | 1·00 | | 0·97 | 0·84 | 0·76 | 0·89 | 1·20 | | 1·20 |
| $2\times10^7$ | 1·00 | 1·07 | 0·99 | 1·32 | 1·22 | 1·70 | 1·39 | 1·31 | 1·70 |
| $2\times10^7$ | 1·00 | 1·11 | 0·98 | 0·94 | 0·74 | 0·78 | 0·49 | 0·91 | 1·11 |
| $2\times10^7$ | 1·00 | 1·29 | 1·71 | 1·57 | 1·02 | 0·98 | 0·94 | 1·06 | 1·71 |
| $2\times10^7$ | 1·00 | 1·44 | 2·78 | 8·55 | 8·11 | 3·77 | 2·26 | 1·68 | 8·55 |
| $2\times10^7$ | 1·00 | 0·97 | 1·39 | 1·40 | 1·33 | 1·38 | 1·32 | 1·10 | 1·40 |
| $2\times10^7$ | 1·00 | 1·09 | 2·03 | 2·75 | 1·76 | 1·17 | 1·10 | 1·01 | 2·75 |
| $2\times10^7$ | 1·00 | 0·69 | 1·18 | 0·99 | 1·04 | 1·21 | | 0·97 | 1·21 |
| $2\times10^8$ | 1·00 | 1·03 | 1·20 | 3·24 | 1·46 | 0·92 | 24·19 | 1·43 | 24·19 |
| $2\times10^8$ | 1·00 | 0·98 | 1·22 | 1·46 | 1·16 | 1·28 | 0·97 | 0·45 | 1·46 |
| $2\times10^8$ | 1·00 | 0·99 | 1·15 | 1·40 | 0·87 | 1·48 | 1·05 | 0·94 | 1·48 |
| $2\times10^8$ | 1·00 | 1·09 | 1·00 | 0·55 | 0·72 | 0·73 | 0·59 | 0·42 | 1·09 |
| $2\times10^8$ | 1·00 | 1·11 | 1·26 | 1·28 | 1·45 | 1·15 | 1·34 | 1·18 | 1·45 |
| $2\times10^8$ | 1·00 | 1·05 | 1·14 | 1·92 | | | | | 1·92 |
| $2\times10^8$ | 1·00 | 1·21 | 1·23 | 1·45 | 0·81 | 0·61 | 0·85 | 0·70 | 1·45 |
| $2\times10^8$ | 1·00 | 0·82 | 0·83 | 1·11 | 1·32 | 1·39 | 1·46 | 1·43 | 1·46 |
| $2\times10^8$ | 1·00 | 0·92 | 0·95 | 1·48 | 1·80 | 1·41 | 1·40 | 1·54 | 1·80 |
| $2\times10^8$ | 1·00 | 1·06 | 0·92 | 0·85 | 0·70 | 0·73 | 0·72 | 0·79 | 1·06 |
| $2\times10^8$ | 1·00 | 1·34 | 1·43 | 1·43 | 1·34 | 1·11 | 1·24 | 1·35 | 1·43 |
| $2\times10^8$ | 1·00 | 1·01 | 0·77 | 0·90 | 0·81 | | 0·77 | 0·87 | 1·01 |
| $2\times10^8$ | 1·00 | 0·76 | 1·24 | 1·21 | 1·34 | 1·19 | 1·06 | 1·10 | 1·34 |

| | RAU | | |
|---|---|---|---|
| | D1 | D39 | |
| *V. cholerae* | 8·3096 | 11·9207 | |
| | 22·5324 | 24·1178 | |
| | 5·0720 | 6·7092 | |
| | 2·0096 | 2·3126 | |
| | 412·9516 | 435·5952 | |
| | D1 | D11 | |
| Vaxchora D11 Challenge | 3·0896 | 2·7502 | |
| | 12·4626 | 12·7372 | |
| | 4·5114 | 4·1675 | |
| | 4·9911 | 4·4099 | |
| | 3·1501 | 3·7446 | |
| | 5·8859 | 4·7500 | |
| | D1 | D11 | D91 |
| Vaxchora D91 Challenge | 2·6483 | 2·3649 | 2·5012 |
| | 11·9207 | 5·3322 | 4·9142 |
| | 1·2446 | 1·2915 | 1·5760 |
| | 6·0910 | 4·7309 | 8·1284 |
| | 8·1504 | 10·2717 | 7·9180 |
| | 2·0897 | 2·4033 | 2·6764 |

| | RAU fold change | | | max FC |
|---|---|---|---|---|
| | D1 | D39 | | |
| *V. cholerae* | 1·00 | 1·43 | | 1·43 |
| | 1·00 | 1·07 | | 1·07 |
| | 1·00 | 1·32 | | 1·32 |
| | 1·00 | 1·15 | | 1·15 |
| | 1·00 | 1·05 | | 1·05 |
| | D1 | D11 | | |
| Vaxchora D11 Challenge | 1·00 | 0·89 | | 0·89 |
| | 1·00 | 1·02 | | 1·02 |
| | 1·00 | 0·92 | | 0·92 |
| | 1·00 | 0·88 | | 0·88 |
| | 1·00 | 1·19 | | 1·19 |
| | 1·00 | 0·81 | | 0·81 |
| | D1 | D11 | D91 | |
| Vaxchora D91 Challenge | 1·00 | 0·89 | 0·94 | 0·94 |
| | 1·00 | 0·45 | 0·41 | 0·45 |
| | 1·00 | 1·04 | 1·27 | 1·27 |
| | 1·00 | 0·78 | 1·33 | 1·33 |
| | 1·00 | 1·26 | 0·97 | 1·26 |
| | 1·00 | 1·15 | 1·28 | 1·28 |

**Dose-Escalation Module**

| | TcpA IgG (RAU) | | | | | | | | |
|---|---|---|---|---|---|---|---|---|---|
| Dose (CFU) | D1 | D4 | D7 | D15 | D29 | D57 | D85 | D180 | max |
| $10^5$ | 0·0447 | 0·0294 | 0·0593 | 0·0447 | 0·0593 | 0·4180 | 0·1922 | 0·0788 | 0·4180 |
| $10^5$ | 0·0358 | 0·0349 | 0·0737 | 0·0675 | 0·0506 | 0·1151 | 0·1684 | | 0·1684 |
| $10^5$ | 0·1062 | 0·1195 | 0·0995 | 0·1595 | 0·2076 | 0·5916 | 0·9098 | | 0·9098 |
| $10^6$ | 0·0707 | 0·0619 | 0·0583 | 0·1784 | 0·1291 | 0·1080 | 0·1194 | | 0·1784 |
| $10^6$ | 0·0565 | 0·0583 | 0·0565 | 0·0931 | 0·0777 | 0·0964 | 0·0511 | 0·0981 | 0·0981 |
| $10^6$ | 0·2322 | 0·1918 | 0·1932 | 0·1754 | 0·2222 | 0·2477 | 0·2208 | 0·1121 | 0·2477 |
| $10^7$ | 0·1291 | 0·1338 | 0·1146 | 0·1526 | 0·3602 | 0·7304 | 0·6159 | 0·1666 | 0·7304 |
| $10^7$ | 0·0707 | 0·0829 | 0·0742 | 0·0547 | 0·0829 | 0·0637 | 0·0742 | 0·0448 | 0·0829 |
| $10^7$ | 0·3615 | 0·3010 | 0·3472 | 0·2753 | 0·2889 | 0·2463 | 0·2616 | 0·0755 | 0·3615 |
| $10^8$ | 0·2136 | 0·2364 | 0·2546 | 0·2265 | 0·2698 | 0·1417 | 0·1754 | 0·0652 | 0·2698 |
| $10^8$ | 0·1146 | 0·1064 | 0·0760 | 0·0742 | 0·0742 | 0·0846 | 0·0652 | 0·0716 | 0·1146 |
| $10^8$ | 0·3024 | 0·2643 | 0·2983 | 0·3498 | 0·5667 | 6·5286 | 10·2342 | 9·4380 | 10·2342 |
| $10^9$ | 0·1194 | 0·1307 | 0·1918 | 0·1769 | 0·1495 | 0·0677 | 0·0911 | 0·0768 | 0·1918 |
| $10^9$ | 0·1510 | 0·1572 | 0·1784 | 0·1068 | 0·1200 | 0·1068 | 0·2579 | | 0·2579 |
| $10^9$ | 0·0537 | 0·0524 | 0·0537 | 0·0575 | 0·0613 | 0·0613 | 0·0587 | 0·0486 | 0·0613 |
| $10^{10}$ | 0·0136 | 0·0312 | 0·0448 | 0·1162 | 0·0716 | | 0·0639 | 0·0511 | 0·1162 |
| $10^{10}$ | 0·0264 | 0·0312 | 0·0312 | 0·0386 | | | 0·0373 | 0·0361 | 0·0386 |
| $10^{10}$ | 0·1506 | 0·1613 | 0·1519 | 0·1280 | 0·2421 | 0·3881 | 0·3073 | 0·2502 | 0·3881 |

| | Fold change TcpA IgG | | | | | | | | |
|---|---|---|---|---|---|---|---|---|---|
| Dose (CFU) | D1 | D4 | D7 | D15 | D29 | D57 | D85 | D180 | max FC |
| $10^5$ | 1·00 | 0·66 | 1·33 | 1·00 | 1·33 | 9·35 | 4·30 | 1·76 | 9·35 |
| $10^5$ | 1·00 | 0·97 | 2·06 | 1·88 | 1·41 | 3·21 | 4·70 | | 4·70 |
| $10^5$ | 1·00 | 1·12 | 0·94 | 1·50 | 1·95 | 5·57 | 8·56 | | 8·56 |
| $10^6$ | 1·00 | 0·88 | 0·82 | 2·52 | 1·82 | 1·53 | 1·69 | | 2·52 |
| $10^6$ | 1·00 | 1·03 | 1·00 | 1·65 | 1·37 | 1·71 | 0·90 | 1·74 | 1·74 |
| $10^6$ | 1·00 | 0·83 | 0·83 | 0·76 | 0·96 | 1·07 | 0·95 | 0·48 | 1·07 |
| $10^7$ | 1·00 | 1·04 | 0·89 | 1·18 | 2·79 | 5·66 | 4·77 | 1·29 | 5·66 |
| $10^7$ | 1·00 | 1·17 | 1·05 | 0·77 | 1·17 | 0·90 | 1·05 | 0·63 | 1·17 |
| $10^7$ | 1·00 | 0·83 | 0·96 | 0·76 | 0·80 | 0·68 | 0·72 | 0·21 | 0·96 |
| $10^8$ | 1·00 | 1·11 | 1·19 | 1·06 | 1·26 | 0·66 | 0·82 | 0·30 | 1·26 |
| $10^8$ | 1·00 | 0·93 | 0·66 | 0·65 | 0·65 | 0·74 | 0·57 | 0·62 | 0·93 |
| $10^8$ | 1·00 | 0·87 | 0·99 | 1·16 | 1·87 | 21·59 | 33·85 | 31·21 | 33·85 |
| $10^9$ | 1·00 | 1·09 | 1·61 | 1·48 | 1·25 | 0·57 | 0·76 | 0·64 | 1·61 |
| $10^9$ | 1·00 | 1·04 | 1·18 | 0·71 | 0·79 | 0·71 | 1·71 | | 1·71 |
| $10^9$ | 1·00 | 0·98 | 1·00 | 1·07 | 1·14 | 1·14 | 1·09 | 0·91 | 1·14 |
| $10^{10}$ | 1·00 | 2·30 | 3·30 | 8·56 | 5·27 | | 4·70 | 3·76 | 8·56 |
| $10^{10}$ | 1·00 | 1·18 | 1·18 | 1·46 | | | 1·42 | 1·37 | 1·46 |
| $10^{10}$ | 1·00 | 1·07 | 1·01 | 0·85 | 1·61 | 2·58 | 2·04 | 1·66 | 2·58 |

**Dose- Expansion Module**

| | TcpA IgG (RAU) | | | | | | | | |
|---|---|---|---|---|---|---|---|---|---|
| Dose (CFU) | D1 | D4 | D7 | D15 | D29 | D57 | D85 | D180 | max |
| 0 | 0·0867 | 0·1050 | 0·0935 | 0·0946 | 0·1016 | 0·0985 | 0·1072 | 0·1142 | 0·1142 |
| 0 | 0·1565 | 0·1776 | 0·1621 | 0·1373 | 0·1595 | 0·1634 | 0·1618 | 0·1300 | 0·1776 |
| 0 | 0·2299 | 0·2438 | 0·2660 | 0·2412 | 0·2512 | 0·2561 | 0·2471 | 0·0525 | 0·2660 |
| 0 | 0·2942 | 0·2671 | 0·2978 | 0·2674 | 0·2908 | 0·1107 | 0·0746 | 0·0442 | 0·2978 |
| 0 | 0·1106 | 0·1204 | 0·1106 | 0·1307 | 0·0993 | 0·0280 | 0·0403 | 0·0182 | 0·1307 |
| 0 | 0·0553 | 0·0747 | 0·0642 | 0·0033 | | 0·0024 | | 0·0002 | 0·0747 |
| 0 | 0·1049 | 0·0872 | 0·0746 | 0·1146 | 0·1273 | 0·0545 | 0·0670 | 0·0890 | 0·1273 |
| $2\times10^7$ | 0·7588 | 0·7588 | 0·6883 | 0·8107 | 2·0121 | 0·9726 | 0·8715 | 0·8879 | 2·0121 |
| $2\times10^7$ | 0·1578 | 0·1637 | 0·1522 | 0·1893 | 0·1709 | 0·1699 | 0·1858 | 0·1676 | 0·1893 |
| $2\times10^7$ | 0·1001 | 0·1001 | 0·0989 | 0·1261 | 0·1087 | 0·1575 | 0·1240 | 0·1057 | 0·1575 |
| $2\times10^7$ | 0·0974 | 0·1211 | 0·1168 | 0·1390 | 0·1095 | 0·2008 | 0·1959 | 0·1529 | 0·2008 |
| $2\times10^7$ | 0·1608 | 0·1641 | 0·1427 | 0·2482 | 0·2657 | 0·5390 | 0·5512 | 0·4582 | 0·5512 |
| $2\times10^7$ | 0·1562 | 0·1421 | 0·1397 | 0·1355 | 0·1488 | 0·1912 | 0·2257 | 0·1018 | 0·2257 |
| $2\times10^7$ | 0·1833 | | 0·1909 | 0·2002 | 0·2100 | 0·1808 | 0·1320 | | 0·2100 |
| $2\times10^7$ | 0·0993 | 0·1027 | 0·0997 | 0·1345 | 0·1400 | 0·0718 | 0·0378 | 0·0427 | 0·1400 |
| $2\times10^7$ | 0·1502 | 0·1268 | 0·1296 | 0·1407 | 0·1282 | 0·0235 | 0·0442 | 0·0329 | 0·1502 |
| $2\times10^7$ | 0·1581 | 0·1508 | 0·1750 | 0·1618 | 0·3640 | 0·5295 | 0·5178 | 0·4763 | 0·5295 |
| $2\times10^7$ | 0·0545 | 0·0260 | 0·0516 | 0·0486 | 0·0904 | 0·2353 | 0·3134 | 0·2839 | 0·3134 |
| $2\times10^7$ | 0·0477 | 0·0467 | 0·0349 | 0·0555 | 0·0755 | 0·0774 | 0·0858 | 0·0486 | 0·0858 |
| $2\times10^7$ | 0·0270 | 0·0388 | 0·0442 | 0·0344 | 0·0457 | 0·0353 | 0·0491 | 0·0447 | 0·0491 |
| $2\times10^7$ | 0·0383 | 0·0437 | 0·0574 | 0·0275 | 0·0289 | 0·0447 | | 0·0231 | 0·0574 |
| $2\times10^8$ | 0·1247 | 0·1444 | 0·1208 | 0·2435 | 0·1471 | 0·2106 | 0·2224 | 0·1731 | 0·2435 |
| $2\times10^8$ | 0·0591 | 0·0526 | 0·0610 | 0·0469 | 0·0291 | 0·0530 | 0·0286 | 0·0391 | 0·0610 |
| $2\times10^8$ | 0·0997 | 0·1275 | 0·1050 | 0·1390 | 0·1502 | 0·2287 | 0·2182 | 0·1780 | 0·2287 |
| $2\times10^8$ | 0·1562 | 0·1588 | 0·1558 | 0·1410 | 0·2042 | 0·7168 | 0·7542 | 0·2353 | 0·7542 |
| $2\times10^8$ | 0·0619 | 0·0806 | 0·0814 | 0·0997 | 0·1153 | 0·0164 | 0·0083 | 0·0043 | 0·1153 |
| $2\times10^8$ | 0·1397 | 0·1349 | 0·1300 | 0·1451 | | | | | 0·1451 |
| $2\times10^8$ | 0·1495 | 0·1545 | 0·1673 | 0·2134 | 0·1476 | 0·1836 | 0·2456 | 0·1463 | 0·2456 |
| $2\times10^8$ | 0·0426 | 0·0297 | 0·0468 | 0·0568 | 0·0066 | 0·0043 | 0·0078 | 0·0066 | 0·0568 |
| $2\times10^8$ | 0·1012 | 0·1072 | 0·0935 | 0·0462 | 0·0353 | 0·0398 | 0·0521 | 0·0319 | 0·1072 |
| $2\times10^8$ | 0·0718 | 0·1004 | 0·0812 | 0·0904 | 0·1337 | 0·2250 | 0·2015 | 0·1632 | 0·2250 |
| $2\times10^8$ | 0·0314 | 0·0270 | 0·0211 | 0·0280 | 0·0226 | 0·0240 | 0·0211 | 0·0245 | 0·0314 |
| $2\times10^8$ | 0·0646 | 0·0477 | 0·0289 | 0·0211 | 0·0187 | | 0·0339 | 0·0250 | 0·0646 |
| $2\times10^8$ | 0·0231 | 0·0187 | 0·0589 | 0·1816 | 0·1430 | 0·1844 | 0·1595 | 0·1753 | 0·1844 |

| | Fold change TcpA IgG | | | | | | | | |
|---|---|---|---|---|---|---|---|---|---|
| Dose (CFU) | D1 | D4 | D7 | D15 | D29 | D57 | D85 | D180 | max FC |
| 0 | 1·00 | 1·21 | 1·08 | 1·09 | 1·17 | 1·14 | 1·24 | 1·32 | 1·32 |
| 0 | 1·00 | 1·13 | 1·04 | 0·88 | 1·02 | 1·04 | 1·03 | 0·83 | 1·13 |
| 0 | 1·00 | 1·06 | 1·16 | 1·05 | 1·09 | 1·11 | 1·07 | 0·23 | 1·16 |
| 0 | 1·00 | 0·91 | 1·01 | 0·91 | 0·99 | 0·38 | 0·25 | 0·15 | 1·01 |
| 0 | 1·00 | 1·09 | 1·00 | 1·18 | 0·90 | 0·25 | 0·36 | 0·16 | 1·18 |
| 0 | 1·00 | 1·35 | 1·16 | 0·06 | | 0·04 | | 0·00 | 1·35 |
| 0 | 1·00 | 0·83 | 0·71 | 1·09 | 1·21 | 0·52 | 0·64 | 0·85 | 1·21 |
| $2\times10^7$ | 1·00 | 1·00 | 0·91 | 1·07 | 2·65 | 1·28 | 1·15 | 1·17 | 2·65 |
| $2\times10^7$ | 1·00 | 1·04 | 0·96 | 1·20 | 1·08 | 1·08 | 1·18 | 1·06 | 1·20 |
| $2\times10^7$ | 1·00 | 1·00 | 0·99 | 1·26 | 1·09 | 1·57 | 1·24 | 1·06 | 1·57 |
| $2\times10^7$ | 1·00 | 1·24 | 1·20 | 1·43 | 1·12 | 2·06 | 2·01 | 1·57 | 2·06 |
| $2\times10^7$ | 1·00 | 1·02 | 0·89 | 1·54 | 1·65 | 3·35 | 3·43 | 2·85 | 3·43 |
| $2\times10^7$ | 1·00 | 0·91 | 0·89 | 0·87 | 0·95 | 1·22 | 1·45 | 0·65 | 1·45 |
| $2\times10^7$ | 1·00 | | 1·04 | 1·09 | 1·15 | 0·99 | 0·72 | | 1·15 |
| $2\times10^7$ | 1·00 | 1·03 | 1·00 | 1·35 | 1·41 | 0·72 | 0·38 | 0·43 | 1·41 |
| $2\times10^7$ | 1·00 | 0·84 | 0·86 | 0·94 | 0·85 | 0·16 | 0·29 | 0·22 | 0·94 |
| $2\times10^7$ | 1·00 | 0·95 | 1·11 | 1·02 | 2·30 | 3·35 | 3·27 | 3·01 | 3·35 |
| $2\times10^7$ | 1·00 | 0·48 | 0·95 | 0·89 | 1·66 | 4·32 | 5·75 | 5·21 | 5·75 |
| $2\times10^7$ | 1·00 | 0·98 | 0·73 | 1·16 | 1·59 | 1·62 | 1·80 | 1·02 | 1·80 |
| $2\times10^7$ | 1·00 | 1·44 | 1·64 | 1·27 | 1·69 | 1·31 | 1·82 | 1·66 | 1·82 |
| $2\times10^7$ | 1·00 | 1·14 | 1·50 | 0·72 | 0·76 | 1·17 | | 0·60 | 1·50 |
| $2\times10^8$ | 1·00 | 1·16 | 0·97 | 1·95 | 1·18 | 1·69 | 1·78 | 1·39 | 1·95 |
| $2\times10^8$ | 1·00 | 0·89 | 1·03 | 0·79 | 0·49 | 0·90 | 0·48 | 0·66 | 1·03 |
| $2\times10^8$ | 1·00 | 1·28 | 1·05 | 1·39 | 1·51 | 2·29 | 2·19 | 1·79 | 2·29 |
| $2\times10^8$ | 1·00 | 1·02 | 1·00 | 0·90 | 1·31 | 4·59 | 4·83 | 1·51 | 4·83 |
| $2\times10^8$ | 1·00 | 1·30 | 1·32 | 1·61 | 1·86 | 0·26 | 0·13 | 0·07 | 1·86 |
| $2\times10^8$ | 1·00 | 0·97 | 0·93 | 1·04 | | | | | 1·04 |
| $2\times10^8$ | 1·00 | 1·03 | 1·12 | 1·43 | 0·99 | 1·23 | 1·64 | 0·98 | 1·64 |
| $2\times10^8$ | 1·00 | 0·70 | 1·10 | 1·33 | 0·16 | 0·10 | 0·18 | 0·16 | 1·33 |
| $2\times10^8$ | 1·00 | 1·06 | 0·92 | 0·46 | 0·35 | 0·39 | 0·51 | 0·32 | 1·06 |
| $2\times10^8$ | 1·00 | 1·40 | 1·13 | 1·26 | 1·86 | 3·13 | 2·81 | 2·27 | 3·13 |
| $2\times10^8$ | 1·00 | 0·86 | 0·67 | 0·89 | 0·72 | 0·77 | 0·67 | 0·78 | 0·89 |
| $2\times10^8$ | 1·00 | 0·74 | 0·45 | 0·33 | 0·29 | | 0·52 | 0·39 | 0·74 |
| $2\times10^8$ | 1·00 | 0·81 | 2·55 | 7·87 | 6·20 | 7·99 | 6·92 | 7·60 | 7·99 |

| | RAU | | |
|---|---|---|---|
| | D1 | D39 | |
| *V. cholerae* | 0·3223 | 1·0861 | |
| | 0·2726 | 0·4767 | |
| | 0·2983 | 0·6321 | |
| | 0·2712 | 1·0099 | |
| | 1·5925 | 1·4285 | |
| | D1 | D11 | |
| Vaxchora D11 Challenge | 0·6414 | 0·8354 | |
| | 1·0390 | 1·1117 | |
| | 0·9341 | 0·7932 | |
| | 0·6356 | 0·6827 | |
| | 0·6124 | 0·6655 | |
| | 0·5537 | 0·3947 | |
| | D1 | D11 | D91 |
| Vaxchora D91 Challenge | 0·5832 | 0·8243 | 0·8519 |
| | 0·1541 | 0·2546 | 0·4621 |
| | 0·3769 | 0·5513 | 0·4645 |
| | 0·8915 | 0·9287 | 0·8597 |
| | 0·1962 | 0·4694 | 0·5820 |
| | 0·1275 | 0·1814 | 0·1769 |

| | RAU fold change | | | max FC |
|---|---|---|---|---|
| | D1 | D39 | | |
| *V. cholerae* | 1·00 | 3·37 | | 3·37 |
| | 1·00 | 1·75 | | 1·75 |
| | 1·00 | 2·12 | | 2·12 |
| | 1·00 | 3·72 | | 3·72 |
| | 1·00 | 0·90 | | 0·90 |
| | D1 | D11 | | |
| Vaxchora D11 Challenge | 1·00 | 1·30 | | 1·30 |
| | 1·00 | 1·07 | | 1·07 |
| | 1·00 | 0·85 | | 0·85 |
| | 1·00 | 1·07 | | 1·07 |
| | 1·00 | 1·09 | | 1·09 |
| | 1·00 | 0·71 | | 0·71 |
| | D1 | D11 | D91 | |
| Vaxchora D91 Challenge | 1·00 | 1·41 | 1·46 | 1·46 |
| | 1·00 | 1·65 | 3·00 | 3·00 |
| | 1·00 | 1·46 | 1·23 | 1·46 |
| | 1·00 | 1·04 | 0·96 | 1·04 |
| | 1·00 | 2·39 | 2·97 | 2·97 |
| | 1·00 | 1·42 | 1·39 | 1·42 |

**Dose-Escalation Module**

| TcpA IgA (RAU) | | | | | | | | | |
|---|---|---|---|---|---|---|---|---|---|
| **Dose (CFU)** | **D1** | **D4** | **D7** | **D15** | **D29** | **D57** | **D85** | **D180** | **max** |
| $10^5$ | 0·2262 | 0·2515 | 0·2399 | 0·1942 | 0·2499 | 0·3304 | 0·1943 | 0·1786 | 0·3304 |
| $10^5$ | 0·1054 | 0·0950 | 0·1079 | 0·1215 | 0·1005 | 0·0975 | 0·1166 | | 0·1215 |
| $10^5$ | 0·0891 | 0·0921 | 0·0863 | 0·0905 | 0·1686 | 0·3347 | 0·2929 | | 0·3347 |
| $10^6$ | 0·0706 | 0·0836 | 0·0734 | 0·0942 | 0·0981 | 0·0967 | 0·1038 | | 0·1038 |
| $10^6$ | 0·2641 | 0·2250 | 0·1791 | 0·1732 | 0·1640 | 0·1876 | 0·1739 | 0·1833 | 0·2641 |
| $10^6$ | 0·1014 | 0·1601 | 0·1142 | 0·1314 | 0·0904 | 0·1098 | 0·0784 | 0·0784 | 0·1601 |
| $10^7$ | 0·1149 | 0·1156 | 0·1180 | 0·1552 | 0·2013 | 0·2098 | 0·2412 | 0·1235 | 0·2412 |
| $10^7$ | 0·0972 | 0·0753 | 0·0768 | 0·0652 | 0·0742 | 0·0528 | 0·0730 | 0·0284 | 0·0972 |
| $10^7$ | 0·1488 | 0·1175 | 0·0986 | 0·1168 | 0·1280 | 0·0892 | 0·1273 | 0·0177 | 0·1488 |
| $10^8$ | 0·1601 | 0·1628 | 0·1381 | 0·1474 | 0·1442 | 0·1430 | 0·1704 | 0·1051 | 0·1704 |
| $10^8$ | 0·1614 | 0·1229 | 0·1416 | 0·2266 | 0·2323 | 0·1906 | 0·1161 | 0·1334 | 0·2323 |
| $10^8$ | 0·0430 | 0·0399 | 0·0614 | 0·0942 | 0·1755 | 0·7457 | 0·9681 | 1·6468 | 1·6468 |
| $10^9$ | 0·1672 | 0·1695 | 0·2713 | 0·2061 | 0·2287 | 0·1293 | 0·1103 | 0·1089 | 0·2713 |
| $10^9$ | 0·3542 | 0·2935 | 0·2804 | 0·1840 | 0·1860 | 0·1515 | 0·2818 | | 0·3542 |
| $10^9$ | 0·1193 | 0·1538 | 0·1357 | 0·1522 | 0·1413 | 0·1198 | 0·1461 | 0·1293 | 0·1538 |
| $10^{10}$ | 0·0396 | 0·0365 | 0·0531 | 0·0645 | 0·1074 | | 0·0723 | 0·1155 | 0·1155 |
| $10^{10}$ | 0·1187 | 0·1227 | 0·1124 | 0·1123 | | | 0·1357 | 0·1256 | 0·1357 |
| $10^{10}$ | 0·0722 | 0·0807 | 0·0709 | 0·1268 | 0·1114 | 0·0939 | 0·1042 | 0·0924 | 0·1268 |

| Fold change TcpA IgA | | | | | | | | | |
|---|---|---|---|---|---|---|---|---|---|
| **Dose (CFU)** | **D1** | **D4** | **D7** | **D15** | **D29** | **D57** | **D85** | **D180** | **max FC** |
| $10^5$ | 1·00 | 1·11 | 1·06 | 0·86 | 1·10 | 1·46 | 0·86 | 0·79 | 1·46 |
| $10^5$ | 1·00 | 0·90 | 1·02 | 1·15 | 0·95 | 0·92 | 1·11 | | 1·15 |
| $10^5$ | 1·00 | 1·03 | 0·97 | 1·02 | 1·89 | 3·76 | 3·29 | | 3·76 |
| $10^6$ | 1·00 | 1·18 | 1·04 | 1·33 | 1·39 | 1·37 | 1·47 | | 1·47 |
| $10^6$ | 1·00 | 0·85 | 0·68 | 0·66 | 0·62 | 0·71 | 0·66 | 0·69 | 0·85 |
| $10^6$ | 1·00 | 1·58 | 1·13 | 1·30 | 0·89 | 1·08 | 0·77 | 0·77 | 1·58 |
| $10^7$ | 1·00 | 1·01 | 1·03 | 1·35 | 1·75 | 1·82 | 2·10 | 1·07 | 2·10 |
| $10^7$ | 1·00 | 0·78 | 0·79 | 0·67 | 0·76 | 0·54 | 0·75 | 0·29 | 0·79 |
| $10^7$ | 1·00 | 0·79 | 0·66 | 0·79 | 0·86 | 0·60 | 0·86 | 0·12 | 0·86 |
| $10^8$ | 1·00 | 1·02 | 0·86 | 0·92 | 0·90 | 0·89 | 1·06 | 0·66 | 1·06 |
| $10^8$ | 1·00 | 0·76 | 0·88 | 1·40 | 1·44 | 1·18 | 0·72 | 0·83 | 1·44 |
| $10^8$ | 1·00 | 0·93 | 1·43 | 2·19 | 4·08 | 17·34 | 22·51 | 38·29 | 38·29 |
| $10^9$ | 1·00 | 1·01 | 1·62 | 1·23 | 1·37 | 0·77 | 0·66 | 0·65 | 1·62 |
| $10^9$ | 1·00 | 0·83 | 0·79 | 0·52 | 0·53 | 0·43 | 0·80 | | 0·83 |
| $10^9$ | 1·00 | 1·29 | 1·14 | 1·28 | 1·18 | 1·00 | 1·22 | 1·08 | 1·29 |
| $10^{10}$ | 1·00 | 0·92 | 1·34 | 1·63 | 2·71 | | 1·83 | 2·92 | 2·92 |
| $10^{10}$ | 1·00 | 1·03 | 0·95 | 0·95 | | | 1·14 | 1·06 | 1·14 |
| $10^{10}$ | 1·00 | 1·12 | 0·98 | 1·76 | 1·54 | 1·30 | 1·44 | 1·28 | 1·76 |

**Dose- Expansion Module**

| TcpA IgA (RAU) | | | | | | | | | |
|---|---|---|---|---|---|---|---|---|---|
| **Dose (CFU)** | **D1** | **D4** | **D7** | **D15** | **D29** | **D57** | **D85** | **D180** | **max** |
| 0 | 0·1085 | 0·1276 | 0·1276 | 0·1207 | 0·1190 | 0·1125 | 0·1113 | 0·1235 | 0·1276 |
| 0 | 0·0733 | 0·0576 | 0·0694 | 0·0589 | 0·0733 | 0·0707 | 0·0723 | 0·0681 | 0·0733 |
| 0 | 0·0520 | 0·0605 | 0·0485 | 0·0385 | 0·0552 | 0·0626 | 0·0520 | 0·0466 | 0·0626 |
| 0 | 0·3047 | 0·2436 | 0·2850 | 0·2520 | 0·1957 | 0·2254 | 0·2164 | 0·2958 | 0·3047 |
| 0 | 0·0605 | 0·0705 | 0·0972 | 0·0744 | 0·0780 | 0·1031 | 0·0842 | 0·0762 | 0·1031 |
| 0 | 0·0404 | 0·0637 | 0·0493 | 0·0266 | 0·0310 | 0·0335 | 0·0365 | 0·0395 | 0·0637 |
| 0 | 0·0647 | 0·0501 | 0·0561 | 0·0547 | 0·0762 | 0·0494 | 0·0306 | 0·0469 | 0·0762 |
| $2\times10^7$ | 0·0589 | 0·0621 | 0·0570 | 0·1115 | 0·0937 | 0·0652 | 0·0733 | 0·0739 | 0·1115 |
| $2\times10^7$ | 0·0431 | 0·0562 | 0·0626 | 0·0757 | 0·0655 | 0·0728 | 0·1123 | 0·0490 | 0·1123 |
| $2\times10^7$ | 0·1133 | 0·1078 | 0·1068 | 0·1058 | 0·1230 | 0·0931 | 0·0987 | 0·0957 | 0·1230 |
| $2\times10^7$ | 0·1755 | 0·2239 | 0·1976 | 0·1738 | 0·1870 | 0·2807 | 0·2588 | 0·1922 | 0·2807 |
| $2\times10^7$ | 0·1788 | 0·1457 | 0·2023 | 0·3098 | 0·3022 | 0·2548 | 0·2053 | 0·1279 | 0·3098 |
| $2\times10^7$ | 0·1185 | 0·1299 | 0·1678 | 0·1997 | 0·2218 | 0·1894 | 0·1915 | 0·1221 | 0·2218 |
| $2\times10^7$ | 0·1535 | | 0·1330 | 0·1472 | 0·1173 | 0·1045 | 0·1120 | | 0·1535 |
| $2\times10^7$ | 0·0514 | 0·0634 | 0·0947 | 0·1428 | 0·1195 | 0·0761 | 0·0626 | 0·0605 | 0·1428 |
| $2\times10^7$ | 0·0896 | 0·0746 | 0·0898 | 0·1108 | 0·1002 | 0·1013 | 0·0997 | 0·1203 | 0·1203 |
| $2\times10^7$ | 0·0525 | 0·0671 | 0·0453 | 0·1068 | 0·0807 | 0·1054 | 0·0876 | 0·0561 | 0·1068 |
| $2\times10^7$ | 0·0364 | 0·0254 | 0·0292 | 0·0373 | 0·0596 | 0·0992 | 0·0689 | 0·0637 | 0·0992 |
| $2\times10^7$ | 0·0822 | 0·0927 | 0·1357 | 0·0974 | 0·0946 | 0·0835 | 0·1208 | 0·1136 | 0·1357 |
| $2\times10^7$ | 0·0200 | 0·0213 | 0·0288 | 0·0458 | 0·0392 | 0·0299 | 0·0332 | 0·0174 | 0·0458 |
| $2\times10^7$ | 0·0920 | 0·0749 | 0·0880 | 0·0543 | 0·0628 | 0·0725 | | 0·0515 | 0·0920 |
| $2\times10^8$ | 0·1235 | 0·1472 | 0·1150 | 0·1733 | 0·1510 | 0·1028 | 0·1367 | 0·1491 | 0·1733 |
| $2\times10^8$ | 0·1480 | 0·1939 | 0·1831 | 0·1533 | 0·1996 | 0·2058 | 0·1742 | 0·0936 | 0·2058 |
| $2\times10^8$ | 0·0471 | 0·0482 | 0·0565 | 0·0637 | 0·0741 | 0·0837 | 0·0463 | 0·0597 | 0·0837 |
| $2\times10^8$ | 0·0967 | 0·0901 | 0·1000 | 0·0952 | 0·1374 | 0·1877 | 0·1196 | 0·1157 | 0·1877 |
| $2\times10^8$ | 0·0439 | 0·0272 | 0·0490 | 0·0455 | 0·0286 | 0·0251 | 0·0197 | 0·0171 | 0·0490 |
| $2\times10^8$ | 0·1296 | 0·1108 | 0·1070 | 0·1217 | | | | | 0·1296 |
| $2\times10^8$ | 0·0546 | 0·0702 | 0·0726 | 0·1681 | 0·0880 | 0·0830 | 0·0875 | 0·0650 | 0·1681 |
| $2\times10^8$ | 0·1088 | 0·0816 | 0·0893 | 0·0875 | 0·0604 | 0·0774 | 0·0774 | 0·0832 | 0·1088 |
| $2\times10^8$ | 0·1068 | 0·0898 | 0·1007 | 0·0863 | 0·1234 | 0·1134 | 0·1120 | 0·0985 | 0·1234 |
| $2\times10^8$ | 0·2204 | 0·2212 | 0·2345 | 0·2227 | 0·2840 | 0·2271 | 0·2103 | 0·1800 | 0·2840 |
| $2\times10^8$ | 0·0517 | 0·0602 | 0·0588 | 0·0656 | 0·0544 | 0·0552 | 0·0546 | 0·0531 | 0·0656 |
| $2\times10^8$ | 0·3359 | 0·4103 | 0·3486 | 0·2951 | 0·3299 | | 0·4045 | 0·4911 | 0·4911 |
| $2\times10^8$ | 0·0398 | 0·0325 | 0·9183 | 1·4229 | 0·1544 | 0·0727 | 0·0779 | 0·0722 | 1·4229 |

| Fold change TcpA IgA | | | | | | | | | |
|---|---|---|---|---|---|---|---|---|---|
| **Dose (CFU)** | **D1** | **D4** | **D7** | **D15** | **D29** | **D57** | **D85** | **D180** | **max FC** |
| 0 | 1·00 | 1·18 | 1·18 | 1·11 | 1·10 | 1·04 | 1·03 | 1·14 | 1·18 |
| 0 | 1·00 | 0·79 | 0·95 | 0·80 | 1·00 | 0·96 | 0·99 | 0·93 | 1·00 |
| 0 | 1·00 | 1·16 | 0·93 | 0·74 | 1·06 | 1·20 | 1·00 | 0·90 | 1·20 |
| 0 | 1·00 | 0·80 | 0·94 | 0·83 | 0·64 | 0·74 | 0·71 | 0·97 | 0·97 |
| 0 | 1·00 | 1·17 | 1·61 | 1·23 | 1·29 | 1·71 | 1·39 | 1·26 | 1·71 |
| 0 | 1·00 | 1·58 | 1·22 | 0·66 | 0·77 | 0·83 | 0·90 | 0·98 | 1·58 |
| 0 | 1·00 | 0·77 | 0·87 | 0·85 | 1·18 | 0·76 | 0·47 | 0·72 | 1·18 |
| $2\times10^7$ | 1·00 | 1·05 | 0·97 | 1·89 | 1·59 | 1·11 | 1·25 | 1·25 | 1·89 |
| $2\times10^7$ | 1·00 | 1·31 | 1·45 | 1·76 | 1·52 | 1·69 | 2·61 | 1·14 | 2·61 |
| $2\times10^7$ | 1·00 | 0·95 | 0·94 | 0·93 | 1·09 | 0·82 | 0·87 | 0·84 | 1·09 |
| $2\times10^7$ | 1·00 | 1·28 | 1·13 | 0·99 | 1·07 | 1·60 | 1·48 | 1·10 | 1·60 |
| $2\times10^7$ | 1·00 | 0·82 | 1·13 | 1·73 | 1·69 | 1·43 | 1·15 | 0·72 | 1·73 |
| $2\times10^7$ | 1·00 | 1·10 | 1·42 | 1·69 | 1·87 | 1·60 | 1·62 | 1·03 | 1·87 |
| $2\times10^7$ | 1·00 | | 0·87 | 0·96 | 0·76 | 0·68 | 0·73 | | 0·96 |
| $2\times10^7$ | 1·00 | 1·23 | 1·84 | 2·78 | 2·32 | 1·48 | 1·22 | 1·18 | 2·78 |
| $2\times10^7$ | 1·00 | 0·83 | 1·00 | 1·24 | 1·12 | 1·13 | 1·11 | 1·34 | 1·34 |
| $2\times10^7$ | 1·00 | 1·28 | 0·86 | 2·03 | 1·54 | 2·01 | 1·67 | 1·07 | 2·03 |
| $2\times10^7$ | 1·00 | 0·70 | 0·80 | 1·03 | 1·64 | 2·73 | 1·90 | 1·75 | 2·73 |
| $2\times10^7$ | 1·00 | 1·13 | 1·65 | 1·18 | 1·15 | 1·02 | 1·47 | 1·38 | 1·65 |
| $2\times10^7$ | 1·00 | 1·06 | 1·44 | 2·29 | 1·95 | 1·49 | 1·66 | 0·87 | 2·29 |
| $2\times10^7$ | 1·00 | 0·81 | 0·96 | 0·59 | 0·68 | 0·79 | | 0·56 | 0·96 |
| $2\times10^8$ | 1·00 | 1·19 | 0·93 | 1·40 | 1·22 | 0·83 | 1·11 | 1·21 | 1·40 |
| $2\times10^8$ | 1·00 | 1·31 | 1·24 | 1·04 | 1·35 | 1·39 | 1·18 | 0·63 | 1·39 |
| $2\times10^8$ | 1·00 | 1·02 | 1·20 | 1·35 | 1·57 | 1·78 | 0·98 | 1·27 | 1·78 |
| $2\times10^8$ | 1·00 | 0·93 | 1·03 | 0·98 | 1·42 | 1·94 | 1·24 | 1·20 | 1·94 |
| $2\times10^8$ | 1·00 | 0·62 | 1·12 | 1·04 | 0·65 | 0·57 | 0·45 | 0·39 | 1·12 |
| $2\times10^8$ | 1·00 | 0·85 | 0·83 | 0·94 | | | | | 0·94 |
| $2\times10^8$ | 1·00 | 1·28 | 1·33 | 3·08 | 1·61 | 1·52 | 1·60 | 1·19 | 3·08 |
| $2\times10^8$ | 1·00 | 0·75 | 0·82 | 0·80 | 0·55 | 0·71 | 0·71 | 0·76 | 0·82 |
| $2\times10^8$ | 1·00 | 0·84 | 0·94 | 0·81 | 1·16 | 1·06 | 1·05 | 0·92 | 1·16 |
| $2\times10^8$ | 1·00 | 1·00 | 1·06 | 1·01 | 1·29 | 1·03 | 0·95 | 0·82 | 1·29 |
| $2\times10^8$ | 1·00 | 1·17 | 1·14 | 1·27 | 1·05 | 1·07 | 1·06 | 1·03 | 1·27 |
| $2\times10^8$ | 1·00 | 1·22 | 1·04 | 0·88 | 0·98 | | 1·20 | 1·46 | 1·46 |
| $2\times10^8$ | 1·00 | 0·82 | 23·08 | 35·76 | 3·88 | 1·83 | 1·96 | 1·82 | 35·76 |

| | RAU | | |
|---|---|---|---|
| | D1 | D39 | |
| *V. cholerae* | 0·3990 | 0·7217 | |
| | 0·3895 | 0·4397 | |
| | 0·3091 | 0·3357 | |
| | 0·1042 | 1·5261 | |
| | 2·4698 | 1·8283 | |
| | D1 | D11 | |
| Vaxchora D11 Challenge | 0·4026 | 0·4608 | |
| | 0·8187 | 0·8196 | |
| | 0·8873 | 0·6844 | |
| | 0·2863 | 0·3028 | |
| | 0·2530 | 0·2752 | |
| | 0·5529 | 0·2441 | |
| | D1 | D11 | D91 |
| Vaxchora D91 Challenge | 0·1284 | 0·1205 | 0·1833 |
| | 0·1284 | 0·1979 | 0·2002 |
| | 0·1910 | 0·3535 | 0·2036 |
| | 0·1442 | 0·2344 | 0·1894 |
| | 0·1289 | 0·2750 | 0·4008 |
| | 0·1706 | 0·1961 | 0·2152 |

| | RAU fold change | | | max FC |
|---|---|---|---|---|
| | D1 | D39 | | |
| *V. cholerae* | 1·00 | 1·81 | | 1·81 |
| | 1·00 | 1·13 | | 1·13 |
| | 1·00 | 1·09 | | 1·09 |
| | 1·00 | 14·64 | | 14·64 |
| | 1·00 | 0·74 | | 0·74 |
| | D1 | D11 | | |
| Vaxchora D11 Challenge | 1·00 | 1·14 | | 1·14 |
| | 1·00 | 1·00 | | 1·00 |
| | 1·00 | 0·77 | | 0·77 |
| | 1·00 | 1·06 | | 1·06 |
| | 1·00 | 1·09 | | 1·09 |
| | 1·00 | 0·44 | | 0·44 |
| | D1 | D11 | D91 | |
| Vaxchora D91 Challenge | 1·00 | 0·94 | 1·43 | 1·43 |
| | 1·00 | 1·54 | 1·56 | 1·56 |
| | 1·00 | 1·85 | 1·07 | 1·85 |
| | 1·00 | 1·63 | 1·31 | 1·63 |
| | 1·00 | 2·13 | 3·11 | 3·11 |
| | 1·00 | 1·15 | 1·26 | 1·26 |

**Table S10: Isotype and antigen specific immune responses in antibody lymphocyte supernatants of vaccine and placebo recipients.** Dark blue: no serum sample; yellow: below limit of detection.

**Dose-Escalation Module**

| | Inaba IgM (RAU) | | | | | | | |
|---|---|---|---|---|---|---|---|---|
| **Dose (CFU)** | **D1** | **D7** | **D15** | **D29** | **D57** | **D85** | **D180** | **max** |
| $10^{5}$ | | 1·95E-06 | | 1·49E-05 | | | | 1·49E-05 |
| $10^{5}$ | | 3·89E-04 | 1·55E-04 | 4·24E-05 | | | | 3·89E-04 |
| $10^{5}$ | 6·55E-06 | 9·26E-03 | 5·88E-05 | 1·55E-05 | | | | 9·26E-03 |
| $10^{6}$ | | 2·71E-01 | | 1·39E-05 | 8·96E-05 | 1·79E-05 | | 2·71E-01 |
| $10^{6}$ | | 9·23E-04 | | | | | | 9·23E-04 |
| $10^{6}$ | | 4·43E-03 | 1·15E-02 | | | | | 1·15E-02 |
| $10^{7}$ | | 9·79E-03 | 4·74E-05 | 3·33E-04 | 6·20E-05 | 1·51E-05 | | 9·79E-03 |
| $10^{7}$ | | 1·97E-02 | 2·78E-05 | | 4·68E-06 | | | 1·97E-02 |
| $10^{7}$ | 3·28E-06 | 2·19E-03 | 3·57E-04 | 6·34E-05 | | 6·23E-06 | | 2·19E-03 |
| $10^{8}$ | | 2·02E-04 | | 1·63E-06 | | 5·29E-06 | | 2·02E-04 |
| $10^{8}$ | | 4·18E-02 | 2·60E-05 | 3·73E-05 | 6·82E-05 | 1·14E-04 | 1·45E-05 | 4·18E-02 |
| $10^{8}$ | | 8·30E-02 | 1·07E-06 | 1·23E-05 | 7·55E-05 | 4·62E-06 | | 8·30E-02 |
| $10^{9}$ | | 2·20E-01 | 1·00E-03 | 5·98E-04 | 7·90E-06 | | 1·64E-04 | 2·20E-01 |
| $10^{9}$ | | 7·76E-04 | 1·40E-03 | 1·27E-03 | 4·06E-04 | 1·00E-04 | | 1·40E-03 |
| $10^{9}$ | | 1·80E-04 | | 1·89E-05 | | 3·05E-05 | 9·70E-06 | 1·80E-04 |
| $10^{10}$ | | 2·60E-03 | | 5·01E-05 | | 3·40E-06 | | 2·60E-03 |
| $10^{10}$ | | 1·13E+00 | 2·52E-04 | | | 3·88E-06 | | 1·13E+00 |
| $10^{10}$ | | 2·59E-01 | 1·64E-04 | 8·76E-04 | 4·48E-04 | 5·87E-05 | 5·30E-07 | 2·59E-01 |

**Dose- Expansion Module**

| | Inaba IgM (RAU) | | | | | | | |
|---|---|---|---|---|---|---|---|---|
| **Dose (CFU)** | **D1** | **D7** | **D15** | **D29** | **D57** | **D85** | **D180** | **max** |
| 0 | 1·59E-03 | 1·65E-03 | 1·47E-03 | 8·73E-05 | 2·79E-03 | | 5·05E-03 | 5·05E-03 |
| 0 | 1·57E-05 | 2·30E-05 | | 3·79E-06 | | | | 2·30E-05 |
| 0 | 3·28E-05 | 9·03E-06 | 2·30E-05 | 3·00E-05 | | | | 3·28E-05 |
| 0 | 2·58E-05 | 1·70E-05 | 6·28E-06 | 2·03E-05 | | | 5·00E-06 | 2·58E-05 |
| 0 | | 6·81E-06 | 2·30E-05 | 4·26E-06 | | | 3·14E-06 | 2·30E-05 |
| 0 | 4·67E-05 | 5·21E-07 | | | | | 3·57E-06 | 4·67E-05 |
| 0 | 6·00E-06 | 7·08E-06 | | | | | | 7·08E-06 |
| $2\times10^{7}$ | 1·47E-03 | | 4·15E-03 | 3·24E-03 | | | 3·31E-03 | 4·15E-03 |
| $2\times10^{7}$ | 1·78E-03 | 1·14E-01 | 7·82E-03 | 6·22E-03 | | | 3·11E-03 | 1·14E-01 |
| $2\times10^{7}$ | 3·34E-06 | 6·67E-01 | 3·23E-04 | 1·68E-03 | | | 4·74E-05 | 6·67E-01 |
| $2\times10^{7}$ | 1·57E-05 | 3·64E-05 | 3·57E-05 | 2·79E-05 | | | 4·50E-06 | 3·64E-05 |
| $2\times10^{7}$ | 2·91E-06 | 1·78E-02 | 1·71E-04 | 8·33E-04 | | | | 1·78E-02 |
| $2\times10^{7}$ | 5·24E-06 | 2·27E-03 | 2·17E-05 | 2·72E-05 | | | | 2·27E-03 |
| $2\times10^{7}$ | 2·65E-05 | 4·37E-05 | 2·37E-05 | 1·71E-04 | | | | 1·71E-04 |
| $2\times10^{7}$ | 1·32E-05 | 1·48E-02 | | 8·46E-06 | | | 1·28E-06 | 1·48E-02 |
| $2\times10^{7}$ | 5·24E-06 | 1·24E-01 | 2·84E-04 | 7·25E-04 | | | 9·74E-07 | 1·24E-01 |
| $2\times10^{7}$ | 2·91E-06 | 9·47E-03 | 2·10E-03 | 4·15E-05 | | | 1·95E-06 | 9·47E-03 |
| $2\times10^{7}$ | 1·97E-05 | 3·06E-03 | 1·11E-05 | 5·03E-05 | | | 6·55E-06 | 3·06E-03 |
| $2\times10^{7}$ | | 3·07E-03 | 4·77E-03 | 6·37E-05 | | | 7·08E-06 | 4·77E-03 |
| $2\times10^{7}$ | 3·14E-06 | 9·34E-02 | 7·79E-05 | 3·39E-05 | | | 1·62E-05 | 9·34E-02 |
| $2\times10^{7}$ | | 4·77E-03 | 1·20E-04 | 1·81E-05 | | | 1·30E-05 | 4·77E-03 |
| $2\times10^{8}$ | 1·11E-03 | 9·13E+01 | 1·66E-01 | 6·08E-02 | 2·15E-02 | | 2·34E-03 | 9·13E+01 |
| $2\times10^{8}$ | 2·15E-03 | 4·64E-01 | 8·79E-04 | 8·77E-03 | | | | 4·64E-01 |
| $2\times10^{8}$ | 6·28E-06 | 4·45E-02 | 1·93E-04 | 4·23E-05 | | | 1·24E-05 | 4·45E-02 |
| $2\times10^{8}$ | 6·81E-06 | 1·43E-03 | 9·77E-04 | 1·92E-04 | | | | 1·43E-03 |
| $2\times10^{8}$ | 7·90E-06 | 2·22E-02 | 2·72E-05 | 1·32E-05 | | | 3·14E-06 | 2·22E-02 |
| $2\times10^{8}$ | | | 1·57E-05 | | | | | 1·57E-05 |
| $2\times10^{8}$ | 1·37E-06 | 3·79E-05 | 3·71E-05 | 7·41E-05 | | | | 7·41E-05 |
| $2\times10^{8}$ | 1·05E-06 | 1·97E-05 | 1·77E-05 | 6·28E-06 | | | | 1·97E-05 |
| $2\times10^{8}$ | 1·02E-05 | 1·76E+00 | 4·79E-03 | 2·22E-04 | | | 4·63E-04 | 1·76E+00 |
| $2\times10^{8}$ | | 3·65E-02 | 2·15E-05 | 5·60E-04 | | | | 3·65E-02 |
| $2\times10^{8}$ | 1·68E-05 | 4·46E-04 | 3·70E-04 | 5·80E-05 | | | 2·76E-05 | 4·46E-04 |
| $2\times10^{8}$ | | 1·93E-03 | 1·95E-06 | 2·42E-05 | | | | 1·93E-03 |
| $2\times10^{8}$ | | 2·37E-02 | 2·21E-05 | 5·88E-05 | | | | 2·37E-02 |

no ALS sample
below limit of detection

D57 and D85 ALS samples were mainly collected during the dose-escalation module.

**Dose-Escalation Module**

| | Inaba IgG (RAU) | | | | | | | |
|---|---|---|---|---|---|---|---|---|
| **Dose (CFU)** | **D1** | **D7** | **D15** | **D29** | **D57** | **D85** | **D180** | **max** |
| $10^{5}$ | | | | 4·61E-04 | | | 7·87E-05 | 4·61E-04 |
| $10^{5}$ | | | 7·33E-04 | | | | | 7·33E-04 |
| $10^{5}$ | | | 7·87E-05 | | | | | 7·87E-05 |
| $10^{6}$ | | 1·27E-04 | 5·84E-05 | | 1·62E-03 | 7·25E-04 | | 1·62E-03 |
| $10^{6}$ | 1·27E-04 | 3·76E-04 | | | 1·44E-04 | | | 3·76E-04 |
| $10^{6}$ | 8·59E-05 | 2·50E-02 | 5·78E-02 | 5·84E-05 | 1·44E-04 | 5·84E-05 | 1·35E-04 | 5·78E-02 |
| $10^{7}$ | | 5·84E-05 | | | 1·27E-04 | 2·29E-04 | 2·10E-03 | 2·10E-03 |
| $10^{7}$ | | 3·66E-03 | 1·08E-04 | 2·97E-04 | | 5·84E-05 | 8·85E-05 | 3·66E-03 |
| $10^{7}$ | | | | | | 2·97E-04 | 1·13E-04 | 2·97E-04 |
| $10^{8}$ | | | 1·27E-04 | | 2·53E-04 | | 4·26E-04 | 4·26E-04 |
| $10^{8}$ | | 2·64E-03 | 5·84E-05 | | | 1·24E-04 | | 2·64E-03 |
| $10^{8}$ | | | | 1·27E-04 | 1·69E-05 | 2·65E-04 | 1·78E-04 | 2·65E-04 |
| $10^{9}$ | 5·84E-05 | | | | 1·78E-04 | 3·34E-05 | | 1·78E-04 |
| $10^{9}$ | | | 4·85E-05 | 3·01E-04 | 8·85E-05 | 2·31E-03 | | 2·31E-03 |
| $10^{9}$ | 1·69E-05 | 7·58E-05 | 8·85E-05 | 1·01E-04 | 4·85E-05 | | 8·85E-05 | 1·01E-04 |
| $10^{10}$ | 6·25E-05 | 2·08E-04 | 1·24E-04 | 6·25E-05 | | 7·58E-05 | 7·58E-05 | 2·08E-04 |
| $10^{10}$ | 3·34E-05 | 1·69E-05 | | | | 1·69E-05 | 4·85E-05 | 4·85E-05 |
| $10^{10}$ | 1·01E-04 | 8·85E-05 | 1·01E-04 | | 1·01E-04 | | 2·83E-04 | 2·83E-04 |

**Dose- Expansion Module**

| | Inaba IgG (RAU) | | | | | | | |
|---|---|---|---|---|---|---|---|---|
| **Dose (CFU)** | **D1** | **D7** | **D15** | **D29** | **D57** | **D85** | **D180** | **max** |
| 0 | 4·67E-03 | 6·48E-03 | 2·16E-03 | 1·78E-03 | 5·79E-03 | | 1·25E-03 | 6·48E-03 |
| 0 | 1·46E-04 | | | 5·22E-05 | | | 1·81E-04 | 1·81E-04 |
| 0 | 8·10E-05 | 8·10E-05 | 4·82E-05 | 4·43E-05 | | | | 8·10E-05 |
| 0 | | 1·51E-04 | 3·30E-05 | 1·42E-04 | | | | 1·51E-04 |
| 0 | 6·02E-05 | 1·95E-04 | 8·95E-05 | 8·53E-05 | | | | 1·95E-04 |
| 0 | 8·53E-05 | 8·10E-05 | | | | | | 8·53E-05 |
| 0 | | | | | | | | |
| $2\times10^{7}$ | 6·35E-04 | 7·63E-03 | 1·78E-03 | 9·27E-04 | | | 1·27E-02 | 1·27E-02 |
| $2\times10^{7}$ | 5·12E-03 | 7·11E-02 | 5·12E-03 | 1·25E-03 | | | 9·02E-03 | 7·11E-02 |
| $2\times10^{7}$ | 5·62E-05 | 5·30E-04 | 1·92E-05 | 2·24E-05 | | | 7·26E-05 | 5·30E-04 |
| $2\times10^{7}$ | | 8·10E-05 | | 5·62E-05 | | | | 8·10E-05 |
| $2\times10^{7}$ | 8·95E-05 | 1·68E-04 | 1·51E-04 | 2·93E-05 | | | 4·40E-04 | 4·40E-04 |
| $2\times10^{7}$ | 1·92E-05 | 6·02E-05 | 8·08E-07 | | | | | 6·02E-05 |
| $2\times10^{7}$ | | 1·07E-04 | 3·67E-05 | 3·30E-05 | | | | 1·07E-04 |
| $2\times10^{7}$ | 4·04E-05 | 1·29E-04 | 6·43E-05 | 4·43E-05 | | | | 1·29E-04 |
| $2\times10^{7}$ | 1·73E-04 | 2·04E-04 | 8·10E-05 | 1·42E-04 | | | | 2·04E-04 |
| $2\times10^{7}$ | | 3·30E-05 | 1·02E-04 | | | | | 1·02E-04 |
| $2\times10^{7}$ | | 2·43E-04 | | | | | 3·10E-04 | 3·10E-04 |
| $2\times10^{7}$ | | | | | | | | |
| $2\times10^{7}$ | | | | | | | | |
| $2\times10^{7}$ | | | | | | | | |
| $2\times10^{8}$ | 5·12E-03 | 5·79E-03 | 7·86E-03 | 7·63E-03 | 4·23E-03 | | 1·37E-02 | 1·37E-02 |
| $2\times10^{8}$ | 2·35E-03 | 6·35E-04 | 7·16E-03 | 9·27E-04 | | | 2·12E-05 | 7·16E-03 |
| $2\times10^{8}$ | 1·92E-05 | 2·70E-04 | | 1·31E-05 | | | | 2·70E-04 |
| $2\times10^{8}$ | 1·02E-04 | 1·59E-04 | 1·46E-04 | 3·48E-04 | | | 7·87E-05 | 3·48E-04 |
| $2\times10^{8}$ | 2·08E-04 | 9·38E-05 | 4·82E-05 | 5·50E-06 | | | | 2·08E-04 |
| $2\times10^{8}$ | | | 6·02E-05 | | | | | 6·02E-05 |
| $2\times10^{8}$ | 1·07E-04 | 2·93E-05 | 2·93E-05 | 1·24E-04 | | | | 1·24E-04 |
| $2\times10^{8}$ | 6·43E-05 | | 6·43E-05 | 1·29E-04 | | | | 1·29E-04 |
| $2\times10^{8}$ | | 1·11E-03 | 2·58E-05 | 1·92E-05 | | | 9·73E-04 | 1·11E-03 |
| $2\times10^{8}$ | | | | | | | | |
| $2\times10^{8}$ | | | | | | | 1·68E-03 | 1·68E-03 |
| $2\times10^{8}$ | | | | 5·42E-04 | | | | 5·42E-04 |
| $2\times10^{8}$ | 6·93E-06 | 4·47E-06 | 3·35E-06 | 1·89E-05 | | | 6·05E-05 | 6·05E-05 |

**Dose-Escalation Module**

| Inaba IgA (RAU) | | | | | | | | |
|---|---|---|---|---|---|---|---|---|
| Dose (CFU) | D1 | D7 | D15 | D29 | D57 | D85 | D180 | max |
| $10^5$ | 1·10E-05 | 1·18E-04 | 9·82E-06 | 1·10E-05 | | | 1·93E-05 | 1·18E-04 |
| $10^5$ | 1·57E-05 | 3·10E-05 | 2·25E-04 | 5·78E-05 | | | | 2·25E-04 |
| $10^5$ | 2·28E-05 | 7·36E-05 | 3·50E-05 | 1·28E-04 | | | | 1·28E-04 |
| $10^6$ | 7·03E-06 | 6·81E-03 | 6·48E-05 | 8·60E-05 | 9·36E-04 | 8·67E-04 | | 6·81E-03 |
| $10^6$ | 6·58E-06 | 1·14E-01 | 1·28E-04 | 5·57E-05 | 2·25E-04 | 3·58E-05 | 2·21E-05 | 1·14E-01 |
| $10^6$ | 1·31E-05 | 1·89E-03 | 1·34E-01 | 2·51E-05 | 1·12E-05 | 2·44E-06 | 1·90E-06 | 1·34E-01 |
| $10^7$ | 7·75E-06 | 1·15E-06 | 1·41E-06 | 6·80E-06 | 1·92E-05 | 4·47E-04 | 3·06E-04 | 4·47E-04 |
| $10^7$ | 9·68E-06 | 1·46E-02 | 5·01E-05 | 2·63E-05 | 7·82E-06 | 4·87E-05 | 9·37E-06 | 1·46E-02 |
| $10^7$ | 1·64E-06 | 4·70E-04 | 2·61E-04 | 1·22E-05 | 9·06E-06 | 7·82E-06 | 4·16E-06 | 4·70E-04 |
| $10^8$ | 9·06E-06 | 2·90E-05 | 1·41E-05 | 2·45E-05 | | 1·16E-05 | 5·26E-05 | 5·26E-05 |
| $10^8$ | 1·72E-05 | 9·70E-04 | 1·28E-05 | 1·22E-05 | 5·01E-05 | 2·02E-05 | 9·29E-05 | 9·70E-04 |
| $10^8$ | 5·10E-07 | 2·61E-02 | 1·92E-05 | 5·13E-05 | 3·29E-04 | 2·06E-06 | | 2·61E-02 |
| $10^9$ | 9·06E-06 | 1·20E-04 | 3·17E-05 | 3·65E-04 | 8·93E-05 | 3·11E-05 | 9·06E-06 | 3·65E-04 |
| $10^9$ | 3·20E-06 | 1·05E-03 | 2·67E-04 | 1·54E-03 | 2·71E-04 | 9·60E-04 | | 1·54E-03 |
| $10^9$ | 2·44E-06 | 1·72E-05 | 8·44E-06 | 1·84E-05 | 3·29E-05 | 8·44E-06 | 3·40E-05 | 3·40E-05 |
| $10^{10}$ | 5·75E-07 | 4·92E-03 | 2·04E-05 | 1·68E-05 | | | | 4·92E-03 |
| $10^{10}$ | 5·97E-06 | 3·87E-05 | 2·52E-04 | | | 1·27E-04 | 2·57E-04 | 2·57E-04 |
| $10^{10}$ | 2·16E-06 | 4·97E-03 | 1·08E-03 | 2·56E-03 | 8·32E-04 | 6·45E-04 | 1·54E-03 | 4·97E-03 |

**Dose- Expansion Module**

| Inaba IgA (RAU) | | | | | | | | |
|---|---|---|---|---|---|---|---|---|
| Dose (CFU) | D1 | D7 | D15 | D29 | D57 | D85 | D180 | max |
| 0 | 1·90E-06 | 4·16E-06 | 3·28E-06 | | 2·99E-06 | | 2·44E-06 | 4·16E-06 |
| 0 | 4·75E-06 | 5·36E-06 | 1·16E-05 | 4·75E-06 | | | 8·13E-06 | 1·16E-05 |
| 0 | 2·16E-06 | 2·99E-06 | | 5·97E-06 | | | 1·28E-05 | 1·28E-05 |
| 0 | 5·29E-06 | 1·10E-05 | 7·52E-06 | 2·93E-05 | | | 1·93E-05 | 2·93E-05 |
| 0 | 1·93E-05 | 9·24E-06 | 1·45E-05 | 9·82E-06 | | | 1·22E-05 | 1·93E-05 |
| 0 | 1·69E-05 | 7·52E-06 | 2·69E-05 | 4·23E-06 | | | 1·10E-05 | 2·69E-05 |
| 0 | 1·69E-05 | 6·95E-06 | 3·21E-06 | 1·22E-05 | | | 2·05E-05 | 2·05E-05 |
| $2\times10^7$ | 7·51E-06 | 1·54E-03 | 4·16E-06 | 7·82E-06 | | | 3·57E-06 | 1·54E-03 |
| $2\times10^7$ | | 2·06E-01 | 5·60E-02 | 8·28E-02 | | | 9·81E-05 | 2·06E-01 |
| $2\times10^7$ | 1·59E-05 | 1·97E-03 | 4·47E-04 | 7·07E-07 | | | 1·16E-05 | 1·97E-03 |
| $2\times10^7$ | 6·89E-06 | 1·12E-05 | 8·13E-06 | 6·58E-06 | | | 9·06E-06 | 1·12E-05 |
| $2\times10^7$ | 1·57E-05 | 1·75E-03 | 1·42E-04 | 3·83E-04 | | | 2·64E-04 | 1·75E-03 |
| $2\times10^7$ | 5·29E-06 | 5·45E-04 | 7·36E-04 | 6·26E-05 | | | 9·13E-05 | 7·36E-04 |
| $2\times10^7$ | 7·52E-06 | 4·57E-05 | 2·10E-05 | 2·52E-05 | | | | 4·57E-05 |
| $2\times10^7$ | 8·66E-06 | 3·62E-05 | 1·81E-05 | 2·16E-05 | | | 5·84E-06 | 3·62E-05 |
| $2\times10^7$ | 2·16E-05 | 1·26E-03 | 4·50E-04 | 3·20E-03 | | | 1·93E-04 | 3·20E-03 |
| $2\times10^7$ | 6·39E-06 | 7·80E-04 | 1·42E-03 | 2·98E-05 | | | 1·38E-04 | 1·42E-03 |
| $2\times10^7$ | 1·51E-05 | 2·40E-02 | 1·07E-03 | 7·95E-04 | | | 1·20E-03 | 2·40E-02 |
| $2\times10^7$ | 2·93E-05 | 6·73E-05 | 2·40E-05 | 1·19E-04 | | | 2·09E-04 | 2·09E-04 |
| $2\times10^7$ | 3·71E-06 | 1·71E-02 | 4·01E-05 | 2·28E-05 | | | 1·81E-05 | 1·71E-02 |
| $2\times10^7$ | 1·40E-06 | 3·14E-04 | 1·70E-04 | 1·81E-06 | | | 9·24E-06 | 3·14E-04 |
| $2\times10^8$ | 1·06E-03 | 3·33E-01 | 2·88E-02 | 6·17E-02 | 3·35E-03 | | 8·42E-03 | 3·33E-01 |
| $2\times10^8$ | 2·97E-03 | 1·24E-02 | 7·81E-04 | | | | 1·59E-05 | 1·24E-02 |
| $2\times10^8$ | 1·10E-05 | 9·67E-05 | 7·51E-05 | 9·18E-05 | | | 8·38E-05 | 9·67E-05 |
| $2\times10^8$ | 1·26E-05 | 4·89E-03 | 2·50E-04 | 7·04E-04 | | | 1·18E-04 | 4·89E-03 |
| $2\times10^8$ | 9·05E-06 | 1·15E-04 | 5·79E-06 | 4·32E-06 | | | | 1·15E-04 |
| $2\times10^8$ | | | 2·43E-05 | | | | | 2·43E-05 |
| $2\times10^8$ | 4·32E-05 | 7·38E-06 | 7·19E-05 | 5·64E-05 | | | | 7·19E-05 |
| $2\times10^8$ | 2·43E-05 | 1·63E-05 | | 2·43E-05 | | | | 2·43E-05 |
| $2\times10^8$ | 1·44E-05 | 1·12E-02 | 4·58E-04 | 2·22E-05 | | | 2·36E-04 | 1·12E-02 |
| $2\times10^8$ | 1·04E-05 | 8·40E-04 | 2·19E-05 | 1·84E-04 | | | 3·45E-05 | 8·40E-04 |
| $2\times10^8$ | 1·10E-05 | 4·23E-06 | 2·28E-05 | 1·45E-05 | | | 7·46E-05 | 7·46E-05 |
| $2\times10^8$ | 3·16E-05 | 2·39E-04 | 2·69E-05 | 6·52E-05 | | | 1·40E-06 | 2·39E-04 |
| $2\times10^8$ | 2·87E-05 | 2·14E-04 | 1·63E-05 | 1·39E-05 | | | 2·40E-05 | 2·14E-04 |

**Dose-Escalation Module**

| | Ogawa IgM (RAU) | | | | | | | |
|---|---|---|---|---|---|---|---|---|
| **Dose (CFU)** | **D1** | **D7** | **D15** | **D29** | **D57** | **D85** | **D180** | **max** |
| $10^5$ | | 8·94E-05 | | 8·31E-05 | | | | 8·94E-05 |
| $10^5$ | | 8·01E-05 | | 5·27E-04 | | | | 5·27E-04 |
| $10^5$ | | | | | | | | |
| $10^6$ | | 1·03E+00 | 7·42E-05 | 2·84E-06 | 4·55E-05 | 1·73E-05 | | 1·03E+00 |
| $10^6$ | 7·88E-07 | 1·61E-03 | | | | 1·27E-06 | | 1·61E-03 |
| $10^6$ | | 8·79E-02 | 2·95E-02 | 3·23E-05 | 1·14E-05 | 1·54E-06 | | 8·79E-02 |
| $10^7$ | | 4·71E-02 | 9·45E-04 | 4·07E-04 | 8·80E-05 | 2·04E-05 | 1·05E-05 | 4·71E-02 |
| $10^7$ | 5·88E-07 | 9·15E-02 | 6·99E-05 | 1·31E-05 | 3·23E-05 | | 2·53E-06 | 9·15E-02 |
| $10^7$ | | 1·76E-03 | 1·07E-04 | | | | 3·42E-06 | 1·76E-03 |
| $10^8$ | 5·88E-07 | 3·29E-03 | 1·25E-04 | 2·96E-05 | 1·27E-06 | 8·20E-06 | | 3·29E-03 |
| $10^8$ | 1·37E-05 | 8·91E-03 | 2·23E-04 | 1·23E-04 | 6·22E-06 | 5·46E-05 | 3·11E-06 | 8·91E-03 |
| $10^8$ | | 2·86E-03 | 2·49E-06 | 7·88E-07 | | | | 2·86E-03 |
| $10^9$ | | 2·99E-02 | 8·51E-05 | 1·51E-04 | 1·71E-05 | | 9·34E-05 | 2·99E-02 |
| $10^9$ | | 2·70E-05 | 6·09E-05 | 4·06E-04 | 4·89E-05 | 5·67E-05 | | 4·06E-04 |
| $10^9$ | | 1·93E-03 | 5·31E-05 | | 4·74E-06 | 1·71E-05 | 4·32E-05 | 1·93E-03 |
| $10^{10}$ | | 1·74E-03 | 4·07E-06 | | | | | 1·74E-03 |
| $10^{10}$ | | 2·40E-02 | 8·05E-05 | | | | 5·25E-05 | 2·40E-02 |
| $10^{10}$ | | 3·09E-01 | 1·30E-04 | 1·41E-04 | 1·90E-04 | 2·57E-05 | 1·25E-06 | 3·09E-01 |

**Dose- Expansion Module**

| | Ogawa IgM (RAU) | | | | | | | |
|---|---|---|---|---|---|---|---|---|
| **Dose (CFU)** | **D1** | **D7** | **D15** | **D29** | **D57** | **D85** | **D180** | **max** |
| 0 | | | | | | | | |
| 0 | | | | | | | 5·06E-06 | 5·06E-06 |
| 0 | 2·79E-05 | | | 1·71E-05 | | | | 2·79E-05 |
| 0 | 1·05E-06 | | 5·06E-06 | | | | | 5·06E-06 |
| 0 | 1·26E-05 | 6·39E-06 | | | | | | 1·26E-05 |
| 0 | | | 1·63E-05 | | | | | 1·63E-05 |
| 0 | | 2·02E-05 | | | | | | 2·02E-05 |
| $2{\times}10^7$ | | 1·76E-02 | | | | | 1·51E-04 | 1·76E-02 |
| $2{\times}10^7$ | | 9·57E+00 | 6·64E-03 | | | | 5·24E-03 | 9·57E+00 |
| $2{\times}10^7$ | 2·39E-05 | 1·54E-01 | | 8·31E-05 | | | | 1·54E-01 |
| $2{\times}10^7$ | 1·05E-06 | 1·03E-04 | 5·06E-06 | | | | | 1·03E-04 |
| $2{\times}10^7$ | | 1·75E-02 | 2·22E-04 | 2·59E-05 | | | | 1·75E-02 |
| $2{\times}10^7$ | 7·83E-06 | 5·25E-02 | 4·18E-04 | 1·50E-04 | | | | 5·25E-02 |
| $2{\times}10^7$ | 1·44E-05 | 1·43E-03 | 3·21E-05 | 4·99E-05 | | | | 1·43E-03 |
| $2{\times}10^7$ | 1·81E-06 | 1·81E-02 | 8·67E-05 | | | | | 1·81E-02 |
| $2{\times}10^7$ | 1·01E-05 | 5·80E-05 | | 1·81E-06 | | | | 5·80E-05 |
| $2{\times}10^7$ | 4·86E-07 | 1·23E-02 | 2·86E-03 | 1·53E-05 | | | | 1·23E-02 |
| $2{\times}10^7$ | | 9·87E-03 | 2·68E-04 | | | | | 9·87E-03 |
| $2{\times}10^7$ | | 2·24E-03 | 3·66E-04 | 8·47E-05 | | | | 2·24E-03 |
| $2{\times}10^7$ | | 7·81E-04 | | 9·66E-07 | | | | 7·81E-04 |
| $2{\times}10^7$ | | 1·56E-03 | 2·22E-05 | | | | 2·02E-05 | 1·56E-03 |
| $2{\times}10^8$ | | 2·07E-01 | 3·50E-03 | 3·57E-02 | | | | 2·07E-01 |
| $2{\times}10^8$ | 5·09E-04 | 2·41E-01 | | 6·81E-04 | | | | 2·41E-01 |
| $2{\times}10^8$ | 1·05E-06 | 1·34E-02 | 2·75E-06 | 1·01E-05 | | | | 1·34E-02 |
| $2{\times}10^8$ | 4·43E-06 | 7·74E-02 | 2·42E-04 | 6·15E-05 | | | | 7·74E-02 |
| $2{\times}10^8$ | 3·28E-06 | 7·82E-03 | | | | | | 7·82E-03 |
| $2{\times}10^8$ | | | 2·26E-04 | | | | | 2·26E-04 |
| $2{\times}10^8$ | 2·75E-06 | 1·48E-03 | 1·45E-04 | 4·86E-07 | | | | 1·48E-03 |
| $2{\times}10^8$ | | 2·18E-03 | 1·62E-05 | 2·75E-06 | | | | 2·18E-03 |
| $2{\times}10^8$ | 7·43E-07 | 2·15E+00 | 5·79E-03 | 3·05E-04 | | | 5·55E-04 | 2·15E+00 |
| $2{\times}10^8$ | | 1·58E-01 | 2·51E-04 | 5·85E-04 | | | | 1·58E-01 |
| $2{\times}10^8$ | | 3·82E-02 | 8·24E-04 | 1·63E-05 | | | | 3·82E-02 |
| $2{\times}10^8$ | | 2·42E-03 | 1·54E-04 | | | | | 2·42E-03 |
| $2{\times}10^8$ | | | | | | | | |

**Dose-Escalation Module**

| | Ogawa IgG (RAU) | | | | | | | |
|---|---|---|---|---|---|---|---|---|
| Dose (CFU) | D1 | D7 | D15 | D29 | D57 | D85 | D180 | max |
| $10^5$ | | 4·63E-06 | | 2·59E-04 | | | 8·68E-06 | 2·59E-04 |
| $10^5$ | | | | | | | | |
| $10^5$ | | | | | | | | |
| $10^6$ | 1·30E-05 | 2·02E-05 | 3·50E-05 | | 8·50E-05 | 9·86E-05 | | 9·86E-05 |
| $10^6$ | 6·26E-06 | 4·24E-05 | 1·30E-05 | | 2·76E-05 | | 4·97E-05 | 4·97E-05 |
| $10^6$ | | 4·15E-02 | 2·02E-01 | 2·11E-04 | 1·12E-04 | 2·02E-05 | 7·28E-05 | 2·02E-01 |
| $10^7$ | 4·24E-05 | 2·02E-05 | 6·26E-06 | 3·50E-05 | 9·92E-07 | 4·24E-05 | 1·45E-04 | 1·45E-04 |
| $10^7$ | 9·92E-07 | 3·05E-04 | 2·02E-05 | 1·05E-04 | 1·30E-05 | 1·30E-05 | | 3·05E-04 |
| $10^7$ | | 1·30E-05 | | 6·26E-06 | 4·97E-05 | 9·86E-05 | 4·93E-05 | 9·86E-05 |
| $10^8$ | 3·50E-05 | | 9·92E-07 | 3·50E-05 | 2·02E-05 | | | 3·50E-05 |
| $10^8$ | 5·69E-05 | 1·19E-04 | 2·02E-05 | 2·02E-05 | | 4·34E-05 | 1·33E-05 | 1·19E-04 |
| $10^8$ | 4·97E-05 | 3·50E-05 | 6·26E-06 | 1·30E-05 | 5·53E-05 | 6·11E-05 | 1·33E-05 | 6·11E-05 |
| $10^9$ | | 3·50E-05 | 2·76E-05 | | 1·92E-05 | | 1·33E-05 | 3·50E-05 |
| $10^9$ | 9·92E-07 | | | 8·42E-05 | 4·93E-05 | 6·07E-04 | | 6·07E-04 |
| $10^9$ | 1·33E-05 | | | | 3·74E-05 | | 7·47E-06 | 3·74E-05 |
| $10^{10}$ | 1·92E-05 | 6·93E-02 | | 1·29E-04 | | | | 6·93E-02 |
| $10^{10}$ | | 3·13E-05 | 1·92E-05 | | | 7·47E-06 | | 3·13E-05 |
| $10^{10}$ | 4·34E-05 | 2·53E-05 | 1·33E-05 | | | 2·31E-06 | 2·53E-05 | 4·34E-05 |

**Dose- Expansion Module**

| | Ogawa IgG (RAU) | | | | | | | |
|---|---|---|---|---|---|---|---|---|
| Dose (CFU) | D1 | D7 | D15 | D29 | D57 | D85 | D180 | max |
| 0 | | | | | | | | |
| 0 | | 1·08E-04 | | | | | 2·26E-05 | 1·08E-04 |
| 0 | | 7·71E-05 | | 1·53E-05 | | | | 7·71E-05 |
| 0 | 8·98E-05 | 9·29E-05 | | 4·77E-05 | | | | 9·29E-05 |
| 0 | 1·27E-04 | 8·36E-05 | 1·50E-04 | 8·67E-05 | | | | 1·50E-04 |
| 0 | | 9·89E-05 | | | | | | 9·89E-05 |
| 0 | | | 1·10E-05 | | | | | 1·10E-05 |
| $2\times10^7$ | | 3·74E-03 | | | | | 7·10E-03 | 7·10E-03 |
| $2\times10^7$ | | 7·46E-01 | 4·72E-03 | | | | | 7·46E-01 |
| $2\times10^7$ | 4·34E-05 | 4·77E-05 | 3·88E-05 | 2·26E-05 | | | 5·18E-05 | 5·18E-05 |
| $2\times10^7$ | 8·67E-05 | 2·86E-05 | | 2·86E-05 | | | | 8·67E-05 |
| $2\times10^7$ | 1·30E-04 | 1·45E-04 | 2·86E-05 | 7·71E-05 | | | 1·10E-05 | 1·45E-04 |
| $2\times10^7$ | 8·36E-05 | 1·13E-04 | 7·71E-05 | | | | | 1·13E-04 |
| $2\times10^7$ | | 8·98E-05 | 5·18E-05 | | | | | 8·98E-05 |
| $2\times10^7$ | 1·19E-04 | | 8·36E-05 | | | | | 1·19E-04 |
| $2\times10^7$ | | 7·71E-05 | | 5·57E-05 | | | | 7·71E-05 |
| $2\times10^7$ | | 9·59E-05 | 7·71E-05 | 3·40E-05 | | | | 9·59E-05 |
| $2\times10^7$ | | 1·53E-05 | | | | | | 1·53E-05 |
| $2\times10^7$ | | 2·10E-05 | | | | | | 2·10E-05 |
| $2\times10^7$ | | | 2·10E-05 | | | | | 2·10E-05 |
| $2\times10^7$ | 1·84E-05 | | | | | | 1·84E-05 | 1·84E-05 |
| $2\times10^8$ | | 5·20E-03 | 2·53E-03 | 3·53E-04 | | | | 5·20E-03 |
| $2\times10^8$ | 6·40E-03 | 7·18E-04 | | 3·26E-03 | | | 1·10E-05 | 6·40E-03 |
| $2\times10^8$ | 1·05E-04 | 1·08E-04 | 5·95E-05 | | | | | 1·08E-04 |
| $2\times10^8$ | | 2·14E-04 | 6·68E-05 | 1·27E-04 | | | | 2·14E-04 |
| $2\times10^8$ | | 1·71E-03 | 4·34E-05 | 5·95E-05 | | | 1·84E-05 | 1·71E-03 |
| $2\times10^8$ | | | 7·37E-05 | | | | | 7·37E-05 |
| $2\times10^8$ | 3·40E-05 | 7·37E-05 | | 1·82E-04 | | | | 1·82E-04 |
| $2\times10^8$ | | | | | | | | |
| $2\times10^8$ | | 5·14E-04 | 7·03E-05 | | | | 4·72E-04 | 5·14E-04 |
| $2\times10^8$ | | 5·31E-04 | | 4·63E-06 | | | | 5·31E-04 |
| $2\times10^8$ | 2·91E-06 | 2·53E-05 | 3·27E-05 | 6·58E-06 | | | 9·16E-04 | 9·16E-04 |
| $2\times10^8$ | | | 6·58E-06 | 1·10E-05 | | | | 1·10E-05 |
| $2\times10^8$ | | | | 6·92E-05 | | | 1·10E-05 | 6·92E-05 |

**Dose-Escalation Module**

| | Ogawa IgA (RAU) | | | | | | | |
|---|---|---|---|---|---|---|---|---|
| **Dose (CFU)** | **D1** | **D7** | **D15** | **D29** | **D57** | **D85** | **D180** | **max** |
| $10^5$ | 4·06E-05 | 7·70E-05 | 4·26E-05 | 3·27E-05 | | | 3·07E-05 | 7·70E-05 |
| $10^5$ | 3·07E-05 | 2·66E-05 | 7·04E-05 | 8·53E-05 | | | | 8·53E-05 |
| $10^5$ | 5·62E-05 | 6·95E-05 | 5·04E-05 | 5·04E-05 | | | | 6·95E-05 |
| $10^6$ | 5·59E-05 | 6·64E-03 | 5·80E-05 | 9·82E-05 | 1·85E-04 | 4·10E-04 | | 6·64E-03 |
| $10^6$ | 1·88E-05 | 1·39E-02 | 3·49E-05 | 5·79E-05 | 6·89E-05 | 2·17E-06 | 4·66E-05 | 1·39E-02 |
| $10^6$ | 6·16E-05 | 2·52E-02 | 6·08E-02 | 5·87E-05 | 8·96E-06 | 1·63E-05 | 4·27E-06 | 6·08E-02 |
| $10^7$ | 4·22E-05 | 1·91E-05 | 1·47E-05 | 1·91E-05 | 2·76E-05 | 3·56E-04 | 3·51E-04 | 3·56E-04 |
| $10^7$ | 2·94E-05 | 3·67E-01 | 1·63E-04 | 3·07E-04 | 4·27E-05 | 1·47E-05 | 3·54E-06 | 3·67E-01 |
| $10^7$ | 5·79E-06 | 8·96E-06 | 1·25E-04 | 6·57E-06 | 3·33E-05 | 1·30E-05 | 1·88E-05 | 1·25E-04 |
| $10^8$ | 5·85E-06 | 3·31E-05 | 2·30E-05 | 2·83E-05 | 1·87E-04 | 1·35E-05 | 2·84E-05 | 1·87E-04 |
| $10^8$ | 4·51E-05 | 7·84E-03 | 6·16E-05 | 3·41E-05 | 1·10E-04 | 3·89E-05 | 3·57E-05 | 7·84E-03 |
| $10^8$ | 2·85E-05 | 3·80E-04 | 5·87E-05 | 1·38E-04 | 2·69E-04 | 2·12E-05 | 6·57E-06 | 3·80E-04 |
| $10^9$ | 4·04E-05 | 2·01E-04 | 2·83E-06 | 1·73E-04 | 3·96E-05 | 1·16E-04 | 2·29E-05 | 2·01E-04 |
| $10^9$ | 6·38E-05 | 8·72E-05 | 2·79E-04 | 1·89E-03 | 1·63E-04 | 6·93E-04 | | 1·89E-03 |
| $10^9$ | 1·96E-05 | 2·45E-05 | 5·12E-05 | 1·71E-05 | 6·31E-05 | 2·61E-05 | 1·80E-05 | 6·31E-05 |
| $10^{10}$ | 5·57E-05 | 2·34E-03 | 1·22E-05 | 2·17E-06 | | 1·47E-05 | 2·21E-05 | 2·34E-03 |
| $10^{10}$ | 5·02E-06 | 5·37E-04 | 3·09E-04 | | | 2·61E-05 | 1·09E-04 | 5·37E-04 |
| $10^{10}$ | | 6·15E-04 | 3·15E-04 | 7·41E-04 | 3·06E-04 | 5·03E-04 | 1·82E-03 | 1·82E-03 |

**Dose- Expansion Module**

| | Ogawa IgA (RAU) | | | | | | | |
|---|---|---|---|---|---|---|---|---|
| **Dose (CFU)** | **D1** | **D7** | **D15** | **D29** | **D57** | **D85** | **D180** | **max** |
| 0 | 2·53E-05 | | 2·12E-05 | 1·63E-05 | 1·54E-06 | | 9·84E-07 | 2·53E-05 |
| 0 | 1·23E-04 | 2·14E-05 | 1·15E-04 | 1·07E-04 | | | 1·36E-04 | 1·36E-04 |
| 0 | 1·88E-05 | 9·77E-06 | 1·06E-05 | 5·02E-06 | | | 4·58E-05 | 4·58E-05 |
| 0 | 2·05E-05 | 3·07E-05 | 5·52E-05 | 4·06E-05 | | | 5·33E-05 | 5·52E-05 |
| 0 | 2·14E-05 | 2·53E-05 | 4·95E-05 | 3·48E-06 | | | | 4·95E-05 |
| 0 | 4·75E-05 | 2·05E-05 | 5·81E-05 | 4·26E-05 | | | 4·26E-05 | 5·81E-05 |
| 0 | 5·91E-05 | 4·26E-05 | 2·46E-05 | 2·56E-05 | | | 4·94E-05 | 5·91E-05 |
| $2\times10^7$ | 1·06E-05 | 1·80E-05 | 3·73E-05 | | | | 2·12E-05 | 3·73E-05 |
| $2\times10^7$ | 3·25E-05 | 2·94E-04 | 3·65E-05 | 3·25E-05 | | | 3·25E-05 | 2·94E-04 |
| $2\times10^7$ | 2·14E-05 | 2·38E-03 | 2·93E-05 | 8·23E-05 | | | 1·37E-05 | 2·38E-03 |
| $2\times10^7$ | 3·48E-06 | 3·33E-05 | 3·48E-06 | 9·47E-05 | | | | 9·47E-05 |
| $2\times10^7$ | 3·48E-06 | 4·91E-03 | 1·60E-04 | 9·47E-05 | | | 1·52E-04 | 4·91E-03 |
| $2\times10^7$ | 3·48E-06 | 9·59E-02 | 9·16E-04 | 8·86E-04 | | | 2·71E-05 | 9·59E-02 |
| $2\times10^7$ | 1·44E-05 | 6·93E-04 | 3·72E-04 | 3·47E-05 | | | | 6·93E-04 |
| $2\times10^7$ | 2·86E-05 | 2·71E-02 | 2·76E-05 | 2·23E-04 | | | 1·23E-05 | 2·71E-02 |
| $2\times10^7$ | 3·73E-05 | 6·18E-05 | 6·59E-05 | 2·53E-05 | | | 6·54E-06 | 6·59E-05 |
| $2\times10^7$ | 1·03E-05 | 3·91E-03 | 2·84E-03 | 7·42E-05 | | | 4·84E-05 | 3·91E-03 |
| $2\times10^7$ | 3·48E-06 | 1·93E-02 | 5·16E-04 | 1·73E-04 | | | 3·63E-04 | 1·93E-02 |
| $2\times10^7$ | 4·06E-05 | 6·03E-04 | 8·71E-05 | 4·07E-04 | | | 7·32E-05 | 6·03E-04 |
| $2\times10^7$ | 3·27E-05 | 8·07E-05 | 1·04E-04 | 5·04E-05 | | | 4·26E-05 | 1·04E-04 |
| $2\times10^7$ | 2·66E-05 | 3·03E-04 | 1·25E-04 | 3·47E-05 | | | 2·26E-05 | 3·03E-04 |
| $2\times10^8$ | 5·79E-06 | 3·98E-04 | 8·46E-05 | 2·22E-04 | 1·30E-05 | | 1·14E-05 | 3·98E-04 |
| $2\times10^8$ | 1·14E-05 | 6·09E-05 | 4·35E-05 | 8·16E-06 | | | 9·77E-06 | 6·09E-05 |
| $2\times10^8$ | 2·26E-05 | 7·14E-05 | 4·36E-05 | 9·26E-05 | | | 1·33E-04 | 1·33E-04 |
| $2\times10^8$ | 1·37E-05 | 1·00E-01 | 2·48E-04 | 2·93E-05 | | | | 1·00E-01 |
| $2\times10^8$ | 5·36E-05 | 5·08E-03 | 5·77E-05 | 2·14E-05 | | | | 5·08E-03 |
| $2\times10^8$ | | | 4·13E-05 | | | | | 4·13E-05 |
| $2\times10^8$ | 7·41E-05 | 1·84E-04 | 1·19E-04 | 9·05E-05 | | | | 1·84E-04 |
| $2\times10^8$ | 3·57E-05 | 1·74E-04 | 3·07E-05 | 1·53E-04 | | | 1·47E-04 | 1·74E-04 |
| $2\times10^8$ | | 8·95E-03 | 5·53E-04 | 3·33E-05 | | | 3·69E-04 | 8·95E-03 |
| $2\times10^8$ | 1·60E-05 | 1·32E+00 | 1·68E-03 | 4·79E-03 | | | 2·32E-05 | 1·32E+00 |
| $2\times10^8$ | 6·54E-06 | 5·74E-04 | 1·46E-06 | 6·56E-05 | | | | 5·74E-04 |
| $2\times10^8$ | 2·57E-05 | 2·19E-04 | 7·48E-06 | 1·01E-04 | | | 1·48E-05 | 2·19E-04 |
| $2\times10^8$ | 7·14E-05 | 4·06E-05 | 4·06E-05 | 4·26E-05 | | | 4·94E-05 | 7·14E-05 |

**Dose-Escalation Module**

| CT-B IgM (RAU) | | | | | | | | |
|---|---|---|---|---|---|---|---|---|
| **Dose (CFU)** | **D1** | **D7** | **D15** | **D29** | **D57** | **D85** | **D180** | **max** |
| $10^{5}$ | 1·10E-03 | 4·13E-03 | 3·13E-05 | | | | | 4·13E-03 |
| $10^{5}$ | | | | 4·50E-02 | | | | 4·50E-02 |
| $10^{5}$ | 5·75E-04 | 1·19E-03 | 3·13E-05 | 9·17E-05 | | | | 1·19E-03 |
| $10^{6}$ | 2·73E-06 | 1·46E-02 | 1·25E-03 | | | | | 1·46E-02 |
| $10^{6}$ | | 3·60E-03 | | 3·10E-04 | | 1·20E-04 | | 3·60E-03 |
| $10^{6}$ | 4·18E-04 | 3·44E-02 | 8·79E-03 | 2·58E-03 | | 1·20E-04 | | 3·44E-02 |
| $10^{7}$ | 5·19E-03 | 1·00E-02 | 1·40E-02 | 1·01E-03 | 3·60E-03 | 5·88E-03 | | 1·40E-02 |
| $10^{7}$ | | 1·01E-03 | | | 7·66E-04 | | 7·68E-05 | 1·01E-03 |
| $10^{7}$ | | 3·38E-03 | 5·31E-04 | | 2·81E-03 | 1·65E-02 | 1·30E-04 | 1·65E-02 |
| $10^{8}$ | | 4·25E-03 | 1·20E-04 | | | 4·18E-04 | | 4·25E-03 |
| $10^{8}$ | 5·68E-02 | 2·34E-02 | 2·21E-01 | 1·62E-01 | 5·72E-02 | 1·36E-02 | 3·10E-04 | 2·21E-01 |
| $10^{8}$ | 5·31E-04 | 2·09E-02 | 1·50E-03 | | | | | 2·09E-02 |
| $10^{9}$ | 4·04E-03 | 1·64E-02 | 7·66E-04 | 4·57E-03 | | | 1·27E-02 | 1·64E-02 |
| $10^{9}$ | | 5·31E-04 | | 4·71E-04 | | | | 5·31E-04 |
| $10^{9}$ | | | 8·96E-04 | | | | | 8·96E-04 |
| $10^{10}$ | | 2·93E-03 | | | | | | 2·93E-03 |
| $10^{10}$ | | 3·10E-04 | | | | | | 3·10E-04 |
| $10^{10}$ | | 8·99E-03 | | | | 1·06E-03 | 6·17E-05 | 8·99E-03 |

**Dose- Expansion Module**

| CT-B IgM (RAU) | | | | | | | | |
|---|---|---|---|---|---|---|---|---|
| **Dose (CFU)** | **D1** | **D7** | **D15** | **D29** | **D57** | **D85** | **D180** | **max** |
| 0 | | | | | | | 1·59E-01 | 1·59E-01 |
| 0 | 7·27E-04 | 1·91E-02 | 4·36E-02 | 2·78E-04 | | | | 4·36E-02 |
| 0 | 2·75E-03 | 5·26E-05 | 7·82E-04 | | | | | 2·75E-03 |
| 0 | | | | 8·29E-05 | | | 1·17E-04 | 1·17E-04 |
| 0 | | 1·18E-03 | | | | | | 1·18E-03 |
| 0 | | | | | | | | |
| 0 | 1·44E-04 | 8·72E-04 | | 3·04E-04 | | | 5·75E-04 | 8·72E-04 |
| $2\times10^{7}$ | 1·05E-02 | 4·25E-03 | | 7·34E-02 | | | 3·50E-02 | 7·34E-02 |
| $2\times10^{7}$ | | | | | | | | |
| $2\times10^{7}$ | | | | | | | | |
| $2\times10^{7}$ | 6·21E-04 | 8·93E-04 | 2·78E-04 | 1·54E-03 | | | 2·31E-03 | 2·31E-03 |
| $2\times10^{7}$ | 1·54E-04 | 1·01E-02 | 1·73E-03 | | | | | 1·01E-02 |
| $2\times10^{7}$ | 1·12E-03 | | | 4·18E-04 | | | | 1·12E-03 |
| $2\times10^{7}$ | 3·70E-04 | | 1·36E-03 | | | | | 1·36E-03 |
| $2\times10^{7}$ | | 2·04E-03 | | 3·08E-03 | | | | 3·08E-03 |
| $2\times10^{7}$ | | | 5·17E-04 | 5·17E-04 | | | | 5·17E-04 |
| $2\times10^{7}$ | 3·70E-04 | 1·24E-03 | | 4·18E-04 | | | 5·75E-04 | 1·24E-03 |
| $2\times10^{7}$ | 2·35E-04 | 4·72E-02 | | 2·70E-04 | | | | 4·72E-02 |
| $2\times10^{7}$ | | 1·17E-02 | 1·29E-02 | 9·18E-04 | | | | 1·29E-02 |
| $2\times10^{7}$ | 7·12E-06 | | 1·98E-03 | 3·13E-05 | | | 1·73E-04 | 1·98E-03 |
| $2\times10^{7}$ | | 3·13E-05 | | | | | | 3·13E-05 |
| $2\times10^{8}$ | 1·41E-02 | | | | | | 1·04E-01 | 1·04E-01 |
| $2\times10^{8}$ | | 7·84E-02 | | | | | 1·51E-03 | 7·84E-02 |
| $2\times10^{8}$ | | 2·69E-03 | | 2·49E-03 | | | | 2·69E-03 |
| $2\times10^{8}$ | | 4·37E-03 | 1·60E-03 | | | | | 4·37E-03 |
| $2\times10^{8}$ | | 6·54E-03 | 6·21E-04 | 8·29E-05 | | | 2·50E-03 | 6·54E-03 |
| $2\times10^{8}$ | | | 3·24E-04 | | | | | 3·24E-04 |
| $2\times10^{8}$ | | 6·19E-03 | | 1·91E-03 | | | 7·19E-04 | 6·19E-03 |
| $2\times10^{8}$ | | 1·54E-04 | 5·17E-04 | | | | | 5·17E-04 |
| $2\times10^{8}$ | | 2·69E-03 | 2·95E-03 | 1·14E-02 | | | 5·14E-03 | 1·14E-02 |
| $2\times10^{8}$ | | 3·48E-03 | | | | | | 3·48E-03 |
| $2\times10^{8}$ | 3·13E-05 | 2·53E-03 | 1·42E-03 | | | | | 2·53E-03 |
| $2\times10^{8}$ | | | | | | | | |
| $2\times10^{8}$ | | | 7·00E-04 | | | | | 7·00E-04 |

**Dose-Escalation Module**

| | CT-B IgG (RAU) | | | | | | | |
|---|---|---|---|---|---|---|---|---|
| **Dose (CFU)** | **D1** | **D7** | **D15** | **D29** | **D57** | **D85** | **D180** | **max** |
| $10^{5}$ | | 5·43E-04 | 8·19E-03 | 1·68E-03 | | | | 8·19E-03 |
| $10^{5}$ | | | | 2·49E-04 | | | | 2·49E-04 |
| $10^{5}$ | | 3·60E-04 | 6·83E-04 | 8·26E-05 | | | | 6·83E-04 |
| $10^{6}$ | 8·38E-07 | 1·96E-02 | 1·50E-05 | 1·57E-06 | | 1·57E-06 | | 1·96E-02 |
| $10^{6}$ | | 3·46E-02 | 1·57E-06 | | 8·38E-07 | | | 3·46E-02 |
| $10^{6}$ | | 2·54E-01 | 7·72E-03 | 2·21E-04 | 7·43E-05 | 4·68E-06 | | 2·54E-01 |
| $10^{7}$ | | 3·26E-04 | 3·97E-04 | 1·08E-04 | 2·47E-06 | | | 3·97E-04 |
| $10^{7}$ | | 3·27E-05 | 3·35E-04 | 7·62E-05 | 1·33E-05 | | | 3·35E-04 |
| $10^{7}$ | | 4·42E-04 | | | | 2·00E-05 | | 4·42E-04 |
| $10^{8}$ | | 4·20E-05 | 8·64E-04 | 5·53E-05 | 1·57E-06 | | | 8·64E-04 |
| $10^{8}$ | | 1·34E-02 | 8·52E-04 | 4·68E-06 | | 3·63E-05 | | 1·34E-02 |
| $10^{8}$ | | 7·48E-03 | 3·02E-04 | 4·96E-05 | | | | 7·48E-03 |
| $10^{9}$ | | 3·33E-02 | 1·59E-04 | 9·32E-05 | | 5·68E-06 | 2·06E-05 | 3·33E-02 |
| $10^{9}$ | | | | | | | | |
| $10^{9}$ | | 3·75E-05 | 1·27E-04 | | 5·91E-05 | | | 1·27E-04 |
| $10^{10}$ | | 5·73E-04 | 2·37E-06 | | | | | 5·73E-04 |
| $10^{10}$ | | 6·86E-03 | 1·98E-04 | | | | | 6·86E-03 |
| $10^{10}$ | | 4·75E-02 | 1·26E-03 | 9·00E-05 | 5·30E-05 | 2·38E-05 | | 4·75E-02 |

**Dose- Expansion Module**

| | CT-B IgG (RAU) | | | | | | | |
|---|---|---|---|---|---|---|---|---|
| **Dose (CFU)** | **D1** | **D7** | **D15** | **D29** | **D57** | **D85** | **D180** | **max** |
| 0 | | | | | | | | |
| 0 | 1·65E-05 | 2·83E-05 | 3·37E-06 | 7·85E-06 | | | 3·28E-05 | 3·28E-05 |
| 0 | | | 3·06E-05 | 2·27E-06 | | | | 3·06E-05 |
| 0 | | | | | | | | |
| 0 | 3·37E-06 | 2·09E-05 | | | | | | 2·09E-05 |
| 0 | 2·80E-06 | | | | | | | 2·80E-06 |
| 0 | | | | | | | | |
| $2\times10^{7}$ | | 4·85E-01 | | 1·10E-03 | | | 1·38E-03 | 4·85E-01 |
| $2\times10^{7}$ | | | | | | | | |
| $2\times10^{7}$ | | | 1·50E-05 | 5·20E-06 | | | | 1·50E-05 |
| $2\times10^{7}$ | | 2·03E-03 | 4·61E-04 | 7·17E-06 | | | 1·58E-04 | 2·03E-03 |
| $2\times10^{7}$ | | 2·67E-04 | 1·13E-04 | 1·87E-05 | | | | 2·67E-04 |
| $2\times10^{7}$ | 5·85E-06 | 6·79E-04 | 5·96E-04 | | | | | 6·79E-04 |
| $2\times10^{7}$ | | 3·96E-06 | 3·63E-04 | 2·61E-04 | | | | 3·63E-04 |
| $2\times10^{7}$ | | 1·22E-02 | 1·35E-04 | 1·22E-04 | | | | 1·22E-02 |
| $2\times10^{7}$ | | | 2·91E-05 | 8·88E-05 | | | | 8·88E-05 |
| $2\times10^{7}$ | | 1·43E-04 | 2·31E-05 | | | | | 1·43E-04 |
| $2\times10^{7}$ | 5·95E-05 | 1·72E-01 | 5·55E-04 | 1·99E-04 | | | 8·68E-05 | 1·72E-01 |
| $2\times10^{7}$ | | 4·69E-03 | 1·95E-03 | 9·07E-05 | | | | 4·69E-03 |
| $2\times10^{7}$ | | | | | | | | |
| $2\times10^{7}$ | | 9·20E-04 | | | | | | 9·20E-04 |
| $2\times10^{8}$ | 3·03E-03 | | | | | | 1·31E-03 | 3·03E-03 |
| $2\times10^{8}$ | | 1·74E-01 | | | | | | 1·74E-01 |
| $2\times10^{8}$ | 5·85E-06 | 1·10E-02 | 3·31E-04 | | | | | 1·10E-02 |
| $2\times10^{8}$ | 1·28E-05 | 6·44E-02 | 2·31E-04 | 5·20E-06 | | | 1·09E-05 | 6·44E-02 |
| $2\times10^{8}$ | | 3·34E-01 | 1·60E-04 | 2·39E-05 | | | 1·77E-04 | 3·34E-01 |
| $2\times10^{8}$ | | | 1·95E-03 | | | | | 1·95E-03 |
| $2\times10^{8}$ | 1·43E-05 | 5·76E-02 | 1·17E-03 | 9·93E-05 | | | | 5·76E-02 |
| $2\times10^{8}$ | | 5·73E-05 | 1·10E-03 | 8·14E-05 | | | | 1·10E-03 |
| $2\times10^{8}$ | | | 5·88E-05 | 7·30E-04 | | | 4·85E-04 | 7·30E-04 |
| $2\times10^{8}$ | 2·54E-06 | 7·28E-02 | 5·05E-04 | 5·81E-04 | | | | 7·28E-02 |
| $2\times10^{8}$ | | | 1·86E-05 | 4·95E-06 | | | | 1·86E-05 |
| $2\times10^{8}$ | | | | | | | | |
| $2\times10^{8}$ | | 2·73E-04 | | 7·79E-06 | | | | 2·73E-04 |

**Dose-Escalation Module**

| | CT-B IgA (RAU) | | | | | | | |
|---|---|---|---|---|---|---|---|---|
| **Dose (CFU)** | **D1** | **D7** | **D15** | **D29** | **D57** | **D85** | **D180** | **max** |
| $10^{5}$ | 7·40E-04 | 1·61E-03 | 4·56E-02 | 1·78E-03 | | | 3·32E-04 | 4·56E-02 |
| $10^{5}$ | 2·96E-04 | 1·43E-05 | 1·33E-04 | 6·85E-04 | | | | 6·85E-04 |
| $10^{5}$ | 2·45E-05 | 2·56E-04 | 2·07E-03 | 1·69E-04 | | | | 2·07E-03 |
| $10^{6}$ | 8·65E-05 | 6·30E-01 | 6·25E-04 | 1·68E-04 | 2·29E-04 | 6·30E-05 | | 6·30E-01 |
| $10^{6}$ | 4·12E-04 | 4·13E+00 | 4·17E-03 | 5·17E-04 | 1·07E-04 | 2·71E-05 | 6·13E-05 | 4·13E+00 |
| $10^{6}$ | 6·36E-05 | 1·65E+00 | 4·11E-02 | 1·26E-03 | 5·36E-04 | 2·08E-04 | 2·50E-05 | 1·65E+00 |
| $10^{7}$ | 1·79E-04 | 6·70E-03 | 6·63E-03 | 2·12E-03 | 1·12E-04 | 6·69E-05 | 3·78E-05 | 6·70E-03 |
| $10^{7}$ | 7·52E-05 | 2·03E-03 | 5·70E-03 | 1·55E-04 | 8·76E-05 | 2·05E-05 | 1·04E-05 | 5·70E-03 |
| $10^{7}$ | 4·06E-05 | 4·46E-03 | 1·23E-04 | 6·27E-05 | 1·92E-05 | 3·52E-05 | 3·73E-05 | 4·46E-03 |
| $10^{8}$ | 5·02E-05 | 1·24E-02 | 4·44E-02 | 1·01E-03 | 2·92E-04 | 1·48E-04 | 1·29E-05 | 4·44E-02 |
| $10^{8}$ | 3·22E-04 | 1·86E-02 | 3·19E-03 | 1·23E-04 | 2·17E-04 | 4·03E-04 | 4·07E-05 | 1·86E-02 |
| $10^{8}$ | 7·93E-05 | 6·02E-01 | 1·28E-02 | 3·13E-03 | 4·66E-04 | 2·15E-04 | | 6·02E-01 |
| $10^{9}$ | 8·76E-05 | 2·91E-01 | 5·49E-04 | 1·01E-04 | 5·64E-05 | 6·37E-05 | 1·92E-05 | 2·91E-01 |
| $10^{9}$ | 1·09E-04 | 2·42E-04 | 1·38E-04 | 7·72E-05 | 7·14E-05 | 1·21E-04 | | 2·42E-04 |
| $10^{9}$ | 9·70E-06 | 1·12E-03 | 2·45E-03 | 1·40E-05 | 1·74E-04 | 2·44E-05 | 1·07E-05 | 2·45E-03 |
| $10^{10}$ | 6·68E-04 | 1·41E-03 | 2·64E-04 | 1·72E-04 | | 5·02E-05 | 4·18E-05 | 1·41E-03 |
| $10^{10}$ | 1·60E-05 | 1·71E-01 | 1·06E-03 | | | 9·02E-06 | 5·35E-06 | 1·71E-01 |
| $10^{10}$ | | 3·65E-02 | 1·27E-03 | 4·77E-04 | 1·55E-04 | 1·24E-04 | 3·03E-05 | 3·65E-02 |

**Dose- Expansion Module**

| | CT-B IgA (RAU) | | | | | | | |
|---|---|---|---|---|---|---|---|---|
| **Dose (CFU)** | **D1** | **D7** | **D15** | **D29** | **D57** | **D85** | **D180** | **max** |
| 0 | 1·01E-03 | 3·71E-04 | 1·14E-04 | 6·15E-04 | 7·91E-05 | | 5·56E-05 | 1·01E-03 |
| 0 | 3·42E-04 | 1·47E-04 | 4·27E-04 | 2·40E-04 | | | 4·70E-04 | 4·70E-04 |
| 0 | 1·31E-05 | 2·08E-05 | 1·68E-05 | 5·78E-05 | | | 1·28E-04 | 1·28E-04 |
| 0 | 1·91E-04 | 5·33E-04 | | 9·63E-06 | | | 2·33E-04 | 5·33E-04 |
| 0 | 5·48E-04 | 3·92E-06 | 7·30E-05 | 6·79E-05 | | | 1·17E-04 | 5·48E-04 |
| 0 | 6·55E-06 | 5·14E-07 | 1·84E-05 | 2·05E-05 | | | 1·60E-04 | 1·60E-04 |
| 0 | 4·17E-05 | 6·58E-05 | | | | | 2·94E-04 | 2·94E-04 |
| $2\times10^{7}$ | 1·97E-05 | 2·69E-03 | 7·95E-04 | | | | 4·60E-05 | 2·69E-03 |
| $2\times10^{7}$ | 6·56E-05 | 9·23E-03 | 5·80E-04 | 3·16E-04 | | | 7·08E-05 | 9·23E-03 |
| $2\times10^{7}$ | 4·79E-05 | 1·15E-04 | 1·68E-05 | 1·86E-06 | | | 1·36E-04 | 1·36E-04 |
| $2\times10^{7}$ | 4·79E-05 | 3·07E-03 | 2·30E-03 | 1·91E-04 | | | 3·07E-04 | 3·07E-03 |
| $2\times10^{7}$ | 2·87E-05 | 4·06E-03 | 7·86E-04 | 1·58E-03 | | | 1·75E-05 | 4·06E-03 |
| $2\times10^{7}$ | 3·32E-05 | 1·46E-02 | 5·23E-03 | 1·32E-04 | | | | 1·46E-02 |
| $2\times10^{7}$ | 7·81E-05 | 2·12E-04 | 1·14E-03 | 1·96E-04 | | | | 1·14E-03 |
| $2\times10^{7}$ | | 2·63E-01 | 3·45E-03 | 6·79E-05 | | | 2·25E-06 | 2·63E-01 |
| $2\times10^{7}$ | 2·00E-04 | 1·52E-04 | 1·02E-03 | 1·70E-03 | | | 1·53E-04 | 1·70E-03 |
| $2\times10^{7}$ | 1·05E-04 | 1·35E-04 | 1·14E-04 | 6·89E-04 | | | 9·83E-06 | 6·89E-04 |
| $2\times10^{7}$ | 6·79E-05 | 3·17E-01 | 1·79E-03 | 3·46E-04 | | | 1·18E-03 | 3·17E-01 |
| $2\times10^{7}$ | 1·87E-04 | 8·05E-02 | 1·46E-02 | 7·99E-04 | | | 1·03E-04 | 8·05E-02 |
| $2\times10^{7}$ | 5·39E-06 | 1·70E-04 | 9·62E-05 | 7·46E-05 | | | 5·29E-05 | 1·70E-04 |
| $2\times10^{7}$ | | 1·03E-03 | 3·78E-04 | 1·56E-07 | | | 7·53E-06 | 1·03E-03 |
| $2\times10^{8}$ | | 8·05E-04 | 1·22E-03 | 4·68E-04 | 7·46E-05 | | 4·25E-05 | 1·22E-03 |
| $2\times10^{8}$ | 7·40E-05 | 6·81E-03 | 3·34E-03 | 1·86E-03 | | | 5·02E-05 | 6·81E-03 |
| $2\times10^{8}$ | 1·42E-04 | 3·49E-02 | 6·67E-04 | 1·25E-03 | | | 1·14E-04 | 3·49E-02 |
| $2\times10^{8}$ | 7·30E-05 | 6·24E-01 | 1·03E-03 | 4·54E-04 | | | 1·69E-04 | 6·24E-01 |
| $2\times10^{8}$ | 5·14E-06 | 4·27E-01 | 8·09E-05 | 3·19E-04 | | | 3·06E-04 | 4·27E-01 |
| $2\times10^{8}$ | | | 8·17E-03 | | | | | 8·17E-03 |
| $2\times10^{8}$ | 1·80E-04 | 1·13E-01 | 1·92E-03 | 1·69E-04 | | | 8·33E-05 | 1·13E-01 |
| $2\times10^{8}$ | 9·38E-05 | 2·93E-05 | 3·55E-03 | 1·04E-03 | | | 2·37E-04 | 3·55E-03 |
| $2\times10^{8}$ | 2·02E-05 | 1·39E-03 | 2·29E-02 | 1·24E-02 | | | 1·40E-04 | 2·29E-02 |
| $2\times10^{8}$ | 9·23E-05 | 9·34E+00 | 1·14E-03 | 2·17E-03 | | | 5·06E-05 | 9·34E+00 |
| $2\times10^{8}$ | 5·39E-06 | 2·58E-05 | 8·38E-05 | 1·16E-04 | | | 1·73E-03 | 1·73E-03 |
| $2\times10^{8}$ | 8·48E-05 | 1·52E-04 | 2·43E-04 | 2·21E-04 | | | 2·25E-06 | 2·43E-04 |
| $2\times10^{8}$ | | 3·61E-05 | 5·92E-05 | 5·23E-05 | | | | 5·92E-05 |

**Dose-Escalation Module**

| TcpA IgM (RAU) | | | | | | | | |
|---|---|---|---|---|---|---|---|---|
| Dose (CFU) | D1 | D7 | D15 | D29 | D57 | D85 | D180 | max |
| $10^5$ | 1·40E-05 | 2·49E-05 | 2·74E-04 | 6·54E-04 | | | 2·49E-05 | 6·54E-04 |
| $10^5$ | | 1·68E-02 | 5·57E-03 | | | | | 1·68E-02 |
| $10^5$ | 1·40E-05 | 1·23E-02 | 2·44E-03 | 1·63E-04 | | | | 1·23E-02 |
| $10^6$ | | 1·67E-02 | 1·58E-03 | | 3·26E-03 | | | 1·67E-02 |
| $10^6$ | | 1·92E-02 | | 2·46E-03 | | | | 1·92E-02 |
| $10^6$ | | 9·44E-03 | 5·67E-03 | | | | | 9·44E-03 |
| $10^7$ | | 3·84E-03 | | | | | 5·16E-04 | 3·84E-03 |
| $10^7$ | | | | | | 1·47E-03 | | 1·47E-03 |
| $10^7$ | | 1·44E-02 | 2·39E-03 | | | 1·09E-02 | | 1·44E-02 |
| $10^8$ | 1·05E-03 | | | | | | | 1·05E-03 |
| $10^8$ | 1·15E-01 | 1·67E-02 | 5·36E+00 | 3·18E+00 | 1·03E-01 | 5·73E-03 | 1·64E-03 | 5·36E+00 |
| $10^8$ | | 7·86E-02 | 1·47E-03 | | | | 2·66E-05 | 7·86E-02 |
| $10^9$ | 9·48E-03 | 1·71E-02 | 2·54E-03 | 1·56E-02 | 8·29E-04 | 5·94E-04 | 1·41E-02 | 1·71E-02 |
| $10^9$ | | | | 1·77E-04 | | | | 1·77E-04 |
| $10^9$ | | 1·10E-04 | | | 3·24E-04 | 2·12E-04 | | 3·24E-04 |
| $10^{10}$ | | 1·31E-02 | 1·41E-03 | 7·51E-04 | | 2·66E-05 | | 1·31E-02 |
| $10^{10}$ | | 2·27E-03 | | | | | 4·38E-04 | 2·27E-03 |
| $10^{10}$ | | 3·10E-02 | | 3·24E-04 | | 1·45E-03 | | 3·10E-02 |

**Dose- Expansion Module**

| TcpA IgM (RAU) | | | | | | | | |
|---|---|---|---|---|---|---|---|---|
| Dose (CFU) | D1 | D7 | D15 | D29 | D57 | D85 | D180 | max |
| 0 | | 3·20E-02 | | | 5·51E-02 | | | 5·51E-02 |
| 0 | 2·24E-04 | 7·02E-05 | 3·30E-03 | 1·23E-04 | | | 6·16E-03 | 6·16E-03 |
| 0 | 6·36E-04 | 3·06E-04 | 9·48E-05 | 1·22E-05 | | | 1·42E-06 | 6·36E-04 |
| 0 | 2·44E-04 | 2·44E-04 | | 5·79E-04 | | | 4·47E-03 | 4·47E-03 |
| 0 | | 6·43E-07 | 1·76E-05 | 3·97E-04 | | | 2·49E-05 | 3·97E-04 |
| 0 | 1·54E-04 | | | 2·74E-04 | | | | 2·74E-04 |
| 0 | 7·32E-04 | 1·61E-03 | 1·02E-03 | 1·28E-03 | | | 5·46E-05 | 1·61E-03 |
| $2\times10^7$ | | 1·14E-01 | | 5·51E-02 | | | | 1·14E-01 |
| $2\times10^7$ | | 6·06E-01 | | | | | 9·55E-02 | 6·06E-01 |
| $2\times10^7$ | 7·80E-06 | 3·58E-02 | 5·51E-04 | 4·90E-05 | | | 1·01E-03 | 3·58E-02 |
| $2\times10^7$ | 4·42E-06 | 1·08E-03 | 7·24E-04 | 3·14E-05 | | | 7·32E-04 | 1·08E-03 |
| $2\times10^7$ | 1·76E-05 | 1·76E-02 | 7·80E-06 | 6·07E-04 | | | 7·32E-04 | 1·76E-02 |
| $2\times10^7$ | 2·06E-04 | 1·01E-03 | 3·14E-05 | 3·06E-04 | | | | 1·01E-03 |
| $2\times10^7$ | | 1·70E-04 | 4·72E-04 | | | | | 4·72E-04 |
| $2\times10^7$ | 2·40E-05 | 6·07E-04 | 1·70E-04 | | | | 6·17E-04 | 6·17E-04 |
| $2\times10^7$ | | 1·38E-04 | 4·90E-05 | 8·49E-04 | | | 1·40E-05 | 8·49E-04 |
| $2\times10^7$ | 3·97E-04 | 8·99E-03 | 1·88E-04 | 3·06E-04 | | | 1·15E-04 | 8·99E-03 |
| $2\times10^7$ | | 4·91E-02 | 3·02E-03 | 2·40E-03 | | | 1·38E-04 | 4·91E-02 |
| $2\times10^7$ | 4·02E-04 | 2·97E-03 | 2·16E-04 | 5·46E-05 | | | 2·49E-05 | 2·97E-03 |
| $2\times10^7$ | 3·85E-05 | 3·09E-01 | 1·70E-03 | 9·27E-05 | | | | 3·09E-01 |
| $2\times10^7$ | 1·63E-04 | 5·81E-03 | 3·04E-04 | 5·07E-04 | | | 7·27E-05 | 5·81E-03 |
| $2\times10^8$ | | | 4·01E-02 | | | | | 4·01E-02 |
| $2\times10^8$ | 7·59E-02 | | 6·01E-01 | | | | | 6·01E-01 |
| $2\times10^8$ | 4·42E-06 | 2·62E-03 | 1·76E-05 | 1·54E-04 | | | 4·02E-04 | 2·62E-03 |
| $2\times10^8$ | 6·07E-04 | 3·74E-04 | 8·21E-05 | 9·81E-04 | | | 1·63E-04 | 9·81E-04 |
| $2\times10^8$ | 7·80E-06 | 1·84E-02 | 4·46E-04 | 1·54E-04 | | | 1·19E-03 | 1·84E-02 |
| $2\times10^8$ | | | 9·81E-04 | | | | | 9·81E-04 |
| $2\times10^8$ | | 2·24E-04 | 1·01E-03 | 9·48E-05 | | | 5·93E-04 | 1·01E-03 |
| $2\times10^8$ | 5·24E-04 | 2·40E-05 | 1·76E-05 | 2·24E-04 | | | | 5·24E-04 |
| $2\times10^8$ | | 3·06E-04 | 1·38E-04 | 7·80E-06 | | | 7·85E-03 | 7·85E-03 |
| $2\times10^8$ | | 4·70E-04 | 1·42E-06 | | | | 3·36E-04 | 4·70E-04 |
| $2\times10^8$ | | 4·92E-03 | 6·66E-04 | | | | 8·53E-04 | 4·92E-03 |
| $2\times10^8$ | 1·42E-06 | | 1·89E-04 | 6·01E-06 | | | 2·16E-04 | 2·16E-04 |
| $2\times10^8$ | 4·70E-04 | 4·36E-04 | 2·49E-05 | 3·69E-04 | | | 1·40E-05 | 4·70E-04 |

**Dose-Escalation Module**

| TcpA IgG (RAU) | | | | | | | | |
|---|---|---|---|---|---|---|---|---|
| Dose (CFU) | D1 | D7 | D15 | D29 | D57 | D85 | D180 | max |
| $10^{5}$ | 2·71E-04 | 3·76E-04 | 1·41E-04 | 1·57E-04 | | | 1·33E-04 | 3·76E-04 |
| $10^{5}$ | 2·95E-04 | | 3·20E-04 | 4·40E-04 | | | | 4·40E-04 |
| $10^{5}$ | 5·68E-05 | 8·64E-05 | 1·49E-04 | 1·02E-04 | | | | 1·49E-04 |
| $10^{6}$ | | 1·39E-04 | | | | | | 1·39E-04 |
| $10^{6}$ | 3·61E-05 | 5·29E-04 | | | | | | 5·29E-04 |
| $10^{6}$ | | 6·79E-05 | | 1·58E-04 | 1·21E-04 | 1·06E-05 | | 1·58E-04 |
| $10^{7}$ | 1·21E-04 | | | 8·51E-05 | 2·22E-05 | 3·61E-05 | 8·20E-05 | 1·21E-04 |
| $10^{7}$ | | | | 8·51E-05 | 1·06E-05 | | 1·76E-05 | 8·51E-05 |
| $10^{7}$ | | | | 3·61E-05 | | 1·06E-05 | 4·84E-06 | 3·61E-05 |
| $10^{8}$ | 2·22E-05 | 1·06E-05 | 2·22E-05 | | | | 9·41E-07 | 2·22E-05 |
| $10^{8}$ | | 1·21E-04 | | 1·06E-05 | | 4·84E-06 | 4·84E-06 | 1·21E-04 |
| $10^{8}$ | 1·95E-04 | | | 3·08E-04 | 1·93E-04 | 9·41E-07 | 9·24E-05 | 3·08E-04 |
| $10^{9}$ | | 2·32E-04 | | 1·06E-05 | 5·21E-05 | 9·41E-07 | 1·06E-05 | 2·32E-04 |
| $10^{9}$ | | | 4·28E-05 | 2·54E-05 | 1·06E-05 | 3·38E-05 | | 4·28E-05 |
| $10^{9}$ | | 5·21E-05 | 1·25E-04 | 1·06E-05 | 4·28E-05 | 4·84E-06 | 1·06E-05 | 1·25E-04 |
| $10^{10}$ | 8·20E-05 | 3·38E-05 | 2·54E-05 | 9·41E-07 | | 5·21E-05 | | 8·20E-05 |
| $10^{10}$ | 4·84E-06 | | 1·06E-05 | | | | 4·28E-05 | 4·28E-05 |
| $10^{10}$ | | 3·38E-05 | 1·76E-05 | 1·81E-04 | 6·18E-05 | 7·18E-05 | 1·06E-05 | 1·81E-04 |

**Dose- Expansion Module**

| TcpA IgG (RAU) | | | | | | | | |
|---|---|---|---|---|---|---|---|---|
| Dose (CFU) | D1 | D7 | D15 | D29 | D57 | D85 | D180 | max |
| 0 | | | | | 2·35E-02 | | | 2·35E-02 |
| 0 | 1·63E-04 | | 4·05E-05 | 1·29E-04 | | | | 1·63E-04 |
| 0 | | | | 1·28E-06 | | | 4·56E-04 | 4·56E-04 |
| 0 | 1·28E-06 | 1·28E-06 | 2·09E-04 | | | | 1·81E-05 | 2·09E-04 |
| 0 | 1·55E-04 | | | | | | 1·82E-04 | 1·82E-04 |
| 0 | | 1·12E-05 | 1·33E-04 | 4·28E-05 | | | 1·02E-04 | 1·33E-04 |
| 0 | 4·28E-05 | 2·63E-04 | 1·02E-04 | 2·30E-04 | | | 1·17E-04 | 2·63E-04 |
| $2\times10^{7}$ | | | | 1·81E-02 | | | | 1·81E-02 |
| $2\times10^{7}$ | | 3·76E-02 | | | | | 1·54E-02 | 3·76E-02 |
| $2\times10^{7}$ | | 1·50E-05 | | | | | 6·66E-05 | 6·66E-05 |
| $2\times10^{7}$ | 5·30E-05 | 1·81E-04 | 1·28E-06 | 1·28E-06 | | | 1·98E-04 | 1·98E-04 |
| $2\times10^{7}$ | 1·12E-05 | 2·37E-04 | 2·85E-06 | | | | 8·64E-05 | 2·37E-04 |
| $2\times10^{7}$ | | 1·25E-05 | 1·96E-06 | 4·92E-06 | | | 3·04E-06 | 1·25E-05 |
| $2\times10^{7}$ | | 4·66E-05 | | 1·29E-04 | | | | 1·29E-04 |
| $2\times10^{7}$ | | 1·01E-05 | 5·60E-06 | | | | 2·79E-04 | 2·79E-04 |
| $2\times10^{7}$ | 1·21E-04 | 1·29E-04 | | | | | 1·49E-04 | 1·49E-04 |
| $2\times10^{7}$ | 2·92E-05 | 2·92E-05 | 1·12E-05 | 5·05E-06 | | | 2·14E-04 | 2·14E-04 |
| $2\times10^{7}$ | | | 2·06E-04 | 5·04E-04 | | | 1·49E-04 | 5·04E-04 |
| $2\times10^{7}$ | 3·36E-04 | 2·38E-04 | 1·65E-04 | 1·17E-04 | | | | 3·36E-04 |
| $2\times10^{7}$ | 3·44E-04 | 2·98E-05 | | 9·42E-05 | | | 2·71E-04 | 3·44E-04 |
| $2\times10^{7}$ | 2·22E-04 | 2·06E-04 | 8·64E-05 | | | | 1·49E-04 | 2·22E-04 |
| $2\times10^{8}$ | | | 2·57E-02 | | | | | 2·57E-02 |
| $2\times10^{8}$ | 6·05E-03 | | | | | | 3·28E-04 | 6·05E-03 |
| $2\times10^{8}$ | | 4·05E-05 | 5·05E-06 | 1·12E-05 | | | 1·82E-04 | 1·82E-04 |
| $2\times10^{8}$ | | 4·66E-05 | | | | | 3·92E-04 | 3·92E-04 |
| $2\times10^{8}$ | | 5·48E-04 | 3·47E-05 | | | | 2·55E-04 | 5·48E-04 |
| $2\times10^{8}$ | | | | | | | | |
| $2\times10^{8}$ | | 4·66E-05 | | | | | 1·73E-04 | 1·73E-04 |
| $2\times10^{8}$ | | 7·38E-05 | 4·66E-05 | 2·92E-05 | | | 1·65E-04 | 1·65E-04 |
| $2\times10^{8}$ | | 1·93E-05 | | 8·11E-05 | | | 1·73E-04 | 1·73E-04 |
| $2\times10^{8}$ | 2·55E-04 | 1·10E-04 | 1·33E-04 | 1·98E-04 | | | 1·17E-04 | 2·55E-04 |
| $2\times10^{8}$ | 2·47E-04 | 1·65E-04 | 2·63E-04 | 1·17E-04 | | | 2·14E-04 | 2·63E-04 |
| $2\times10^{8}$ | 7·87E-05 | 1·73E-04 | 1·33E-04 | 9·42E-05 | | | 1·65E-04 | 1·73E-04 |
| $2\times10^{8}$ | 1·33E-04 | 1·96E-03 | 5·27E-04 | 1·49E-04 | | | 5·68E-05 | 1·96E-03 |

**Dose-Escalation Module**

| TcpA IgA (RAU) | | | | | | | | |
|---|---|---|---|---|---|---|---|---|
| Dose (CFU) | D1 | D7 | D15 | D29 | D57 | D85 | D180 | max |
| $10^5$ | 2·34E-04 | 5·28E-04 | 6·40E-04 | 1·40E-03 | | | 2·90E-04 | 1·40E-03 |
| $10^5$ | 2·03E-05 | 7·72E-05 | 7·03E-05 | 6·37E-05 | | | | 7·72E-05 |
| $10^5$ | 1·76E-04 | 1·99E-04 | 4·74E-04 | 5·62E-04 | | | | 5·62E-04 |
| $10^6$ | 5·21E-04 | 7·39E-04 | 2·60E-04 | 6·28E-04 | 1·19E-04 | 1·95E-04 | | 7·39E-04 |
| $10^6$ | 8·15E-05 | 2·86E-03 | 6·55E-05 | 1·21E-04 | 2·04E-04 | 6·30E-04 | 7·00E-05 | 2·86E-03 |
| $10^6$ | 2·86E-04 | 7·56E-04 | 1·93E-04 | 3·85E-04 | 2·25E-05 | 6·33E-05 | 2·44E-05 | 7·56E-04 |
| $10^7$ | 2·16E-04 | 2·12E-03 | 3·80E-04 | 3·96E-04 | 1·00E-04 | 9·54E-05 | 3·77E-04 | 2·12E-03 |
| $10^7$ | 1·31E-04 | 2·79E-04 | 1·31E-04 | 5·21E-05 | 1·16E-04 | 8·84E-05 | 3·48E-05 | 2·79E-04 |
| $10^7$ | 1·61E-04 | 1·50E-04 | 1·19E-04 | 1·88E-04 | 1·45E-04 | 9·77E-05 | 9·28E-05 | 1·88E-04 |
| $10^8$ | 5·65E-05 | 1·45E-04 | 5·95E-04 | 1·19E-04 | 3·75E-04 | 1·02E-04 | 9·66E-05 | 5·95E-04 |
| $10^8$ | 6·52E-04 | 1·73E-03 | 3·65E-04 | 3·77E-04 | 2·88E-04 | 2·12E-04 | 7·93E-05 | 1·73E-03 |
| $10^8$ | 1·85E-04 | 1·90E-03 | 5·76E-04 | 1·45E-03 | 1·66E-03 | 1·41E-04 | 1·47E-04 | 1·90E-03 |
| $10^9$ | 1·42E-04 | 6·16E-04 | 9·77E-05 | 2·21E-04 | 1·15E-04 | 1·81E-04 | 5·48E-05 | 6·16E-04 |
| $10^9$ | 4·40E-04 | 2·52E-04 | 1·64E-04 | 6·16E-05 | 1·23E-05 | 5·70E-05 | | 4·40E-04 |
| $10^9$ | 2·09E-05 | 2·56E-04 | 2·10E-04 | 2·62E-05 | 1·68E-05 | 4·19E-05 | 8·42E-05 | 2·56E-04 |
| $10^{10}$ | 7·57E-05 | 2·77E-04 | 2·19E-03 | 7·59E-04 | | 1·45E-05 | 1·71E-06 | 2·19E-03 |
| $10^{10}$ | 2·03E-06 | 2·37E-04 | 1·52E-05 | | | 7·03E-07 | 1·60E-05 | 2·37E-04 |
| $10^{10}$ | 4·01E-06 | 1·11E-03 | 2·99E-05 | 2·12E-04 | 5·04E-05 | 6·27E-05 | 8·31E-06 | 1·11E-03 |

**Dose- Expansion Module**

| TcpA IgA (RAU) | | | | | | | | |
|---|---|---|---|---|---|---|---|---|
| Dose (CFU) | D1 | D7 | D15 | D29 | D57 | D85 | D180 | max |
| 0 | 1·40E-04 | 1·18E-04 | 1·30E-04 | 1·98E-04 | 3·63E-05 | | 9·63E-05 | 1·98E-04 |
| 0 | 4·46E-04 | 4·02E-04 | 7·43E-04 | 8·07E-04 | | | 8·71E-04 | 8·71E-04 |
| 0 | 1·35E-04 | 7·38E-05 | 6·68E-05 | 1·79E-04 | | | 2·88E-04 | 2·88E-04 |
| 0 | 1·99E-04 | 3·30E-04 | 2·75E-04 | 3·54E-04 | | | 3·98E-04 | 3·98E-04 |
| 0 | 2·96E-04 | 2·21E-04 | 3·30E-04 | 2·81E-04 | | | 3·30E-04 | 3·30E-04 |
| 0 | 3·57E-04 | 1·96E-04 | 1·33E-04 | 1·30E-04 | | | 2·40E-04 | 3·57E-04 |
| 0 | 4·25E-04 | 2·56E-04 | 1·53E-04 | 3·48E-04 | | | 2·18E-04 | 4·25E-04 |
| $2\times10^7$ | 8·77E-05 | 8·07E-04 | 1·49E-04 | 3·63E-05 | | | 7·55E-05 | 8·07E-04 |
| $2\times10^7$ | 2·78E-04 | 2·02E-04 | 1·05E-04 | 3·22E-04 | | | 2·62E-04 | 3·22E-04 |
| $2\times10^7$ | 2·14E-04 | 1·21E-03 | 9·21E-04 | 2·48E-04 | | | 3·88E-04 | 1·21E-03 |
| $2\times10^7$ | 3·43E-04 | 4·74E-04 | 1·40E-04 | 7·30E-04 | | | 1·19E-04 | 7·30E-04 |
| $2\times10^7$ | 2·80E-04 | 4·86E-03 | 8·07E-04 | 1·85E-03 | | | 6·37E-05 | 4·86E-03 |
| $2\times10^7$ | 2·48E-04 | 1·25E-03 | 4·72E-05 | 1·60E-04 | | | | 1·25E-03 |
| $2\times10^7$ | 1·40E-04 | 2·34E-03 | 3·73E-04 | 2·14E-04 | | | | 2·34E-03 |
| $2\times10^7$ | 2·31E-04 | 3·05E-04 | 2·31E-04 | 2·87E-04 | | | 2·25E-04 | 3·05E-04 |
| $2\times10^7$ | 6·73E-04 | 3·12E-04 | 5·31E-04 | 2·09E-04 | | | 1·70E-04 | 6·73E-04 |
| $2\times10^7$ | 1·78E-04 | 2·14E-04 | 1·20E-04 | 1·96E-04 | | | | 2·14E-04 |
| $2\times10^7$ | 3·75E-04 | 9·05E-04 | 4·01E-04 | 4·37E-04 | | | 1·36E-03 | 1·36E-03 |
| $2\times10^7$ | 3·42E-04 | 4·83E-04 | 3·48E-04 | 6·54E-04 | | | 1·33E-04 | 6·54E-04 |
| $2\times10^7$ | 5·81E-05 | 7·03E-04 | 5·62E-04 | 3·05E-04 | | | 2·47E-04 | 7·03E-04 |
| $2\times10^7$ | 2·56E-04 | 5·20E-04 | 2·87E-04 | 2·81E-04 | | | 3·18E-04 | 5·20E-04 |
| $2\times10^8$ | 9·37E-06 | 6·33E-05 | 1·13E-04 | 2·85E-04 | 1·55E-04 | | 3·12E-04 | 3·12E-04 |
| $2\times10^8$ | 1·30E-04 | 4·24E-04 | 2·68E-04 | 9·97E-05 | | | 1·37E-04 | 4·24E-04 |
| $2\times10^8$ | 1·20E-04 | 3·12E-04 | | 2·48E-04 | | | 3·96E-04 | 3·96E-04 |
| $2\times10^8$ | 2·96E-04 | 4·60E-04 | 3·43E-04 | 5·98E-04 | | | 2·46E-04 | 5·98E-04 |
| $2\times10^8$ | 1·78E-04 | 3·10E-03 | 2·64E-04 | | | | 3·00E-06 | 3·10E-03 |
| $2\times10^8$ | | | 3·42E-04 | | | | | 3·42E-04 |
| $2\times10^8$ | 5·71E-04 | 2·08E-03 | 1·02E-03 | 6·38E-04 | | | 8·10E-07 | 2·08E-03 |
| $2\times10^8$ | 1·53E-04 | 5·14E-04 | 2·37E-04 | 4·60E-04 | | | 8·37E-04 | 8·37E-04 |
| $2\times10^8$ | 2·18E-04 | 9·49E-04 | 3·78E-04 | 3·57E-04 | | | 2·18E-04 | 9·49E-04 |
| $2\times10^8$ | 7·03E-05 | 1·35E-03 | 1·54E-04 | 1·40E-03 | | | 2·25E-04 | 1·40E-03 |
| $2\times10^8$ | 3·12E-04 | 3·69E-04 | 5·34E-04 | 4·04E-04 | | | 1·06E-04 | 5·34E-04 |
| $2\times10^8$ | 2·52E-04 | 4·29E-04 | 1·65E-04 | 8·05E-04 | | | 1·40E-04 | 8·05E-04 |
| $2\times10^8$ | 5·11E-04 | 5·64E-02 | 3·48E-04 | 3·39E-04 | | | 1·23E-04 | 5·64E-02 |
